# A systematic review of Zika virus disease: epidemiological parameters, mathematical models, and outbreaks

**DOI:** 10.1101/2025.07.10.25331254

**Authors:** Kelly McCain, Anna Vicco, Christian Morgenstern, Thomas Rawson, Tristan M. Naidoo, Sangeeta Bhatia, Dominic P. Dee, Patrick Doohan, Keith Fraser, Anna-Maria Hartner, Sequoia I. Leuba, Shazia Ruybal-Pesántez, Richard J. Sheppard, H. Juliette T. Unwin, Kelly Charniga, Zulma M. Cucunubá, Gina Cuomo-Dannenburg, Natsuko Imai-Eaton, Edward S. Knock, Adam Kucharski, Mantra Kusumgar, Paul Liétar, Rebecca K. Nash, Sabine van Elsland, Pathogen Epidemiology Review Group, Nuno R. Faria, Anne Cori, Ruth McCabe, Ilaria Dorigatti

## Abstract

**Background:** Zika virus (ZIKV) is an Aedes-borne arbovirus that can cause serious risks of neurological complications and birth defects in newborns of mothers infected during pregnancy. ZIKV is classified as a priority pathogen by the World Health Organization.

**Methods:** We conducted a systematic review of peer-reviewed studies reporting ZIKV epidemiological parameters, transmission models, and outbreaks (PROSPERO #CRD42023393345) to characterise its transmissibility, seroprevalence, risk factors, disease sequelae, and natural history. We performed a meta-analysis of Congenital Zika Syndrome (CZS), pregnancy loss probabilities amongst ZIKV-infected mothers and the proportion of symptomatic cases.

**Findings:** We extracted information from 574 of 27,491 identified studies. Across 418 included studies assigned a high-quality score, we extracted 969 parameters, 127 outbreak records, and 154 models. We estimated a pooled total random effect of CZS probability (4.65%), pregnancy loss probability (2.48%), and proportion of symptomatic cases (51.20%). Seroprevalence estimates (n=354) were retrieved beyond South America and French Polynesia. The basic reproduction number estimates (n=77) ranged between 1.12 and 7.4. We found 66 human epidemiological delay estimates, ranging 4-12.1 days for the intrinsic incubation period (n=11), 3-50 days for the infectious period (n=15), 5.1-24.2 days for the extrinsic incubation period (n=22), and 7.4-32.9 days for the serial interval (n=27).

These data are available in the R package *epireview*.

**Interpretation:** This study provides the most comprehensive systematic summary of the ZIKV epidemiological information currently available in the literature. Large heterogeneities and inconsistencies in the reporting of parameter estimates, study designs, and parameter definitions were found, underscoring the need for standardised epidemiological definitions in future publications.

**Funding:** Academy of Medical Sciences, British Heart Foundation, Community Jameel, Diabetes UK, Imperial College London, National Institute for Health and Care Research, Royal Society, Schmidt Sciences, UK Department for Business, Energy, and Industrial Strategy, UK Medical Research Council, United Nations Foundation, Wellcome Trust.

**Research in Context:** *Evidence before this study:* We searched Web of Science and PubMed from database inception to October 31, 2024 to collate epidemiological parameters, mathematical models, and outbreaks of Zika virus (ZIKV). We found 66 systematic reviews on ZIKV since 2016: of these, 29 focussed on Congenital Zika Syndrome (CZS) and/or Guillain-Barré Syndrome, 19 on ZIKV transmission, 16 on the distribution and burden of ZIKV infection and disease and the remaining on clinical presentation, severity, forecasting, time-to-event, risk mapping and vaccines and therapeutics. The published systematic reviews included between 4 to 204 publications, and most (75%) had a global scope. Only 5 systematic reviews conducted a meta-analysis assessing the overall probability of CZS upon infection in the mother, and only 1 review investigated the probability of CZS by trimester of pregnancy (using 2 articles).

*Added value of this study:* In this systematic review, we extracted information from 574 relevant studies, which makes this the most comprehensive synthesis of Zika parameters for outbreak response to date. From 418 studies with a high-quality assessment score, we extracted 969 epidemiological parameters, 127 outbreak records and 154 transmission models, most of which were published after the 2016 outbreak. We extracted and analysed 354 geolocated and time-stamped seroprevalence estimates; 100 basic and 15 effective reproduction number estimates from 21 countries; 66 human epidemiological delays and 22 extrinsic incubation periods. Our meta-analysis provides robust pooled estimates of the proportion of symptomatic cases (51.20%) and CZS probability (4.65%), also stratified by trimester based on maternal ZIKV infection (based on 7 publications), addressing a gap present in most previous reviews.

*Implications of all the available evidence:* This study provides the most comprehensive overview of ZIKV epidemiology to date, serving as a valuable resource for future modelling studies and stakeholders developing preparedness and response plans to control future outbreaks.

## Introduction

Arboviruses pose a substantial threat to public health, with the World Health Organization (WHO) reporting close to 5.5 billion people at risk globally.^1,2^ Zika virus (ZIKV) is an arbovirus of concern that is mainly transmitted via *Aedes* mosquitoes but also human-to-human via sexual transmission.^3^ Initially discovered in Uganda in 1947, until the mid-2000s, ZIKV was only detected within the African and Asian continents.^4^ The first outbreak of ZIKV outside of these regions occurred in the Federated States of Micronesia in 2007,^5^ with the virus subsequently spreading to the Americas.^6,7^ An increase in cases of ZIKV, microcephaly, Congenital Zika Syndrome (CZS) and Guillain-Barré syndrome (GBS) in Brazil in 2015 led to the declaration of a Public Health Emergency of International Concern (PHEIC) by WHO from February to November 2016.^8–10^

Whilst most ZIKV infections are asymptomatic,^3,5,11^ the serious complications in the foetus and negative pregnancy outcomes associated with ZIKV infection during pregnancy, including CZS and its association with GBS and other neurological disorders, underscore why future ZIKV re-emergence remains a global public health concern.^10^ To date, no licensed vaccines or therapeutics exist, and although ZIKV cases have declined globally since 2017, they continue to be detected worldwide.^12^ ZIKV remains listed among the pathogens with pandemic potential and is prioritised for research and development by WHO^13,14^ and the UK Health Security Agency.^15^

Mathematical models play a critical role in outbreak response and risk assessment by providing insights into the potential location, timing, and magnitude of disease transmission. These tools can help guide the selection of sites for vaccine or therapeutic trials and inform the prepositioning of protocols and other preparedness activities. Central to these activities is the need for quantitative estimates of key epidemiological parameters, such as the reproduction number, epidemiological delays and seroprevalence, together with their variation and a catalogue of existing mathematical models. Here, we conducted a systematic review to generate a comprehensive database to aid the global modelling and public health community in their response to future ZIKV outbreaks. This systematic review is part of a set of reviews of nine WHO priority pathogens^16–19^ and contributes to a wider central, up-to-date and accessible database of information critical to mathematical modelling for timely outbreak response.

## Methods

We followed the Preferred Reporting Items for Systematic Reviews and Meta-Analyses (PRISMA) guidelines and registered our study protocol with PROSPERO (International Prospective Register of Systematic Reviews, #CRD42023393345). The checklist is included in the Appendix (pp 116-119).

### Search and screening

We searched PubMed and Web of Science for studies published from database inception to October 31, 2024. Results were imported into Covidence^20^ and de-duplicated. Titles, abstracts, and then full texts were independently screened by two reviewers, and conflicts were resolved by consensus. The Cohen’s Kappa scores for screening and full-text review are reported in Appendix Figure A1. Non-peer-reviewed literature and non-English language studies were excluded. Systematic reviews (n=66) were excluded from extraction (Appendix: Section A), but peer-reviewed literature identified through backwards citation screening meeting the selection criteria was included. Further details are provided in the Appendix (pp 6).

### Data extraction

Of the full texts meeting the inclusion criteria, 16% (n=91) were randomly selected for double extraction to ensure concordance between a team of 16 extractors. After consensus was reached, reviewers independently conducted single extractions on the remaining full texts.

Data were extracted using a custom-made Microsoft Access database (version 2305) (Appendix Tables A4–A6). We collected information on publication details, transmission models, outbreaks and epidemiological parameters including basic and effective reproduction numbers, epidemiological delays (the incubation period, the time between symptom onset to hospitalisation or from symptom onset to outcome, the extrinsic incubation period), case fatality ratios (CFRs), attack rates, growth rates, evolutionary, mutation and substitution rates, the proportion of symptomatic cases, overdispersion (quantifying heterogeneity in transmission), seroprevalence, probability of CZS/microcephaly in newborns born to infected mothers, probability of pregnancy loss in infected pregnant women, relative contributions to transmission from different routes and risk factors related to Zika or CZS. Pregnancy loss probability was extracted as either the probability of miscarriage if the study explicitly reported this, or general pregnancy loss, which included both the sum of miscarriage and stillbirth and pregnancy loss with no specified information on trimester of loss.

We extracted estimates of the basic and effective reproduction numbers (*R*_0_ and *R*_e_, representing the average number of secondary infections generated by a case in a fully susceptible and partially susceptible population, respectively), the basic vector-borne reproduction number (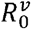, representing the average number of new infections generated via vector-borne transmission in a fully susceptible population), and the basic and effective sexual reproduction number (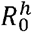and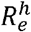, representing the average number of new cases generated via sexual transmission in a fully susceptible and partially susceptible population, respectively).

We used a custom quality assessment tool to assess the quality of each study (Appendix Table A2). Full details on the data extraction process and extracted data are provided in the Appendix (pp 6-8).

### Data analysis

The analysis was conducted in R using the *orderly2* workflow package.^21^ The quality score of each study was calculated as the proportion of ‘yes’ answers to the total number of applicable questions in the quality assessment tool for that study. Only parameters with a quality score ≥ 50% were included in the analyses presented in the main text. Sensitivity analyses on the results, regardless of the quality assessment of the studies, are presented in the Appendix (Section B4).

We conducted meta-analyses for the proportion of symptomatic cases, probability of CZS and probability of pregnancy loss among confirmed ZIKV-infected mothers. For the latter two, we only included estimates with at least 10 pregnant women with confirmed ZIKV infection and with a study design that did not select for the outcome (CZS or miscarriage), using the meta R package.^22^ We used random (RE) and fixed (FE) effects models using the logit link function to calculate pooled estimates for both parameters, with 95% confidence intervals and I2 heterogeneity estimates. We report the RE in the paper, the FE is reported in meta-analysis plots (Figure 5 and Appendix: Section B5). We estimated the pooled CZS and pregnancy loss probabilities by country, continent, and sample population type using maximum likelihood to estimate the between-studies variance. Subgroup analyses were considered to assess differences in CZS and pregnancy loss probabilities. Funnel plots assessing publication bias are shown in the Appendix (Figures B29 and B30).

## Results

The PRISMA flowchart is shown in Figure 1. From the 27,491 studies identified through the database search, 11,845 studies were screened following de-duplication, of which 101 were identified through backwards citation screening of the systematic reviews. A total of 10,500 studies were excluded at the abstract/title screening stage, and we undertook full text review of the remaining 1,343 studies, of which 574 met the criteria for inclusion (Figure 1, Table A1). We extracted 159 outbreak records, 229 models, and 1,334 parameters, of which 127 outbreak records, 154 models, and 969 parameters belonged to publications that met the quality score ≥ 50% and are included in our main analysis. All extracted data and the full list of publications included in the main text of this review are available in the R package *epireview*.^23^ The full analysis, including all publications, regardless of their QA score, is provided in the Appendix (Figures B.25-B28). Information about the extracted models is provided in Table B7 and Figure B4.

**Figure 1.**
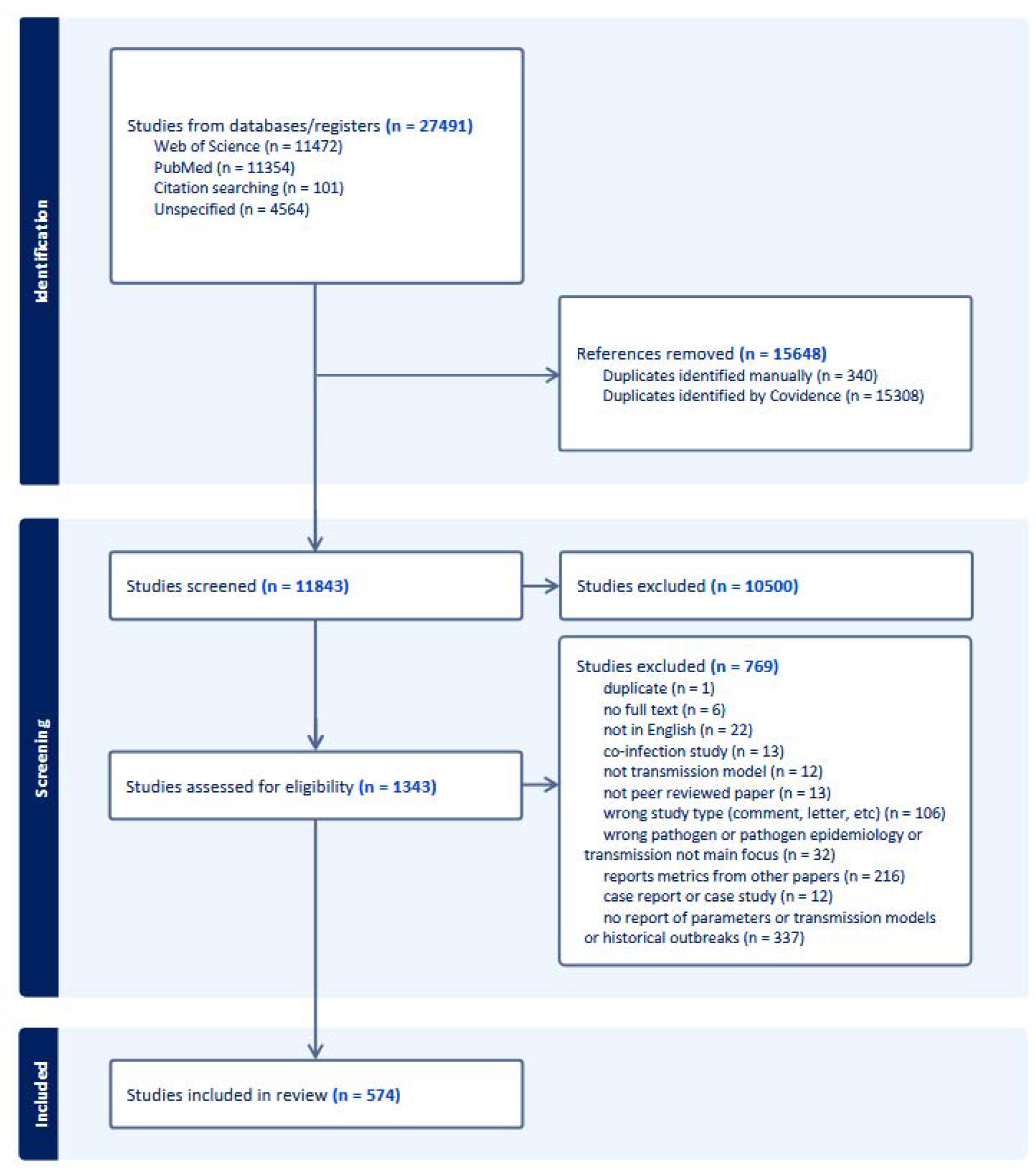
Flowchart for study selection. Studies were screened according to the PRISMA guidelines (see Appendix pp 116-118). Reasons for exclusion at the abstract/title stage were not collected by the Covidence software.

### Outbreaks

Forty studies reported 127 outbreak records between 2006 (Thailand) and 2021 (India) from 29 countries across the Americas (n=97), Oceania (n=13), Asia (n=7), Africa (n=4), and Europe (n=2) (Figure 2, Appendix: Table B8). The largest number of records was reported in Colombia across several subregions (n=55), followed by French Polynesia (n=12) and Suriname (n=11). The largest ZIKV outbreaks were reported in Puerto Rico between 2015 and 2016 (with 39,717 suspected cases, of which 36,390 were laboratory-confirmed) and in Colombia between 2015 and 2017 (with 108,087 suspected cases, 62% of which were among women, and 9,802 of which were laboratory-confirmed).

**Figure 2:**
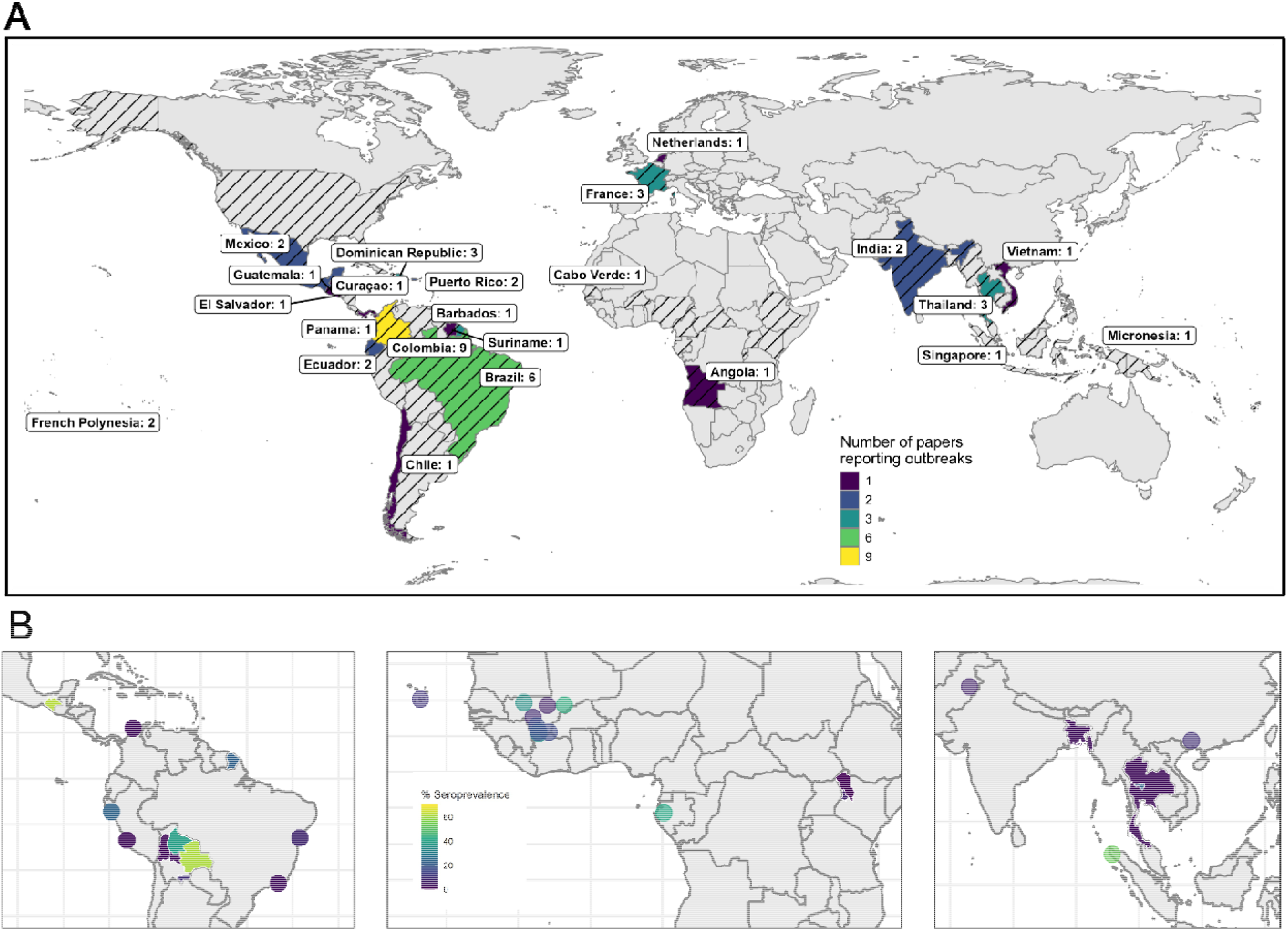
ZIKV outbreak and seroprevalence mapping: (A) Countries with studies reporting ZIKV outbreak information, coloured by number of studies. Outbreaks reported in France and the Netherlands reflect outbreaks in overseas regions. Countries with black diagonal stripes indicate locations where ZIKV transmission has been reported by the WHO 77. (B) Geolocated areas or regions with ZIKV seroprevalence studies (using IgG assay, HAI/HI, MIA, NS1 BOB ELISA, IFA, capture ELISA and neutralisation assays) conducted in the general population in the Americas (left), Africa (centre) and Asia (right). Each dot represents a location-specific estimate, while shaded areas indicate estimates at the administrative unit level (region, province, district, or entire country).

### Parameters

The 969 extracted parameter estimates came from 281 studies, comprising seroprevalence (n=354), risk factors (n=242), reproduction numbers (n=117), CZS probability (n=53), epidemiological delays in humans (n=66), attack rates (n=52), extrinsic incubation period (n=22), evolutionary rate (n=15), substitution rate (n=3), pregnancy loss probability (n=23), CFR (n=6), proportion of symptomatic cases (n=10), relative contributions to transmission (n=2) and growth rates (n=4) (Appendix: Figure B3).

### Seroprevalence

A total of 139 studies reported 354 ZIKV seroprevalence estimates from serosurveys conducted between 1980 (Nigeria^24^) and 2022 (El Salvador^25^) in 59 countries across the Americas (n=143), Africa (n=97), Asia (n=62), Europe (n=36) and Oceania (n=16). All seroprevalence estimates extracted in this study are reported in Appendix Table B10. Over one-third of these estimates were from surveys of the general population (n=107), while the rest was from pregnant women (n=58), suspected ZIKV infection (n=64), children (n=35), mixed population groups (n=35), blood donors (n=10) and other at-risk populations (n=43) (Appendix: Figures B9, B10). The timing of serosurveys coincided with the major epidemics (Appendix: Figure B9, B10). The majority of ZIKV seroprevalence estimates were derived from Immunoglobulin G (IgG) assays (n=139) and neutralisation tests (NT, n=109) (Appendix: Figure B11).

Seroprevalence estimates varied widely from 100% in a hospital-based study of 29 individuals in Cúcuta, Colombia,^26^ to 0% in local regions in Austria,^27^ Iran,^28^ and Taiwan,^29^ and specific locations within Brazil,^30,31^ Bolivia,^32^ Kenya,^33^ Indonesia,^34^ and Thailand.^35^ Within Brazil, ZIKV exposure in Salvador (Bahia) ranged between 7% (95%CI 4-10%) in 2014 and 63% (95%CI 60-65%) in 2015,^36^ consistent with the timing of the major outbreak in the Americas. In contrast, ZIKV seroprevalence in Rio de Janeiro was reported to be 3% in 2018.^37^ In Asia, a national serosurvey conducted in Thailand during 2017-2020 yielded an average seroprevalence of 2.8%^38^ but estimates were as high as 23.5% from a serosurvey conducted in 2011-2012 in Nakhon Sawan city.^39^ In Africa, serosurveys conducted from 2013 to 2017 reported ZIKV seroprevalence less than 11.5% in several sites of Kenya,^33^ 42.1% in Gabon,^40^ 10.9% in Cabo Verde^41^ and spanning 3.1% to 43.3% in Mali.^28^

### Transmissibility

We extracted estimates of the basic reproduction number R_0_ (n=77), human-to-human (sexual)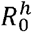, and vector-borne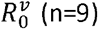, effective reproduction number R_e_(n=15)and sexual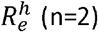,from 40 studies. The methods used to estimate R_0_ and R_e_ varied across the different studies. The next generation matrix method was the one used most frequently (n=27), followed by compartmental models (n=18), and branching processes (n=16). Most estimates were from the 2013-2014 outbreaks in French Polynesia (n=24), and the 2015-2016 epidemics in Brazil (n=26) and Colombia (n=15).

The majority (63/77) of the central estimates of R_0_ were between 1.5 and 4 (Figure 3). Estimates of the other transmission type-specific reproduction numbers are shown in the Appendix (Figure B5 and B7, Table B12).

**Figure 3.**
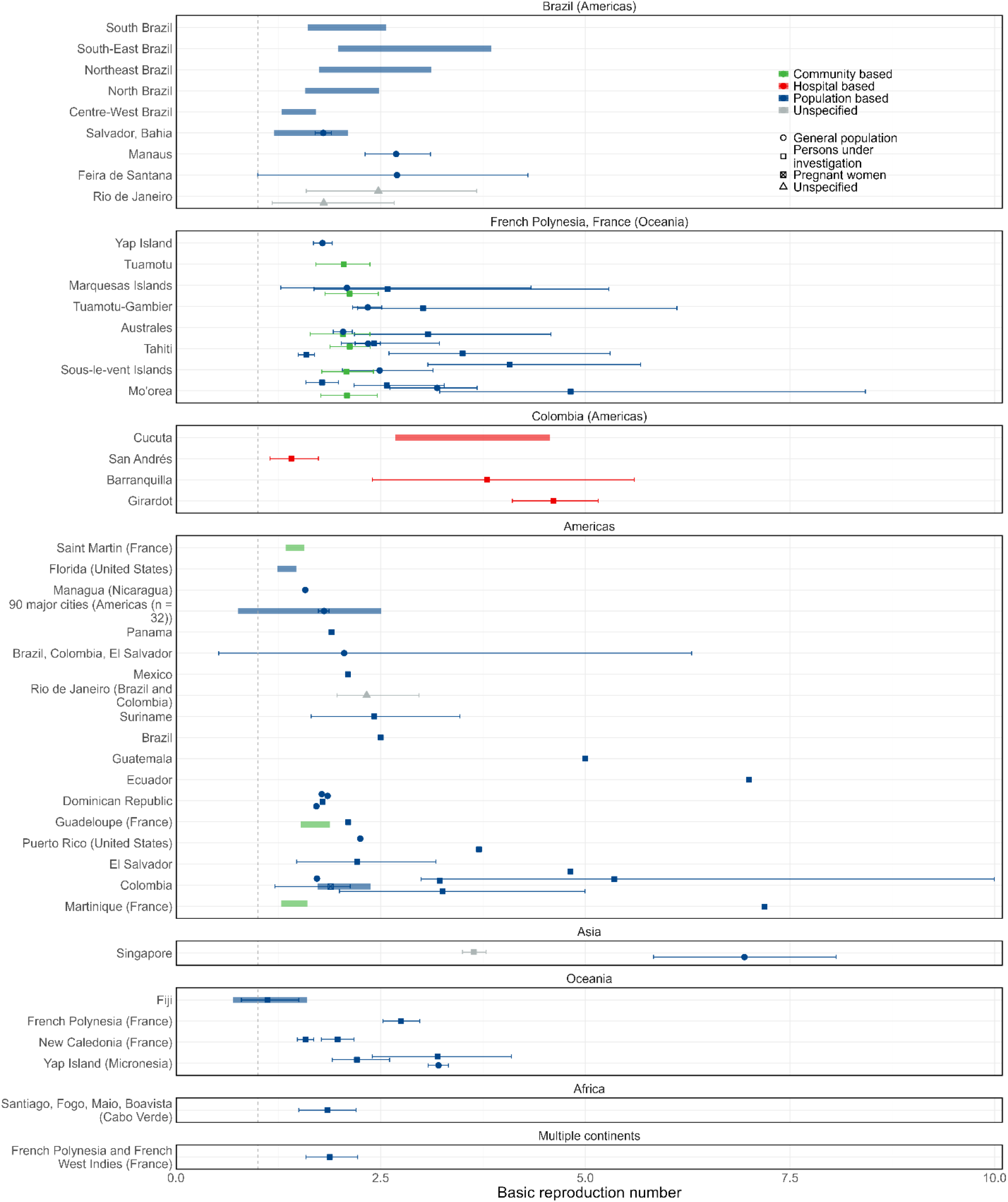
ZIKV basic reproduction number estimates. R_0_ estimates by geographic location, type of sample, and population group. Sub-national estimates for Brazil, Colombia, and French Polynesia are shown in separate panels because of the high number of estimates; all other estimates are shown in the respective continent panels. Overall country estimates for Brazil, Colombia, and French Polynesia are shown in the corresponding continent panels. Points are central estimates reported in the published studies, error bars are 95% confidence or credible intervals, thicker shaded bars are ranges of central estimates over disaggregated groups. The grey vertical dotted line marks 1. When multiple estimates for the same location were available, the estimates were jittered.

### Epidemiological delays

We analysed 66 human epidemiological delays from 29 studies and 22 extrinsic incubation period (EIP) estimates from 12 studies (Figure 4). The 11 central intrinsic incubation period estimates ranged from 4 days (95%CI 5.48-3.15) in Australes, French Polynesia in 2013-2014^42^ to 12.1 days in Brazil in 2016^43^, though roughly half of the reported estimates (n=6/11) were between 5 and 7 days. The 15 estimates of the human infectious period were highly variable, ranging from central estimates of 3 days in Brazil in 2016^43^ to 50 days among asymptomatic cases in Florida, United States^44^ in July to September 2016. We extracted only 2 estimates from 2 studies of the serial interval: a mean of 7.4 days (95%CI 4.59-10.2) in Singapore in August to November 2016^45^ and of 32.9 days in French Polynesia between October 2013 and October 2016.^46^ The single extracted estimate for the generation time was estimated from data collected in the French West Indies (Guadeloupe, Martinique, and Saint-Martin) in 2015-2017 and was described by a mean of 2.5 weeks (SD 0.7).

**Figure 4.**
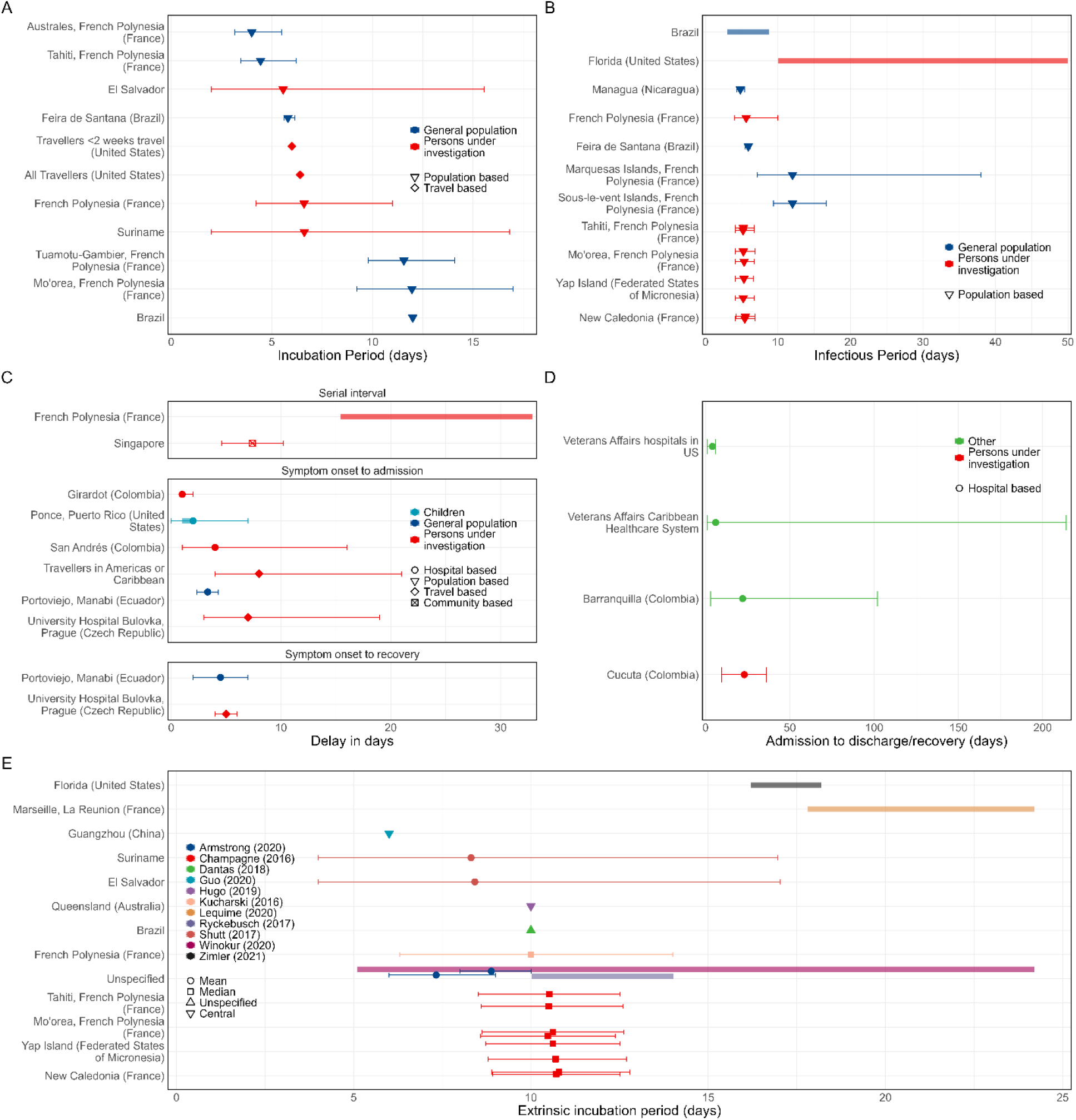
Epidemiological delays in human and mosquitoes. Estimates by location and type of sample of Zika (A) intrinsic incubation period, (B) infectious period, (C) serial interval, symptom onset to admission, and symptom onset to recovery, and (D) admission to discharge/recovery. Estimates by location and study of ZIKV extrinsic incubation period in the mosquito (E). Points are the central estimates reported in the studies, error bars are 95% confidence or credible intervals, and shaded bars are ranges of central estimates over disaggregated groups.

The 22 central estimates of the EIP ranged from 5.1^47^ to 24.2 days.^47,48^ The central estimates of the time from symptom onset to admission to care ranged from 1 (IQR 1-2) day among hospitalised patients in Colombia^49^ to 8 (IQR 4-21) among travellers.^50^ The central estimates of time from admission to recovery or discharge ranged from 4^51^ to 23 (IQR 9.5-36)^26^ days. All epidemiological delays extracted in this study are listed in Table B9 of the Appendix.

### Severity and adverse birth outcomes

We extracted 53 estimates of CZS probability given a ZIKV-infected mother from 36 studies. The CZS probability was highly variable across studies (Figure 5A), 92.5% (49/53) of estimates were from the Americas, and all estimates were obtained from data collected between January 2015 and February 2020.

**Figure 5.**
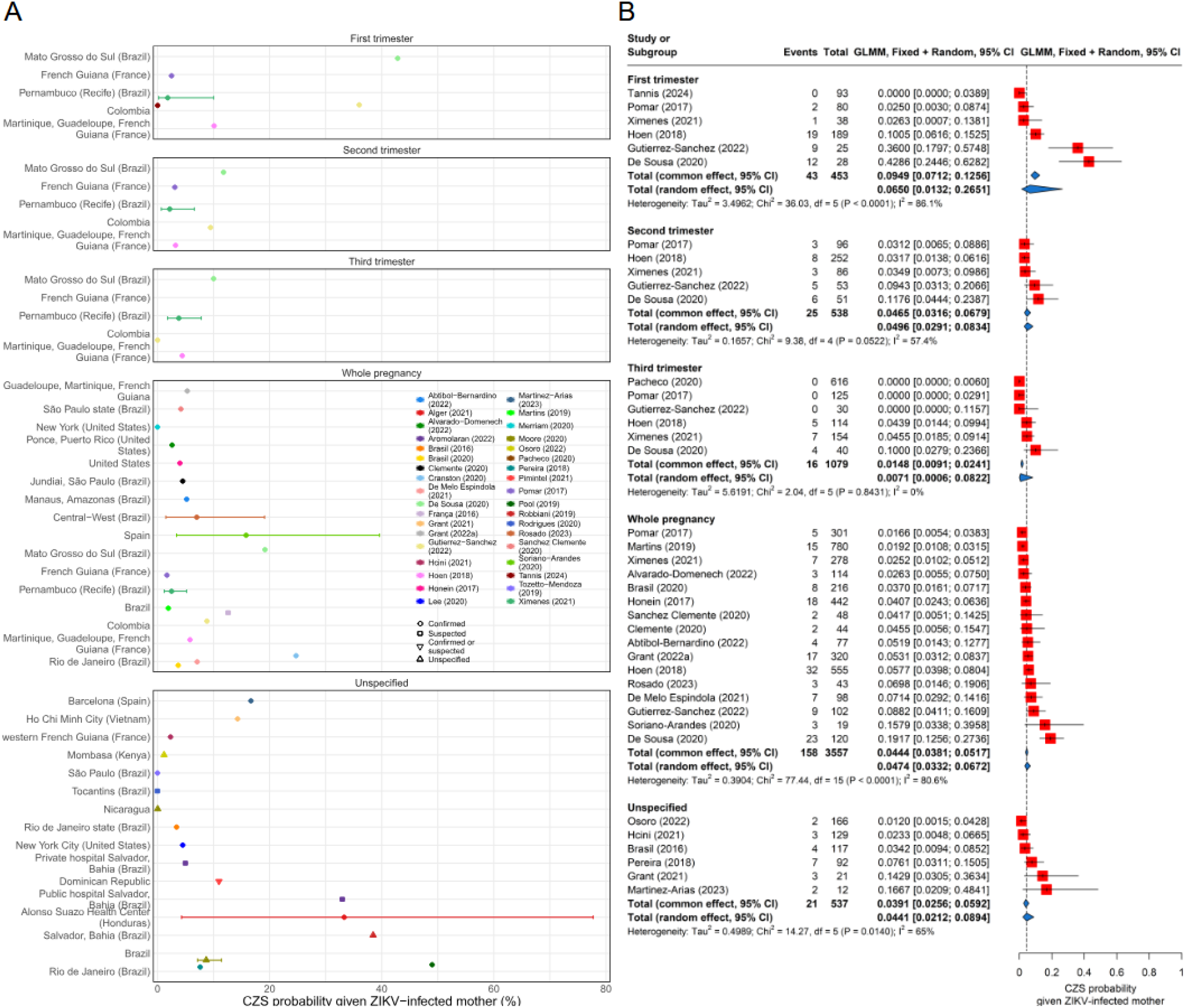
Risk of Zika congenital syndrome (CZS). (A) Estimates of reported CZS risks from Brazil studies (top) and from the rest of the world (bottom). Points are central estimates, solid lines are confidence or credible intervals, and shaded segments are ranges of central estimates across multiple groups. (B) Meta-analysis of CZS risk stratified by population sample type. Red squares represent the observed study effect sizes, the solid black horizontal lines are confidence intervals, and blue diamonds are the pooled estimates for each sub-group and overall. The vertical dashed line is the overall pooled estimate.

Under our stringent inclusion criteria, 39 estimates from 24 studies were eligible for meta-analysis. The pooled CZS probability, restricted to non-trimester-specific estimates, was 4.65% (95%CI: 3.38%-6.37%) using the random effects (RE) model (I^2^=77.1%) (Appendix: Figure B33). Stratifying the meta-analysis by trimester of maternal ZIKV exposure shows a higher pooled estimate of CZS probability in the first trimester (6.50% (95%CI: 1.31%-26.51%, I^2^=86.1%) compared to the second (4.96% [95%CI: 2.91%-8.34%]) or third (0.71% [95%CI: 0.06%-8.22%]) trimester (Figure 5B). Sensitivity analyses of the CZS probability by continent showed that the estimate for the Americas (4.43% [95%CI: 3.21%-6.07%]) was lower than that from travel-based studies in Europe (16.13% [95%CI: 6.88%-33.37%]) (Appendix: Figure B31). The RE estimate for Brazil (5.80% [95%CI: 3.74%-8.91%]) was marginally higher than the overall estimate (Appendix: Figure B32).

We extracted 23 central estimates of the probability of pregnancy loss associated with maternal ZIKV infection, which were derived from 17 studies and ranged between 0%^52–54^-25%^55^ (Appendix: Figure B14). Of these, we included only 14 studies (20 estimates) in the meta-analysis because of our more stringent selection criteria. The pooled overall estimates of the probability of pregnancy loss were 2.48% (95%CI: 1.62%-3.78%) using the RE model (Appendix: Figure B33). When we stratified based on the time of pregnancy loss, the pooled estimate of miscarriage probability (2.04% [95%CI: 1.21%-3.43%]) was marginally lower than the pooled estimate for general pregnancy loss (3.16% [1.76%-5.61%]) (Appendix: Figure B34).

The CFR estimates (Appendix: Figure B15, Table B14) extracted from 5 studies conducted between 2015 and 2017 in the Dominican Republic,^56^ the West Indies,^57^ Brazil,^58,59^ and Colombia^60^ varied from 0.04% reported in a population-based study of 5,161 individuals in the Dominican Republic in 2016,^56^ to 38% in a small hospital-based cohort of 16 persons under investigation in Brazil in 2016.^59^ The proportion of symptomatic individuals (Appendix: Figure B16, Table B14) was reported in 3 studies, yielding 10 estimates ranging from 23.2% in pregnant women in French Guiana^61^ to 71% (95%CI: 66-76%) in children in the Society Islands (French Polynesia).^62^ Of the 3 studies, 2 had sample size information and thus were included in our meta-analysis, which showed a pooled estimate of the proportion of symptomatic cases of 51.20% (95%CI: 38.00%-64.23%, I^2^=96.7%) (Appendix: Figures B35-B36).

## Discussion

In this systematic review, we collated and analysed epidemiological parameters, transmission models, and outbreak records of ZIKV to inform future modelling in response to ZIKV outbreaks. One of the clearest findings of this review is the concentration of Zika-related publications within a narrow time window—from the 2015 outbreak in Brazil to the onset of the COVID-19 pandemic in 2020. The 2015-

2016 epidemic in Latin America marked a turning point in ZIKV research: before this, only 6 of the 574 studies included in this review were published, none of which involved modelling. With the emergence of ZIKV as a global health concern, research activity surged, particularly following the outbreak in Brazil in 2015 and the subsequent PHEIC declaration, resulting in a sharp increase in publications (often with a QA score < 50%) (Appendix: Figure B2). Since 2020, however, the volume of new studies has declined, likely due to reduced data availability and shits in funding priorities after the end of the PHEIC.

Among the 354 estimates characterising ZIKV exposure through seroprevalence included in this review, there is a marked heterogeneity in geographic representation, with the Americas accounting for the majority of the published seroprevalence estimates. While these statistics reflect the attention that ZIKV received during the 2015–2016 epidemic in the Americas, they also underscore gaps in our understanding of the population-level immunity of populations outside these regions, which is critical to evaluate global outbreak risks now and in the future, and to design clinical trials for future vaccine and therapeutics. Seroprevalence estimates should also be interpreted with caution, as variations in assay sensitivity, specificity, and cross-reactivity—especially with other flaviviruses like dengue—can bias the results. Our analysis has demonstrated that ZIKV is moderately to highly transmissible (R_0_ typically 1.5–4, up to >7), with both vector-borne and sexual transmission contributing to infection. In combination with a pooled CZS probability of 4.65% and a pooled pregnancy loss probability of 2.48%, continued monitoring is essential to track outbreak potential and protect vulnerable populations. Moreover, our meta-analysis on the proportion of ZIKV-infected symptomatic cases suggests that up to half of ZIKV infections may go undetected in surveillance systems that rely on symptomatic reporting (51.20% (95%CI: 38.00%-64.23%, I^2^=96.7%)).

Estimates of CZS probability vary widely due to differences in study design (i.e. recruitment based on confirmed infection vs retrospective serology), population characteristics, case definitions, and methods used for the identification of ZIKV infections during pregnancy. Several studies from which we extracted CZS probability were not specifically designed to assess the CZS probability in recently infected mothers (i.e. studies tested for past exposure by IgG assays rather than recent infections through IgM detection). Our meta-analysis produced an overall estimate of 4.65% (95%CI: 3.38%-6.37%) which is slightly higher than other published estimates ^63–66^, though the confidence intervals overlap. We also found only one published review that conducted a trimester-specific meta-analysis of the probability of CZS^67^ including only 2 studies^68,69^. Our meta-analysis of CZS probability by trimester, based on 7 studies, yielded higher estimates than those published in Gallo et al.^67^, with consistent higher probability estimates in earlier trimesters.

Although we extracted 969 parameters from the literature, it was not possible to perform meta-analyses across most parameters (other than CZS, pregnancy loss probabilities and proportion of symptomatic cases) due to low numbers of comparable estimates, unavailability of sample size information and substantial heterogeneity in study contexts and methodologies. For example, only one estimate of the generation time and two estimates of the serial interval have been published. We also noted substantial variation in the terminology and methods used for estimation for studies reporting attack rates and reproduction number estimates, where basic and effective reproduction numbers were occasionally defined interchangeably, and in studies reporting genomic information, where mutation rates (per generation) and evolutionary or substitution rates (per year) were often conflated.

This study has limitations. We included only peer-reviewed publications in English, which may have led to the exclusion of relevant studies published in other languages and in grey literature or outbreak reports. Studies reporting outbreak dynamics or seroprevalence estimates which did not meet the formal inclusion criteria were excluded,^70–74^ hence, the outbreak data reported in this systematic review should not be considered a complete compendium, as several sources of ZIKV outbreaks exist outside peer-reviewed publications, specifically PAHO^75^ and national Ministries of Health. Moreover, while we attempted to extract disaggregated parameter estimates where available, for feasibility purposes, we prioritised summarising data over fully disaggregated information (e.g., by age or sex) (Appendix pp 6). All information is however included in our open-source database (also in Appendix: Tables B7-B16) that is accessible via the R package *epireview* to support future reuse. Finally, data extraction was conducted by a team of 16 reviewers using detailed, standardised guidelines^76^. Although double extractions were conducted at early stages, a residual risk of inconsistency and human error remains.

Despite these limitations, this study has several strengths as it summarises the information published in 418 studies on 984 epidemiological parameters, 154 models, and 127 outbreak records, which provides the most thorough quantitative overview of ZIKV epidemiology and disease risk available to date. As well as documenting key epidemiological parameters needed for outbreak response and preparedness planning, the richness of the data collated in this systematic review allows us to provide global, regional and country-level estimates of the probability of adverse effects of ZIKV, including CZS, pregnancy loss and the proportion of symptomatic cases.

In summary, this study presents a dynamic, open-source resource designed to support continuous updates by the research community to provide timely quantitative information on ZIKV epidemiology. By enabling the development of data-driven and evidence-based analytical, modelling and computational tools, this resource will aid mathematical modellers, public health officials and policymakers in outbreak prevention, preparedness, response and disease control planning.

## Contributions

SB, SvE, NI-E, and AC conceptualised this systematic review. KM, AV, CM, TR, SIL, HJTU, KC, ZMC, ESK and RM, searched the literature and screened the titles and abstracts. KM, AV, CM, TR, TMN, KF, A-MH, SIL, KC, ZMC, GC-D, ESK and RM reviewed all full-text publications. KM, AV, CM, TR, TMN, SB, DPD, PD, KF, A-MH, SIL, SR-P, RJS, HJTU, RM and ID extracted the data. KM and AV did formal analysis of, visualised, and validated the data, with supervision from RM and ID. KM, AV, TMN and SB were responsible for software infrastructure. AC acquired funding. AC, RM, and ID were responsible for project administration. PD, GC-D, RKN, DN, HJTU, and SvE were responsible for training individuals on and accessing Covidence and designing the Access system. AV and KM accessed and verified the data. KM, AV, RM, and ID wrote the original draft of the manuscript. All authors debated, discussed, edited, and approved the final version of the manuscript. All authors had full access to all the data in the study and had final responsibility for the decision to submit the manuscript for publication.

## Data Availability

Data and code are available from: https://github.com/mrc-ide/epireview/ and https://github.com/mrc-ide/priority-pathogens.

## Declarations of interest

PD and RM report payment from WHO for consulting on integrated modelling and MERS-CoV, respectively. HJTU reports payment from the Moderna Charitable Foundation (paid directly to institution for an unrelated project). AC received personal consulting fees from Munich Re for work unrelated to this project. All other authors declare no competing interests. The views expressed are those of the authors and not necessarily those of the National Institute for Health and Care Research (NIHR), UK Health Security Agency, or the Department of Health and Social Care.

## Role of the funding source

This study was funded by the Medical Research Council (MRC) Centre for Global Infectious Disease Analysis (MR/X020258/1) funded by the UK MRC and carried out in the frame of the Global Health EDCTP3 Joint Undertaking supported by the EU; and the National Institute for Health Research (NIHR) Health Protection Research Unit in Health Analytics & Modelling (NIHR207404), a partnership between UK Health Security Agency (UKHSA), London School of Hygiene & Tropical Medicine, and Imperial College of Science, Technology, & Medicine. The views expressed are those of the author(s) and not necessarily those of the NIHR, UKHSA, or the Department of Health and Social Care.

KM, AV, CM, TR, TMN, SB, SR-P acknowledge funding from data.org (Epiverse-TRACE). KM acknowledges research funding from the Imperial College President’s PhD Scholarship. ID and AV acknowledge funding from the Wellcome Trust (213494/Z/18/Z, 226072/Z/22/Z and 228185/Z/23/Z). CM acknowledges the Schmidt Sciences for research funding (grant code 6–22–63345). TR, TMN and PD acknowledge funding from Community Jameel. SB acknowledges funding from the Wellcome Trust (223120/Z/21/Z). A-MH acknowledges additional funding from Gavi, the Vaccine Alliance, the Bill and Melinda Gates Foundation (INV-034281), previously (OPP1157270/INV-009125), and with ID from the Wellcome Trust (226727/Z/22/Z). GC-D acknowledges funding from the Royal Society. MK and PL acknowledge funding from the Imperial Policy Forum. NRF acknowledges funding from Wellcome Trust (316633/Z/24/Z) and the United Nations Foundation (G-23279). AC was supported by the Academy of Medical Sciences Springboard scheme, funded by the Academy of Medical Sciences, the Wellcome Trust, the UK Department for Business, Energy, and Industrial Strategy, the British Heart Foundation, and Diabetes UK (SBF005\1044).

The funders of this study had no role in study design, data collection, data analysis, data interpretation, or writing of the report.

