## Appendix for "A systematic review of Zika virus disease: epidemiological parameters, mathematical models, and outbreaks"

**C PRISMA 2020 Checklists** **118**

**D Pathogen Epidemiology Review Group (PERG) Membership** **121**

### Overview

We provide further details of the methodology (section A) and additional results (section B), including tables of all of the information extracted as part of the review (section B.7). We report the PRISMA checklists in Section C. Section D lists all members of the Pathogen Epidemiology Review Group who contributed to this work.

### A Extended Methods

This systematic review and meta-analysis for Zika Virus Disease is part of a series of systematic reviews of nine priority pathogens on WHO's 2018 R&D Blueprint List [1] and was registered on PROSPERO (International Prospective Register of Systematic Reviews, CRD42023393345). Other systematic reviews in this series completed to date studied Marburg virus disease [2], Ebola virus disease [3], Lassa fever [4], and SARS-CoV-1[5].

The code to reproduce the results presented in this manuscript is available at `https://github.com/mrc-ide/priority-pathogens`.

#### A.1 Search strategy and screening

**Database search term:**

```
zika AND ((transmissi* OR epidemiolog*) OR (model* NOT imag*) OR (severity OR "case
    fatality ratio*" OR CFR OR "case fatality rate*" OR "mortality rate*" OR "attack
    rate*") OR ("infectious period*" OR "serial interval*" OR "incubation period*" OR
    "generation time*" OR "generation interval*" OR "latent period*" OR latency) OR
    (heterogeneit* OR superspread* OR "super spread*" OR super-spread* OR
    overdispersion OR overdispersed OR over-dispersion OR over-dispersed OR "over
    dispersion" OR "over dispersed") OR (infectivity OR infectiousness OR "growth
    rate*" OR "reproduction number*" OR "reproductive number*" OR R0 OR "reproduction
    ratio*" OR "reproductive rate*") OR ("pre-existing immunity" OR serological OR
    serology OR serosurvey*) OR (evolution* OR mutation* OR substitution*) OR
    (outbreak* OR cluster* OR epidemic*) OR (risk factor*") OR ("extrinsic incubation
    period" OR EIP))
```

This search term is analogous for all pathogens in the series of systematic reviews. For this review, "extrinsic incubation period OR EIP" was added to the end. The database search was conducted on 6 March 2019 and repeated on 31 October 2024. All articles were imported into *Covidence*, a systematic review management software for screening.

We included peer-reviewed original research articles in English if they reported ZIKV disease parameters (including seroprevalence, reproduction numbers, genomic information, severity, risk factors, Zika congenital syndrome/microcephaly probability, pregnancy loss probability, or delays), outbreaks, and/or transmission models. The inclusion and exclusion criteria are listed below A.1.

| Inclusion | Exclusion |
|---|---|
| Measures/estimates of human: Reproduction numbers (R, $R_0$, $R_t$, r, $R_e$), growth rate (r), doubling times, generation time, serial interval, incubation/latent period, case fatality ratio (CFR), attack rate, mutation rate (e.g. from phylogenetic study), overdispersion, risk factors (risk and the measure), microcephaly or pregnancy loss probability. | Non-English language publication |
| Mention of historical or any outbreak in humans: size, year, location, duration, spatial scale | Studies of co-infections. (local, regional, national, international). |
| Measures/estimates of animal: R, $R_0$, $R_t$, r, $R_e$, growth rate, mutation rate. | Animal studies that do not report R, $R_t$ etc. |
| Mathematical or statistical model of transmission. | Qualitative studies, e.g., KAP studies. |
| Measures of seroprevalence and negative seroprevalence in humans. | Pathogen not the primary focus of study. |
| Relative ratio of human-human vs animal introductions. | Duplicates. |
| Reviews that report inclusion criteria for reference checking. | Does not match any of the inclusion criteria. |
| For "small" pathogens[1], include case reports to potentially reconstruct serial interval distribution, etc. | In-vitro studies. |
| | Non-peer reviewed publications, conference proceedings, abstracts, posters, letters to the editor |

Table A.1: Inclusion and exclusion criteria for papers.

Two independent reviewers for both the abstract screening and full-text review stages screened the identified articles to assess eligibility and accordance with the inclusion/exclusion criteria (see list of the reviewers in the Methods section of main text). Any disagreements between reviewers were resolved by consensus between the two reviewers. We show the Cohen's Kappa statistic of the inter-rater reliability in A.1.

Per the original research criterion in A.1, all reviews were excluded. From the systematic reviews identified in our search (**n=66**), we extracted the citations from these reviews and cross-referenced them with those already screened in Covidence. Among the citations that had not been previously screened in Covidence, we filtered the list to include only those with the word "Zika" in the title (n=350), which we manually checked for duplicates, relevance, and publication type. In total, 103 new papers were added to Covidence to be screened, of which some were duplicates or not found. Of the 69 left, we included 6 studies from which we extracted data. This list of studies is provided in epireview.

We provide a csv of articles included in the study hosted in the epireview package. The below code can be used to generate this list from epireview.

```
require(tidyverse)

is_epireview_available <- require("epireview")

if(!is_epireview_available)
{
  is_remotes_available   <- require('remotes')

  if(!is_remotes_available)
    install.packages('remotes')

  remotes::install_github('mrc-ide/epireview')
}

zika     <- epireview::load_epidata('zika')
```

The epireview package has multiple vignettes available, listed below.

[1]The inclusion/exclusion criteria are reported as stated in the PROSPERO registration for the overall priority pathogen project. The threshold is determined on a case-by-case basis for each pathogen, and ZIKV is not considered a "small" pathogen.

**Title and abstract screening**

| Reviewer A | Reviewer B | n | Proportionate Agreement | Yes Probability | No Probability | Random Agreement Probability | Cohen's Kappa |
|---|---|---|---|---|---|---|---|
| P3 | P1 | 2547 | 98.23 | 1.07 | 80.38 | 81.45 | 90.48 |
| P14 | P10 | 1644 | 98.36 | 0.09 | 94.07 | 94.16 | 71.88 |
| P3 | P5 | 1586 | 93.88 | 5.31 | 59.22 | 64.53 | 82.76 |
| P2 | P3 | 1446 | 98.06 | 0.01 | 97.66 | 97.67 | 16.7 |
| P10 | P11 | 1272 | 91.59 | 0.9 | 81.48 | 82.39 | 52.24 |
| P14 | P11 | 1271 | 95.12 | 0.35 | 88.55 | 88.89 | 56.08 |
| P1 | P5 | 1213 | 83.76 | 5.25 | 59 | 64.25 | 54.56 |
| P2 | P1 | 422 | 89.57 | 2.44 | 71.16 | 73.6 | 60.5 |
| P2 | P5 | 167 | 78.44 | 5.72 | 57.82 | 63.54 | 40.87 |
| P13 | P10 | 156 | 94.87 | 0.43 | 86.33 | 86.76 | 61.27 |
| P15 | P5 | 95 | 80 | 57.91 | 5.27 | 63.18 | 45.68 |
| P2 | P8 | 12 | 100 | 0 | 100 | 100 | NA |
| P3 | P8 | 5 | 100 | 0 | 100 | 100 | NA |
| P1 | P8 | 5 | 100 | 0 | 100 | 100 | NA |
| P5 | P4 | 2 | 50 | 0 | 50 | 50 | 0 |

**Full text review**

| Reviewer A | Reviewer B | n | Proportionate Agreement | Yes Probability | No Probability | Random Agreement Probability | Cohen's Kappa |
|---|---|---|---|---|---|---|---|
| P3 | P1 | 430 | 93.26 | 21.66 | 28.4 | 50.06 | 86.5 |
| P2 | P1 | 221 | 77.38 | 28.74 | 20.15 | 48.89 | 55.73 |
| P10 | P11 | 209 | 72.73 | 34.89 | 16.71 | 51.6 | 43.66 |
| P3 | P5 | 143 | 82.52 | 15.03 | 37.41 | 52.44 | 63.24 |
| P2 | P3 | 116 | 97.41 | 13.41 | 40.13 | 53.54 | 94.43 |
| P3 | P4 | 81 | 74.07 | 5.76 | 57.61 | 63.37 | 29.21 |
| P9 | P1 | 31 | 74.19 | 29.97 | 20.29 | 50.26 | 48.12 |
| P12 | P1 | 22 | 81.82 | 24.79 | 24.79 | 49.59 | 63.93 |
| P1 | P8 | 20 | 90 | 8.75 | 48.75 | 57.5 | 76.47 |
| P1 | P4 | 16 | 100 | 0 | 100 | 100 | NA |
| P1 | P6 | 15 | 73.33 | 52 | 5.33 | 57.33 | 37.5 |
| P1 | P5 | 8 | 100 | 1.56 | 76.56 | 78.12 | 100 |
| P7 | P1 | 7 | 57.14 | 24.49 | 24.49 | 48.98 | 16 |
| P2 | P8 | 5 | 100 | 100 | 0 | 100 | NA |
| P12 | P5 | 5 | 100 | 0 | 100 | 100 | NA |
| P5 | P4 | 4 | 100 | 0 | 100 | 100 | NA |
| P2 | P4 | 2 | 100 | 0 | 100 | 100 | NA |
| P3 | P7 | 2 | 50 | 0 | 50 | 50 | 0 |
| P2 | P5 | 2 | 50 | 50 | 0 | 50 | 0 |
| P3 | P8 | 1 | 0 | 0 | 0 | 0 | 0 |
| P12 | P3 | 1 | 100 | 0 | 100 | 100 | NA |
| P12 | P7 | 1 | 100 | 0 | 100 | 100 | NA |
| P2 | P7 | 1 | 0 | 0 | 0 | 0 | 0 |

Figure A.1: Cohen's Kappa for screening and full text review. P1-P15 are members of PERG (see appendix D), and we have provided who participated at each stage in the methods section of the main text. $n$ is the number of studies considered by each Reviewer A/B pair. We note that two people screened every study at both the Abstract/title and full-text review stages and any conflicts were resolved by discussion and mutual agreement of Reviewer A and B.

Table A.2: Available Vignettes in the `epireview` Package

| Title | Description |
|---|---|
| Disease Overview | Vignette for Marburg Virus Disease, Lassa fever, SARS-CoV-1 and Zika with tables and figures from the respective papers. This will be updated as data are added to the database. |
| Field Options | Vignette listing the options for each outbreak, model, or parameter field and explaining how to access them using functions in the package. |
| Database Update | Vignette explaining the process of updating the database with new article, outbreak, model, or parameter data. |

#### A.2 Data extraction

Once studies were identified as eligible for data extraction, they moved to the data extraction stage. We used a custom Microsoft Access database (Version 2503) that collected information on article metadata (including first author, title, journal, publication year, volume) and a quality assessment score (see A.2.4 below). We also extracted information on models, outbreaks, and parameters. See the wiki on Github for more details.

To validate data extraction, 16% (e.g. >10% required in our protocol) of identified papers were randomly selected for double extraction by two independent extractors to ensure concordance in the team of 16 extractors (see Methods in main text). Any discordance was resolved between the pairs of extractors. All of the remaining studies were extracted by a single extractor, but data were further validated during cleaning and analysis.

##### A.2.1 Outbreaks

Because of the complexity of the definition of ZIKV outbreaks that may not be well-defined, outbreaks were extracted from studies only if defined as such (either as "outbreaks" or "epidemics") by the study authors. We note that many outbreaks are reported in non-peer-reviewed literature, such as reports or correspondences, which would have been excluded from this review due to the inclusion/exclusion criteria (Table A.1).

##### A.2.2 Models

Information about transmission models for ZIKV disease was extracted from eligible papers. We collected information including model type, whether it modelled a stochastic or deterministic process, transmission pathways and interventions modelled, and compartments (e.g. SEIR-SEI) of compartmental models. We also extracted information about model assumptions, whether they were fitted to data or theoretical, and code availability.

##### A.2.3 Parameters

We extracted parameter values and types of values (e.g. mean, median), uncertainty intervals and types (e.g. confidence or credible intervals), variability (e.g. ranges or IQRs around a central estimate), and ranges of central tendency values across multiple estimates obtained from some form of disaggregation of data (e.g. age-stratified estimates or estimates from different populations). Parameter contexts were also recorded (e.g. population sample type, population group, location, country, ages, survey dates, sample size, and trimester of pregnancy where available).

For any genomic information, we recorded the part of the genome studied and if any new sequences were available. We recorded information about the method used to estimate reproduction numbers. Additionally, for reproduction numbers, we investigated estimates that were outliers from the others; in doing this, we found two estimates of the basic reproduction number that were in fact effective reproduction numbers (estimates of R after control measures were implemented). These two estimates were relabelled as Effective Reproduction numbers in the database.

For case fatality ratios, CZS probability, seroprevalence, and pregnancy loss probability, we recorded the numerators and denominators if reported and whether estimates were adjusted, either for censoring, test sensitivity/specificity or in any other way, or not.

Risk factors are highly dependent on the context of the data used to estimate it, so we extracted only the outcome (e.g. ZIKV infection, microcephaly) and risk factor (e.g. age, ZIKV-infected mother, sex, occupation), the type of occupation if specified, and whether the estimate(s) were statistically significant and/or adjusted. We chose not to extract the values of the risk factor estimates (e.g. odds or risk ratios) or the direction of the estimate for simplicity, as estimates are dependent on the context of the study and the reference group used, which requires many further fields for extraction. Levels of statistical significance were taken as reported in the paper. We show an overview of reported risk factors for a range of outcomes that may be useful for parameterising or designing transmission models. For each study, we combined risk factors and outcomes together; for example, if both age and sex were significant, adjusted risk factors for ZIKV infection and death, we would have included age and sex in the risk factor column and infection and death in the risk factor outcome column, resulting in a single risk factor entry in the database. For each study, the maximum number of risk factors that could have been extracted was 4 - significant/adjusted, significant/unadjusted, non-significant/adjusted, non-significant/unadjusted.

We extracted multiple forms of the reproduction number. The overall reproduction number can be separated into multiple routes of transmission, where the vector-borne reproduction number represents the average number of new human infections from a single infectious human via vector-borne transmission, while the sexual reproduction number refers to the average number of new cases via sexual transmission. The overall reproduction number is the sum of the transmission-route-specific estimates, assuming independence in transmission route.

Many papers reported multiple parameter values for the same parameter (e.g. across multiple locations, time periods, assumptions, sample types, etc.). For these, we applied the 'rule of 3', where if more than 3 parameter values were reported across these disaggregated groups, we only captured the range of central estimates reported for this parameter. We applied the 'rule of 3' to all disaggregation groups except locations (e.g. if $R_0$ estimates were reported across 10 admin 1 units, we extracted each estimate separately, whereas if there were 6 estimates corresponding to different age groups, we would only extract the range of the central estimates).

##### A.2.4 Quality assessment

For each study, we provided a quality assessment (QA) score derived using a custom questionnaire (A.3, the wiki on Github). The overall QA score was calculated as the mean number of "Yes" answers, excluding any non-applicable questions. We show the distribution of QA scores and trends of QA scores by publication year (based on a fit of a local polynomial regression using `ggplot2::geom_smooth()` in R) in B.2.

#### A.3 Data analysis

##### A.3.1 Meta-Analysis

The meta-analyses for the probability of CZS and pregnancy loss among ZIKV-infected mothers (in Figure 5 in the main text) followed a standard approach and was conducted using the `meta` R package [6]. Estimates were included in the meta-analyses if they were assigned a QA score of ≥50%, had ≥10 pregnant women with confirmed ZIKV infection in the denominator, and used a study design that was deemed by the authors to not bias estimates by selecting for the outcome (pregnancy loss or CZS). We combined the 3 cluster-level estimates from Robbiani et al. [7] into a single overall estimate for the meta-analysis.

A mixed-effects model is a linear model $y_i = \beta_0 + \sum_{j=1} \beta_j x_{ij} + u_i + \epsilon_i$, where $y_i$ are the observed data, $x_{ij}$ are explanatory variables, $\beta_j$ are the fixed effects, $u_i$ are the random effects (centered around zero and independent across $i$), and $\epsilon_i$ are error terms. Meta-analysis is a special case of the above mixed-effects model with only an intercept term $\beta_0$ (a fixed-effects/common-effects model) or only

Table A.3: Quality assessment questionnaire: possible responses for each question listed were: Yes, No, and Non-applicable (NA).

| Theme | Question |
|---|---|
| Is the methodological/statistical approach suitable? (how the data are used) | 1. Clear and reproducible<br>2. Robust and appropriate for the aim [subjective criteria] |
| Are the assumptions appropriate? (input parameters/assumptions - what goes into the methodology) | 3. Clear and reproducible<br>4. Justified (published study or analysis of data)[objective criteria] |
| Are the data appropriate for the selected methodological approach? | 5. Clearly described and reproducible<br>6. Are issues in the data clearly discussed and acknowledged?<br>7. Are issues in the data accounted for in the chosen methodological approach? |

an intercept term $\beta_0$ and a random-effects term $u_i$ associated with that intercept (a random-effects model).

We used a generalised logistic mixed-effects model in which the individual CZS probability estimates were transformed using the logit-transformation $y_i = \log\left(\frac{CZS_i}{1-CZS_i}\right)$ to ensure that the distribution was approximately normal. A generalised linear mixed-effects model was then applied to the transformed values to estimate the pooled effect. A comprehensive overview of the methodology is provided by [8].

In Figure 5 of the main text, we perform a sub-group analysis of CZS probability, in which estimates are grouped by population sample type. The meta-analysis in Panel B reports the common and random effects for each sub-group to investigate any patterns between groups.

The risk of publication bias was assessed through funnel plots (see Figures B.29, B.30) for the meta-analysis of CZS probability and pregnancy loss probability.

The code to reproduce our results is available on our GitHub repository.

##### A.3.2 Doubling time calculation

We report doubling times derived from extracted growth rates per day. Doubling times in days were calculated as: $T_d = \frac{log(2)}{r}$ where $r$ is the daily growth rate.

##### A.3.3 Reporting Uncertainty

There was a wide variety of formats reported for uncertainty and ranges in the extracted studies. While most studies reported a central estimate for parameters of interest, some reported a range of central values, or we applied the 'rule of 3' as described in appendix A.2.3. When central estimates across more than 3 groups were reported, this is shown in our plots as thick shaded colour bars. When we refer to a range of central estimates in the main text, the maximum and minimum values of these are included.

We also extracted uncertainty values (e.g. standard deviation or variance) as well as paired uncertainty (e.g. confidence or credible intervals). Uncertainty intervals are represented as thin solid line error bars, and we did not plot standard deviation, variance, or standard error in our figures.

#### A.4 Extraction Fields

##### A.4.1 Model extraction fields

| Data field | Expected data type | Variable name | Notes |
|---|---|---|---|
| Article ID | integer | article_id | ID to connect to article form |
| Model data ID | integer | model_data_id | ID assigned by database |
| Model type | character | model_type | General type of model - from dropdown list |
| Stochastic or deterministic | character | stoch_deter | Stochastic or deterministic model as reported |
| Transmission route | character | transmission_route | Transmission route(s) modelled - from dropdown list |
| Assumptions | character | assumptions | General assumptions for the model - from dropdown list |
| Compartmental type | character | compartmental_type | Specific type of compartmental model - from dropdown list |
| Theoretical model | logical | theoretical_model | Tick box whether the model was fitted to data (TRUE) or just theoretical (NA) |
| Intervention type | character | interventions_type | Type of intervention(s) modelled - from dropdown list |
| Code available | logical | code_available | Tick box whether code for model was publicly available and reported in the paper |

Table A.4: Model form fields: refer to epireview package documentation for dropdown options.

##### A.4.2 Parameter extraction fields

| Data field | Expected type | Variable name | Notes |
|---|---|---|---|
| Article ID | integer | article_ID | ID to connect to article form |
| Parameter data ID | integer | parameter_data_ID | ID assigned by database |
| Parameter type | character | parameter_type | category of parameter - see dropdown list |
| Parameter value | numeric | parameter_value | central parameter value |
| Parameter exponent | integer | exponent | parameter value exponent (base 10) |
| Inverse parameter | logical | inverse_param | tick box to indicate that only inverse of parameter is reported (e.g., recovery rate from fitted model instead of infectious period) |
| Parameter unit | character | parameter_unit | units for parameter value, applies to central estimate and ranges/uncertainty intervals - see dropdown list |
| Parameter value type | character | parameter_value_type | type of central parameter value - see dropdown list |
| Parameter lower bound | numeric | parameter_lower_bound | lower bound of the parameter range if a range was reported or if data are disaggregated |
| Parameter upper bound | numeric | parameter_upper_bound | upper bound of the parameter range if a range was reported or if data are disaggregated |
| Statistical approach | character | statistical_approach | Parameter estimated or observed - see dropdown list |
| Parameter uncertainty - single type | character | parameter_uncertainty_singe_type | type of uncertainty for central parameter value if single value was reported - see dropdown list |
| Parameter uncertainty - single value | numeric | parameter_uncertainty_single_value | value for uncertainty for central parameter value if a single value was reported (e.g. value of std. dev.) |
| Parameter uncertainty paired type | character | parameter_uncertainty_type | type of uncertainty for central parameter value if paired values were reported - see dropdown list |
| Parameter uncertainty - lower value | numeric | parameter_uncertainty_lower_value | lower bound for uncertainty for central parameter value if paired values were reported |
| Parameter uncertainty - upper value | numeric | parameter_uncertainty_upper_value | upper bound for uncertainty for central parameter value if paired values were reported |
| Distribution type | logical | distribution_type | type of distribution for estimated parameter - see dropdown list |
| First distribution parameter type | logical | distribution_par1_type | type of value for first distribution parameter - see dropdown list |
| First distribution parameter value | logical | distribution_par1_value | value for first distribution parameter (e.g. shape or scale parameter for a gamma distribution) |
| First distribution parameter uncertainty | logical | distribution_par1_uncertainty | tick box for whether uncertainty is estimated for the first distribution parameter (true) or not (false) |
| Second distribution parameter type | logical | distribution_par2_type | type of value for second distribution parameter - see dropdown list |

| | | | |
|---|---|---|---|
| Second distribution parameter value | logical | distribution_par2_value | value for second distribution parameter (e.g. shape or scale parameter for a gamma distribution) |
| Second distribution parameter uncertainty | logical | distribution_par2_uncertainty | tick box for whether uncertainty is estimated for the second distribution parameter (true) or not (false) |
| Disaggregated data available | logical | method_disaggregated | tick box if disaggregated estimates are available (true) or not (false) |
| Parameter estimates disaggregated by | character | method_disaggregated_by | categories for disaggregation of parameter estimates |
| Only disaggregated data available | logical | method_disaggregated_only | tick box if only disaggregated estimates are available (true) or if a central estimate is also available (false) |
| Is parameter from supplement? | logical | method_from_supplement | tick box for whether parameter was extracted from supplement (true) or not (false) |
| Is parameter in a figure only? | logical | parameter_fromfigure | tick box to indicate that parameter is plotted in a figure but not reported numerically in the text/table |
| Study population country | character | population_country | country of the survey population - see dropdown list |
| Study population location | character | population_location | region/district/province/city of the survey population - see dropdown list |
| Start day of study | integer | population_study_start_day | study start day |
| Start month of study | character | population_study_start_month | study start month - see dropdown list |
| Start year of study | integer | population_study_start_year | study start year - see dropdown list |
| End day of study | integer | population_study_end_day | study end day |
| End month of study | character | population_study_end_month | study end month - see dropdown list |
| End year of study | integer | population_study_end_year | study end year - see dropdown list |
| Survey timing related to outbreak | character | method_moment_value | timing of the survey in relation to the outbreak, if specified in paper - see dropdown list |
| Study population sample size | integer | population_sample_size | sample size of the population used for parameter estimation |
| Study population minimum age (years) | numeric | population_age_min | minimum age of the survey population in years |
| Study population maximum age (years) | numeric | population_age_max | maximum age of the survey population in years |
| Sex of study population | character | population_sex | sex of survey population - see dropdown list |
| Population sample setting | character | population_sample_type | general setting of the survey - see dropdown list |
| Population group | character | population_group | specific group of the survey population - see dropdown list |
| genome site | character | genome_site | site of genome or gene studied |
| Trimester of pregnant woman | character | trimester | trimester of pregnant woman's ZIKV exposure - see dropdown list |
| Urban/Rural area | character | urban_rural | urban or rural setting of ZIKV circulation - see dropdown list |
| Data availability | character | data_availability | Statement about availability of the data presented in the paper - see dropdown list |
| Gene | logical | Gene | Name of the genomic sequence or gene under analysis |
| Genomic sequence available? | logical | genomic_sequence_available | tick box whether genomic sequence data are available (true) or not (false) |
| Reproduction number pathway | character | R_pathway | transmission pathway that reproduction number is based on for vector-borne diseases |
| Method to estimate R | character | method_R | method used for estimation of the reproduction number - see dropdown list |
| Other delay start point | character | other_delay_start | start point for delays not in the parameter type dropdown list (e.g., delay from ... to ...) |
| Other delay end point | character | other_delay_end | end point for delays not in the parameter type dropdown list (e.g., delay from ... to ...) |
| Numerator | integer | cfr_ifr_numerator | numerator of either cfr/ifr (deaths) or seroprevalence (number seropositive) estimates |
| Denominator | integer | cfr_ifr_denominator | denominator of either cfr/ifr (cases) or seroprevalence (number tested) estimates |
| Is the cfr/ifr estimate adjusted? | character | cfr_ifr_method | is the cfr/ifr estimate adjusted, unadjusted, or unspecified - see dropdown list |
| Is the seroprevalence estimate adjusted? | character | serop_method | is the seroprevalence estimate adjusted (accounting for assay's sensitivity and specificity) - see dropdown list |
| Outcome for risk factor(s) | character | riskfactor_outcome | outcome for risk factor(s) - see dropdown list |
| Risk factor name | character | riskfactor_name | risk factor name - see dropdown list |
| Risk factor occupation | character | riskfactor_occupation | if risk factor is an occupation, then specified occupation as risk factor - see dropdown list |

| | | | |
|---|---|---|---|
| Risk factor adjusted | character | riskfactor_adjusted | adjustment status of risk factor(s) - see dropdown list |
| Risk factor significant | character | riskfactor_significant | statistical significance of risk factor(s) - see dropdown list |

Table A.5: Parameters form fields: refer to epireview package documentation for dropdown options.

##### A.4.3 Outbreak extraction fields

| Data field | Expected type | Variable name | Notes |
|---|---|---|---|
| Article ID | integer | article_id | ID to connect to article form |
| Outbreak data ID | integer | outbreak_data_id | ID assigned by database |
| Pre-outbreak baseline | Logical | pre_outbreak_baseline | Dropdown list for whether a baseline period was specified before the outbreak |
| Outbreak start | Date (day, month, year) | outbreak_start_day, outbreak_start_month, outbreak_start_year | Start date of the outbreak |
| Outbreak end | Date (day, month, year) | outbreak_end_day, outbreak_end_month, outbreak_end_year | End date of the outbreak |
| Outbreak ongoing | Logical | outbreak_ongoing | Tick if the outbreak was ongoing at time of reporting |
| Duration (months) | Numeric | outbreak_duration_months | Duration of the outbreak in months |
| Outbreak country | Character | outbreak_country | Country where the outbreak occurred (dropdown list) |
| Outbreak location | Character | outbreak_location | Subnational region or city of the outbreak |
| Number of cases confirmed | Integer | cases_confirmed | Total confirmed cases |
| Mode detection | Character | mode_detection | How a case was detected (dropdown list) |
| Number of cases suspected | Integer | cases_suspected | Total suspected cases |
| Number of unspecified cases | Integer | cases_unspecified | Number of cases without classification |
| Number of asymptomatic cases | Integer | cases_asymptomatic | Number of reported asymptomatic infections |
| Asymptomatic transmission described | Logical | asymptomatic_transmission | Tick if asymptomatic transmission was described |
| Number of severe cases | Integer | cases_severe | Total number of severe cases |
| Number of deaths | Integer | cases_deaths | Total number of deaths due to the outbreak |
| Population size of geographical area | Integer | population_size | Total population of the affected area |
| Type of cases disaggregated by sex | Character | sex_disaggregation_type | Type of sex disaggregation (dropdown list) |
| Male cases | Integer | male_cases | Number of male cases |
| Proportion of cases in males | Numeric | male_proportion | Proportion of cases that are male |
| Female cases | Integer | female_cases | Number of female cases |
| Proportion of cases in females | Numeric | female_proportion | Proportion of cases that are female |

Table A.6: Outbreak form fields: refer to epireview package documentation for dropdown options.

### B Supplementary results

#### B.1 Quality assessment results

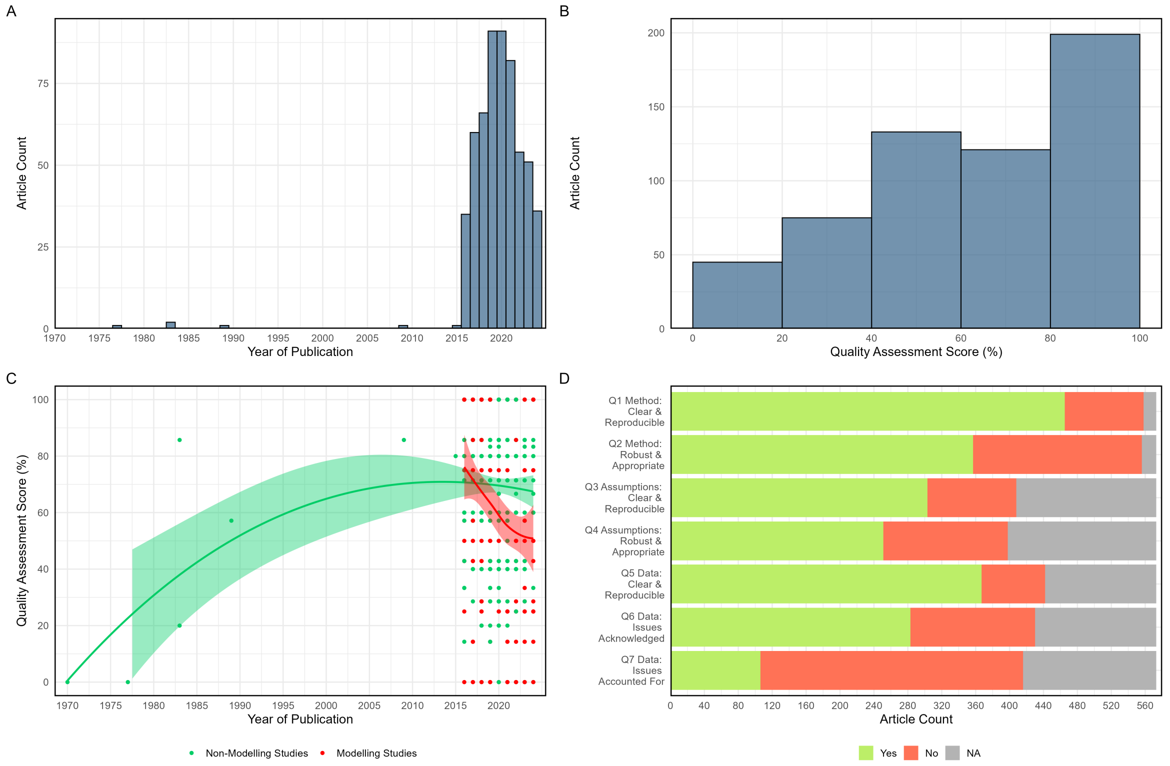

Figure B.2: (A) Article count by year of publication. (B) Article count by quality assessment score (defined as the proportion of "Yes" answers relative to the sum of "Yes" and "No" answers for each paper, excluding non-applicable questions). (C) Quality assessment score by year of publication (time trends for articles with and without transmission models fitted via local polynomial regression). (D) Article count for each quality assessment question scoring 'Yes', 'No' or 'Non-Applicable'.

#### B.2 Summary of extracted models and parameters

Figures B.3 and B.4 present an overview of the types of parameters and models extracted in the review, respectively.

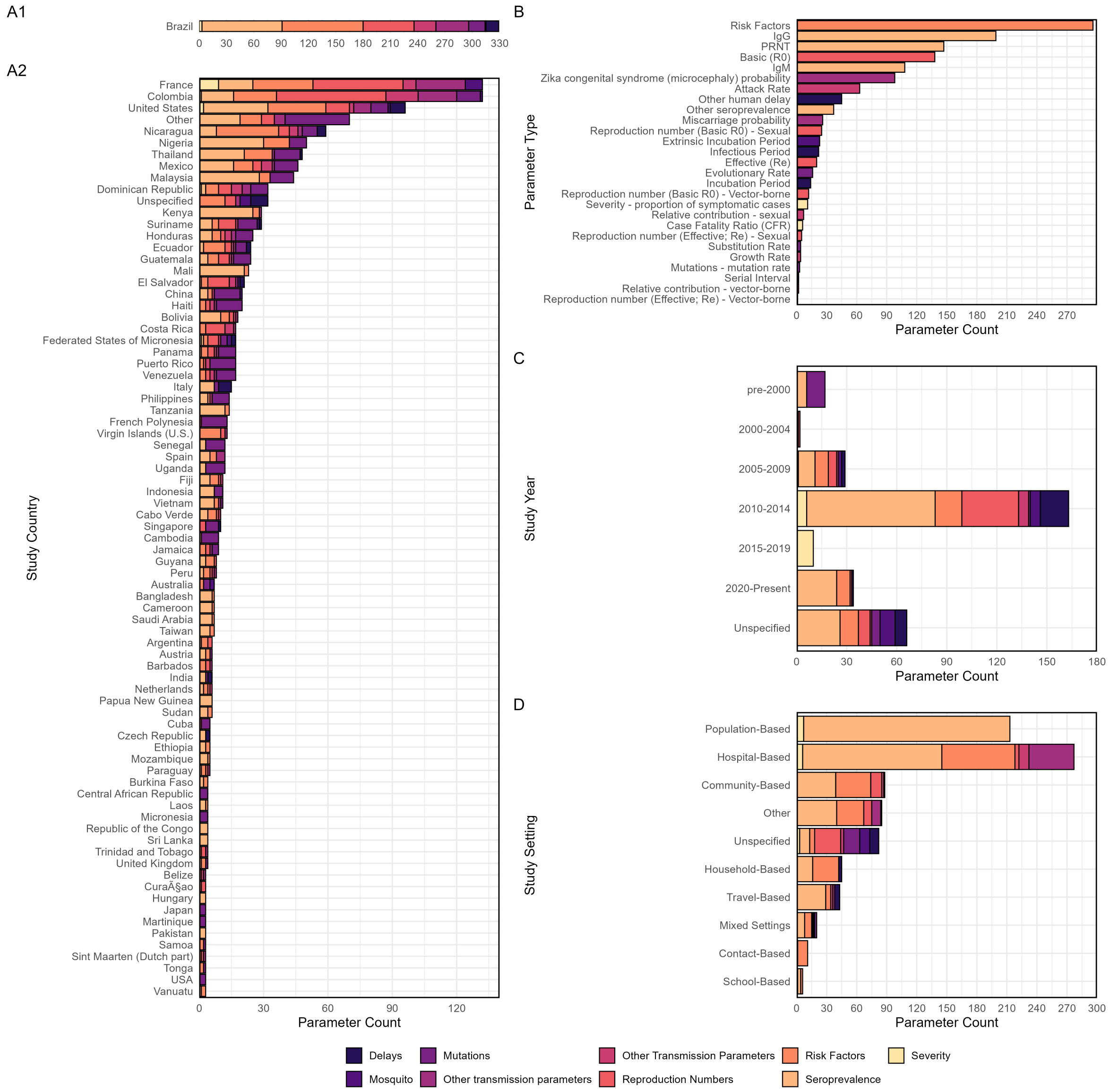

Figure B.3: Summary of extracted parameters by country, parameter type, study year, and study setting. Brazil is separate because of the large number of parameters compared to the other countries. Countries with less than three estimates are grouped into the "Other" label in Panel A2.

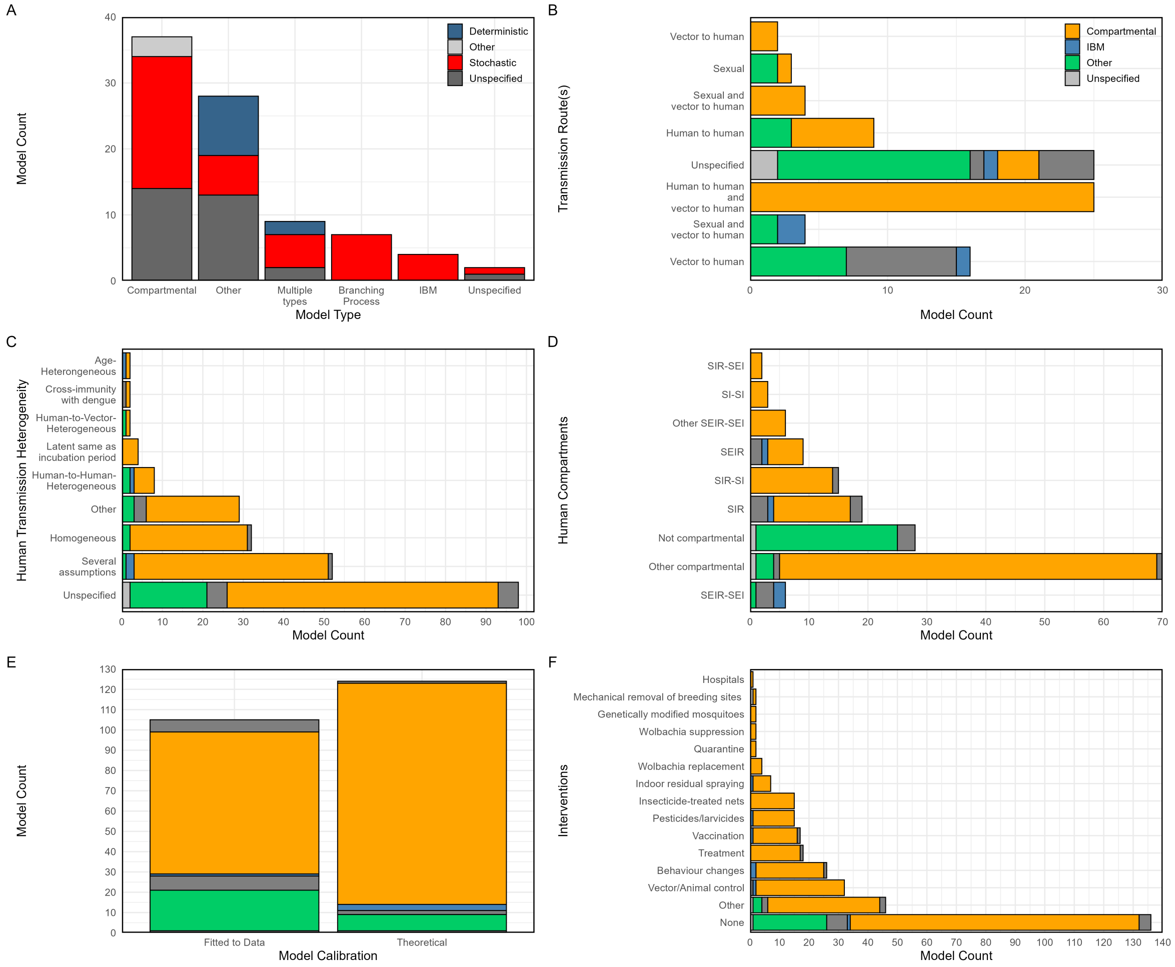

Figure B.4: Summary of extracted models by model type, transmission route, assumptions, compartments, calibration type, and interventions.

#### B.3 Additional plots of parameters

Unless otherwise specified, data in all plots within this section have been filtered to studies with a QA score of at least 50%.

##### B.3.1 Basic reproduction numbers

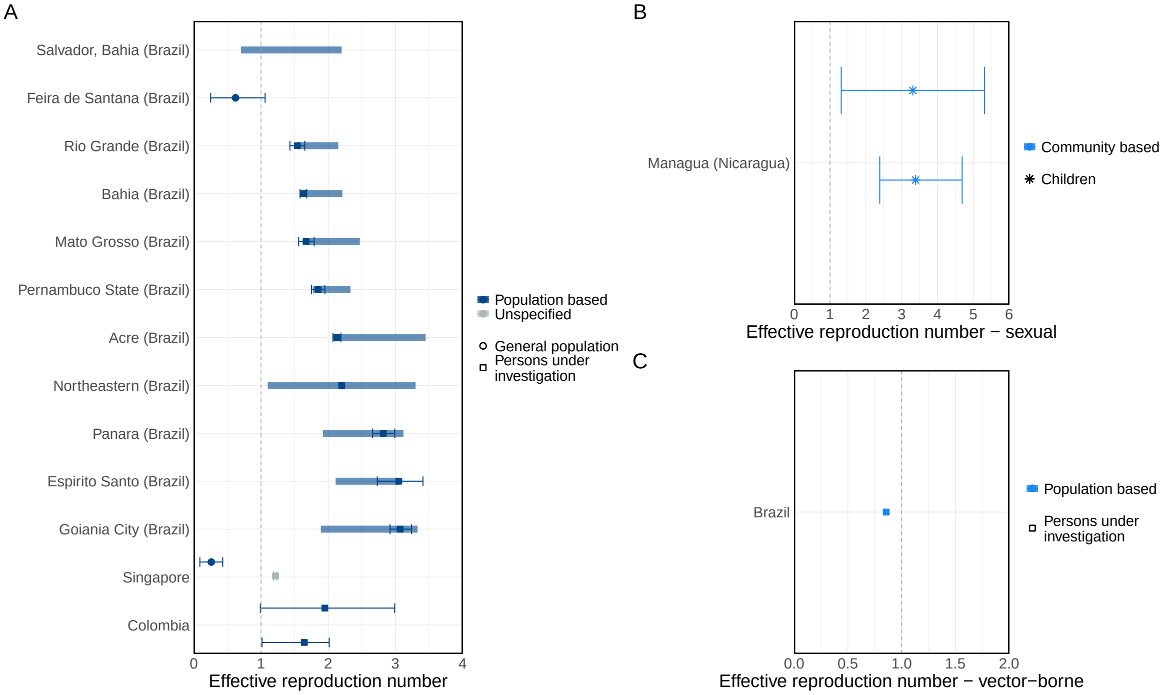

Figure B.5: Basic reproduction numbers, sexual transmission (A), and vector-borne (B). Note: the plot of overall basic reproduction numbers from studies with high QA scores is Figure 3 in the main text.

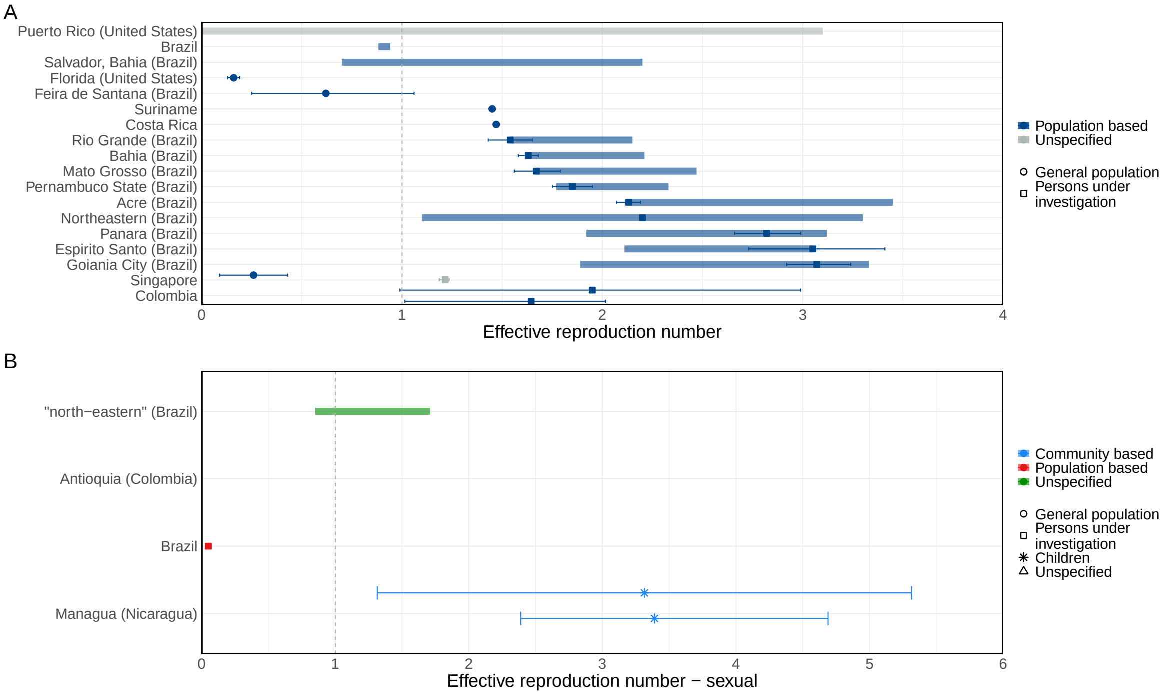

Figure B.6: Basic reproduction numbers from all studies, sexual transmission (A) and vector-borne (B). This figure includes all studies. Note: the plot of overall basic reproduction numbers from all studies (regardless of QA score) is Figure figure B.26.

##### B.3.2 Effective reproduction numbers

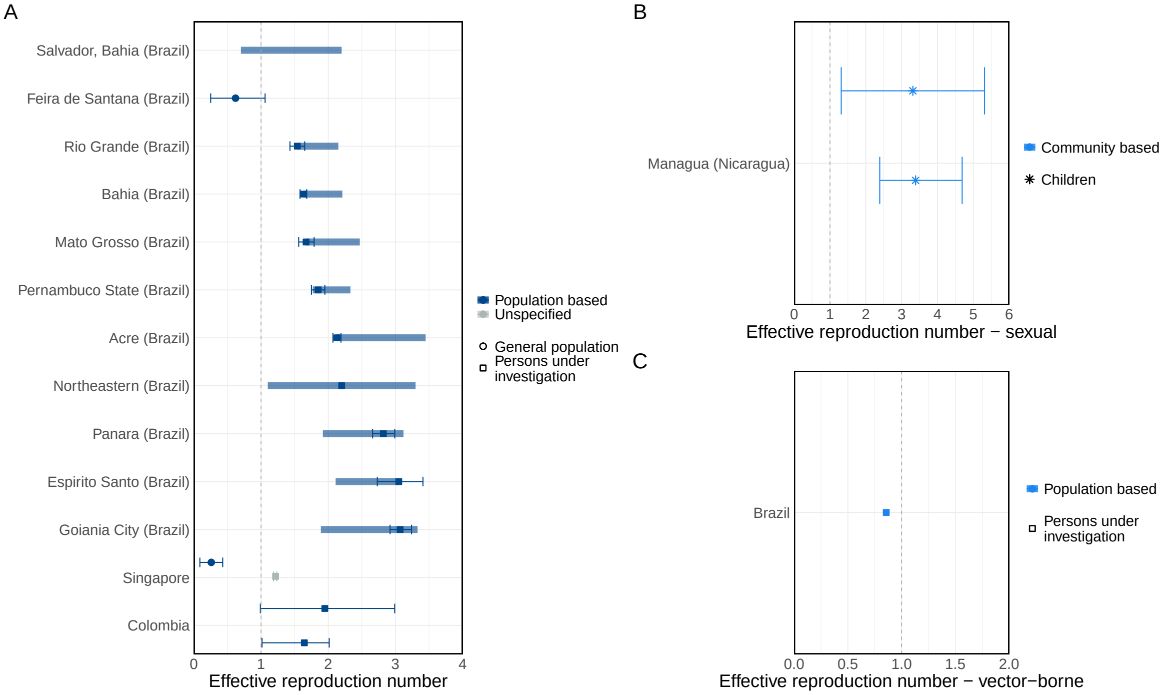

Figure B.7: Effective reproduction numbers, overall (A), sexual (B), and vector-borne (C).

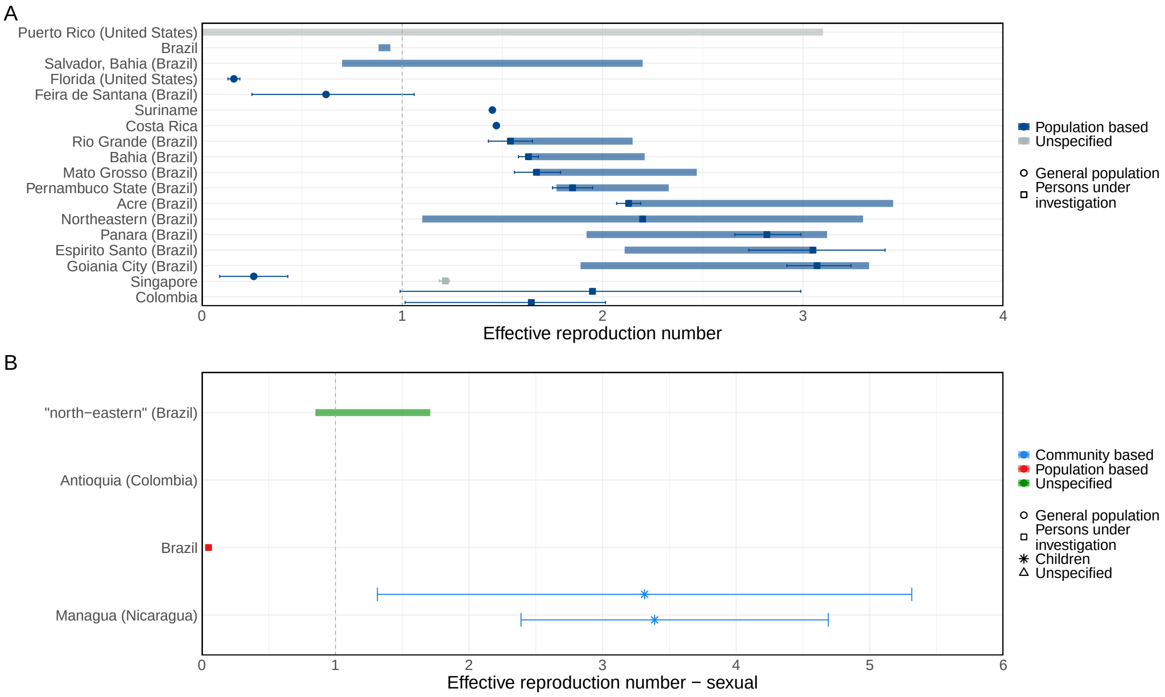

Figure B.8: Effective reproduction numbers from all studies, overall (A), sexual (B), and vector-borne (C). This figure includes all studies.

##### B.3.3 Seroprevalence

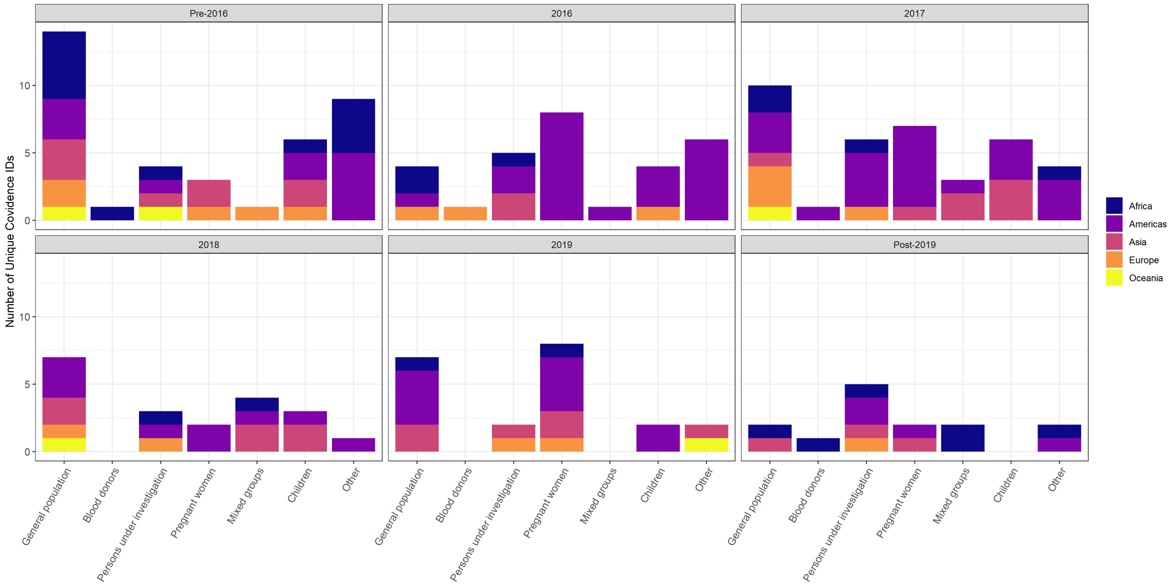

Figure B.9: Number of seroprevalence-related publications by population group. Number of unique studies in each population group, coloured by continent of the sero-survey.

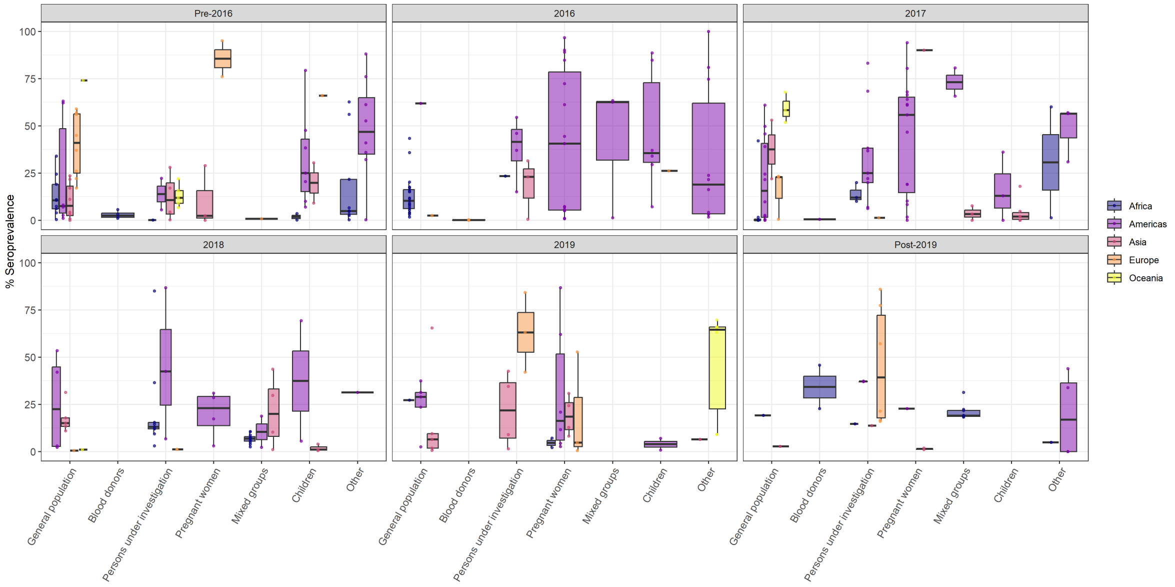

Figure B.10: Seroprevalence estimate by population group. Distribution of seroprevalence estimates in each population group, coloured by continent of the sero-survey.

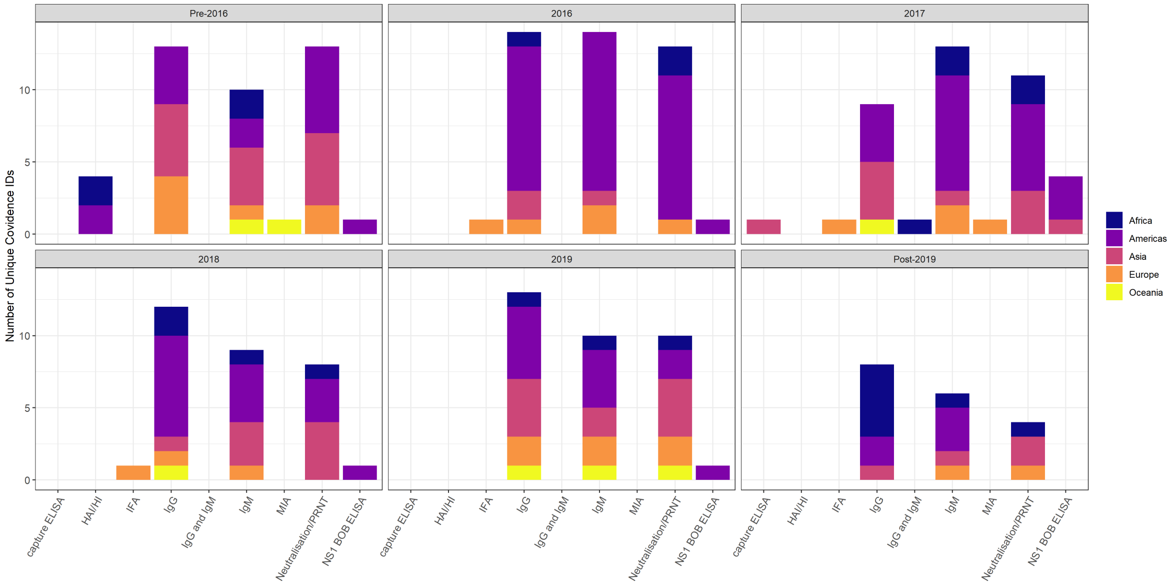

Figure B.11: Seroprevalence by assay. Number of unique studies for each type of assay, coloured by continent of the sero-survey.

##### B.3.4 Genomic parameters

We extracted 15 evolutionary rates and 3 substitution rates from 15 studies, of which 3 were for Brazil, 1 for the United States, while the majority came from multi-country analyses including countries in Europe, the Americas, Asia, Africa and Oceania (n=14). Figures B.12 and B.13 and Table B.13 summarise these estimates across different sampling intervals (in years). The evolutionary rate estimates tended to decrease as the variance in sampling dates increased, but most values clustered around $1 \times 10^{-3}$ substitutions per site per year, with substitution rates showing higher uncertainty at longer sampling intervals.

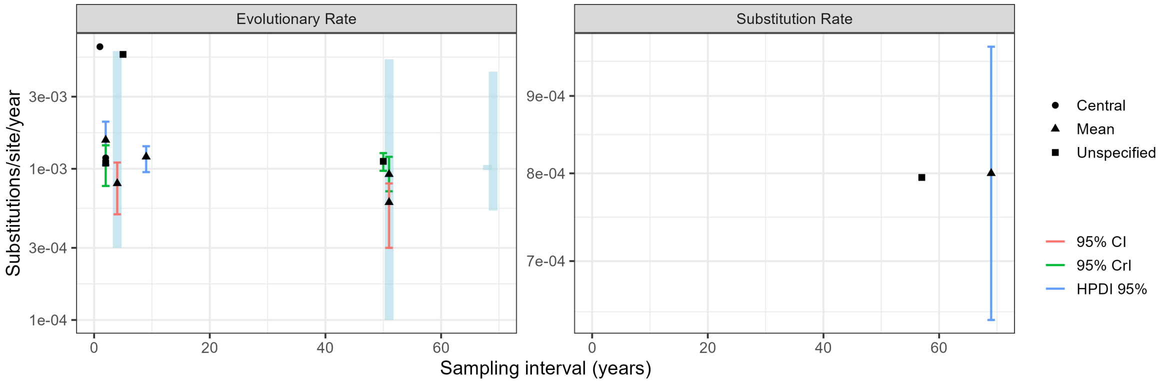

Figure B.12: Genomic information reported in studies with high QA scores. Points are central estimates, solid lines are confidence, credible, or high posterior density intervals, and shaded segments are ranges of central estimates across multiple groups.

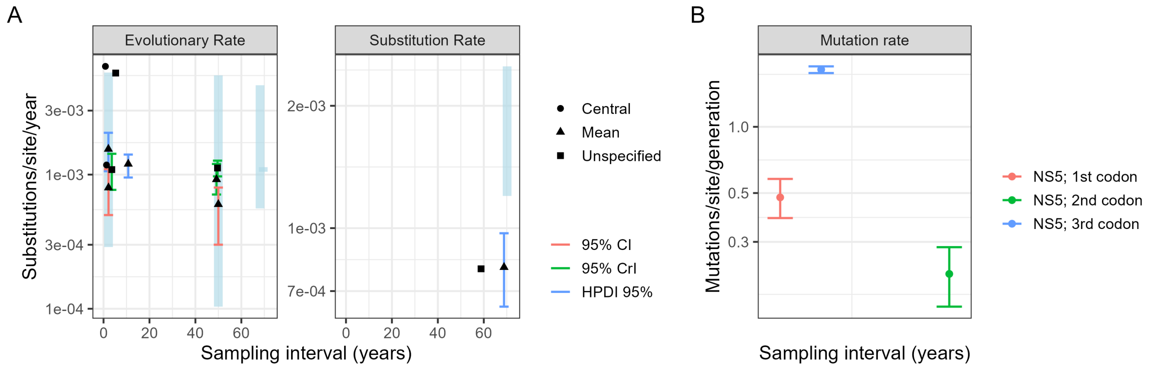

Figure B.13: Genomic information from all included studies. Points are central estimates, solid lines are confidence, credible, or high posterior density intervals, and shaded segments are ranges of central estimates across multiple groups.

##### B.3.5 Pregnancy loss probability

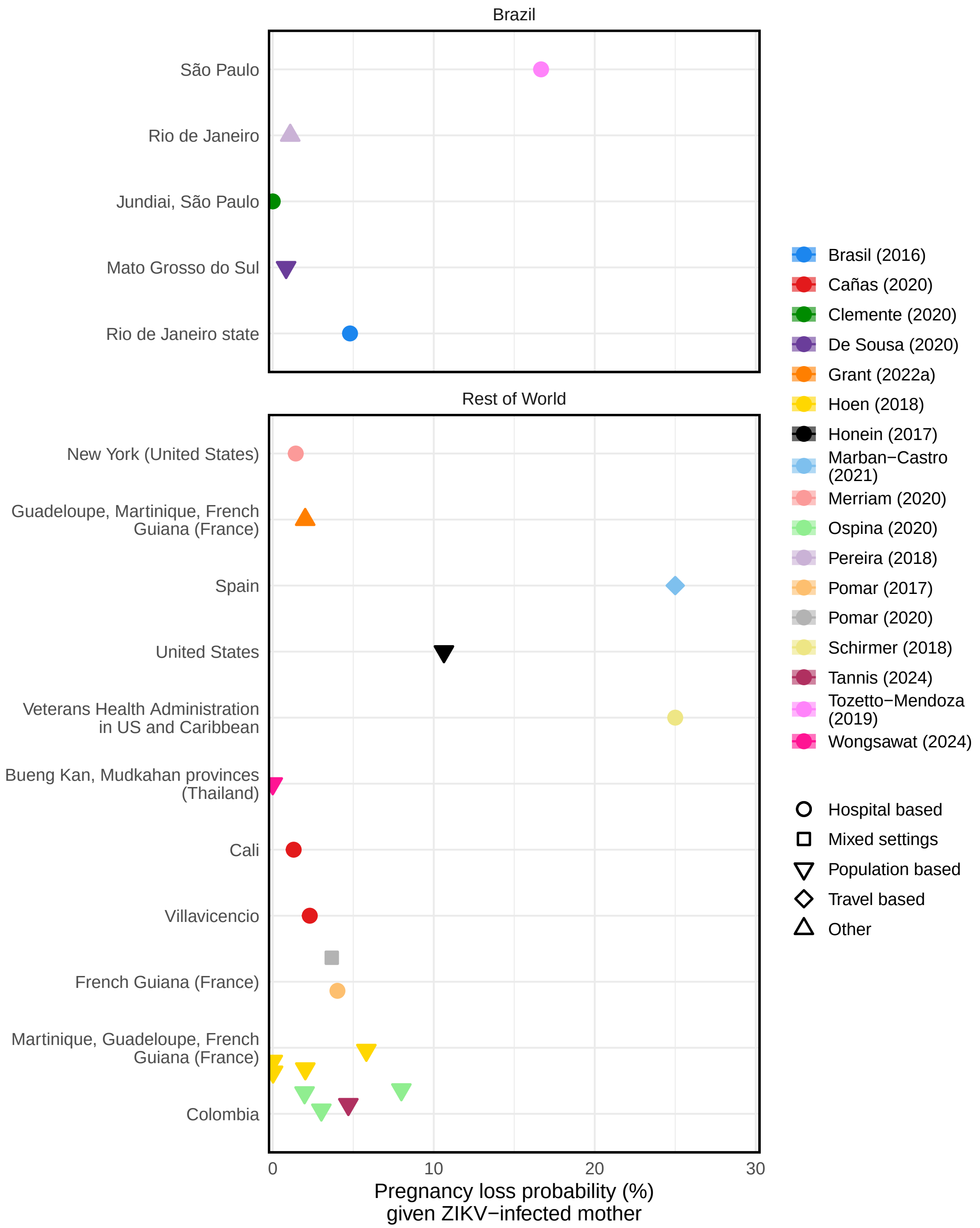

Figure B.14: Estimates of reported pregnancy loss probabilities. The top panel shows estimates from Brazil, while the bottom panel shows estimates from the rest of the world. Points are central estimates, solid lines are confidence or credible intervals, and shaded segments are ranges of central estimates across multiple groups.

##### B.3.6 Severity

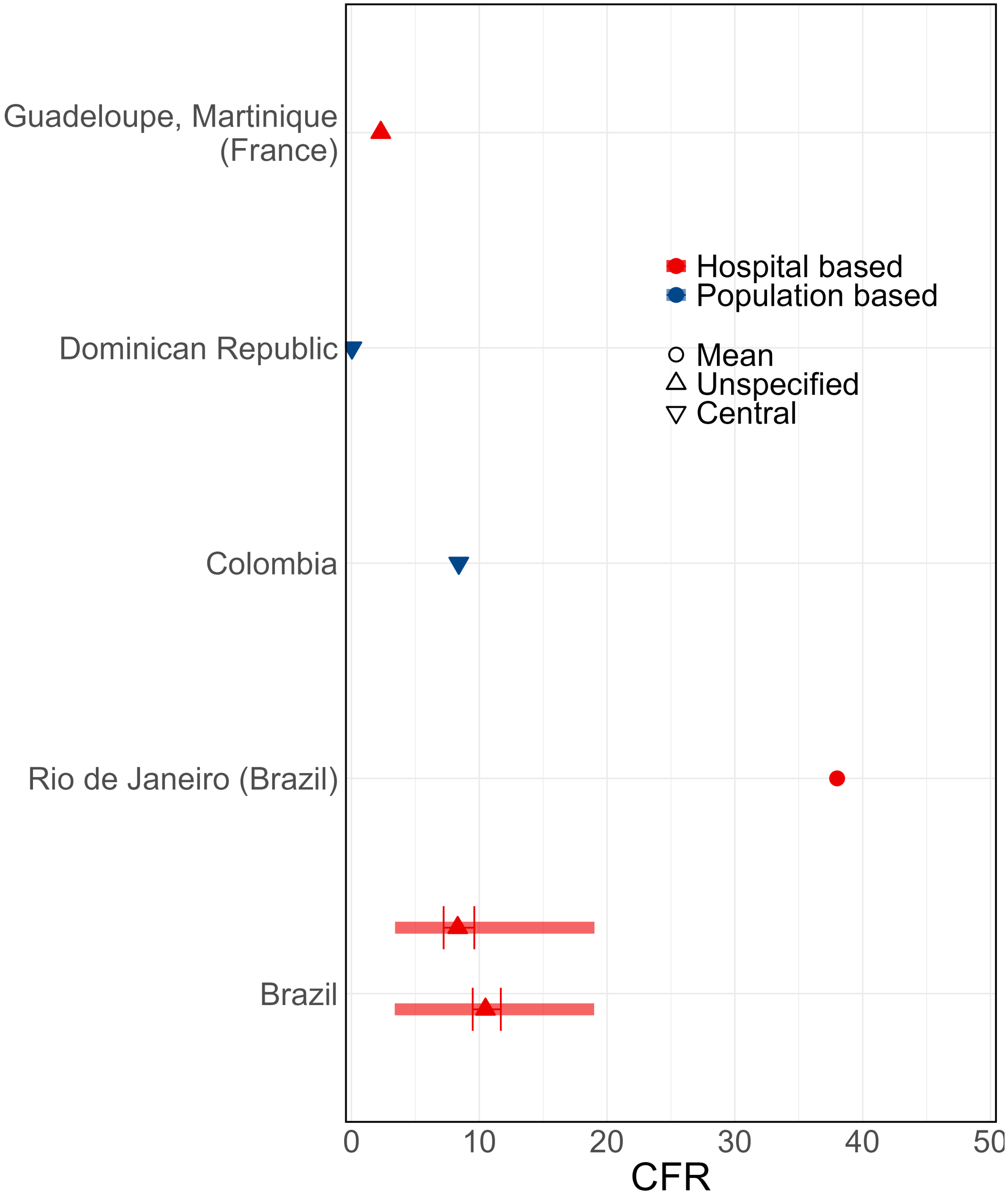

Figure B.15: Case fatality ratio estimates from all included studies, which all had high QA scores. Points are central estimates, solid lines are confidence or credible intervals, and shaded segments are ranges of central estimates across multiple groups.

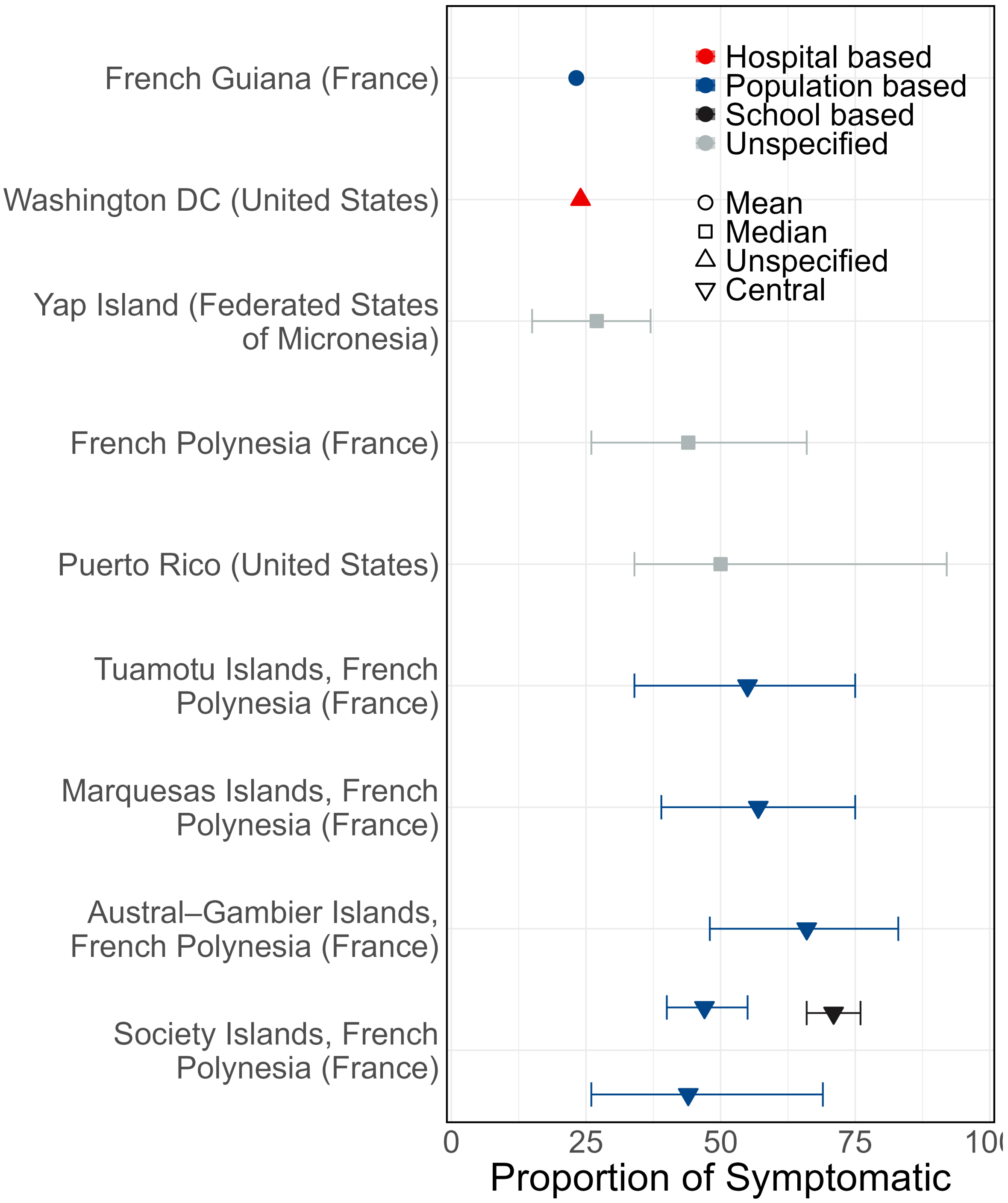

Figure B.16: Proportion of cases that are symptomatic from all included studies. Points are central estimates, solid lines are confidence or credible intervals, and shaded segments are ranges of central estimates across multiple groups.

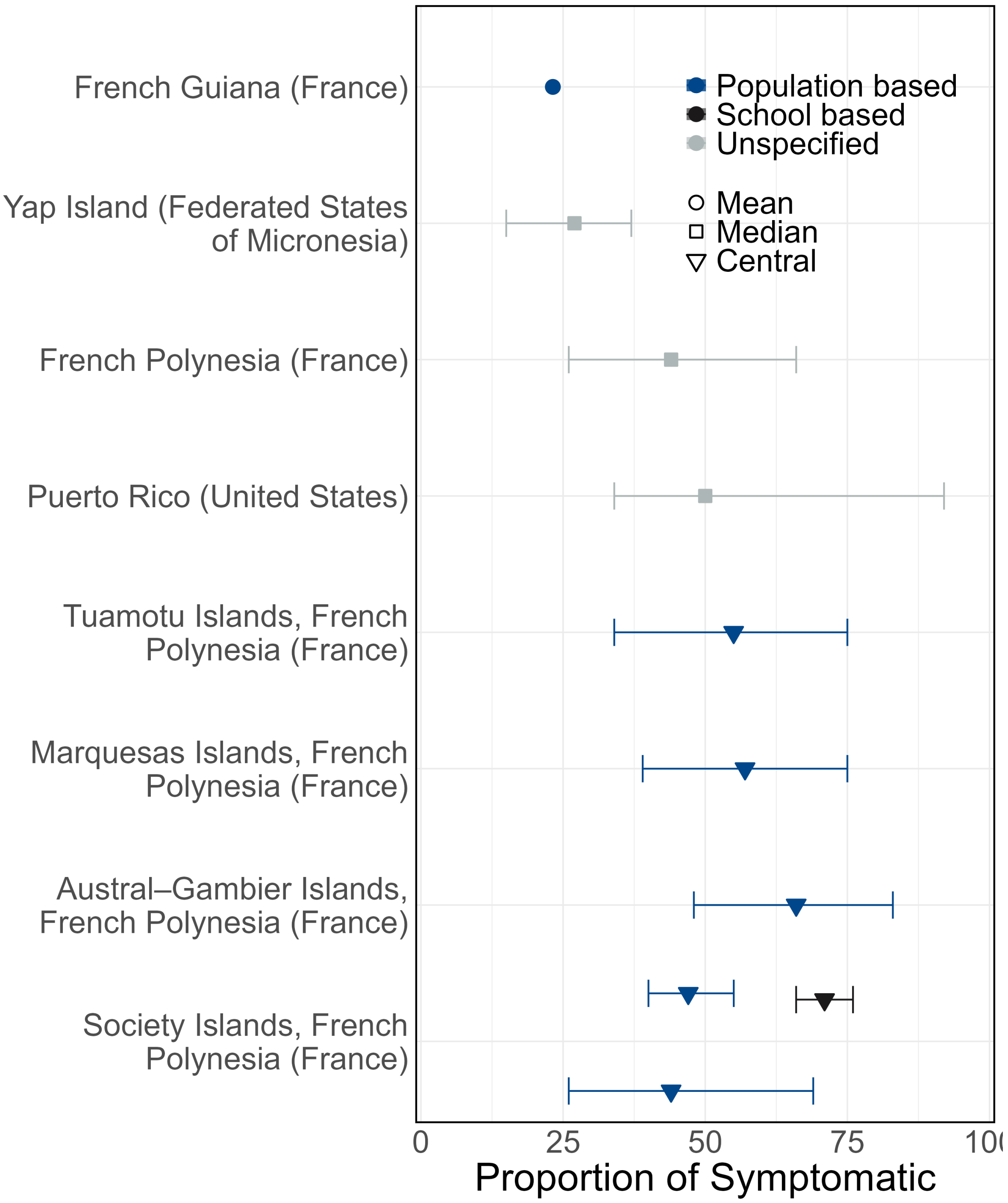

Figure B.17: Proportion of cases that are symptomatic from studies with high QA scores. Points are central estimates, solid lines are confidence or credible intervals, and shaded segments are ranges of central estimates across multiple groups.

##### B.3.7 Other transmission parameters

We considered estimates of the attack rate or final size of the epidemic. Of the 18 studies reporting estimates of the attack rate (Figures B.18 and B.19, Table B.12), we extracted a total of 51 estimates reported from Micronesia, Philippines, French Polynesia, Cabo Verde and several countries of Central and South America. The central estimates of the attack rate ranged from 0% in the Federal District of Mexico in 2015-2018 [9] to 94% estimated by a modelling study in French Polynesia in 2013-2014, [10] underscoring the variability in ZIKV transmission and burden across geographic regions.

In addition, we identified estimates of the growth rate (Figure B.20) and of the contribution of sexual transmission to overall ZIKV spread (Figures B.21 and B.22, Table B.12). Two studies in Colombia quantified the proportion of transmission attributable to sexual transmission, reporting estimates of 15.36% (95%CI 12.83-17.14)50 and 23% (95%CI 1-47%) [11]. We characterised growth rate estimates (n=4) (Appendix: Figure B.20, Table B.12) from 3 studies conducted in Brazil, Colombia, and Nicaragua, with corresponding doubling times spanning from 0.85 (95%CI 0.12-0.85) days in a paediatric cohort in Rio de Janeiro, Brazil [12] to 8.7 (95%CI 8.0-10.5) days in a hospital setting in Barranquilla, Colombia [13].

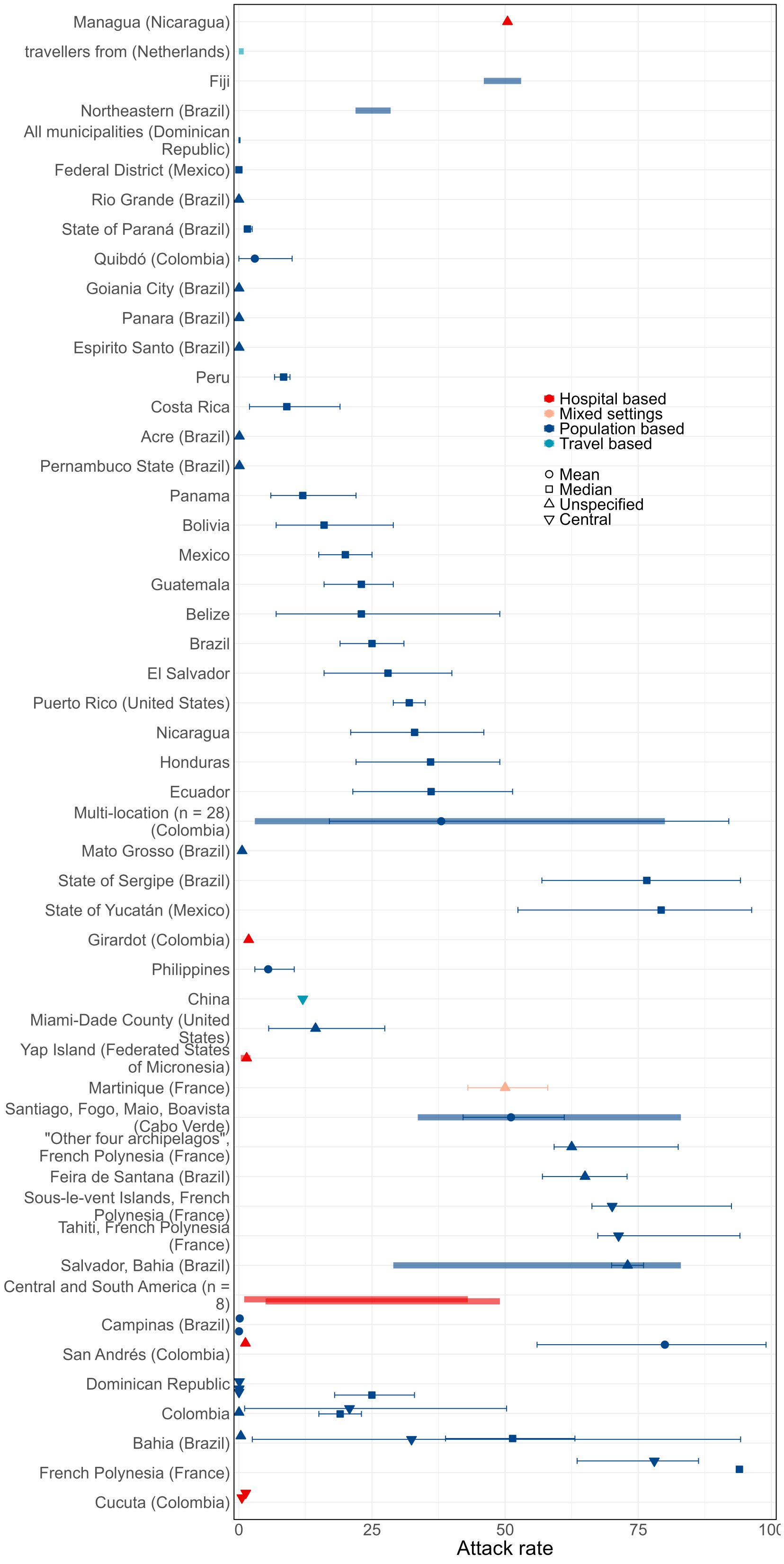

Figure B.18: Attack rate from all included studies. Points are central estimates, solid lines are confidence or credible intervals, and shaded segments are ranges of central estimates across multiple groups.

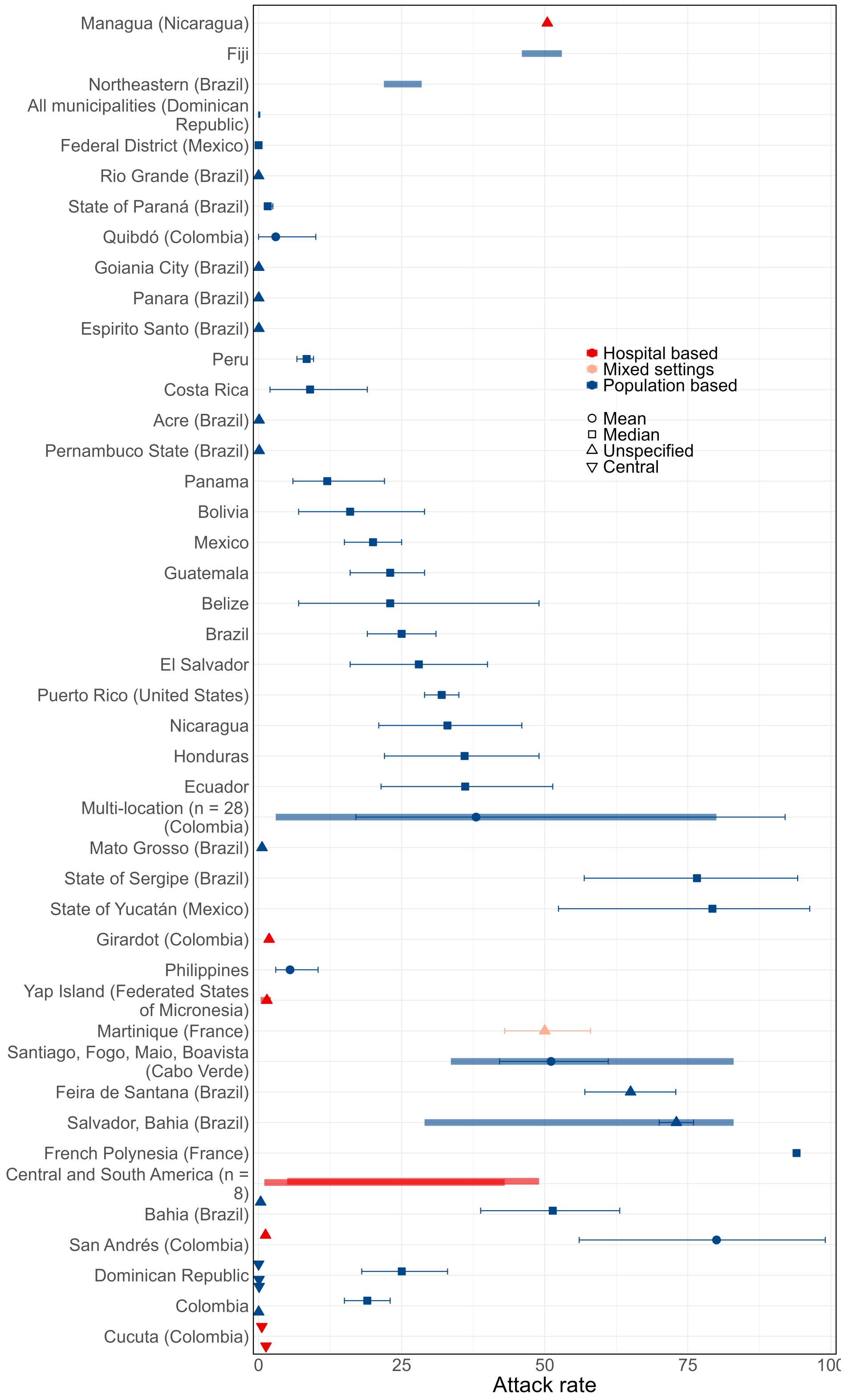

Figure B.19: Attack rate from studies with high QA scores. Points are central estimates, solid lines are confidence or credible intervals, and shaded segments are ranges of central estimates across multiple groups.

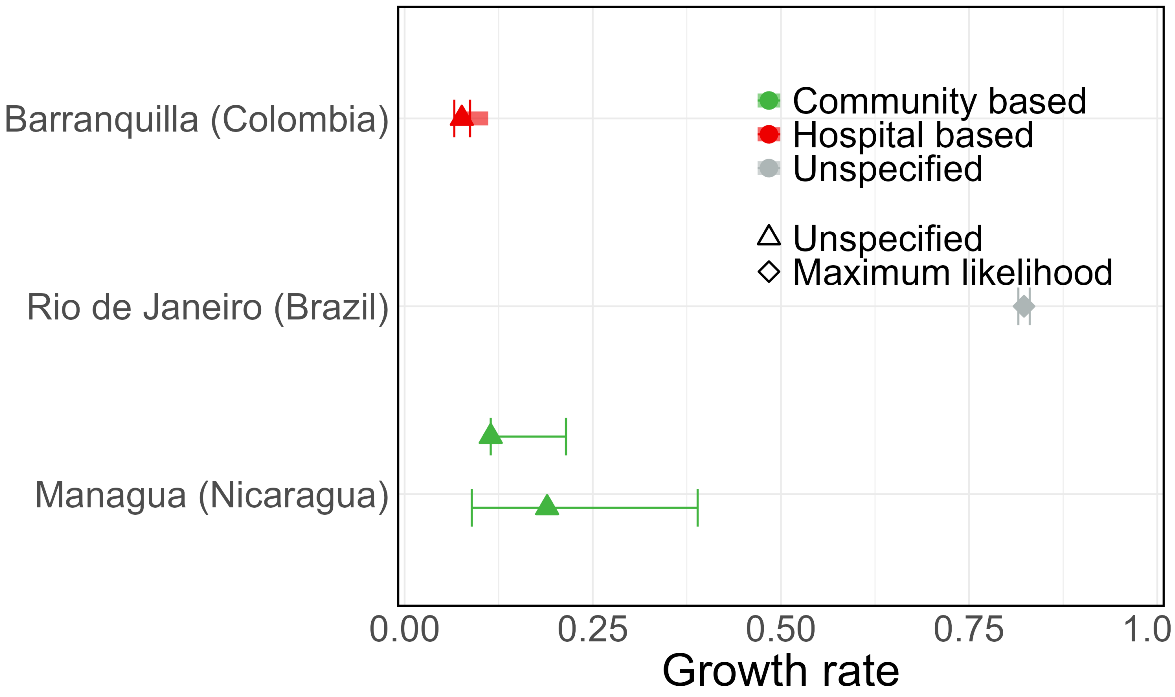

Figure B.20: Growth rate from all included studies, which all had high QA scores. Points are central estimates, solid lines are confidence or credible intervals, and shaded segments are ranges of central estimates across multiple groups.

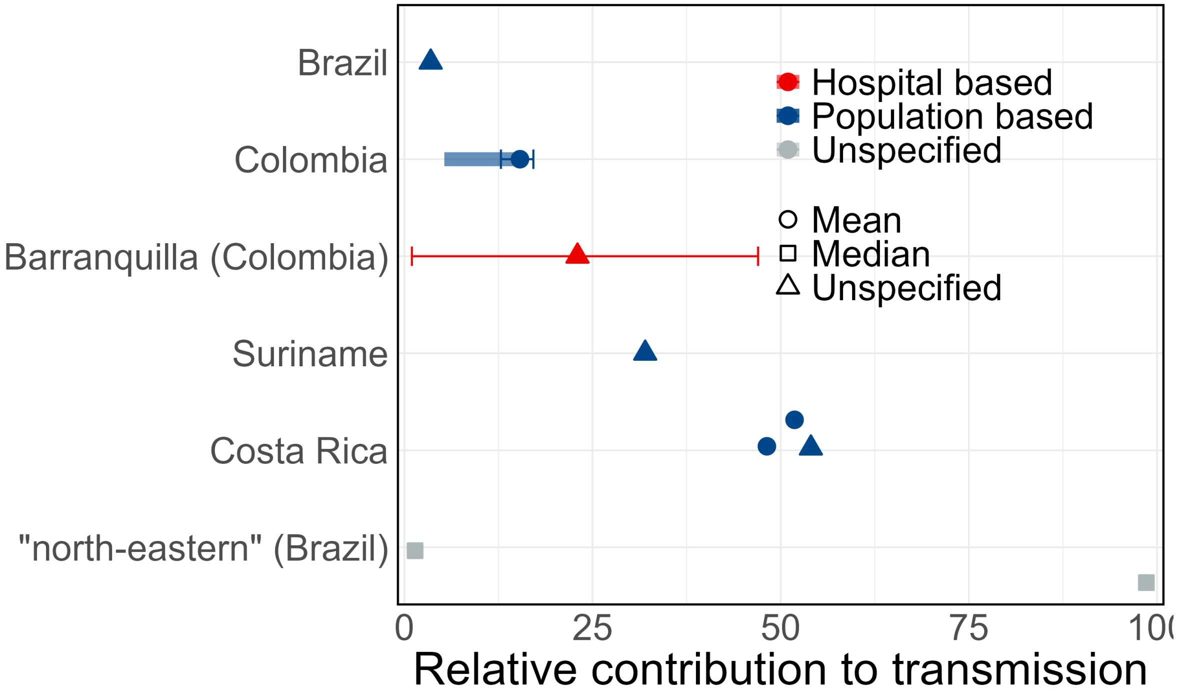

Figure B.21: Sexual relative contribution to transmission from all included studies. Points are central estimates, solid lines are confidence or credible intervals, and shaded segments are ranges of central estimates across multiple groups.

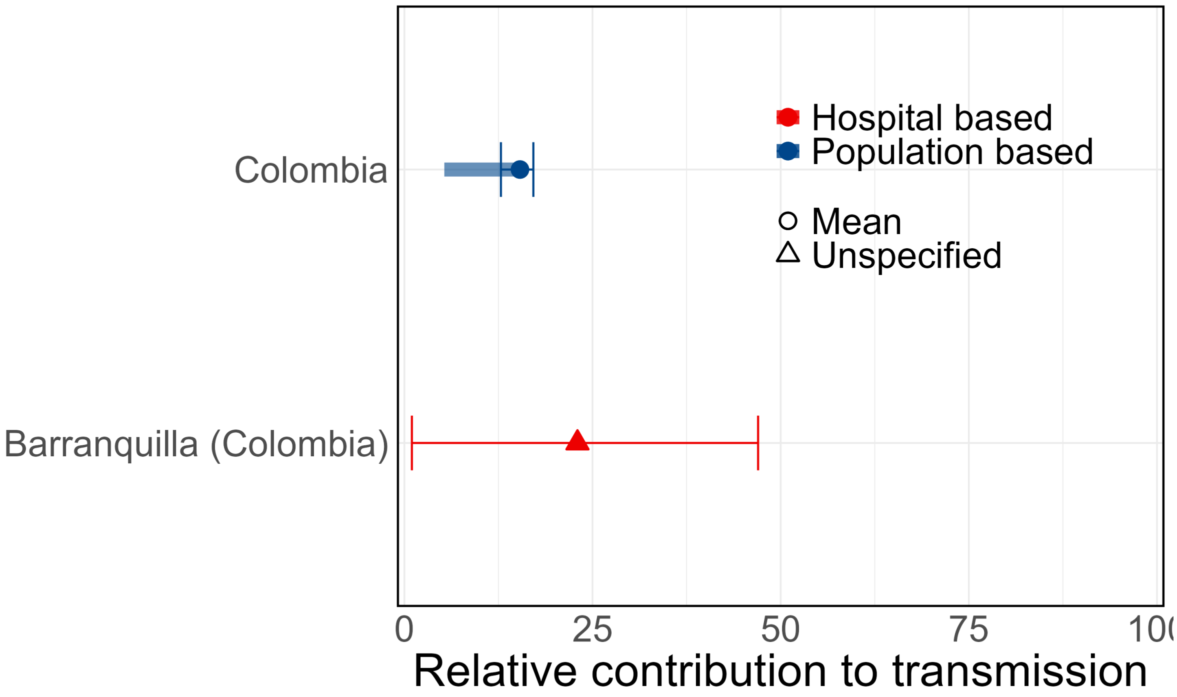

Figure B.22: Sexual relative contribution to transmission from studies with high QA scores. Points are central estimates, solid lines are confidence or credible intervals, and shaded segments are ranges of central estimates across multiple groups.

##### B.3.8 Risk factors

Ten unique risk factor types with 14 different Zika-related outcomes were reported in 137 studies (n=486 entries of 172 unique combinations of risk factor, outcome, statistical significance, and adjustment), of which 415 entries of 157 unique combinations were from 111 studies with quality assessments 50% or higher (Figures B.23 and B.24). ZIKV-related risk factors among the studies wiht high QA scores were most frequently investigated against serological outcomes (n=80) and ZIKV infection (n=65). The most commonly reported risk factors across all outcomes were age (n=87), sex (n=62), and infection during pregnancy (n=40). Most risk factors were reported as both significant and non-significant in different studies (B.11).

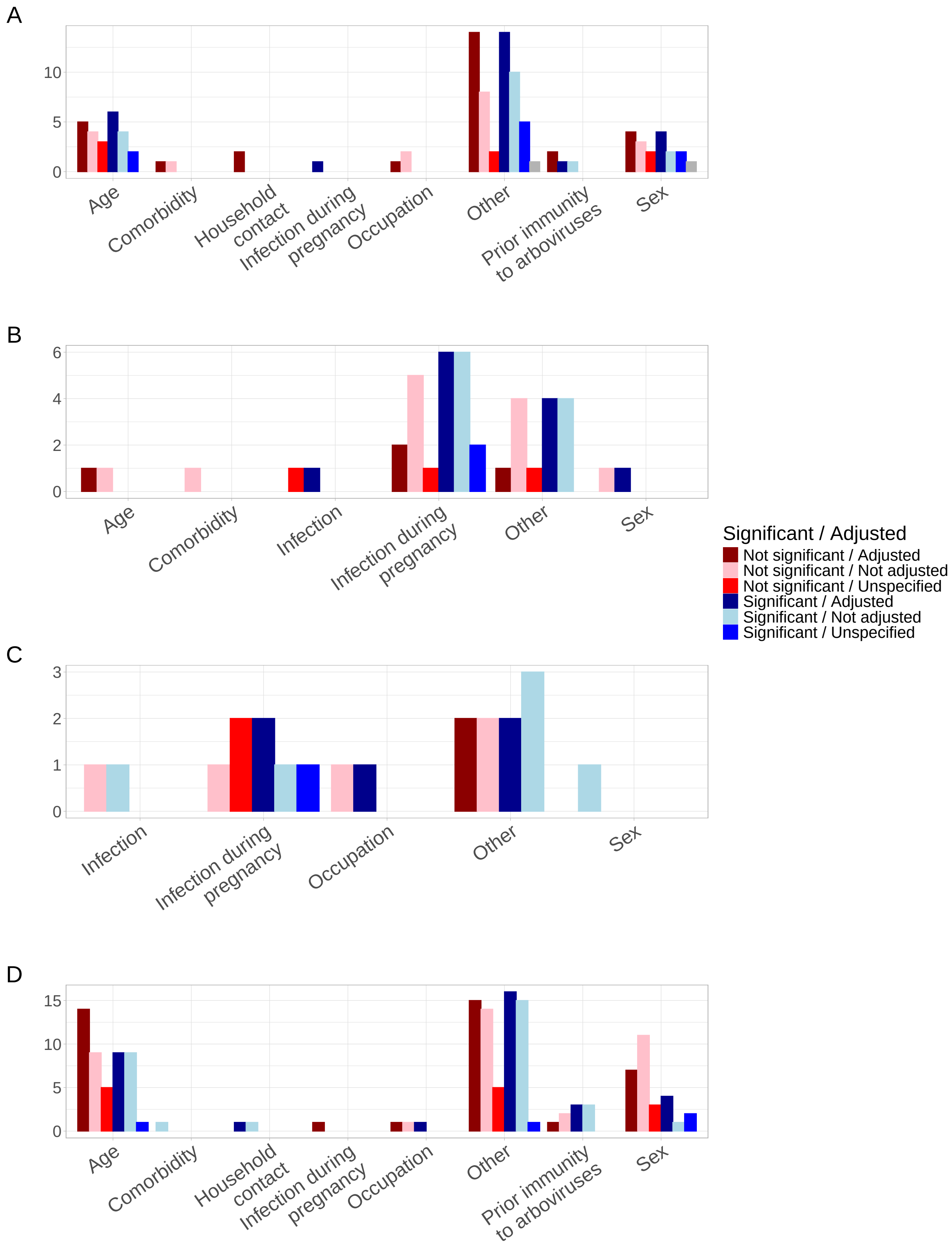

Figure B.23: Commonly reported outcomes and risk factors. A. Infection; B. Zika congenital syndrome; C. Microcephaly; D. Serology

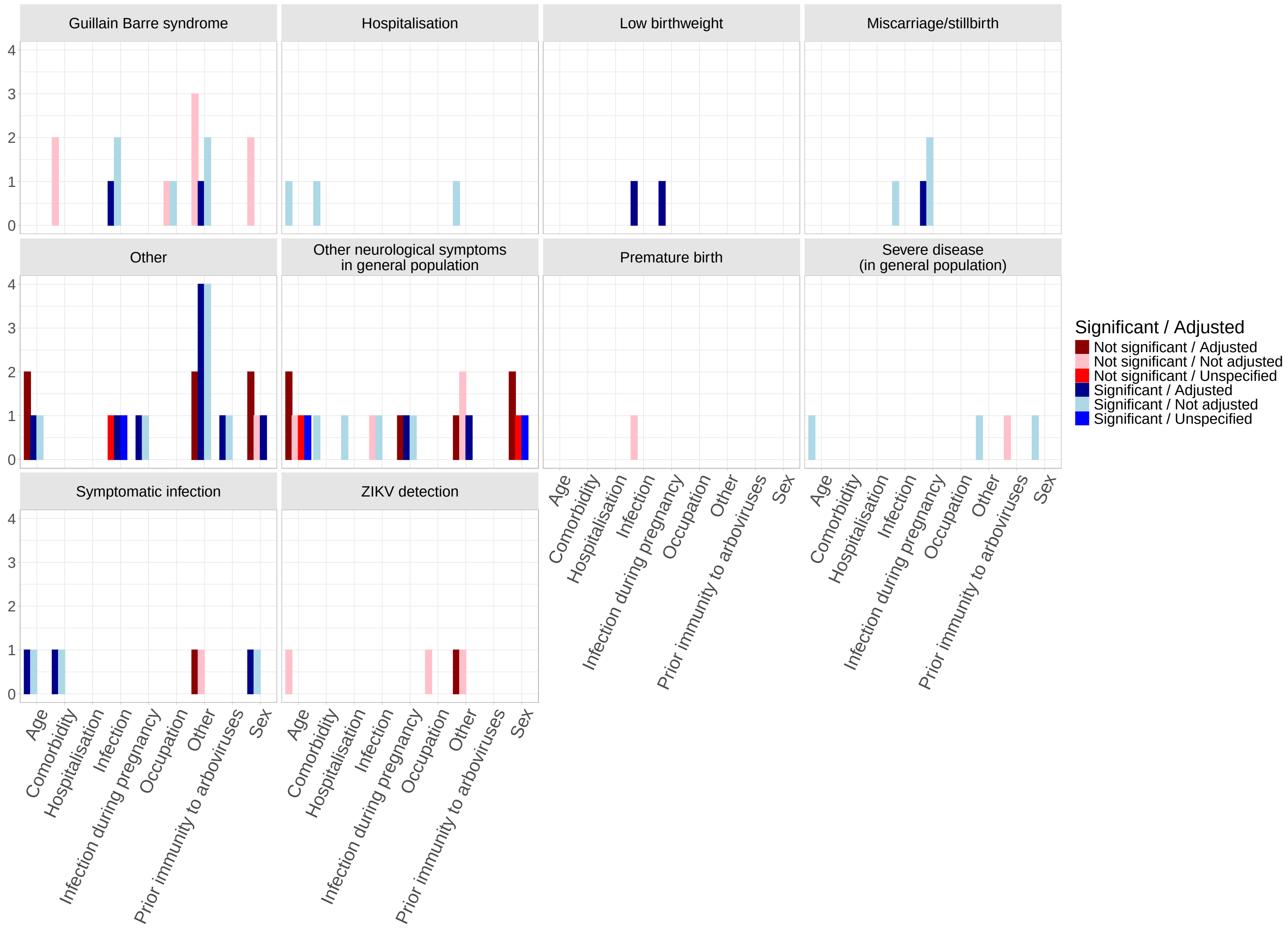

Figure B.24: Other reported outcomes and risk factors

#### B.4 Main figures with no QA filtering

Below, we present analogous versions of Figures 2-5 from the main text, but without filtering for studies with high QA scores.

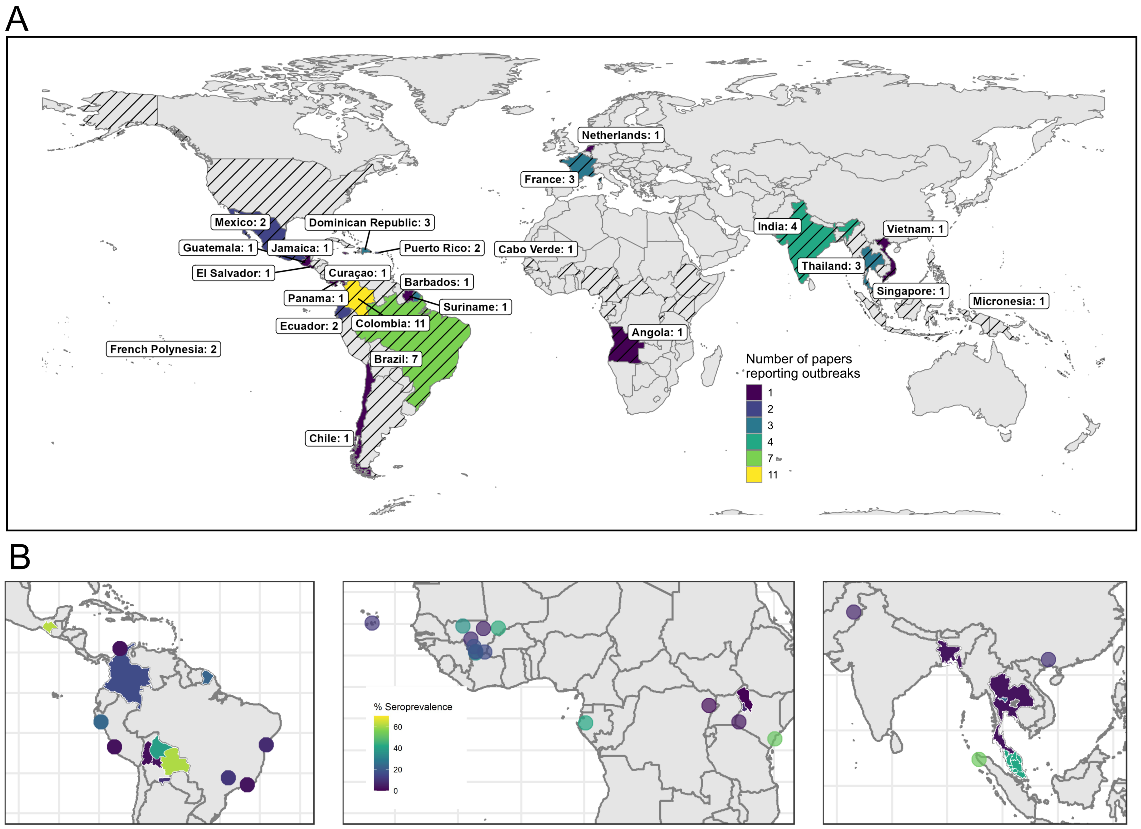

Figure B.25: ZIKV outbreak and seroprevalence mapping from all included studies: (A) Countries with papers reporting ZIKV outbreak information, coloured by number of publications. Outbreaks reported in France and Netherlands reflected outbreaks in overseas regions. Countries with black diagonal stripes indicate locations where ZIKV transmission has been reported by the WHO. (B) Geolocated areas or regions with ZIKV seroprevalence studies (using IgG assay, HAI/HI, MIA, NS1 BOB ELISA, IFA, capture ELISA and neutralisation assays) conducted in the general population in the Americas (left), Africa (centre) and Asia (right). Each dot represents a location-specific estimate, while shaded areas indicate estimates at the administrative unit level (region, province, district, or entire country).

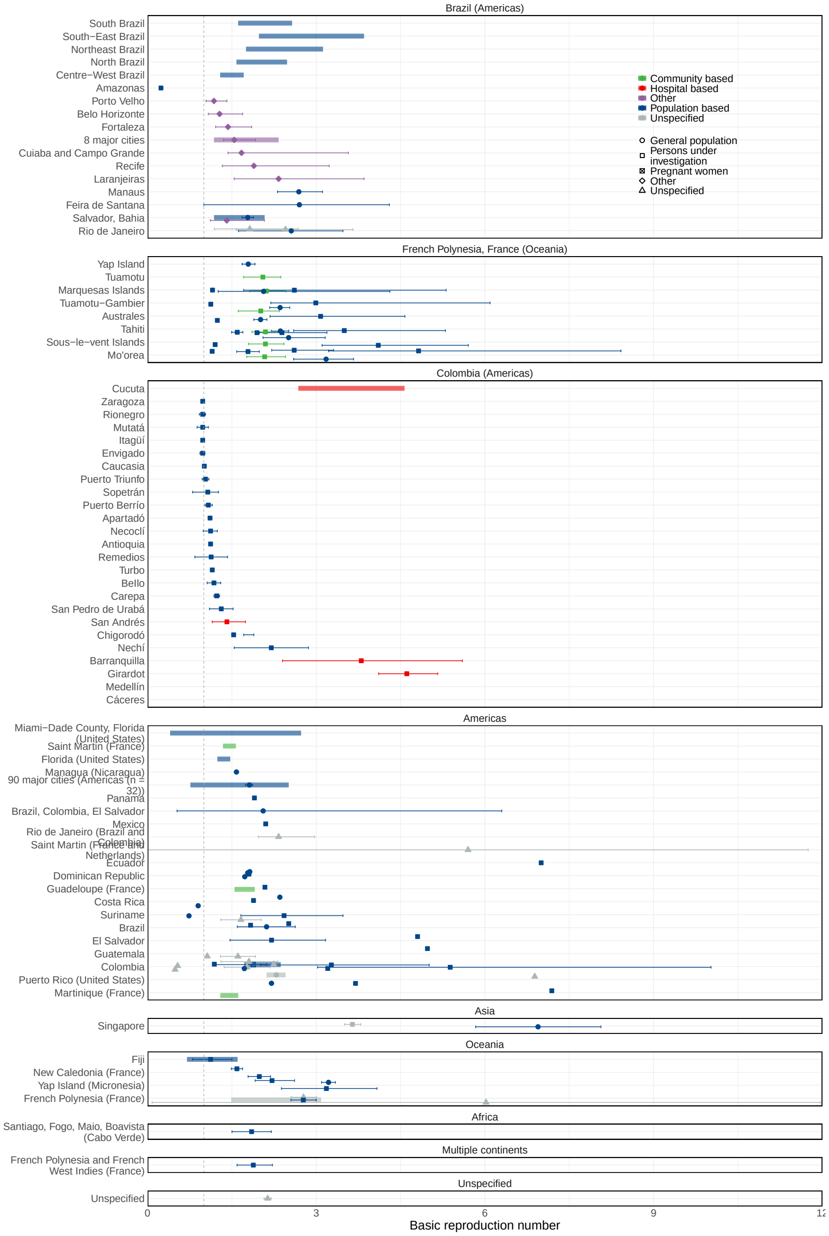

Figure B.26: Estimates by location, type of sample, and population group of the basic reproduction number of ZIKV. Points are central estimates, error bars are 95% confidence or credible intervals, and shaded bars are ranges of central estimates over disaggregated groups. When there were multiple estimates for the same location, points, error bars, and/or ranges were jittered. Note that the point estimates for Medellín (median 22.2) and Cáceres (median 56.38) are not shown because they are outside of the plot bounds.

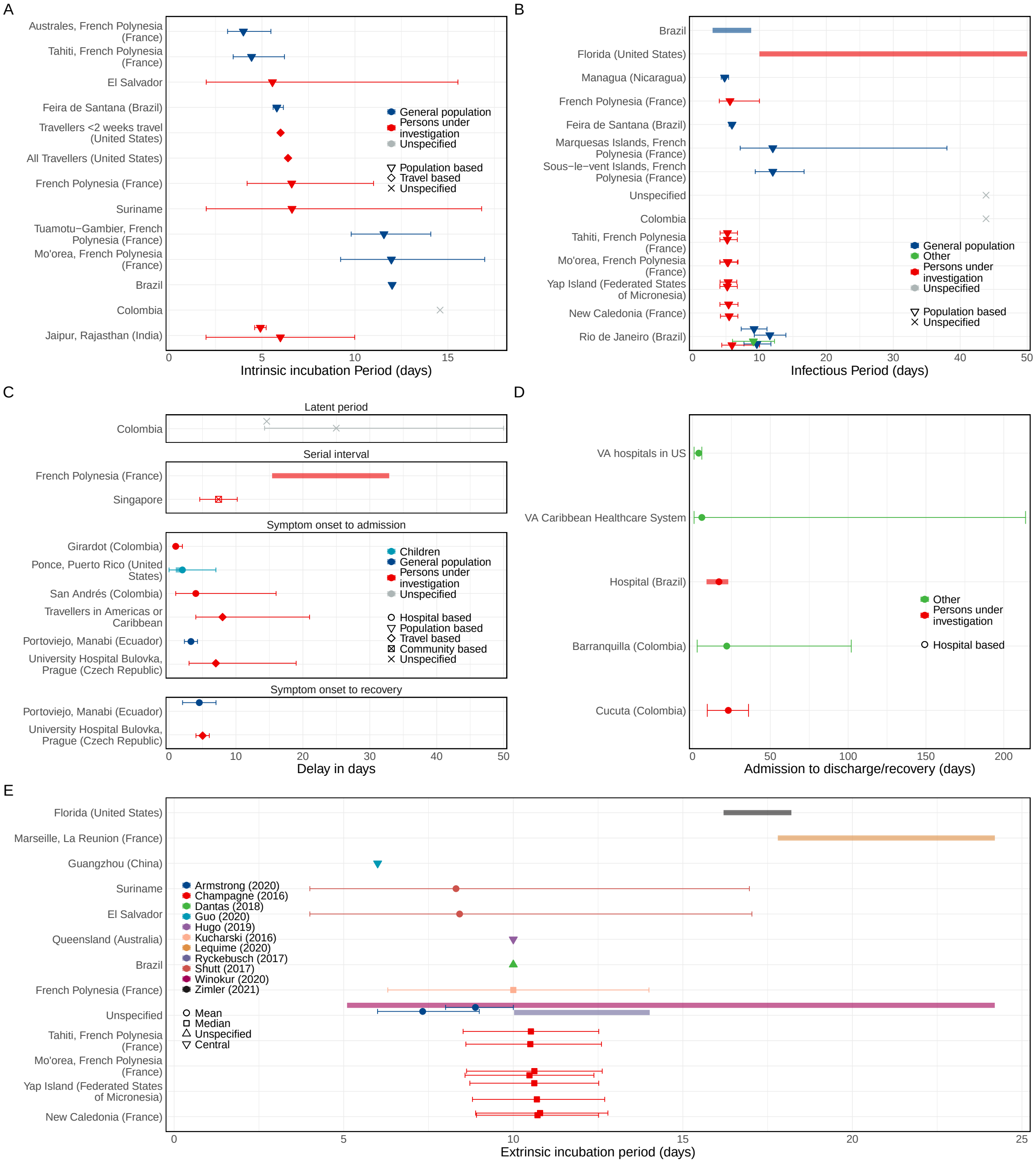

Figure B.27: Estimates by location and type of sample of the infectious period of ZIKV (A), incubation period (B), admission to discharge/recovery (C), and serial interval, symptom onset to admission, and symptom onset to recovery (D), and estimates by location and study of the extrinsic incubation period in the mosquito (E). Points are the central estimates reported in the studies, error bars are 95% confidence or credible intervals, and shaded bars are ranges of central estimates over disaggregated groups.

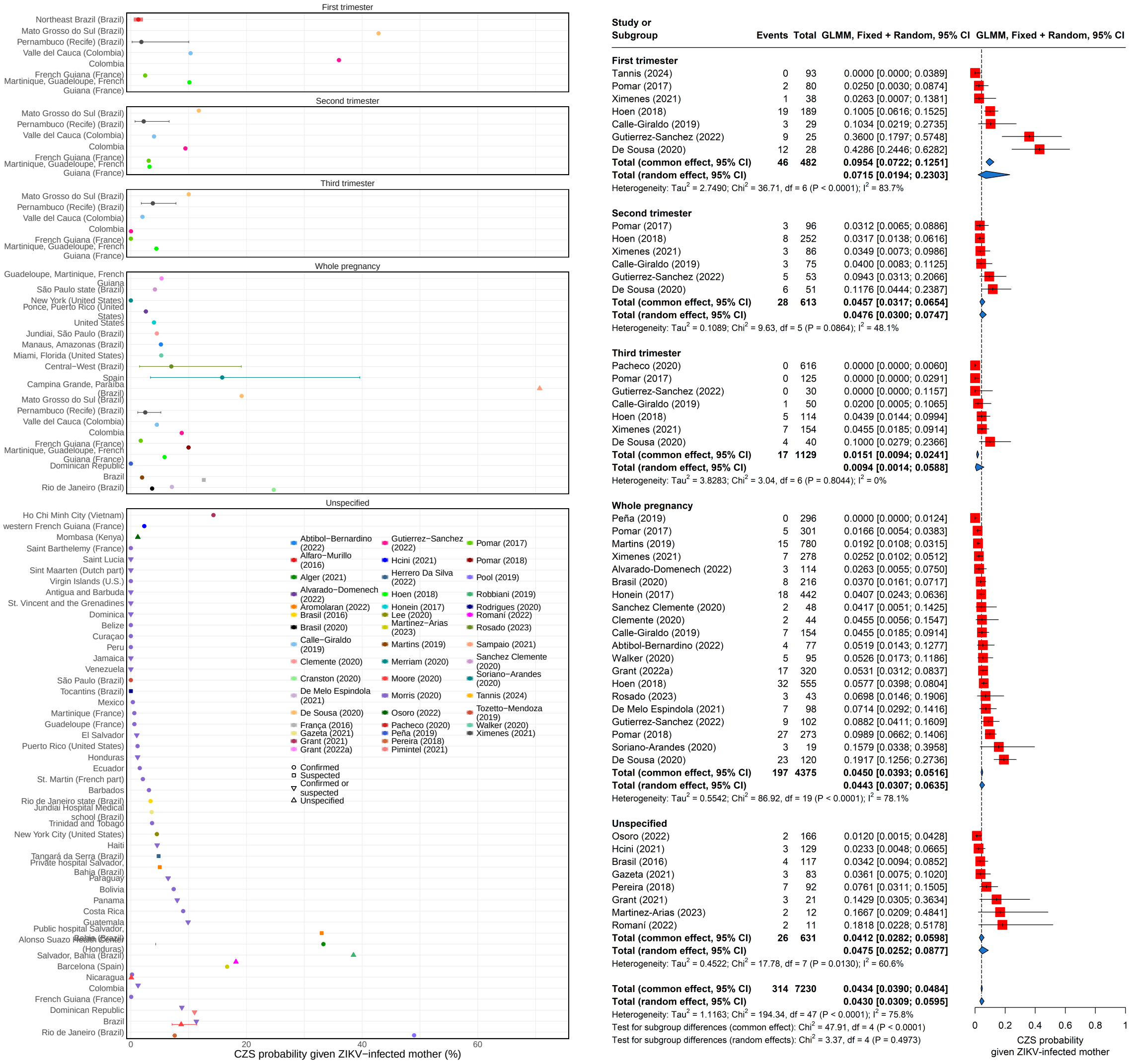

Figure B.28: (A) Estimates of reported CZS risks. The top panel shows estimates from Brazil, while the bottom panel shows estimates from the rest of the world. Points are central estimates, solid lines are confidence or credible intervals, and shaded segments are ranges of central estimates across multiple groups. (B) Meta-analysis of CZS risk stratified by population sample type. Red squares represent the observed study effect sizes, the solid black horizontal lines are confidence intervals, and blue diamonds are the pooled estimates for each sub-group and overall. The vertical dashed line is the overall pooled estimate.

#### B.5 Extended meta-analyses on CZS probability and pregnancy loss probability

In the main text, we presented our meta-analysis of CZS probability by trimester. We also show the meta-analysis of the pregnancy loss probability. We performed subgroup analyses by country (for Brazil), continent, and trimester for CZS probability and by country (for Brazil) and continent for pregnancy loss probability to explore the heterogeneity in estimates.

The CZS probability funnel plot (Figure B.29) and the pregnancy loss probability funnel plot (Figure B.30) are relatively symmetric around the overall pooled random effects estimates.

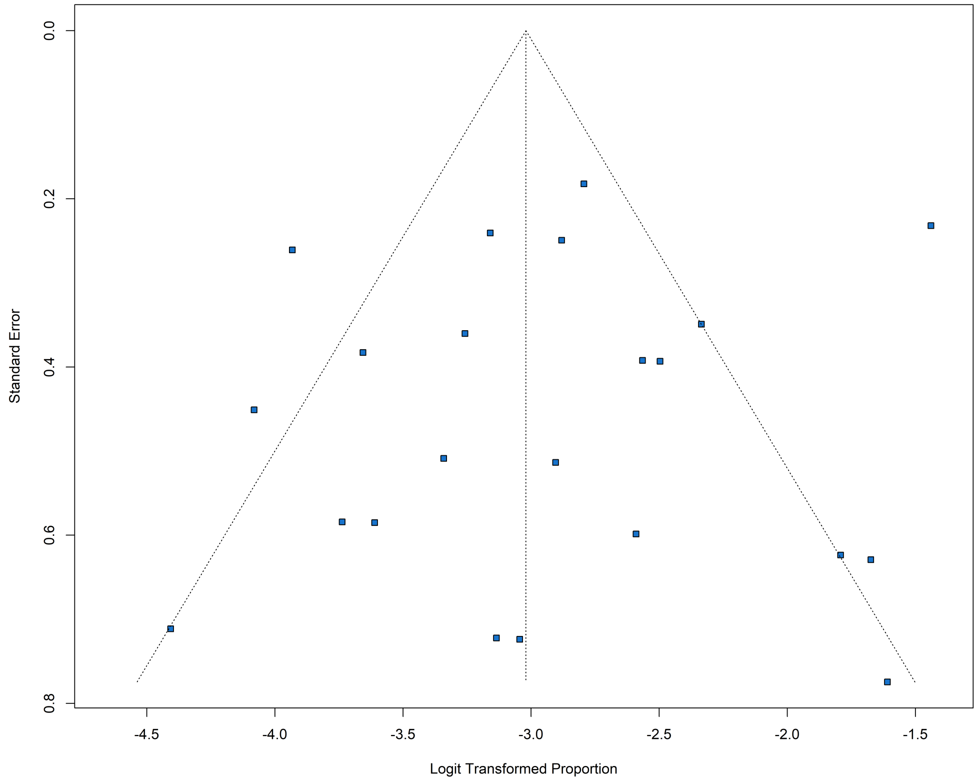

Figure B.29: Funnel plot for the CZS probability meta-analysis. Blue squares represent individual study estimates.

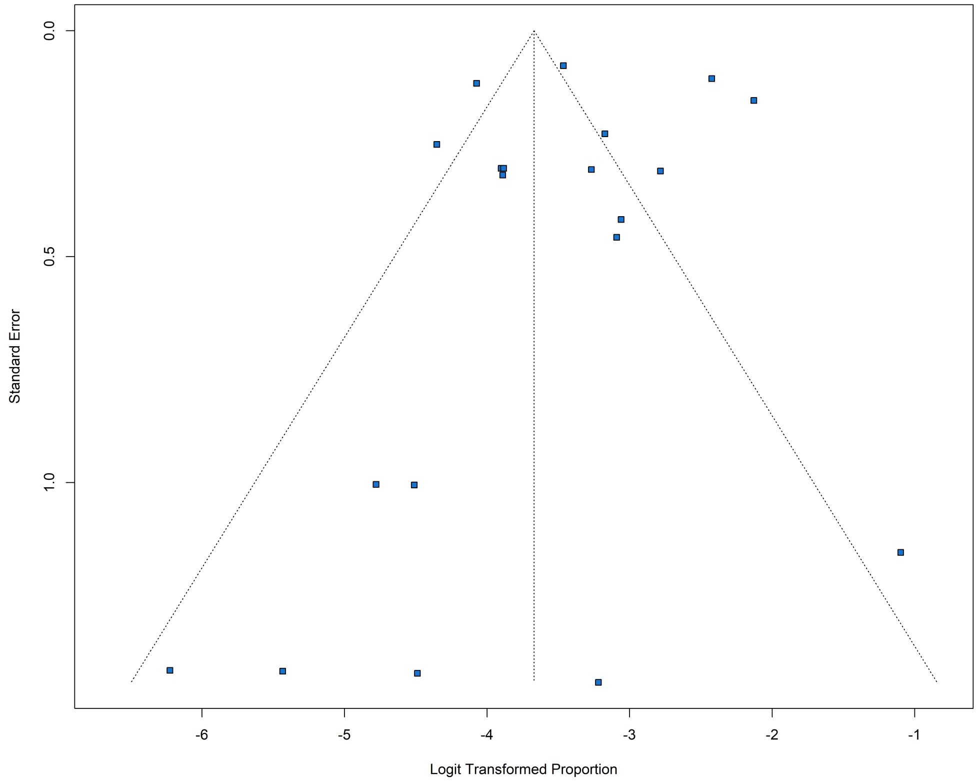

Figure B.30: Funnel plot for the pregnancy loss probability meta-analysis. Blue squares represent individual study estimates.

| Study or Subgroup | Events | Total | GLMM, Fixed + Random, 95% CI |
|---|---|---|---|
| **Africa** | | | |
| Osoro (2022) | 2 | 166 | 0.0120 [0.0015; 0.0428] |
| **Americas** | | | |
| Pomar (2017) | 5 | 301 | 0.0166 [0.0054; 0.0383] |
| Martins (2019) | 15 | 780 | 0.0192 [0.0108; 0.0315] |
| Hcini (2021) | 3 | 129 | 0.0233 [0.0048; 0.0665] |
| Ximenes (2021) | 7 | 278 | 0.0252 [0.0102; 0.0512] |
| Alvarado-Domenech (2022) | 3 | 114 | 0.0263 [0.0055; 0.0750] |
| Brasil (2016) | 4 | 117 | 0.0342 [0.0094; 0.0852] |
| Brasil (2020) | 8 | 216 | 0.0370 [0.0161; 0.0717] |
| Honein (2017) | 18 | 442 | 0.0407 [0.0243; 0.0636] |
| Sanchez Clemente (2020) | 2 | 48 | 0.0417 [0.0051; 0.1425] |
| Clemente (2020) | 2 | 44 | 0.0455 [0.0056; 0.1547] |
| Abtibol-Bernardino (2022) | 4 | 77 | 0.0519 [0.0143; 0.1277] |
| Grant (2022a) | 17 | 320 | 0.0531 [0.0312; 0.0837] |
| Hoen (2018) | 32 | 555 | 0.0577 [0.0398; 0.0804] |
| Rosado (2023) | 3 | 43 | 0.0698 [0.0146; 0.1906] |
| De Melo Espindola (2021) | 7 | 98 | 0.0714 [0.0292; 0.1416] |
| Pereira (2018) | 7 | 92 | 0.0761 [0.0311; 0.1505] |
| Gutierrez-Sanchez (2022) | 9 | 102 | 0.0882 [0.0411; 0.1609] |
| De Sousa (2020) | 23 | 120 | 0.1917 [0.1256; 0.2736] |
| **Total (common effect, 95% CI)** | **169** | **3876** | **0.0436 [0.0376; 0.0505]** |
| **Total (random effect, 95% CI)** | | | **0.0443 [0.0321; 0.0607]** |
| Heterogeneity: $Tau^2 = 0.3425$; $Chi^2 = 77.68$, df = 17 ($P < 0.0001$); $I^2 = 78.1\%$ | | | |
| **Asia** | | | |
| Grant (2021) | 3 | 21 | 0.1429 [0.0305; 0.3634] |
| **Europe (travel-based)** | | | |
| Soriano-Arandes (2020) | 3 | 19 | 0.1579 [0.0338; 0.3958] |
| Martinez-Arias (2023) | 2 | 12 | 0.1667 [0.0209; 0.4841] |
| **Total (common effect, 95% CI)** | **5** | **31** | **0.1613 [0.0688; 0.3337]** |
| **Total (random effect, 95% CI)** | | | **0.1613 [0.0688; 0.3337]** |
| Heterogeneity: $Tau^2 = 0$; $Chi^2 = 0$, df = 1 ($P = 0.9484$); $I^2 = 0\%$ | | | |
| **Total (common effect, 95% CI)** | **179** | **4094** | **0.0437 [0.0379; 0.0504]** |
| **Total (random effect, 95% CI)** | | | **0.0465 [0.0338; 0.0637]** |
| Heterogeneity: $Tau^2 = 0.4077$; $Chi^2 = 91.71$, df = 21 ($P < 0.0001$); $I^2 = 77.1\%$ | | | |
| Test for subgroup differences (common effect): $Chi^2 = 16.18$, df = 3 ($P = 0.0010$) | | | |
| Test for subgroup differences (random effects): $Chi^2 = 15.22$, df = 3 ($P = 0.0016$) | | | |

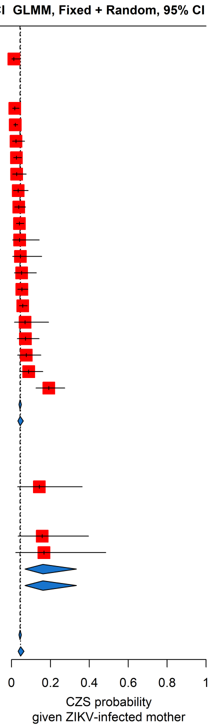

Figure B.31: CZS probability meta-analysis stratified by continent.

| Study or Subgroup | Events | Total | GLMM, Fixed + Random, 95% CI |
|---|---|---|---|
| **First trimester** | | | |
| Ximenes (2021) | 1 | 38 | 0.0263 [0.0007; 0.1381] |
| De Sousa (2020) | 12 | 28 | 0.4286 [0.2446; 0.6282] |
| **Total (common effect, 95% CI)** | **13** | **66** | **0.1970 [0.1180; 0.3103]** |
| **Total (random effect, 95% CI)** | | | **0.1313 [0.0126; 0.6421]** |
| Heterogeneity: $Tau^2 = 2.6414$; $Chi^2 = 9.42$, df = 1 (P = 0.0022); $I^2 = 89.4\%$ | | | |
| **Second trimester** | | | |
| Ximenes (2021) | 3 | 86 | 0.0349 [0.0073; 0.0986] |
| De Sousa (2020) | 6 | 51 | 0.1176 [0.0444; 0.2387] |
| **Total (common effect, 95% CI)** | **9** | **137** | **0.0657 [0.0345; 0.1214]** |
| **Total (random effect, 95% CI)** | | | **0.0654 [0.0276; 0.1474]** |
| Heterogeneity: $Tau^2 = 0.1763$; $Chi^2 = 3.19$, df = 1 (P = 0.0741); $I^2 = 68.6\%$ | | | |
| **Third trimester** | | | |
| Ximenes (2021) | 7 | 154 | 0.0455 [0.0185; 0.0914] |
| De Sousa (2020) | 4 | 40 | 0.1000 [0.0279; 0.2366] |
| **Total (common effect, 95% CI)** | **11** | **194** | **0.0567 [0.0317; 0.0995]** |
| **Total (random effect, 95% CI)** | | | **0.0567 [0.0317; 0.0995]** |
| Heterogeneity: $Tau^2 = 0$; $Chi^2 = 1.68$, df = 1 (P = 0.1950); $I^2 = 40.5\%$ | | | |
| **Unspecified** | | | |
| Brasil (2016) | 4 | 117 | 0.0342 [0.0094; 0.0852] |
| Pereira (2018) | 7 | 92 | 0.0761 [0.0311; 0.1505] |
| **Total (common effect, 95% CI)** | **11** | **209** | **0.0526 [0.0294; 0.0925]** |
| **Total (random effect, 95% CI)** | | | **0.0526 [0.0294; 0.0925]** |
| Heterogeneity: $Tau^2 = 0$; $Chi^2 = 1.72$, df = 1 (P = 0.1891); $I^2 = 42\%$ | | | |
| **Whole pregnancy** | | | |
| Martins (2019) | 15 | 780 | 0.0192 [0.0108; 0.0315] |
| Ximenes (2021) | 7 | 278 | 0.0252 [0.0102; 0.0512] |
| Brasil (2020) | 8 | 216 | 0.0370 [0.0161; 0.0717] |
| Sanchez Clemente (2020) | 2 | 48 | 0.0417 [0.0051; 0.1425] |
| Clemente (2020) | 2 | 44 | 0.0455 [0.0056; 0.1547] |
| Abtibol-Bernardino (2022) | 4 | 77 | 0.0519 [0.0143; 0.1277] |
| Rosado (2023) | 3 | 43 | 0.0698 [0.0146; 0.1906] |
| De Melo Espindola (2021) | 7 | 98 | 0.0714 [0.0292; 0.1416] |
| De Sousa (2020) | 23 | 120 | 0.1917 [0.1256; 0.2736] |
| **Total (common effect, 95% CI)** | **71** | **1704** | **0.0417 [0.0331; 0.0523]** |
| **Total (random effect, 95% CI)** | | | **0.0476 [0.0279; 0.0799]** |
| Heterogeneity: $Tau^2 = 0.4998$; $Chi^2 = 60.74$, df = 8 (P < 0.0001); $I^2 = 86.8\%$ | | | |
| **Total (common effect, 95% CI)** | **115** | **2310** | **0.0498 [0.0416; 0.0594]** |
| **Total (random effect, 95% CI)** | | | **0.0580 [0.0374; 0.0891]** |
| Heterogeneity: $Tau^2 = 0.7011$; $Chi^2 = 108.44$, df = 16 (P < 0.0001); $I^2 = 85.2\%$ | | | |
| Test for subgroup differences (common effect): $Chi^2 = 27.62$, df = 4 (P < 0.0001) | | | |
| Test for subgroup differences (random effects): $Chi^2 = 1.05$, df = 4 (P = 0.9022) | | | |

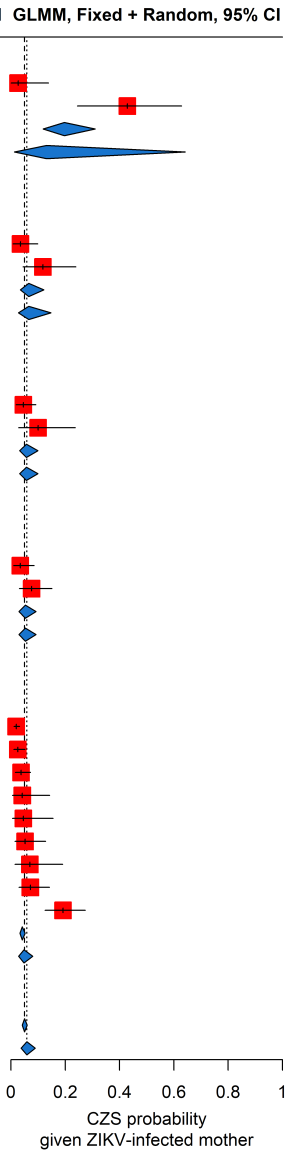

Figure B.32: CZS probability meta-analysis of estimates from Brazil, stratified by trimester of ZIKV exposure. Red squares represent the observed study effect sizes, the solid black horizontal lines are confidence intervals, and blue diamonds are the pooled estimates for each sub-group and overall. The vertical dashed line is the overall pooled estimate.

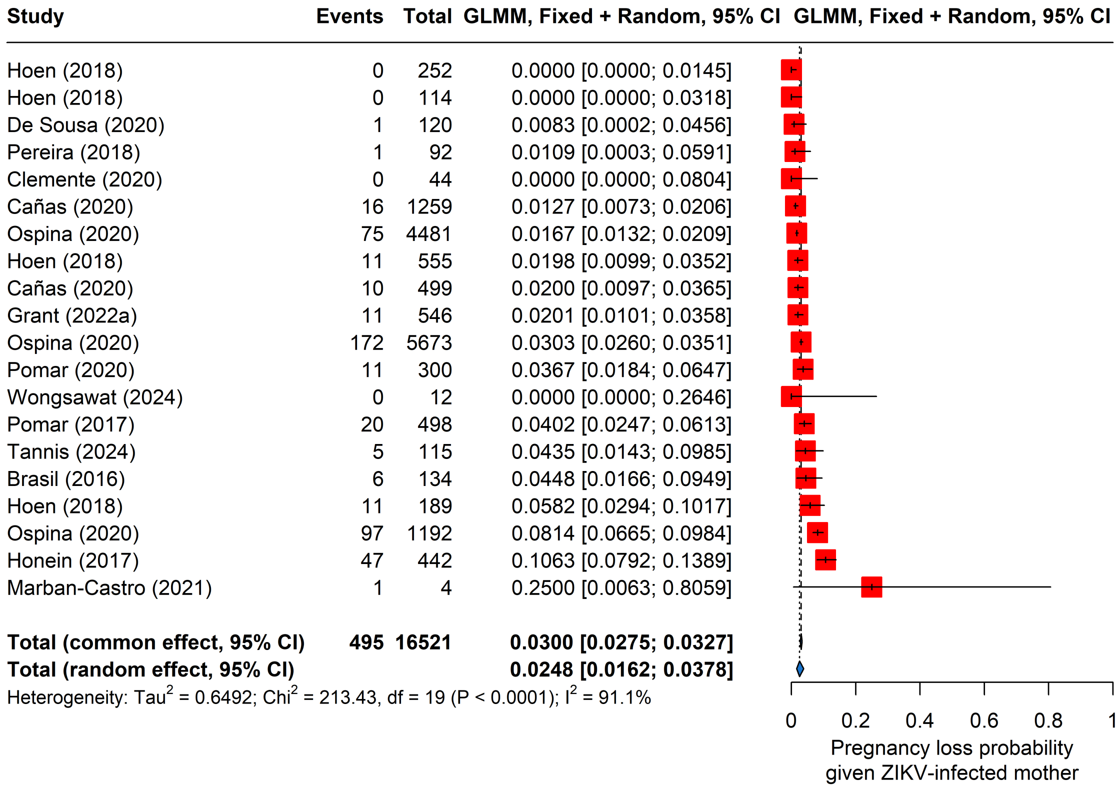

Figure B.33: Pregnancy loss probability meta-analysis, non-stratified. Red squares represent the observed study effect sizes, the solid black horizontal lines are confidence intervals, and blue diamonds are the pooled estimates for each sub-group and overall. The vertical dashed line is the overall pooled estimate.

| Study or Subgroup | Events | Total | GLMM, Fixed + Random, 95% CI |
|---|---|---|---|
| **Miscarriage** | | | |
| Hoen (2018) | 0 | 252 | 0.0000 [0.0000; 0.0145] |
| Hoen (2018) | 0 | 114 | 0.0000 [0.0000; 0.0318] |
| Cañas (2020) | 16 | 1259 | 0.0127 [0.0073; 0.0206] |
| Hoen (2018) | 11 | 555 | 0.0198 [0.0099; 0.0352] |
| Cañas (2020) | 10 | 499 | 0.0200 [0.0097; 0.0365] |
| Grant (2022a) | 11 | 546 | 0.0201 [0.0101; 0.0358] |
| Wongsawat (2024) | 0 | 12 | 0.0000 [0.0000; 0.2646] |
| Pomar (2017) | 20 | 498 | 0.0402 [0.0247; 0.0613] |
| Brasil (2016) | 6 | 134 | 0.0448 [0.0166; 0.0949] |
| Hoen (2018) | 11 | 189 | 0.0582 [0.0294; 0.1017] |
| Marban-Castro (2021) | 1 | 4 | 0.2500 [0.0063; 0.8059] |
| **Total (common effect, 95% CI)** | **86** | **4062** | **0.0212 [0.0172; 0.0261]** |
| **Total (random effect, 95% CI)** | | | **0.0204 [0.0121; 0.0343]** |
| Heterogeneity: $Tau^2$ = 0.4763; $Chi^2$ = 28.37, df = 10 (P = 0.0016); $I^2$ = 64.7% | | | |
| **General pregnancy loss** | | | |
| De Sousa (2020) | 1 | 120 | 0.0083 [0.0002; 0.0456] |
| Pereira (2018) | 1 | 92 | 0.0109 [0.0003; 0.0591] |
| Clemente (2020) | 0 | 44 | 0.0000 [0.0000; 0.0804] |
| Ospina (2020) | 75 | 4481 | 0.0167 [0.0132; 0.0209] |
| Ospina (2020) | 172 | 5673 | 0.0303 [0.0260; 0.0351] |
| Pomar (2020) | 11 | 300 | 0.0367 [0.0184; 0.0647] |
| Tannis (2024) | 5 | 115 | 0.0435 [0.0143; 0.0985] |
| Ospina (2020) | 97 | 1192 | 0.0814 [0.0665; 0.0984] |
| Honein (2017) | 47 | 442 | 0.1063 [0.0792; 0.1389] |
| **Total (common effect, 95% CI)** | **409** | **12459** | **0.0328 [0.0298; 0.0361]** |
| **Total (random effect, 95% CI)** | | | **0.0316 [0.0176; 0.0561]** |
| Heterogeneity: $Tau^2$ = 0.5861; $Chi^2$ = 174.11, df = 8 (P < 0.0001); $I^2$ = 95.4% | | | |
| **Total (common effect, 95% CI)** | **495** | **16521** | **0.0300 [0.0275; 0.0327]** |
| **Total (random effect, 95% CI)** | | | **0.0248 [0.0162; 0.0378]** |
| Heterogeneity: $Tau^2$ = 0.6492; $Chi^2$ = 213.43, df = 19 (P < 0.0001); $I^2$ = 91.1% | | | |
| Test for subgroup differences (common effect): $Chi^2$ = 14.09, df = 1 (P = 0.0002) | | | |
| Test for subgroup differences (random effects): $Chi^2$ = 1.21, df = 1 (P = 0.2717) | | | |

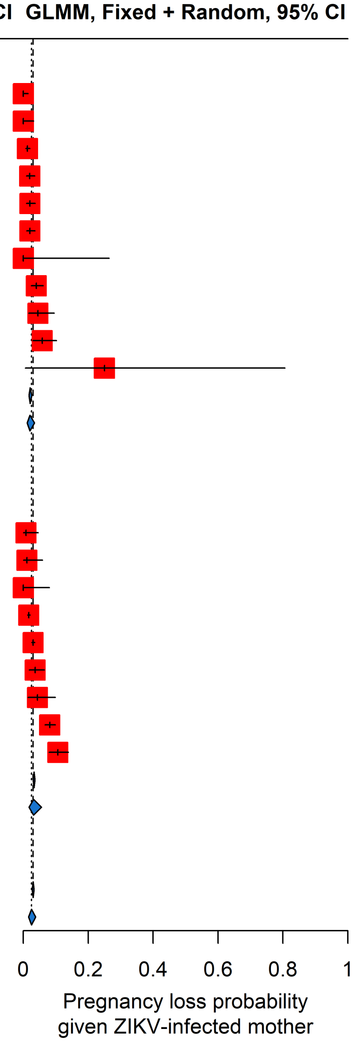

Figure B.34: Pregnancy loss probability meta-analysis, stratified by pregnancy outcome.Red squares represent the observed study effect sizes, the solid black horizontal lines are confidence intervals, and blue diamonds are the pooled estimates for each sub-group and overall. The vertical dashed line is the overall pooled estimate.

#### B.6 Meta-analyses on proportion of symptomatic individuals

In the main text, we presented the results of our meta-analysis of the proportion of symptomatic individuals. This analysis includes 7 estimates (from 2 papers) from overseas territories of France. In more details, 6 estimates come from a study in the general population and children of the French Polynesia (Society Islands, Tuamotu Islands, Marquesas islands and Austral-Gamber islands) and one from a study in pregnant women in French Guiana. The meta-analysis is shown in Figure B.35 and the funnel plot (Figure B.36) is relatively symmetric around the overall pooled random effects estimates.

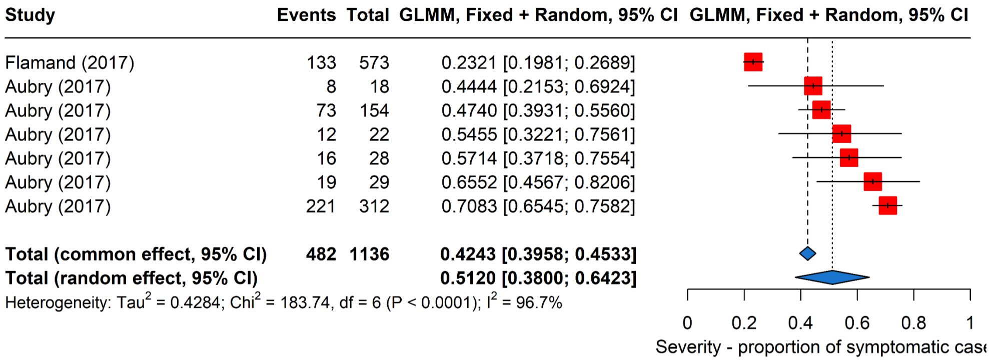

Figure B.35: Meta-analysis of the proportion of symptomatic individuals.

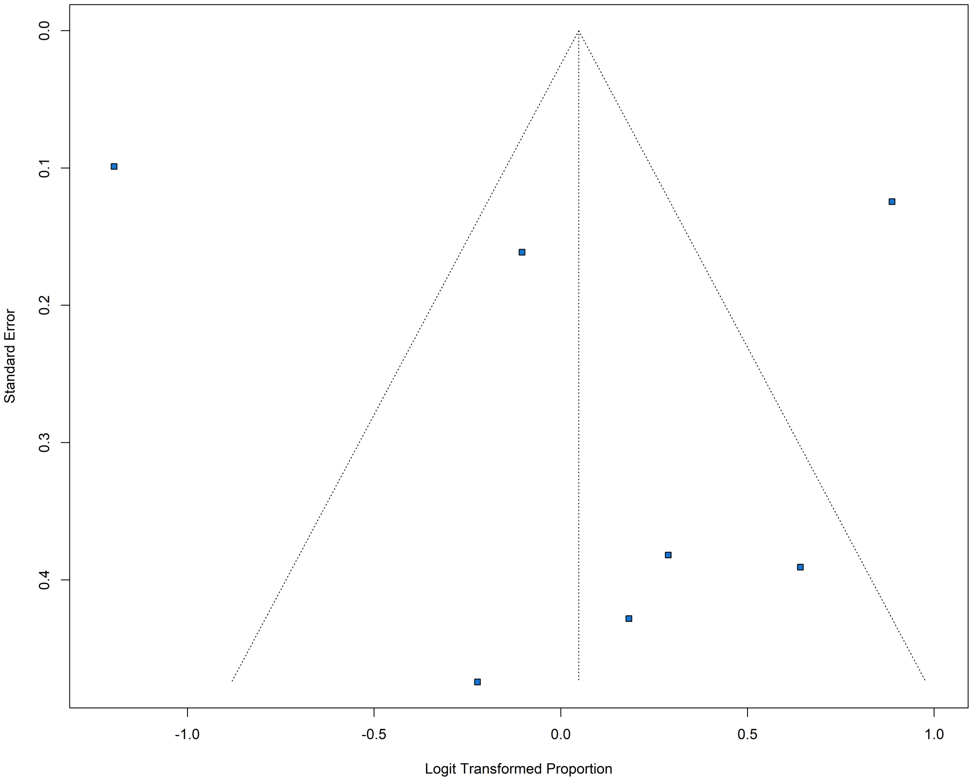

Figure B.36: Funnel plot for the or proportion of symptomatic individuals meta-analysis. Blue squares represent individual study estimates.

#### B.7 Tables of extracted parameters, models, and outbreaks

##### B.7.1 Models

Information on transmission models was extracted from 154 studies ( Appendix: Table B4, Table B8, Figure B4), the majority of which were compartmental (n=120). Other published models included branching processes (n=6) and individual-based models (n=6). Most models accounted for vector-borne transmission (n=127), and some also included human-to-human sexual transmission (n=45). The most common compartmental type used was the SEIR-SEI model (n=47), with Susceptible–Exposed–Infectious–Recovered (SEIR) compartments for humans, and Susceptible–Exposed-Infectious (SEI) compartments for vectors. Notably, no modelling studies were published before the outbreak in Brazil in 2015, but there was a large surge in modelling publications in the following years, with 11 modelling studies published in 2024. Among the 154 studies reporting ZIKV transmission models, only 26 studies (17%) published the associated code.

| Transmission Route(s) | Model Assumptions | Model Compartments | Fitted | Interventions | Source |
|---|---|---|---|---|---|
| **Branching Process** | | | | | |
| Unspecified | Unspecified | | Fitted | | Chowell (2016) |
| Unspecified | Unspecified | | Fitted | | Zhao (2019) |
| Unspecified | | | Fitted | | Chowell (2021) |
| Mosquito-Human | Unspecified | SIR-SI | Theoretical | Behaviour Changes, Other | Soewono (2021) |
| Mosquito-Human | Heterogenity in transmission over time, Homogeneous mixing, Other | SIR | Fitted | | Riou (2017) |
| Unspecified | Unspecified | SIR | Fitted | | Dinh (2016) |
| Mosquito-Human | Homogeneous mixing | Other | Fitted | Treatment, Vaccination | Carmona (2020) |
| **Branching process;Compartmental - Stochastic** | | | | | |
| Mosquito-Human | Other | SEIR | Theoretical | | Castro (2017) |
| **Compartmental - Deterministic** | | | | | |
| Mosquito-Human | Heterogenity in transmission between human and vector, Heterogenity in transmission over time | SEIR-SEI | Theoretical | Behaviour Changes, Pesticides/larvicides, Treatment, Vaccination, Wolbachia replacement, Wolbachia suppression | Ngonghala (2021) |
| Mosquito-Human | Heterogenity in transmission between human and vector, Heterogenity in transmission over time, Latent and incubation periods are same | SEIR-SEI | Theoretical | | Huber (2018) |
| Mosquito-Human | Heterogenity in transmission over time, Homogeneous mixing | SEIR-SEI | Theoretical | | Roy (2020) |
| Mosquito-Human | Homogeneous mixing | SEIR-SEI | Theoretical | | Suantai (2023) |
| Mosquito-Human | Homogeneous mixing | SEIR-SEI | Theoretical | | Kumar (2022) |
| Mosquito-Human | Homogeneous mixing | SEIR-SEI | Fitted | | Prasad (2023) |
| Mosquito-Human | Homogeneous mixing | SEIR-SEI | Fitted | | Kumar (2017) |
| Mosquito-Human | Homogeneous mixing | SEIR-SEI | Fitted | | Dantas (2018) |
| Mosquito-Human | Homogeneous mixing | SEIR-SEI | Theoretical | Wolbachia replacement | Xue (2018) |
| Mosquito-Human | Homogeneous mixing, Latent and incubation periods are same | SEIR-SEI | Fitted | Other, Pesticides/larvicides, Treatment | Alzahrani (2021) |
| Mosquito-Human | Homogeneous mixing, Other | SEIR-SEI | Fitted | | Funk (2016) |
| Mosquito-Human | Homogeneous mixing, Other | SEIR-SEI | Fitted | Insecticide-treated nets, Other, Quarantine, Mosquito Control | Yue (2023) |
| Mosquito-Human | Homogeneous mixing, Other | SEIR-SEI | Fitted | | Rahman (2019) |
| Mosquito-Human | Other | SEIR-SEI | Fitted | | Chen (2018) |
| Mosquito-Human | Other | SEIR-SEI | Theoretical | | Yue (2022) |
| Mosquito-Human | Other | SEIR-SEI | Theoretical | Mosquito Control | Yue (2020) |
| Mosquito-Human | Unspecified | SEIR-SEI | Theoretical | | Imran (2021) |
| Mosquito-Human | Unspecified | SEIR-SEI | Theoretical | | Gao (2019) |
| Mosquito-Human | | SEIR-SEI | Theoretical | | Moreno (2017) |
| Mosquito-Human | | SEIR-SEI | Theoretical | Other, Mosquito Control | Agusto (2017a) |
| Mosquito-Human | | SEIR-SEI | Theoretical | Insecticide-treated nets, Treatment, Mosquito Control | Bonyah (2017) |
| Mosquito-Human | | SEIR-SEI | Theoretical | Mosquito Control | Stone (2019) |
| Mosquito-Human | | SEIR-SEI | Fitted | | Caldwell (2021) |
| Mosquito-Human | | SEIR-SEI | Theoretical | | Nisar (2024) |
| Sexual, Mosquito-Human | Heterogenity in transmission between human and vector, Heterogenity in transmission between human groups | SEIR-SEI | Fitted | Behaviour Changes, Insecticide-treated nets, Mosquito Control | Kumar Biswas (2020) |
| Sexual, Mosquito-Human | Heterogenity in transmission between human and vector, Heterogenity in transmission between human groups | SEIR-SEI | Theoretical | | Thaiprayoon (2022) |

continued on next page

continued from previous page

| Model Compartments | Transmission Route(s) | Model Assumptions | Fitted | Interventions | Source |
|---|---|---|---|---|---|
| Sexual, Mosquito-Human | Heterogenity in transmission between human and vector, Homogeneous mixing, Latent and incubation periods are same | SEIR-SEI | Theoretical | | Towers (2016) |
| Sexual, Mosquito-Human | Homogeneous mixing | SEIR-SEI | Theoretical | | Yamazaki (2019) |
| Sexual, Mosquito-Human | Homogeneous mixing | SEIR-SEI | Fitted | Behaviour Changes, Pesticides/larvicides, Vaccination | Valega-Mackenzie (2023) |
| Sexual, Mosquito-Human | Latent and incubation periods are same | SEIR-SEI | Theoretical | Indoor residual spraying, Insecticide-treated nets | Padmanabhan (2017b) |
| Sexual, Mosquito-Human | Other | SEIR-SEI | Theoretical | Behaviour Changes, Genetically modified mosquitoes, Indoor residual spraying, Mosquito Control, Wolbachia replacement | Bi (2020) |
| Sexual, Mosquito-Human | Unspecified | SEIR-SEI | Theoretical | Indoor residual spraying, Insecticide-treated nets, Treatment | Ali (2022b) |
| Sexual, Mosquito-Human | Unspecified | SEIR-SEI | Theoretical | | Khan (2021) |
| Sexual, Mosquito-Human | Unspecified | SEIR-SEI | Fitted | Other, Vaccination, Mosquito Control | Burgess (2021) |
| Sexual, Mosquito-Human | Unspecified | SEIR-SEI | Fitted | | Sow (2022) |
| Sexual, Mosquito-Human | Unspecified | SEIR-SEI | Theoretical | Vaccination | Hussain (2021) |
| Sexual, Mosquito-Human | | SEIR-SEI | Fitted | Mosquito Control | Wang (2017) |
| Sexual, Mosquito-Human | | SEIR-SEI | Fitted | | Ibrahim (2021) |
| Sexual, Mosquito-Human | | SEIR-SEI | Theoretical | | Harvim (2019) |
| Human-Human, Sexual, Mosquito-Human | Other | SEIR-SEI | Theoretical | | Hasan (2019) |
| Human-Human, Mosquito-Human | Heterogenity in transmission between human groups | SEIR-SEI | Theoretical | Behaviour Changes, Indoor residual spraying, Other, Vaccination | Okyere (2020) |
| Human-Human, Mosquito-Human | Homogeneous mixing | SEIR-SEI | Fitted | | Sadeghieh (2021) |
| Human-Human, Mosquito-Human | Homogeneous mixing, Latent and incubation periods are same, Other | SEIR-SEI | Fitted | | Kucharski (2016) |
| Human-Human, Mosquito-Human | Other | SEIR-SEI | Theoretical | Hospitals | Imran (2018) |
| Human-Human, Mosquito-Human | Unspecified | SEIR-SEI | Theoretical | Treatment, Vaccination | Jan (2023) |
| Human-Human, Mosquito-Human | Unspecified | SEIR-SEI | Theoretical | | Bekiryazici (2022) |
| Human-Human, Mosquito-Human | Unspecified | SEIR-SEI | Fitted | | Ali (2022a) |
| Human-Human, Mosquito-Human | Unspecified | SEIR-SEI | Theoretical | Behaviour Changes, Insecticide-treated nets, Other, Mosquito Control | Goswami (2021) |
| Human-Human, Mosquito-Human | Unspecified | SEIR-SEI | Theoretical | | Veeresha (2022) |
| Mosquito-Human | Other | SEI-SI | Theoretical | | Martin (2021) |
| Sexual, Mosquito-Human | Other | SEIR, SIR | Theoretical | | Terefe (2018) |
| Human-Human, Mosquito-Human | Unspecified | SI-SI | Theoretical | | Dharmalingam (2024) |
| Mosquito-Human | Homogeneous mixing | SIR-SEI | Theoretical | Vaccination | Massad (2019) |
| Human-Human, Mosquito-Human | Unspecified | SIR-SEI | Theoretical | Vaccination | Sharma (2021) |
| Mosquito-Human | Heterogenity in transmission over time, Other | SIR-SI | Theoretical | | Al Najim (2024) |
| Mosquito-Human | Other | SIR-SI | Theoretical | | Rakkiyappan (2019) |
| Mosquito-Human | Unspecified | SIR-SI | Theoretical | Behaviour Changes, Other | Soewono (2021) |
| Mosquito-Human | Unspecified | SIR-SI | Theoretical | | Alfwzan (2023) |
| Mosquito-Human | Unspecified | SIR-SI | Theoretical | | Alshehri (2022) |
| Mosquito-Human | Unspecified | SIR-SI | Theoretical | | Alkahtani (2017) |
| Mosquito-Human | | SIR-SI | Fitted | | Mishra (2021) |
| Mosquito-Human | | SIR-SI | Theoretical | | Jiao (2021) |
| Sexual, Mosquito-Human | Heterogenity in transmission between human and vector, Heterogenity in transmission between human groups, Other | SIR-SI | Fitted | Treatment | Zhao (2020) |
| Sexual, Mosquito-Human | Other | SIR-SI | Fitted | Vaccination | Miyaoka (2019) |
| Sexual, Mosquito-Human | Unspecified | SIR-SI | Theoretical | Other | Angina (2022) |
| Human-Human, Mosquito-Human | Homogeneous mixing, Unspecified | SIR-SI | Theoretical | | Binder (2019) |
| Human-Human, Mosquito-Human | Unspecified | SIR-SI | Theoretical | Behaviour Changes, Insecticide-treated nets, Other, Pesticides/larvicides | Ali (2021) |
| Mosquito-Human, Unspecified | Other | SIR-SI | Theoretical | | Wang (2021) |
| Mosquito-Human | Homogeneous mixing | SIRS-SI | Theoretical | Other, Pesticides/larvicides | González-Parra (2020) |
| Sexual, Mosquito-Human | Homogeneous mixing | SIRS-SI | Fitted | Behaviour Changes, Insecticide-treated nets, Mosquito Control | Ukanwoke (2022) |
| Human-Human, Mosquito-Human | Other | SLIR-SLI | Theoretical | | Van Wyk (2023) |
| Mosquito-Human | Homogeneous mixing | SIR | Fitted | | O'driscoll (2021) |
| Mosquito-Human | Homogeneous mixing, Other | SIR | Fitted | | Ospina (2017) |
| Human-Human | Homogeneous mixing | SIR | Fitted | | Bastos (2018) |
| Human-Human | Homogeneous mixing | SIR | Fitted | | Angulo (2018) |
| Human-Human | | SIR | Fitted | | Harris (2019) |
| Sexual, Mosquito-Human | Homogeneous mixing | SIR | Theoretical | | Baca-Carrasco (2016) |
| Sexual, Mosquito-Human | Homogeneous mixing, Other | SIR | Theoretical | | Baca-Carrasco (2016) |
| Human-Human, Mosquito-Human | Homogeneous mixing | SIR | Theoretical | Behaviour Changes, Pesticides/larvicides, Mosquito Control | Anggriani (2023) |

continued on next page

continued from previous page

| Model Compartments | Transmission Route(s) | Model Assumptions | Fitted | Interventions | Source |
|---|---|---|---|---|---|
| Mosquito-Human | Latent and incubation periods are same | SEIR | Theoretical | | Zhao (2023) |
| Mosquito-Human | | SEIR | Fitted | | Netto (2017) |
| Sexual, Mosquito-Human | Heterogenity in transmission between human groups | SEIR | Fitted | | De Barros (2019) |
| Human-Human, Mosquito-Human | Homogeneous mixing, Other | SEIR | Theoretical | | Baca-Carrasco (2016) |
| Mosquito-Human | Other | Other SEIR-SEI | Theoretical | | Ren (2024) |
| Mosquito-Human | | Other SEIR-SEI | Theoretical | | Wang (2023) |
| Sexual, Mosquito-Human | Homogeneous mixing | Other SEIR-SEI | Fitted | Mosquito Control | Gao (2016) |
| Sexual, Mosquito-Human | Other | Other SEIR-SEI | Theoretical | Behaviour Changes, Insecticide-treated nets, Pesticides/larvicides, Mosquito Control | Chaikham (2017) |
| Sexual, Mosquito-Human | Unspecified | Other SEIR-SEI | Fitted | | Tonsing (2018) |
| Human-Human, Sexual, Mosquito-Human | Age | Other SEIR-SEI | Theoretical | Behaviour Changes, Insecticide-treated nets, Pesticides/larvicides, Treatment, Mosquito Control | Chaikham (2018) |
| Sexual, Mosquito-Human | Unspecified | SAIR-SEI | Theoretical | Behaviour Changes, Indoor residual spraying, Insecticide-treated nets, Treatment | Momoh (2018) |
| Mosquito-Human | Cross-immunity, Homogeneous mixing | Other | Theoretical | Behaviour Changes, Insecticide-treated nets, Other, Treatment, Mosquito Control | Bonyah (2019) |
| Mosquito-Human | Cross-immunity, Other | Other | Fitted | | Henderson (2021) |
| Mosquito-Human | Heterogenity in transmission between human and vector, Heterogenity in transmission between human groups | Other | Theoretical | | Muñoz (2017) |
| Mosquito-Human | Heterogenity in transmission between human groups, Other | Other | Fitted | | Liang (2019) |
| Mosquito-Human | Homogeneous mixing | Other | Fitted | | Aranda (2019) |
| Mosquito-Human | Homogeneous mixing | Other | Fitted | Other | Kumar (2023) |
| Mosquito-Human | Latent and incubation periods are same | Other | Theoretical | | Zhang (2022) |
| Mosquito-Human | Other | Other | Theoretical | | Fitzgibbon (2017) |
| Mosquito-Human | Other | Other | Theoretical | Behaviour Changes, Other, Pesticides/larvicides, Treatment, Mosquito Control | Djomegni (2021) |
| Mosquito-Human | Other | Other | Theoretical | Other, Vaccination | Kribs (2023) |
| Mosquito-Human | Other, Unspecified | Other | Theoretical | | Caminade (2017) |
| Mosquito-Human | Unspecified | Other | Theoretical | | Khan (2019b) |
| Mosquito-Human | Unspecified | Other | Theoretical | Behaviour Changes, Treatment, Mosquito Control | Manisha (2023) |
| Mosquito-Human | | Other | Fitted | | Villela (2017) |
| Mosquito-Human | | Other | Fitted | | O'Reilly (2018) |
| Mosquito-Human | | Other | Theoretical | | Han (2024) |
| Mosquito-Human | | Other | Theoretical | | Hajji (2024) |
| Mosquito-Human | | Other | Theoretical | | Wang (2024b) |
| Mosquito-Human | | Other | Theoretical | | Danbaba (2018) |
| Mosquito-Human | | Other | Fitted | | Omame (2023b) |
| Mosquito-Human | | Other | Theoretical | | Ndairou (2018) |
| Human-Human | Heterogenity in transmission between human groups | Other | Theoretical | Other, Quarantine | Scatà (2016) |
| Human-Human | Unspecified | Other | Fitted | | Massad (2017) |
| Sexual, Mosquito-Human | Heterogenity in transmission between human and vector | Other | Theoretical | Genetically modified mosquitoes, Treatment, Mosquito Control | Atokolo (2022) |
| Sexual, Mosquito-Human | Heterogenity in transmission between human and vector, Other | Other | Theoretical | | Begum (2021) |
| Sexual, Mosquito-Human | Heterogenity in transmission between human and vector, Other | Other | Theoretical | | Farman (2020) |
| Sexual, Mosquito-Human | Heterogenity in transmission between human and vector, Other | Other | Theoretical | | Rezapour (2020) |
| Sexual, Mosquito-Human | Heterogenity in transmission between human groups | Other | Theoretical | | Villela (2017) |
| Sexual, Mosquito-Human | Heterogenity in transmission between human groups, Homogeneous mixing | Other | Theoretical | | Cruz-Pacheco (2019) |
| Sexual, Mosquito-Human | Homogeneous mixing | Other | Theoretical | Behaviour Changes, Indoor residual spraying, Insecticide-treated nets, Other, Pesticides/larvicides, Mosquito Control | Wang (2019) |
| Sexual, Mosquito-Human | Homogeneous mixing | Other | Fitted | | Dénes (2019) |
| Sexual, Mosquito-Human | Homogeneous mixing | Other | Fitted | Pesticides/larvicides, Mosquito Control, Wolbachia replacement | Wang (2024a) |

continued on next page

continued from previous page

| Model Compartments | Transmission Route(s) | Model Assumptions | Fitted | Interventions | Source |
|---|---|---|---|---|---|
| Sexual, Mosquito-Human | Homogeneous mixing, Unspecified | Other | Fitted | | Li (2019) |
| Sexual, Mosquito-Human | Latent and incubation periods are same | Other | Fitted | Mechanical removal of breeding sites , Pesticides/larvicides | Agudelo (2022) |
| Sexual, Mosquito-Human | Other | Other | Fitted | Behaviour Changes | Luo (2021) |
| Sexual, Mosquito-Human | Unspecified | Other | Theoretical | | Brauer (2016) |
| Sexual, Mosquito-Human | Unspecified | Other | Theoretical | Behaviour Changes, Other, Treatment | Wattanasirikosone (2021) |
| Sexual, Mosquito-Human | Unspecified | Other | Fitted | Behaviour Changes, Other, Mosquito Control | Zhu (2022) |
| Sexual, Mosquito-Human | | Other | Theoretical | | Agusto (2017b) |
| Human-Human, Sexual, Mosquito-Human | Heterogenity in transmission between human and vector, Homogeneous mixing, Other | Other | Fitted | | Yuan (2021) |
| Human-Human, Sexual, Mosquito-Human | | Other | Fitted | Behaviour Changes, Mosquito Control | Huo (2023) |
| Human-Human, Mosquito-Human | Heterogenity in transmission between human groups, Homogeneous mixing | Other | Fitted | | Tuncer (2018) |
| Human-Human, Mosquito-Human | Homogeneous mixing, Other | Other | Fitted | Other | Sanchez (2019) |
| Mosquito-Human, Unspecified | Cross-immunity, Other | Other | Fitted | Vaccination | Omame (2023a) |
| Unspecified | Cross-immunity, Other | Other | Theoretical | Other | Deolia (2024) |
| Unspecified | Unspecified | Other | Fitted | | Zafar (2024) |
| Mosquito-Human | Homogeneous mixing | | Theoretical | | Amaku (2018) |
| Mosquito-Human | Unspecified | | Theoretical | | Maamar (2024) |
| Sexual | Cross-immunity | | Fitted | | Tang (2019) |
| Sexual, Mosquito-Human | Heterogenity in transmission between human groups, Other | | Theoretical | Other, Pesticides/larvicides | Olawoyin (2018) |
| Sexual, Mosquito-Human | | | Theoretical | Mosquito Control | Padmanabhan (2017a) |
| Human-Human, Mosquito-Human | Unspecified | | Theoretical | | Rezapour (2023) |
| **Compartmental - Stochastic** | | | | | |
| Mosquito-Human | Cross-immunity, Homogeneous mixing, Other | SEIR-SEI | Fitted | | Shutt (2017) |
| Mosquito-Human | Heterogenity in transmission between human and vector, Homogeneous mixing | SEIR-SEI | Fitted | | Lourenco (2017) |
| Mosquito-Human | Heterogenity in transmission between human groups, Other | SEIR-SEI | Fitted | | Perrotta (2022) |
| Mosquito-Human | Homogeneous mixing | SEIR-SEI | Fitted | | Morrison (2020) |
| Mosquito-Human | Homogeneous mixing | SEIR-SEI | Theoretical | Behaviour Changes, Vaccination, Mosquito Control | Mina (2020) |
| Mosquito-Human | Other | SEIR-SEI | Theoretical | | Tramonte (2019) |
| Mosquito-Human | Unspecified | SEIR-SEI | Theoretical | | Gokila (2021) |
| Sexual, Mosquito-Human | Heterogenity in transmission between human groups, Other | SEIR-SEI | Fitted | Vaccination | Durham (2018) |
| Sexual, Mosquito-Human | Other | SEIR-SEI | Theoretical | | He (2020) |
| Sexual, Mosquito-Human | Unspecified | SEIR-SEI | Theoretical | Insecticide-treated nets, Other, Treatment | Shao (2023) |
| Human-Human, Mosquito-Human | Other | SEIR-SEI | Theoretical | | Sun (2018) |
| Human-Human, Mosquito-Human | Unspecified | SEIR-SEI | Theoretical | | Bekiryazici (2022) |
| Mosquito-Human | Homogeneous mixing | SI-SI | Theoretical | | Maity (2024) |
| Human-Human, Mosquito-Human | Heterogenity in transmission between human groups | SI-SI | Theoretical | | Bekiryazici (2023) |
| Mosquito-Human | Homogeneous mixing, Latent and incubation periods are same | SIR | Fitted | | Perkins (2016) |
| Unspecified | Unspecified | SIR | Fitted | | Quandelacy (2021) |
| Mosquito-Human | Heterogenity in transmission over time, Other | SEIR | Fitted | | Nguyen-Van-Yen (2021) |
| Human-Human | | SEIR | Theoretical | | Wardle (2024) |
| Mosquito-Human | Cross-immunity, Homogeneous mixing | Other | Fitted | | Hirata (2023) |
| Mosquito-Human | Homogeneous mixing | Other | Theoretical | | Zhu (2024) |
| Sexual, Mosquito-Human | Other | Other | Theoretical | Wolbachia suppression | Xue (2021) |
| Human-Human, Mosquito-Human | Heterogenity in transmission between human and vector, Homogeneous mixing | Other | Theoretical | Treatment | Algehyne (2022) |
| Mosquito-Human | Cross-immunity, Heterogenity in transmission between human groups, Other | | Fitted | | Champagne (2016) |
| **Compartmental - Unspecified** | | | | | |
| Mosquito-Human | Heterogenity in transmission between human and vector, Heterogenity in transmission over time, Homogeneous mixing, Latent and incubation periods are same | SEIR-SEI | Fitted | Pesticides/larvicides, Mosquito Control | Suparit (2018) |
| Mosquito-Human | | SEIR-SEI | Theoretical | | Manore (2017) |
| Mosquito-Human | | SEIR-SEI | Fitted | Vaccination | Bartsch (2019) |

continued on next page

continued from previous page

| Model Compartments | Transmission Route(s) | Model Assumptions | Fitted | Interventions | Source |
|---|---|---|---|---|---|
| Human-Human, Mosquito-Human | Unspecified | SEIR-SEI | Fitted | | Ibrahim (2023) |
| Mosquito-Human | Heterogenity in transmission between human and vector, Other | SIR | Fitted | | Siraj (2017) |
| Mosquito-Human | Heterogenity in transmission between human groups, Homogeneous mixing, Other | Other | Fitted | Mosquito Control | Sasmal (2018) |
| Mosquito-Human | | Other | Fitted | | Fuller (2017) |
| Sexual, Mosquito-Human | Heterogenity in transmission between human groups, Other | Other | Theoretical | | Maxian (2017) |
| Sexual, Mosquito-Human | Homogeneous mixing | Other | Theoretical | Mosquito Control | Saad-Roy (2018) |
| Sexual, Mosquito-Human | Other | Other | Theoretical | Vaccination | Valega-Mackenzie (2018) |
| Sexual, Mosquito-Human | Other, Heterogenity in transmission between human groups | Other | Theoretical | | Saad-Roy (2016) |
| Human-Human, Mosquito-Human | | Other | Theoretical | Insecticide-treated nets, Treatment, Mosquito Control | Khan (2019a) |
| Mosquito-Human | Heterogenity in transmission between human and vector, Homogeneous mixing | | Fitted | Behaviour Changes, Treatment, Mosquito Control | Biswas (2024) |
| Sexual, Mosquito-Human | | | Fitted | | Srivastav (2019) |
| **Compartmental;Other - Deterministic** | | | | | |
| Mosquito-Human | Other | SIR | Fitted | | Claypool (2022) |
| **Compartmental;Other - Stochastic** | | | | | |
| Mosquito-Human | | SIR | Fitted | | Riou (2018) |
| **Compartmental;Other - Unspecified** | | | | | |
| Mosquito-Human | Unspecified | SEIR-SEI | Theoretical | | Sabir (2023) |
| Unspecified | Other | SIR | Fitted | | Rotejanaprasert (2019) |
| **Individual based - Stochastic** | | | | | |
| Sexual, Mosquito-Human | Age | SEIR-SEI | Theoretical | Behaviour Changes, Indoor residual spraying, Pesticides/larvicides, Mosquito Control | Moghadas (2017) |
| Sexual, Mosquito-Human | Heterogenity in transmission between human groups | SEIR-SEI | Theoretical | | Ferdousi (2019) |
| Unspecified | Age, Heterogenity in transmission between human groups | SIR | Fitted | Vaccination | Counotte (2019) |
| Mosquito-Human | Heterogenity in transmission between human groups, Other | SEIR | Theoretical | Behaviour Changes | Ajelli (2017) |
| **Individual based;Compartmental - Deterministic** | | | | | |
| Mosquito-Human | | SEIR-SEI | Fitted | Other, Mosquito Control | Gwalani (2018) |
| **Individual based;Compartmental - Stochastic** | | | | | |
| Mosquito-Human | | SEIR-SEI | Fitted | | Zhang (2017) |
| Mosquito-Human | Cross-immunity | SEIR | Fitted | | Pinotti (2024) |
| Mosquito-Human | | Other | Fitted | | Marini (2017) |
| **Other - Deterministic** | | | | | |
| Mosquito-Human | Homogeneous mixing | | Fitted | | Rocklov (2016) |
| Mosquito-Human | Unspecified | | Fitted | Other | Guzzetta (2016) |
| Mosquito-Human | | | Theoretical | | Chen (2024) |
| Human-Human | | | Fitted | | Ogden (2017) |
| Human-Human | | | Fitted | | Hsieh (2017) |
| Sexual, Mosquito-Human | | | Fitted | | Rojas (2016) |
| Unspecified | Unspecified | | Fitted | | Picinini Freitas (2024) |
| Mosquito-Human | Homogeneous mixing | Other | Theoretical | | Rocklov (2016) |
| Unspecified | Unspecified | | Theoretical | | Sabir (2024) |
| **Other - Stochastic** | | | | | |
| Mosquito-Human | Other | | Fitted | | Aguiar (2018) |
| Mosquito-Human | | | Fitted | | Nisar (2024) |
| Sexual | Heterogenity in transmission between human groups | | Theoretical | | Allard (2017a) |
| Sexual | Heterogenity in transmission between human groups | | Theoretical | | Allard (2017b) |
| Unspecified | Heterogenity in transmission over time, Other | | Fitted | | Charniga (2021b) |
| Unspecified | Other | | Fitted | | Gardner (2018) |
| **Other - Unspecified** | | | | | |
| Mosquito-Human | | | Fitted | | Solimini (2018) |
| Human-Human | | | Fitted | | Cousien (2019) |
| Unspecified | Other | | Fitted | | Bogoch (2016) |
| Unspecified | Unspecified | | Fitted | | Sebrango-Rodriguez (2017) |
| Unspecified | | | Fitted | | Watts (2017) |
| Unspecified | | | Fitted | | Fajardo (2016) |
| Unspecified | | | Fitted | | Li (2021) |
| Unspecified | | | Theoretical | | Farman (2022) |
| Unspecified | | | Fitted | | Carreto (2022) |
| Unspecified | | | Fitted | | Weinstein (2020) |
| Unspecified | | | Fitted | | Zhang (2021) |
| Sexual, Mosquito-Human | Heterogenity in transmission between human and vector | Other | Theoretical | | Wang (2022a) |

continued on next page

continued from previous page

| Model Compartments | Transmission Route(s) | Model Assumptions | Fitted | Interventions | Source |
|---|---|---|---|---|---|
| Unspecified | | Other | Theoretical | | Wang (2022b) |
| **Unspecified** | | | | | |
| Unspecified | | | Fitted | | Omame (2023b) |
| **Unspecified - Stochastic** | | | | | |
| Unspecified | Unspecified | Other | Theoretical | Mechanical removal of breeding sites , Other | Madewell (2019) |

Table B.7: Overview of extracted transmission models.

##### B.7.2 Outbreaks

| Outbreak location | Outbreak dates | Suspected cases | Confirmed cases | Mode of case detection | Female cases | Male cases | References |
|---|---|---|---|---|---|---|---|
| Entire Country | 26 Dec 2015 - Dec 2016 | 454 | 147 | Molecular | 115 | 32 | Ryan (2018) |
| Entire Country | 09 Aug 2015 - 02 Apr 2016 | | 65726 | Confirmed + Suspected | 44133 | 21592 | Pacheco (2020) |
| **Angola** | | | | | | | |
| | Dec 2016 | 54 | 4 | Molecular | 3 | 1 | Hill (2019) |
| **Brazil** | | | | | | | |
| Manaus | Nov 2015 - Sep 2018 | | 6987 | Unspecified | | | Giovanetti (2020) |
| Salvador, Bahia | May 2015 - Aug 2017 | 434 | 78 | Unspecified | | | Bandeira (2020) |
| Tocantins | Jul 2015 - Dec 2016 | 10775 | 1825 | Molecular | 1553 | 682 | Rodrigues (2020) |
| Rio De Janeiro | Jan 2015 - 15 Apr 2016 | 25213 | | Symptoms | | | Villela (2017) |
| Rio De Janeiro | Feb 2015 - May 2016 | | 1717 | Molecular | | | Fuller (2017) |
| Minas Gerais State | Dec 2015 - Jul 2017 | | 1723 | Molecular | 1578 | | Iani (2021) |
| Rio De Janeiro | 2015 - 2016 | 39331 | | Unspecified | | | Raymundo (2021) |
| **Cabo Verde** | | | | | | | |
| Santiago, Fogo, Maio, Boavista | 05 Oct 2015 - 29 May 2016 | 7580 | | Symptoms | | | Lourenco (2018) |
| Santiago | 05 Oct 2015 - 29 May 2016 | 4937 | | Symptoms | | | Lourenco (2018) |
| Fogo | 05 Oct 2015 - 29 May 2016 | 1458 | | Symptoms | | | Lourenco (2018) |
| **Chile** | | | | | | | |
| Easter Island | Jan 2014 - 30 May 2014 | 89 | 51 | Molecular | 33 | 18 | Tognarelli (2016) |
| **Colombia** | | | | | | | |
| | Aug 2015 - 2016 | 18364 | | Unspecified | | | Sasmal (2018) |
| Cúcuta | 22 Nov 2015 - 02 Apr 2016 | 4287 | | Symptoms | | | Sebrango-Rodriguez (2017) |
| Nieva | 22 Nov 2015 - 02 Apr 2016 | 1940 | | Symptoms | | | Sebrango-Rodriguez (2017) |
| Arauca | 2015 - 2017 | 788 | | Confirmed + Suspected | | | Charniga (2022) |
| Armenia | 2015 - 2017 | 189 | | Confirmed + Suspected | | | Charniga (2022) |
| Barranquilla | 2015 - 2017 | 4665 | | Confirmed + Suspected | | | Charniga (2022) |
| Bucaramanga | 2015 - 2017 | 4322 | | Confirmed + Suspected | | | Charniga (2022) |
| Cali | 2015 - 2017 | 16279 | | Confirmed + Suspected | | | Charniga (2022) |
| Cartagena | 2015 - 2017 | 1021 | | Confirmed + Suspected | | | Charniga (2022) |
| Cúcuta | 2015 - 2017 | 6485 | | Confirmed + Suspected | | | Charniga (2022) |
| Florencia | 2015 - 2017 | 663 | | Confirmed + Suspected | | | Charniga (2022) |
| Ibagué | 2015 - 2017 | 4076 | | Confirmed + Suspected | | | Charniga (2022) |
| Inírida | 2015 - 2017 | 12 | | Confirmed + Suspected | | | Charniga (2022) |
| Leticia | 2015 - 2017 | 278 | | Confirmed + Suspected | | | Charniga (2022) |
| Medellín | 2015 - 2017 | 549 | | Confirmed + Suspected | | | Charniga (2022) |
| Mitú | 2015 - 2017 | 17 | | Confirmed + Suspected | | | Charniga (2022) |
| Mocoa | 2015 - 2017 | 57 | | Confirmed + Suspected | | | Charniga (2022) |
| Montería | 2015 - 2017 | 1785 | | Confirmed + Suspected | | | Charniga (2022) |
| Neiva | 2015 - 2017 | 3409 | | Confirmed + Suspected | | | Charniga (2022) |
| Pereira | 2015 - 2017 | 463 | | Confirmed + Suspected | | | Charniga (2022) |
| Popayán | 2015 - 2017 | 51 | | Confirmed + Suspected | | | Charniga (2022) |
| Puerto Carreño | 2015 - 2017 | 17 | | Confirmed + Suspected | | | Charniga (2022) |
| Quibdó | 2015 - 2017 | 14 | | Confirmed + Suspected | | | Charniga (2022) |
| Riohacha | 2015 - 2017 | 279 | | Confirmed + Suspected | | | Charniga (2022) |
| San Andrés | 2015 - 2017 | 1109 | | Confirmed + Suspected | | | Charniga (2022) |
| San José Del Guaviare | 2015 - 2017 | 154 | | Confirmed + Suspected | | | Charniga (2022) |
| Santa Marta | 2015 - 2017 | 1913 | | Confirmed + Suspected | | | Charniga (2022) |
| Sincelejo | 2015 - 2017 | 856 | | Confirmed + Suspected | | | Charniga (2022) |
| Valledupar | 2015 - 2017 | 788 | | Confirmed + Suspected | | | Charniga (2022) |
| Villavicencio | 2015 - 2017 | 2377 | | Confirmed + Suspected | | | Charniga (2022) |
| Yopal | 2015 - 2017 | 2121 | | Confirmed + Suspected | | | Charniga (2022) |
| | 2015 - 2016 | 40741 | | Confirmed + Suspected | | | Flórez-Lozano (2020) |
| Girardot | 19 Sep 2015 - 22 Jan 2016 | 1936 | 32 | Molecular | 1138 | | Rojas (2016) |
| Bucaramanga | 13 Dec 2015 - 10 Sep 2016 | 3651 | | Symptoms | | | Sebrango-Rodriguez (2017) |
| Cali | 13 Dec 2015 - 10 Sep 2016 | 12220 | | Symptoms | | | Sebrango-Rodriguez (2017) |
| | 10 Oct 2015 - 24 Jun 2017 | 108087 | 9802 | Molecular | 70478 | 35977 | Méndez (2017) |
| San Andres | 06 Sep 2015 - 30 Jan 2016 | 928 | 52 | Molecular | 589 | | Rojas (2016) |

continued on next page

continued from previous page

| Outbreak location | Outbreak dates | Suspected cases | Confirmed cases | Mode of case detection | Female cases | Male cases | References |
|---|---|---|---|---|---|---|---|
| Barranquilla | 01 Oct 2015 - 31 Dec 2015 | | 1470 | Symptoms | 860 | 610 | Towers (2016) |
| 15 Unspecified Admin Regions | 01 Jan 2016 - 13 Mar 2016 | | 1593 | Unspecified | | | Rocklov (2016) |
| Antioquia: Bajo Cauca: Caceres | 01 Jan 2016 - 11 Apr 2016 | 45 | | Symptoms | | | Ospina (2017) |
| Antioquia: Bajo Cauca: Nechi | 01 Jan 2016 - 11 Apr 2016 | 29 | | Symptoms | | | Ospina (2017) |
| Antioquia: Bajo Cauca: Caucasia | 01 Jan 2016 - 11 Apr 2016 | 59 | | Symptoms | | | Ospina (2017) |
| Antioquia: Bajo Cauca: Zaragoza | 01 Jan 2016 - 11 Apr 2016 | 49 | | Symptoms | | | Ospina (2017) |
| Antioquia: Valle Aburra: Medellin | 01 Jan 2016 - 11 Apr 2016 | 351 | | Symptoms | | | Ospina (2017) |
| Antioquia: Valle Aburra: Bello | 01 Jan 2016 - 11 Apr 2016 | 32 | | Symptoms | | | Ospina (2017) |
| Antioquia: Valle Aburra: Envigado | 01 Jan 2016 - 11 Apr 2016 | 21 | | Symptoms | | | Ospina (2017) |
| Antioquia: Valle Aburra: Itagui | 01 Jan 2016 - 11 Apr 2016 | 40 | | Symptoms | | | Ospina (2017) |
| Antioquia: Valle Aburra: Uraba Chigorodo | 01 Jan 2016 - 11 Apr 2016 | 164 | | Symptoms | | | Ospina (2017) |
| Antioquia: Valle Aburra: San Pedro De Uraba | 01 Jan 2016 - 11 Apr 2016 | 23 | | Symptoms | | | Ospina (2017) |
| Antioquia: Valle Aburra: Carepa | 01 Jan 2016 - 11 Apr 2016 | 102 | | Symptoms | | | Ospina (2017) |
| Antioquia: Valle Aburra: Turbo | 01 Jan 2016 - 11 Apr 2016 | 228 | | Symptoms | | | Ospina (2017) |
| Antioquia: Valle Aburra: Necocli | 01 Jan 2016 - 11 Apr 2016 | 36 | | Symptoms | | | Ospina (2017) |
| Antioquia: Valle Aburra: Apartado | 01 Jan 2016 - 11 Apr 2016 | 315 | | Symptoms | | | Ospina (2017) |
| Antioquia: Valle Aburra: Mutata | 01 Jan 2016 - 11 Apr 2016 | 19 | | Symptoms | | | Ospina (2017) |
| Antioquia: Nordeste: Remedios | 01 Jan 2016 - 11 Apr 2016 | 18 | | Symptoms | | | Ospina (2017) |
| Antioquia: Magdalena Medio: Puerto Berrio | 01 Jan 2016 - 11 Apr 2016 | 61 | | Symptoms | | | Ospina (2017) |
| Antioquia: Magdalena Medio: Puerto Triunfo | 01 Jan 2016 - 11 Apr 2016 | 27 | | Symptoms | | | Ospina (2017) |
| Antioquia: Magdalena Medio: Occidente Sospetran | 01 Jan 2016 - 11 Apr 2016 | 17 | | Symptoms | | | Ospina (2017) |
| Antioquia: Magdalena Medio: Oriente Rionegro | 01 Jan 2016 - 11 Apr 2016 | 18 | | Symptoms | | | Ospina (2017) |
| Florencia | 01 Jan 2015 - 24 Nov 2018 | | 611 | Symptoms | | | Bonilla-Aldana (2020) |
| San Vicente Del Caguan | 01 Jan 2015 - 24 Nov 2018 | | 103 | Symptoms | | | Bonilla-Aldana (2020) |
| San Jose Del Fragua | 01 Jan 2015 - 24 Nov 2018 | | 83 | Symptoms | | | Bonilla-Aldana (2020) |
| El Doncello | 01 Jan 2015 - 24 Nov 2018 | | 75 | Symptoms | | | Bonilla-Aldana (2020) |
| El Paujil | 01 Jan 2015 - 24 Nov 2018 | | 65 | Symptoms | | | Bonilla-Aldana (2020) |
| Montanita | 01 Jan 2015 - 24 Nov 2018 | | 29 | Symptoms | | | Bonilla-Aldana (2020) |
| Belen De Los Andaquies | 01 Jan 2015 - 24 Nov 2018 | | 26 | Symptoms | | | Bonilla-Aldana (2020) |
| Solita | 01 Jan 2015 - 24 Nov 2018 | | 13 | Symptoms | | | Bonilla-Aldana (2020) |
| Cartagena Del Chaira | 01 Jan 2015 - 24 Nov 2018 | | 12 | Symptoms | | | Bonilla-Aldana (2020) |
| Morelia | 01 Jan 2015 - 24 Nov 2018 | | 9 | Symptoms | | | Bonilla-Aldana (2020) |
| Milan | 01 Jan 2015 - 24 Nov 2018 | | 7 | Symptoms | | | Bonilla-Aldana (2020) |
| Albania | 01 Jan 2015 - 24 Nov 2018 | | 5 | Symptoms | | | Bonilla-Aldana (2020) |
| Valparaiso | 01 Jan 2015 - 24 Nov 2018 | | 5 | Symptoms | | | Bonilla-Aldana (2020) |
| Solano | 01 Jan 2015 - 24 Nov 2018 | | 4 | Symptoms | | | Bonilla-Aldana (2020) |
| Curillo | 01 Jan 2015 - 24 Nov 2018 | | 2 | Symptoms | | | Bonilla-Aldana (2020) |
| **Curaçao** | | | | | | | |
| | 16 Dec 2015 - 26 Apr 2017 | 2820 | 781 | Molecular | 574 | 207 | Lim (2019) |
| **Dominican Republic** | | | | | | | |
| | 30 Jan 2016 - 27 Feb 2016 | 573 | | Unspecified | | | Rocklov (2016) |
| | 2015 - 2016 | 5161 | | Unspecified | | | Petrone (2021) |
| | 01 Jan 2016 - 31 Dec 2016 | 5235 | | Unspecified | | | Bowman (2018) |
| **Ecuador** | | | | | | | |
| Portoviejo, Manabi | 01 Jan 2016 - 31 Aug 2016 | 467 | 148 | Molecular | 107 | 41 | Fors (2018) |
| 2 Unspecified Admin Regions | 01 Jan 2016 - 13 Mar 2016 | 96 | | Unspecified | | | Rocklov (2016) |
| **El Salvador** | | | | | | | |
| 11 Unspecified Admin Regions | 01 Jan 2016 - 13 Mar 2016 | 5618 | | Unspecified | | | Rocklov (2016) |
| **Federated States of Micronesia** | | | | | | | |
| Yap Island | 01 Apr 2007 - 31 Jul 2007 | 185 | 108 | Confirmed + Suspected | 66 | | Duffy (2009) |
| **France** | | | | | | | |
| Austral Islands, French Polynesia | Oct 2013 - Mar 2014 | 1208 | | Unspecified | | | Riou (2017) |
| Mo'orea, French Polynesia | Oct 2013 - Mar 2014 | 1235 | | Unspecified | | | Riou (2017) |

continued on next page

continued from previous page

| Outbreak location | Outbreak dates | Suspected cases | Confirmed cases | Mode of case detection | Female cases | Male cases | References |
|---|---|---|---|---|---|---|---|
| Marquesas Islands, French Polynesia | Oct 2013 - Mar 2014 | 994 | | Unspecified | | | Riou (2017) |
| Sous-Le-Vent Islands, French Polynesia | Oct 2013 - Mar 2014 | 3912 | | Unspecified | | | Riou (2017) |
| Tahiti, French Polynesias | Oct 2013 - Mar 2014 | 21406 | | Unspecified | | | Riou (2017) |
| Tuamotus, French Polynesia | Oct 2013 - Mar 2014 | 1211 | | Unspecified | | | Riou (2017) |
| Guadeloupe | Jan 2016 - Oct 2016 | 30454 | | Unspecified | | | Riou (2017) |
| Martinique | Jan 2016 - Oct 2016 | 37295 | | Unspecified | | | Riou (2017) |
| Guadeloupe | Jan 2016 - 25 Feb 2016 | 389 | 35 | Molecular | | | Daudens-Vaysse (2016) |
| Saint-Martin | Feb 2016 - 25 Feb 2016 | 58 | 11 | Molecular | | | Daudens-Vaysse (2016) |
| French Guiana | Dec 2015 - 25 Feb 2016 | 1030 | | Unspecified | | | Daudens-Vaysse (2016) |
| Guadeloupe | 29 Jan 2016 - 10 Mar 2016 | 717 | | Unspecified | | | Rocklov (2016) |
| Martinique | 24 Nov 2015 - 25 Feb 2016 | 7600 | 203 | Unspecified | 142 | 61 | Daudens-Vaysse (2016) |
| Martinique | 21 Dec 2015 - 21 Jan 2016 | | 102 | Unspecified | | | Rocklov (2016) |
| Tahiti, French Polynesias | 11 Oct 2013 - 28 Mar 2014 | 4966 | 128 | Molecular | | | Kucharski (2016) |
| Sous-Le-Vent Islands, French Polynesia | 11 Oct 2013 - 28 Mar 2014 | 1131 | 166 | Molecular | | | Kucharski (2016) |
| Mo'orea, French Polynesia | 11 Oct 2013 - 28 Mar 2014 | 440 | 22 | Molecular | | | Kucharski (2016) |
| Tuamotu-Gambier, French Polynesia | 11 Oct 2013 - 28 Mar 2014 | 612 | 9 | Molecular | | | Kucharski (2016) |
| Marquesas Islands, French Polynesia | 11 Oct 2013 - 28 Mar 2014 | 455 | 21 | Molecular | | | Kucharski (2016) |
| Austral Islands, French Polynesia | 11 Oct 2013 - 28 Mar 2014 | 733 | 36 | Molecular | | | Kucharski (2016) |
| **France and the Netherlands** | | | | | | | |
| Saint Martin | Jan 2016 - Oct 2016 | 2519 | | Unspecified | | | Riou (2017) |
| **Guatemala** | | | | | | | |
| 8 Unspecified Admin Regions | 16 Nov 2015 - 15 Feb 2016 | | | Unspecified | | | Rocklov (2016) |
| **India** | | | | | | | |
| Kerala, Punjab, Rajasthan, Uttar Pradesh, Jharkhand | May 2021 - Oct 2021 | | 67 | Molecular | | | Yadav (2022) |
| Tamil Nadu* | Jun 2017 | | | Unspecified | | | Khan (2022) |
| Maharashtra* | 31 Jul 2021 | | | Unspecified | | | Khan (2022) |
| Madhya Pradesh* | 30 Oct 2018 | | | Unspecified | | | Khan (2022) |
| Uttar Pradesh* | 23 Oct 2021 | | | Unspecified | | | Khan (2022) |
| Rajasthan* | 21 Sep 2018 | | | Unspecified | | | Khan (2022) |
| Jaipur, Rajasthan | 2018 - 16 Nov 2018 | 1925 | 153 | Molecular | | | Sharma (2019) |
| Shastri Nagar And Surrounding Area Jaipur, | 2018 - 16 Nov 2018 | 1925 | 153 | Molecular | | | Sharma (2019) |
| Rajasthan | 15 Sep 2018 - 22 Nov 2018 | | 159 | Molecular | 104 | 55 | Malhotra (2020) |
| Kerala* | 08 Jul 2021 | | | Unspecified | | | Khan (2022) |
| **Jamaica** | | | | | | | |
| | 30 Jan 2016 - 30 Jul 2016 | 4567 | 72 | Unspecified | | | Webster-Kerr (2016) |
| **Mexico** | | | | | | | |
| Chiapas State | 30 Nov 2015 - 18 Dec 2015 | 119 | 25 | Molecular | | | Guerbois (2016) |
| 1 Unspecified Admin Region | 01 Jan 2016 - 13 Mar 2016 | | 129 | Unspecified | | | Rocklov (2016) |
| **Netherlands** | | | | | | | |
| Bonaire | 17 Oct 2016 - 26 Apr 2017 | 382 | 112 | Molecular | 81 | 31 | Lim (2019) |
| **Panama** | | | | | | | |
| 1 Unspecified Admin Region | 01 Jan 2016 - 13 Mar 2016 | | 49 | Unspecified | | | Rocklov (2016) |
| **Singapore** | | | | | | | |
| | 21 Aug 2016 - 30 Nov 2016 | | 455 | Molecular | 192 | 263 | Singapore Zika Study Group (2017) |
| **Suriname** | | | | | | | |
| | 02 Oct 2015 - 23 Aug 2016 | | 791 | Molecular | 553 | 238 | Codrington (2018) |
| Brokopondo | 02 Oct 2015 - 23 Aug 2016 | | 1 | Molecular | | | Codrington (2018) |
| Commewijne | 02 Oct 2015 - 23 Aug 2016 | | 19 | Molecular | | | Codrington (2018) |
| Coronie | 02 Oct 2015 - 23 Aug 2016 | | 5 | Molecular | | | Codrington (2018) |
| Marowoijne | 02 Oct 2015 - 23 Aug 2016 | | 4 | Molecular | | | Codrington (2018) |
| Nickerie | 02 Oct 2015 - 23 Aug 2016 | | 39 | Molecular | | | Codrington (2018) |
| Para | 02 Oct 2015 - 23 Aug 2016 | | 6 | Molecular | | | Codrington (2018) |
| Paramaribo | 02 Oct 2015 - 23 Aug 2016 | | 358 | Molecular | | | Codrington (2018) |
| Saramacca | 02 Oct 2015 - 23 Aug 2016 | | 11 | Molecular | | | Codrington (2018) |
| Sipaliwini | 02 Oct 2015 - 23 Aug 2016 | | 4 | Molecular | | | Codrington (2018) |
| Wanica | 02 Oct 2015 - 23 Aug 2016 | | 121 | Molecular | | | Codrington (2018) |
| **Thailand** | | | | | | | |
| | Jan 2016 - Dec 2017 | 1717 | 368 | Molecular | 224 | | Ruchusatsawat (2019) |
| Samut Songkhram | 2016 - 2018 | | 2300 | Symptoms | | | Sirinam (2022) |
| Ratchaburi, Bangkok | 2006 - 2009 | | 28 | Molecular | 15 | 13 | Sriburin (2021) |
| **United States** | | | | | | | |
| Puerto Rico | 26 Nov 2015 - 24 Feb 2016 | | 198 | Unspecified | | | Rocklov (2016) |

continued on next page

continued from previous page

| Outbreak location | Outbreak dates | Suspected cases | Confirmed cases | Mode of case detection | Female cases | Male cases | References |
|---|---|---|---|---|---|---|---|
| Wynwood Neighborhood, Miami-Dade County, Florida | 26 Jun 2016 - 05 Aug 2016 | | 21 | Unspecified | | | Marini (2017) |
| Hidalgo County, Texas* | 2016 - 2018 | | | Unspecified | | | Hinojosa (2020) |
| Miami-Dade | 07 Jul 2016 - 28 Dec 2016 | | 256 | Molecular | | | Philips (2019) |
| St. Croix, St. John, St. Thomas | 03 Jan 2016 - 24 Jan 2018 | | 1031 | Molecular | | 680 | Browne (2022) |
| St. Croix | 03 Jan 2016 - 24 Jan 2018 | | 257 | Molecular | | | Browne (2022) |
| St. John | 03 Jan 2016 - 24 Jan 2018 | | 89 | Molecular | | | Browne (2022) |
| St. Thomas | 03 Jan 2016 - 24 Jan 2018 | | 685 | Molecular | | | Browne (2022) |
| Puerto Rico | 01 Nov 2015 - 31 Dec 2016 | 39717 | 36390 | Unspecified | | | Sharp (2020) |
| New York City | 01 Jan 2016 - 30 Jun 2017 | 355 | 725 | Molecular | 519 | | Mcgibbon (2018) |
| Puerto Rico | 01 Jan 2015 - 24 Nov 2018 | | 21 | Symptoms | | | Bonilla-Aldana (2020) |
| **Vietnam** | | | | | | | |
| Southern Vietnam | 2016 - 2016 | 2190 | 214 | Molecular | | | Phan (2019) |

Table B.8: Overview of extracted outbreaks. Starred locations indicate outbreaks with reported dates but with no case numbers reported, which are included for completeness.

##### B.7.3 Delays

| Parameter value | Parameter type | Uncertainty | Disaggregation | Sample size | Location | Dates | Population sample type | Population group | Reference |
|---|---|---|---|---|---|---|---|---|---|
| **Admission To Critical Care/ICU To Discharge From Critical Care/Icu** | | | | | | | | | |
| 4 days | Median | Range: 1-30 | | 20 | VA Caribbean Healthcare System | 01 Dec 2015 - 31 Oct 2016 | Hospital | Other | Schirmer (2018) |
| **Admission To Discharge From Care** | | | | | | | | | |
| 4 days | Median | Range: 1-6 | | 3 | VA hospitals in US | 01 Dec 2015 - 31 Oct 2016 | Hospital | Other | Schirmer (2018) |
| 6 days | Median | Range: 1-214 | | 91 | VA Caribbean Healthcare System | 01 Dec 2015 - 31 Oct 2016 | Hospital | Other | Schirmer (2018) |
| 17 days | Median | | | 41 | Hospital (Brazil) | Nov 2015 - Apr 2016 | Hospital | Persons Under Investigation | Ferreira (2020) |
| 22 days | Median | Range: 3-102 | | 47 | Barranquilla (Colombia) | 01 Oct 2015 - 02 Apr 2016 | Hospital | Other | Salinas (2017) |
| 23 days | Median | IQR: 9.5-36 | | 6117 | Cucuta (Colombia) | 29 Jun 2015 - 30 Jul 2016 | Hospital | Persons Under Investigation | Anaya (2017) |
| **Diagnosis/Test Result To Death** | | | | | | | | | |
| 39 days | Median | Range: 3-104 | | 19 | VA Caribbean Healthcare System | 01 Dec 2015 - 31 Oct 2016 | Hospital | Other | Schirmer (2018) |
| **Diagnosis/Test Result To Onset Of Neurologic Symptoms** | | | | | | | | | |
| 7 days | Median | IQR: 2-14.5 | | 29 | Cucuta (Colombia) | 29 Jun 2015 - 30 Jul 2016 | Hospital | Other | Anaya (2017) |
| **Exposure/Infection To Recovery/Non-Infectiousness** | | | | | | | | | |
| 9 days | Median | IQR: 8-10 | | | Marseille, La Reunion (France) | Unspecified | Unspecified | Unspecified | Lequime (2020) |
| **Exposure/Infection To Recovery/Non-Infectiousness (Inverse Parameter)** | | | | | | | | | |
| 0.125 per day | Unspecified | | | | El Salvador | Oct 2015 - Apr 2016 | Population | General Population | Kumar (2017) |
| **Extrinsic Incubation Period** | | | | | | | | | |
| 6 days | Central | | | | Guangzhou (China) | Unspecified | | | Guo (2020) |
| 7.33 days | Mean | 95% CrI: 6-9 | | | Unspecified | Unspecified | Unspecified | Other | Armstrong (2020) |
| 8.311 days | Mean | 95% CrI: 4-16.962 | | | Suriname | 2015 - 22 Jun 2016 | Population | Persons Under Investigation | Shutt (2017) |
| 8.417 days | Mean | 95% CrI: 4-17.035 | | | El Salvador | 2015 - 22 Jun 2016 | Population | Persons Under Investigation | Shutt (2017) |
| 8.88 days | Mean | 95% CrI: 8-10 | | | Unspecified | Unspecified | Unspecified | Other | Armstrong (2020) |
| 10 days | Median | 95% CrI: 6.3-14 | | | French Polynesia (France) | Unspecified | Population | Persons Under Investigation | Kucharski (2016) |
| 10 days | Central | | | | Queensland (Australia) | Apr 2016 | | | Hugo (2019) |
| 10.5 days | Median | 95% CrI: 8.6-12.4 | | | Mo'orea, French Polynesia (France) | Oct 2013 - Apr 2014 | Population | Persons Under Investigation | Champagne (2016) |
| 10.5 days | Median | 95% CrI: 8.6-12.6 | | | Tahiti, French Polynesia (France) | Oct 2013 - Apr 2014 | Population | Persons Under Investigation | Champagne (2016) |
| 10.5 days | Median | 95% CrI: 8.5-12.5 | | | Tahiti, French Polynesia (France) | Oct 2013 - Apr 2014 | Population | Persons Under Investigation | Champagne (2016) |
| 10.6 days | Median | 95% CrI: 8.7-12.5 | | | Yap Island (Federated States of Micronesia) | 01 Apr 2007 - 29 Jul 2007 | Population | Persons Under Investigation | Champagne (2016) |
| 10.6 days | Median | 95% CrI: 8.6-12.6 | | | Mo'orea, French Polynesia (France) | Oct 2013 - Apr 2014 | Population | Persons Under Investigation | Champagne (2016) |

continued on next page

continued from previous page

| Parameter value | Parameter type | Uncertainty | Disaggregation | Sample size | Location | Dates | Population sample type | Population group | Reference |
|---|---|---|---|---|---|---|---|---|---|
| 10.7 days | Median | 95% CrI: 8.9-12.5 | | | New Caledonia (France) | 12 Nov 2013 - Aug 2014 | Population | Persons Under Investigation | Champagne (2016) |
| 10.7 days | Median | 95% CrI: 8.8-12.7 | | | Yap Island (Federated States of Micronesia) | 01 Apr 2007 - 29 Jul 2007 | Population | Persons Under Investigation | Champagne (2016) |
| 10.8 days | Median | 95% CrI: 8.9-12.8 | | | New Caledonia (France) | 12 Nov 2013 - Aug 2014 | Population | Persons Under Investigation | Champagne (2016) |
| 14 days | Central | | | | Queensland (Australia) | Apr 2016 | | | Hugo (2019) |
| 10 - 14 days | | | | | Unspecified | Unspecified | | | Ryckebusch (2017) |
| 5.1 - 24.2 days | | | Other | | Unspecified | Unspecified | | | Winokur (2020) |
| 17.8 - 24.2 days | Mean | | Level of Exposure | | Marseille, La Reunion (France) | Unspecified | Unspecified | Unspecified | Lequime (2020) |
| 16.2 - 18.2 days | | | Other | | Florida (United States) | Unspecified | | | Zimler (2021) |
| **Extrinsic Incubation Period (EIP)** | | | | | | | | | |
| 9.7 - 25.6 E10 | Central | | Other | | Unspecified | Unspecified | Other | Other | Blagrove (2020) |
| **Extrinsic Incubation Period (Inverse Parameter)** | | | | | | | | | |
| 0.1 per day | Unspecified | | | | Brazil | Jan 2016 - Dec 2016 | Population | General Population | Dantas (2018) |
| 0.004 | Maximum likelihood | Other: 0.001-0.014 | | | Colombia | 2015 - 2016 | Unspecified | Unspecified | Tonsing (2018) |
| **First Detection Of Anti-ZIKV IgM To Last Detection Of Anti-Zikv Igm** | | | | | | | | | |
| 124 days | Mean | 95% CI: 139-109 | | 59 | United States | 13 Jul 2016 - 19 Sep 2017 | Hospital | Persons Under Investigation | El Sahly (2019) |
| **First Infection In Country To First Reporting In Country** | | | | | | | | | |
| 12 - 35 weeks | Median | | | | Unspecified | Jan 2014 - Dec 2017 | Population | General Population | Yamamoto (2019) |
| **Generation Time** | | | | | | | | | |
| 2.5 weeks | Mean | SD: 0.7 | | | Guadeloupe, Martinique, Saint-Martin (France) | 2015 - 2017 | | | Riou (2018) |
| **Incubation Period** | | | | | | | | | |
| 4.9 days | Mean | 95% CI: 4.59-5.21 | | 111 | Jaipur, Rajasthan (India) | 22 Sep 2018 - 16 Nov 2018 | Population | Persons Under Investigation | Sharma (2019) |
| 5.563 days | Mean | 95% CrI: 2-15.54 | | 11825 | El Salvador | 2015 - 22 Jun 2016 | Population | Persons Under Investigation | Shutt (2017) |
| 5.8 days | Median | 95% CI: 5.6-6.15 | | 1946 | Feira de Santana (Brazil) | 01 Feb 2015 - 30 Apr 2017 | Population | General Population | Lourenco (2017) |
| 6 days | Mean | 95% CI: 5.2-6.8 | | 79 | Travellers ¡2 weeks travel (United States) | 01 Jan 2015 - 23 Jun 2016 | Travel | Persons Under Investigation | Krow-Lucal (2017) |
| 6 days | Median | Range: 2-10 | | 111 | Jaipur, Rajasthan (India) | 22 Sep 2018 - 16 Nov 2018 | Population | Persons Under Investigation | Sharma (2019) |
| 6.4 days | Mean | 95% CI: 5.7-7 | | 197 | All Travellers (United States) | 01 Jan 2015 - 23 Jun 2016 | Travel | Persons Under Investigation | Krow-Lucal (2017) |
| 6.6 days | Median | 95% CrI: 4.2-11 | | | French Polynesia (France) | 11 Oct 2013 - 28 Mar 2014 | Population | Persons Under Investigation | Kucharski (2016) |
| 6.614 days | Mean | 95% CrI: 2-16.814 | | 3042 | Suriname | 2015 - 22 Jun 2016 | Population | Persons Under Investigation | Shutt (2017) |
| **Incubation Period (Inverse Parameter)** | | | | | | | | | |
| 0.25 days | Central | 95% CI: 0.183-0.317 | | | Australes, French Polynesia (France) | Oct 2013 - Mar 2014 | Population | General Population | Rahman (2019) |

continued on next page

continued from previous page

| Parameter value | Parameter type | Uncertainty | Disaggregation | Sample size | Location | Dates | Population sample type | Population group | Reference |
|---|---|---|---|---|---|---|---|---|---|
| 0.225 days | Central | 95% CI: 0.161-0.29 | | | Tahiti, French Polynesia (France) | Oct 2013 - Mar 2014 | Population | General Population | Rahman (2019) |
| 0.086 days | Central | 95% CI: 0.071-0.102 | | | Tuamotu-Gambier, French Polynesia (France) | Oct 2013 - Mar 2014 | Population | General Population | Rahman (2019) |
| 0.084 days | Central | 95% CI: 0.059-0.108 | | | Mo'orea, French Polynesia (France) | Oct 2013 - Mar 2014 | Population | General Population | Rahman (2019) |
| 0.083 per day | Unspecified | | | | Brazil | Jan 2016 - Dec 2016 | Population | General Population | Dantas (2018) |
| 0.069 per day | Unspecified | | | | Colombia | 2016 - 2016 | Unspecified | Unspecified | Alzahrani (2021) |
| **Infectious Period** | | | | | | | | | |
| 4.8 days | Median | 95% CrI: 4.3-5.4 | | | Managua (Nicaragua) | 2016 - 2016 | Population | General Population | Counotte (2019) |
| 5.2 days | Median | 95% CrI: 4.1-6.7 | | | Tahiti, French Polynesia (France) | Oct 2013 - Apr 2014 | Population | Persons Under Investigation | Champagne (2016) |
| 5.2 days | Median | 95% CrI: 4.1-6.7 | | | Tahiti, French Polynesia (France) | Oct 2013 - Apr 2014 | Population | Persons Under Investigation | Champagne (2016) |
| 5.2 days | Median | 95% CrI: 4.1-6.7 | | | Yap Island (Federated States of Micronesia) | 01 Apr 2007 - 29 Jul 2007 | Population | Persons Under Investigation | Champagne (2016) |
| 5.2 days | Median | 95% CrI: 4.1-6.8 | | | Mo'orea, French Polynesia (France) | Oct 2013 - Apr 2014 | Population | Persons Under Investigation | Champagne (2016) |
| 5.3 days | Median | 95% CrI: 4.1-6.6 | | | Yap Island (Federated States of Micronesia) | 01 Apr 2007 - 29 Jul 2007 | Population | Persons Under Investigation | Champagne (2016) |
| 5.3 days | Median | 95% CrI: 4.1-6.7 | | | Mo'orea, French Polynesia (France) | Oct 2013 - Apr 2014 | Population | Persons Under Investigation | Champagne (2016) |
| 5.4 days | Median | 95% CrI: 4.1-6.8 | | | New Caledonia (France) | 12 Nov 2013 - Aug 2014 | Population | Persons Under Investigation | Champagne (2016) |
| 5.5 days | Median | 95% CrI: 4.2-6.8 | | | New Caledonia (France) | 12 Nov 2013 - Aug 2014 | Population | Persons Under Investigation | Champagne (2016) |
| 5.6 days | Median | 95% CrI: 4-10 | | | French Polynesia (France) | Unspecified | Population | Persons Under Investigation | Kucharski (2016) |
| 5.9 days | Median | 95% CI: 5.47-6.14 | | 1946 | Feira de Santana (Brazil) | 01 Feb 2015 - 30 Apr 2017 | Population | General Population | Lourenco (2017) |
| 9.073 days | Mean | HPDI 95%: 5.956-12.232 | | | Rio de Janeiro (Brazil) | 2015 - 2016 | Population | Other | Bastos (2018) |
| 9.207 days | Mean | HPDI 95%: 7.283-11.134 | | | Rio de Janeiro (Brazil) | 2015 - 2016 | Population | General Population | Bastos (2018) |
| 9.599 days | Mean | 95% CI: 7.673-11.701 | Sex | | Rio de Janeiro (Brazil) | 2015 - 2016 | Population | General Population | Bastos (2018) |
| 11.568 days | Mean | HPDI 95%: 9.246-13.963 | | | Rio de Janeiro (Brazil) | 2015 - 2016 | Population | General Population | Bastos (2018) |
| **Infectious Period (Inverse Parameter)** | | | | | | | | | |
| 0.17 per day | Median | 95% CrI: 0.1-0.23 | | | Rio de Janeiro (Brazil) | 2016 - 2016 | Population | Persons Under Investigation | De Barros (2019) |
| 0.083 days | Central | 95% CI: 0.06-0.107 | | | Sous-le-vent Islands, French Polynesia (France) | Oct 2013 - Mar 2014 | Population | General Population | Rahman (2019) |
| 0.083 days | Central | 95% CI: 0.026-0.14 | | | Marquesas Islands, French Polynesia (France) | Oct 2013 - Mar 2014 | Population | General Population | Rahman (2019) |
| 0.023 per day | Unspecified | | | | Colombia | 2016 - 2016 | Unspecified | Unspecified | Alzahrani (2021) |
| 0.023 per day | Unspecified | | | | Unspecified | Unspecified | Unspecified | Unspecified | Ali (2022a) |
| 0.02 - 0.1 per day | Unspecified | | Method;Other | | Florida (United States) | 19 Jul 2016 - 29 Sep 2016 | Population | Persons Under Investigation | Tuncer (2018) |
| 0.114 - 0.333 per day | Unspecified | | Method | | Brazil | Jan 2016 - Dec 2016 | Population | General Population | Dantas (2018) |
| **Latent Period (Inverse Parameter)** | | | | | | | | | |

continued on next page

continued from previous page

| Parameter value | Parameter type | Uncertainty | Disaggregation | Sample size | Location | Dates | Population sample type | Population group | Reference |
|---|---|---|---|---|---|---|---|---|---|
| 0.069 per day | Unspecified | | | | Colombia | Unspecified | Unspecified | Unspecified | Ali (2022a) |
| 0.04 | Maximum likelihood | Other: 0.02-0.07 | | | Colombia | 2015 - 2016 | Unspecified | Unspecified | Tonsing (2018) |
| **NAAT-Detectable Infection To Seroconversion (IgM)** | | | | | | | | | |
| 7.7 days | Mean | 95% CI: 6.1-9.2 | | 25 | Florida, Puerto Rico, Texas, Nevada (United States) | 06 Jul 2016 - 07 Mar 2017 | Population | Persons Under Investigation | Stone (2020) |
| **NAAT-Detectable Infection To Seroreversion (IgM)** | | | | | | | | | |
| 237 days | Mean | 95% CI: 128.7-459.5 | | 25 | Florida, Puerto Rico, Texas, Nevada (United States) | 06 Jul 2016 - 07 Mar 2017 | Population | Persons Under Investigation | Stone (2020) |
| **Onset Of Antecedent Symptoms To Onset Of Neurologic Symptoms** | | | | | | | | | |
| 6 days | Median | Range: 0-55 | | 47 | Barranquilla (Colombia) | 01 Oct 2015 - 02 Apr 2016 | Hospital | Other | Salinas (2017) |
| **Onset Of Neurologic Symptoms To Admission** | | | | | | | | | |
| 9 days | Median | Range: 0-56 | | 47 | Barranquilla (Colombia) | 01 Oct 2015 - 02 Apr 2016 | Hospital | Other | Salinas (2017) |
| **Onset Of Neurologic Symptoms To Nadir Of Neurologic Symptoms** | | | | | | | | | |
| 6 days | Median | IQR: 4-9 | | 50 | Salvador, Bahia (Brazil) | 01 Jan 2015 - 31 Aug 2015 | Hospital | Other | Styczynski (2017) |
| **Onset Of Previous Illness To Onset Of Neurologic Symptoms** | | | | | | | | | |
| 8 days | Median | IQR: 5-15 | | 50 | Salvador, Bahia (Brazil) | 01 Jan 2015 - 31 Aug 2015 | Hospital | Other | Styczynski (2017) |
| **Onset To Admission** | | | | | | | | | |
| 1 days | Median | IQR: 1-2 | | 1936 | Girardot (Colombia) | 19 Oct 2015 - 22 Jan 2016 | Hospital | Persons Under Investigation | Rojas (2016) |
| 2 days | Median | 95% CI: 0-7 | Age | 351 | Ponce, Puerto Rico (United States) | 2012 - 31 Dec 2016 | Hospital | Children | Read (2018) |
| 3.3 days | Mean | SD: 0.98 | | 467 | Portoviejo, Manabi (Ecuador) | 01 Jan 2016 - 31 Aug 2016 | Hospital | General Population | Fors (2018) |
| 4 days | Median | IQR: 1-16 | | 928 | San Andrés (Colombia) | 06 Sep 2015 - 30 Jan 2016 | Hospital | Persons Under Investigation | Rojas (2016) |
| 7 days | Median | IQR: 3-19 | | 19 | University Hospital Bulovka, Prague (Czech Republic) | Jan 2004 - Dec 2019 | Travel | Persons Under Investigation | Trojanek (2023) |
| 8 days | Median | IQR: 4-21 | | 335 | Cases acquired in travellers in Americas or Caribbean (Brazil) | 01 Mar 2016 - 31 Dec 2019 | Travel | Persons Under Investigation | Angelo (2020) |
| **Onset To Implementation Of Vector Control Measures** | | | | | | | | | |
| 13 days | Median | Range: 4-58 | | 625 | Mainland, travel-related (France) | 01 Jan 2016 - 15 Jul 2016 | Population | Persons Under Investigation | Septfons (2016) |
| **Onset To Negative PCR Test In Saliva** | | | | | | | | | |
| 3.1 days | Mean | SD: 1.5 | | 51 | Managua (Nicaragua) | 31 Aug 2016 - 21 Oct 2016 | Household | Household Contacts Of Survivors | Burger-Calderon (2018) |
| **Onset To Negative PCR Test In Serum/Plasma** | | | | | | | | | |
| 2.56 days | Mean | SD: 1.53 | | 25 | Managua (Nicaragua) | 31 Aug 2016 - 21 Oct 2016 | Household | Household Contacts Of Survivors | Burger-Calderon (2018) |

continued on next page

continued from previous page

| Parameter value | Parameter type | Uncertainty | Disaggregation | Sample size | Location | Dates | Population sample type | Population group | Reference |
|---|---|---|---|---|---|---|---|---|---|
| **Onset To Negative PCR Test In Urine** | | | | | | | | | |
| 5.94 days | Mean | SD: 3.8 | | 86 | Managua (Nicaragua) | 31 Aug 2016 - 21 Oct 2016 | Household | Household Contacts Of Survivors | Burger-Calderon (2018) |
| **Onset To Recovery/Non-Infectiousness** | | | | | | | | | |
| 4.5 days | Mean | Range: 1.8 | | 467 | Portoviejo, Manabi (Ecuador) | 01 Jan 2016 - 31 Aug 2016 | Hospital | General Population | Fors (2018) |
| 5 days | Median | IQR: 4-6 | | 19 | University Hospital Bulovka, Prague (Czech Republic) | Jan 2004 - Dec 2019 | Travel | Persons Under Investigation | Trojanek (2023) |
| **Onset To Return To Area With Active Vectors** | | | | | | | | | |
| 2 days | Median | Range: -7-10 | | 625 | Mainland, travel-related (France) | 01 Jan 2016 - 15 Jul 2016 | Population | Persons Under Investigation | Septfons (2016) |
| **Onset To Test Result** | | | | | | | | | |
| 3.5 days | Median | IQR: 3-5 | | 90 | Bangkok, Samut Prakan, Samut Sakhon, Ratchaburi, Chon Buri (Thailand) | Mar 2020 - Mar 2023 | Hospital | Persons Under Investigation | Khongwichit (2023) |
| 14 days | Other | | | 10 | Recife and Manaus (Brazil) | Unspecified | Hospital | Persons Under Investigation | Batto-Menezes (2019) |
| **Onset To ZIKV IgG Detection** | | | | | | | | | |
| 17 days | Median | IQR: 12-26 | | 30 | Padova and Pavia, travel-related (Italy) | Jan 2016 - Jan 2017 | Hospital | Persons Under Investigation | Barzon (2018) |
| **Onset To ZIKV IgM Detection** | | | | | | | | | |
| 8 days | Median | IQR: 5-15 | | 30 | Padova and Pavia, travel-related (Italy) | Jan 2016 - Jan 2017 | Hospital | Persons Under Investigation | Barzon (2018) |
| **Onset To ZIKV RNA Clearance From Plasma** | | | | | | | | | |
| 11.5 days | Median | IQR: 6-64 | | 30 | Padova and Pavia, travel-related (Italy) | Jan 2016 - Jan 2017 | Hospital | Persons Under Investigation | Barzon (2018) |
| **Onset To ZIKV RNA Clearance From Saliva** | | | | | | | | | |
| 14 days | Median | IQR: 8-31 | | 26 | Padova and Pavia, travel-related (Italy) | Jan 2016 - Jan 2017 | Hospital | Persons Under Investigation | Barzon (2018) |
| **Onset To ZIKV RNA Clearance From Semen** | | | | | | | | | |
| 25 days | Median | IQR: 14-29 | | 10 | Padova and Pavia, travel-related (Italy) | Jan 2016 - Jan 2017 | Hospital | Persons Under Investigation | Barzon (2018) |
| **Onset To ZIKV RNA Clearance From Urine** | | | | | | | | | |
| 24 days | Median | IQR: 17-34 | | 29 | Padova and Pavia, travel-related (Italy) | Jan 2016 - Jan 2017 | Hospital | Persons Under Investigation | Barzon (2018) |
| **Recovery/Non-Infectiousness To Susceptibility (Inverse Parameter)** | | | | | | | | | |
| 0.054 per day | Unspecified | | | | Colombia | 2016 - 2016 | Unspecified | Unspecified | Alzahrani (2021) |
| 0.054 per day | Unspecified | | | | Unspecified | Unspecified | Unspecified | Unspecified | Ali (2022a) |
| **Reporting To Intervention** | | | | | | | | | |
| 5 days | Median | Range: 2-38 | | 625 | Mainland, travel-related (France) | 01 Jan 2016 - 15 Jul 2016 | Population | Persons Under Investigation | Septfons (2016) |
| **Serial Interval** | | | | | | | | | |
| 7.4 days | Mean | 95% CI: 4.59-10.2 | | 10 | Singapore | Unspecified | Community | Persons Under Investigation | Singapore Zika Study Group (2017) |
| 2.2 - 4.7 weeks | Mean | | Region | | French Polynesia (France) | Oct 2013 - Oct 2016 | Population | Persons Under Investigation | Riou (2017) |

Table B.9: Overview of extracted delay parameters.

##### B.7.4 Seroprevalence

| Seroprev- alence type | Parameter value | Uncert- ainty | Disag- gregation | Num- erator | Denom- inator | Sample size | PRNT on ELISA + tests | Location | Dates | Population sample type | Population group | Reference |
|---|---|---|---|---|---|---|---|---|---|---|---|---|
| **Argentina** | | | | | | | | | | | | |
| IgM | | | | 5 | 73 | 104 | | | Apr 2016 - Mar 2017 | Other | Persons Under Investigation | Tellechea (2018) |
| **Austria** | | | | | | | | | | | | |
| IgG | 0.1 % | | | 1 | 1001 | 1001 | | Tyrol | Aug 2016 - Nov 2016 | Population | Blood Donors | Borena (2017) |
| IgM | 0.3 % | | | 3 | 1001 | 1001 | | Tyrol | Aug 2016 - Nov 2016 | Population | Blood Donors | Borena (2017) |
| PRNT | 0 % | | | 0 | 4 | 1001 | TRUE | Tyrol | Aug 2016 - Nov 2016 | Population | Blood Donors | Borena (2017) |
| **Bangladesh** | | | | | | | | | | | | |
| IgG | 17 % | | | 17 | 92 | 92 | | | 2014 - 2014 | Population | Persons Under Investigation | Geurtsvankessel (2018) |
| IgG | 28 % | | | 15 | 52 | 52 | | | 2015 - 2015 | Population | Persons Under Investigation | Geurtsvankessel (2018) |
| IgM | % | | | 1 | 418 | 418 | | | 2011 - 2015 | Population | Persons Under Investigation | Geurtsvankessel (2018) |
| IgM | % | | | 3 | 418 | 418 | | | 2011 - 2015 | Population | General Population | Geurtsvankessel (2018) |
| PRNT | 3.1 % | | Time | 12 | 16 | 418 | TRUE | | 2011 - 2015 | Population | General Population | Geurtsvankessel (2018) |
| PRNT | 4.3 % | | Time | 5 | 15 | 418 | TRUE | | 2011 - 2015 | Population | Persons Under Investigation | Geurtsvankessel (2018) |
| **Bolivia** | | | | | | | | | | | | |
| IgG | 1.9 % | | Age, Sex | 3 | 162 | 162 | | La Paz | Mar 2017 - Apr 2017 | Population | General Population | Villarroel (2018) |
| IgG | 2.6 % | | Age, Sex | 4 | 152 | 152 | | Cochabamba | Mar 2017 - Apr 2017 | Population | General Population | Villarroel (2018) |
| IgG | 9.7 % | | Age, Sex | 19 | 196 | 196 | | Tarija | Mar 2017 - Apr 2017 | Population | General Population | Villarroel (2018) |
| IgG | 61 % | | Age, Sex | 122 | 200 | 200 | | Santa Cruz | Mar 2017 - Apr 2017 | Population | General Population | Villarroel (2018) |
| IgG | 61.9 % | | Age, Sex | 65 | 105 | 105 | | Beni | Dec 2016 - Dec 2016 | Population | General Population | Villarroel (2018) |
| PRNT | 0 % | 95% CI: 0-0 | Age, Sex | 0 | 4 | 4 | | La Paz | Mar 2017 - Apr 2017 | Population | General Population | Villarroel (2018) |
| PRNT | 0 % | 95% CI: 0-0 | Age, Sex | 0 | 7 | 7 | | Cochabamba | Mar 2017 - Apr 2017 | Population | General Population | Villarroel (2018) |
| PRNT | 0.5 % | 95% CI: 0-1.5 | Age, Sex | 1 | 22 | 22 | | Tarija | Mar 2017 - Apr 2017 | Population | General Population | Villarroel (2018) |
| PRNT | 21.5 % | 95% CI: 16-27 | Age, Sex | 43 | 134 | 134 | | Santa Cruz | Mar 2017 - Apr 2017 | Population | General Population | Villarroel (2018) |
| PRNT | 39 % | 95% CI: 30-48 | Age, Sex | 41 | 70 | 70 | | Beni | Mar 2017 - Apr 2017 | Population | General Population | Villarroel (2018) |
| **Brazil** | | | | | | | | | | | | |
| HAI/HI | 27.7 - 36.6 % | | Age | | 298 | 298 | | Manaus (Amazonas state) | Jan 2014 - Dec 2015 | Other | Other | Salgado (2021) |
| HAI/HI | 52.6 % | | Age | 235 | 447 | 447 | | Manaus | Jan 2015 - Dec 2015 | Community | Other | Salgado (2023) |
| IgG | 0 % | | | 1 | 132 | 132 | | Recife | 2016 - 2017 | Population | Children | Alves (2020) |
| IgG | 0.55 % | | | 1 | 182 | 182 | | Rio Grande do Sul | 05 Dec 2016 - 06 Jan 2017 | Hospital | Blood Donors | Diefenbach (2019) |
| IgG | 0 - 3.4 % | | Age | | 135 | 135 | | Diamantina | 2018 - 2019 | Population | Pregnant Women | Santos (2023) |
| IgG | 3 % | | | 63 | 2749 | 2749 | | Rio de Janeiro | Jul 2018 - Oct 2018 | Population | General Population | Perisse (2020) |
| IgG | 4.15 % | | | 7 | 140 | 140 | | São Paulo | May 2016 - May 2018 | Population | Blood Donors | Lira (2022) |
| IgG | 6.8 % | | | 6 | 88 | 88 | | Maranhao state | 2016 - 2018 | Hospital | Persons Under Investigation | Ribeiro (2020) |

continued on next page

continued from previous page

| Seroprev- alence type | Parameter value | Uncert- ainty | Disag- gregation | Num- erator | Denom- inator | Sample size | PRNT on ELISA + tests | Location | Dates | Population sample type | Population group | Reference |
|---|---|---|---|---|---|---|---|---|---|---|---|---|
| IgG | 7 % | 95% CI: 4-10 | | | 249 | 249 | | Salvador, Bahia | Oct 2014 - Oct 2014 | Population | General Population | Rodriguez-Barraquer (2019) |
| IgG | 7.25 % | | | | 262 | 262 | | Recife | Jan 2016 - Dec 2016 | Population | Pregnant Women | Castanha (2019) |
| IgG | 8 % | 95% CI: 6-10 | | | 675 | 675 | | Salvador, Bahia | Mar 2015 - Mar 2015 | Population | General Population | Rodriguez-Barraquer (2019) |
| IgG | 8.3 % | | | 20 | 240 | 240 | | Pernambuco State | Dec 2015 - Jun 2017 | Hospital | Pregnant Women | Ximenes (2019) |
| IgG | 0 - 21.4 % | | | | | | | Ribeirão Preto, São Paulo State | Jul 2010 - May 2017 | Population | General Population | Slavov (2020) |
| IgG | 15 % | 95% CI: 4-29 | | | 751 | 751 | | Porto Velho | Jul 2016 - Jun 2019 | Other | Other | Botosso (2023) |
| IgG | 15 % | | Other, Time | 11 | 72 | 72 | | São Paulo state | Jan 2016 - Sep 2016 | Hospital | Persons Under Investigation | Tozetto-Mendoza (2019) |
| IgG | 17.3 % | | | 34 | 196 | 196 | | Maranhao state | Apr 2017 - Jun 2018 | Hospital | Pregnant Women | Branco (2021) |
| IgG | 19 % | | | 43 | 227 | 227 | | Salvador, Bahia | 01 Jan 2015 - 31 Jan 2017 | Hospital | Pregnant Women | Aromolaran (2022) |
| IgG | 19 % | IQR: 16-22 | | | 227 | 227 | | Salvador, Bahia | 01 Jan 2015 - 31 Jan 2017 | Hospital | Pregnant Women | Aromolaran (2022) |
| IgG | 22 % | 95% CI: 7-41 | | | 817 | 817 | | Belo Horizonte | Jul 2016 - Jun 2019 | Other | Other | Botosso (2023) |
| IgG | 29 % | 95% CI: 11-52 | | | 494 | 494 | | Salvador, Bahia | Jul 2016 - Jun 2019 | Other | Other | Botosso (2023) |
| IgG | 30 % | 95% CI: 17-46 | | | 345 | 345 | | Fortaleza | Jul 2016 - Jun 2019 | Other | Other | Botosso (2023) |
| IgG | 33.87 % | | | 63 | 186 | 186 | | Manaus | May 2021 - Oct 2021 | Population | Other | Neto (2024) |
| IgG | 35 % | 95% CI: 26-48 | Region | | 5108 | 5108 | | Belo Horizonte, Salvador, Laranjeiras, Fortaleza, Recife, Cuiaba and Campo Grande, Porto Velho | Jul 2016 - Jun 2019 | Other | Other | Botosso (2023) |
| IgG | 0.1 - 74.7 % | | Age, Method, Other, Sex | | 2070 | 2070 | | Recife | Aug 2018 - Feb 2019 | Household | General Population | Braga (2023) |
| IgG | 38.6 % | 95% CI: 31.3-47 | | 56 | 144 | 144 | | University Hospital Professor Edgard Santos (UHPES) in Salvador de Bahia | Aug 2017 - Feb 2018 | Hospital | Other | Moreira-Soto (2020) |
| IgG | 40 % | 95% CI: 30-72 | | | 1017 | 1017 | | Cuiaba and Campo Grande | Jul 2016 - Jun 2019 | Other | Other | Botosso (2023) |
| IgG | 43.95 % | | | 98 | 223 | 223 | | Salvador, Bahia | May 2021 - Oct 2021 | Population | Other | Neto (2024) |
| IgG | 46.7 % | | | 163 | 349 | 349 | | Rio de Janeiro | Jul 2017 - Dec 2017 | Hospital | Pregnant Women | Wittlin (2022) |
| IgG | 47 % | 95% CI: 25-69 | | | 1110 | 1110 | | Recife | Jul 2016 - Jun 2019 | Other | Other | Botosso (2023) |
| IgG | | | | 216 | 404 | 404 | | Juazeiro do Norte | Jun 2018 - Dec 2018 | Community | General Population | De Almeida Barreto (2020) |
| IgG | 15 - 94 % | | Time | | | | | | Jan 2016 - Sep 2016 | Hospital | Persons Under Investigation | Tozetto-Mendoza (2019) |
| IgG | 57 % | 95% CI: 35-74 | | | 574 | 574 | | Laranjeiras | Jul 2016 - Jun 2019 | Other | Other | Botosso (2023) |
| IgG | 57 % | | | 172 | 302 | 302 | | Salvador, Bahia | Sep 2017 - Nov 2017 | Hospital | Other | Bastos Filho (2023) |

continued on next page

continued from previous page

| Seroprev- alence type | Parameter value | Uncert- ainty | Disag- gregation | Num- erator | Denom- inator | Sample size | PRNT on ELISA + tests | Location | Dates | Population sample type | Population group | Reference |
|---|---|---|---|---|---|---|---|---|---|---|---|---|
| IgG | 59 % | 95% CI: 50.7-66.7 | | 85 | 144 | 144 | | University Hospital Professor Edgard Santos (UHPES) in Salvador de Bahia | Feb 2016 - May 2016 | Hospital | Other | Moreira-Soto (2020) |
| IgG | | | Age, Other | 393 | 642 | 642 | | Salvador, Bahia | Oct 2015 - Oct 2015 | Population | Other | Rodriguez-Barraquer (2019) |
| IgG | 61.3 % | 95% CI: 52.9-69.7 | | 81 | 132 | 132 | | Recife | 2016 - 2017 | Population | Pregnant Women | Alves (2020) |
| IgG | 63 % | 95% CI: 60-65 | Age, Region | | 1453 | 1453 | | Salvador, Bahia | Oct 2015 - Oct 2015 | Population | General Population | Rodriguez-Barraquer (2019) |
| IgG | 63.3 % | 95% CI: 59.4-66.8 | Other, Time | 401 | 633 | 633 | | Salvador, Bahia | 25 Nov 2015 - 28 May 2016 | Mixed | Mixed Groups | Netto (2017) |
| IgG | 64 % | IQR: 61-67 | | | 188 | 188 | | Salvador, Bahia | 01 Jan 2015 - 31 Jan 2017 | Hospital | Pregnant Women | Aromolaran (2022) |
| IgG | 68 % | | | 127 | 188 | 188 | | Salvador, Bahia | 01 Jan 2015 - 31 Jan 2017 | Hospital | Pregnant Women | Aromolaran (2022) |
| IgG | 72.3 % | | | 73 | 101 | 101 | | Salvador, Bahia | Jan 2016 - Dec 2016 | Population | Pregnant Women | Duarte (2020) |
| IgG | 74.7 % | | | 71 | 95 | 95 | | Salvador, Bahia | 18 Jan 2016 - 16 Dec 2016 | Hospital | Other | Oliveira (2020) |
| IgG | 80.6 % | 95% CI: 76.4-84.1 | Age, Sex | 323 | 401 | 425 | | Pernambuco State | Apr 2017 - Dec 2017 | Household | Mixed Groups | Magalhaes (2021) |
| IgG | 80.9 % | | | 17 | 21 | 21 | | Salvador, Bahia | 18 Jan 2016 - 16 Dec 2016 | Hospital | Other | Oliveira (2020) |
| IgG | 86.7 % | | | 78 | 90 | 90 | | Maranhao state | 2016 - 2018 | Hospital | Persons Under Investigation | Ribeiro (2020) |
| IgG | 88.6 % | | | 62 | 70 | 70 | | Bahia | Mar 2015 - Feb 2016 | Hospital | Children | Venturi (2019) |
| IgG | 96.7 % | | | 87 | 90 | 90 | | Bahia | Mar 2015 - Feb 2016 | Hospital | Pregnant Women | Venturi (2019) |
| IgG and IgM | | | | 54 | 214 | 219 | | Rio de Janeiro | Jan 2015 - Aug 2016 | Hospital | Pregnant Women | João (2018) |
| IgM | 0 % | | | 0 | 132 | 132 | | Recife | 2016 - 2017 | Population | Pregnant Women | Alves (2020) |
| IgM | 0 % | | | 0 | 223 | 223 | | Salvador, Bahia | May 2021 - Oct 2021 | Population | Other | Neto (2024) |
| IgM | 0 % | | | 0 | 186 | 186 | | Manaus | May 2021 - Oct 2021 | Population | Other | Neto (2024) |
| IgM | 1.3 % | | Other, Time | 8 | 633 | 633 | | Salvador, Bahia | 25 Nov 2015 - 28 May 2016 | Mixed | Mixed Groups | Netto (2017) |
| IgM | 1.7 % | | | 6 | 349 | 349 | | Rio de Janeiro | Jun 2017 - Dec 2017 | Hospital | Pregnant Women | Wittlin (2022) |
| IgM | | | | 9 | 404 | 404 | | Juazeiro do Norte | Jun 2018 - Dec 2018 | Community | General Population | De Almeida Barreto (2020) |
| IgM | 2.4 % | | | 1 | 41 | 1289 | | Fortaleza | Feb 2018 - Dec 2018 | Community | Other | Frota (2023) |
| IgM | 2.85 % | | | 4 | 140 | 140 | | São Paulo | May 2016 - May 2018 | Population | Blood Donors | Lira (2022) |
| IgM | 3 % | | | 6 | 196 | 196 | | Maranhao state | Apr 2017 - Jun 2018 | Hospital | Pregnant Women | Branco (2021) |
| IgM | 4.2 % | | | 4 | 95 | 95 | | Salvador, Bahia | 18 Jan 2016 - 16 Dec 2016 | Hospital | Other | Oliveira (2020) |
| IgM | 5.6 % | | | | 262 | 262 | | Recife | Jan 2016 - Dec 2016 | Population | Pregnant Women | Castanha (2019) |
| IgM | | | | 1 | 16 | 16 | | Northeast Brazil | Dec 2014 - Feb 2017 | Hospital | Persons Under Investigation | Leonhard (2021) |

continued on next page

continued from previous page

| Seroprev- alence type | Parameter value | Uncert- ainty | Disag- gregation | Num- erator | Denom- inator | Sample size | PRNT on ELISA + tests | Location | Dates | Population sample type | Population group | Reference |
|---|---|---|---|---|---|---|---|---|---|---|---|---|
| IgM | 7.1 % | | | 5 | 70 | 70 | | Bahia | Mar 2015 - Feb 2016 | Hospital | Children | Venturi (2019) |
| IgM | 10.2 % | | | 71 | 694 | 694 | | Pernambuco State | Dec 2015 - Jun 2017 | Hospital | Pregnant Women | Ximenes (2019) |
| IgM | | | | 3 | 15 | 15 | | Northeast Brazil | Dec 2014 - Feb 2017 | Hospital | Persons Under Investigation | Leonhard (2021) |
| IgM | | | | 9 | 43 | 43 | | Central-West | Jan 2017 - Apr 2019 | Hospital | Pregnant Women | Rosado (2023) |
| IgM | 23.8 % | | | 5 | 21 | 21 | | Salvador, Bahia | 18 Jan 2016 - 16 Dec 2016 | Hospital | Other | Oliveira (2020) |
| IgM | | | | 17 | 68 | 68 | | Northeast Brazil | Dec 2014 - Feb 2017 | Hospital | Persons Under Investigation | Leonhard (2021) |
| IgM | 36 % | | | 30 | 84 | 84 | | Salvador, Bahia | 01 Jan 2015 - 31 Aug 2015 | Hospital | Other | Styczynski (2017) |
| IgM | 37 % | | | 41 | 112 | 112 | | Rio de Janeiro | 2015 - 2016 | Population | Persons Under Investigation | Brasil (2020) |
| IgM | 41 % | | | 17 | 41 | 41 | | Salvador, Bahia | 01 Jan 2015 - 31 Aug 2015 | Hospital | Other | Styczynski (2017) |
| IgM | 44.4 % | | | 40 | 90 | 90 | | Bahia | Mar 2015 - Feb 2016 | Hospital | Pregnant Women | Venturi (2019) |
| IgM | 6 - 86 % | | Time | | | | | São Paulo state | Jan 2016 - Sep 2016 | Hospital | Persons Under Investigation | Tozetto-Mendoza (2019) |
| IgM | | | | 19 | 41 | 41 | | Hospital da Restauração, Recife, Pernambuco, | Nov 2015 - Apr 2016 | Hospital | Persons Under Investigation | Ferreira (2020) |
| IgM | 84.8 % | | | 39 | 48 | 48 | | Salvador, Bahia | Jan 2015 - Dec 2016 | Community | Pregnant Women | Aguilar Ticona (2021) |
| PRNT | 1 % | 95% CI: 0-4 | | | 101 | 101 | | Salvador, Bahia | Mar 2015 - Mar 2015 | Population | General Population | Rodriguez-Barraquer (2019) |
| PRNT | 0 - 16.9 % | | | | | | TRUE | Ribeirão Preto, São Paulo State | Jul 2010 - May 2017 | Population | General Population | Slavov (2020) |
| PRNT | | | | 5 | 43 | 43 | | Central-West | Jan 2017 - Apr 2019 | Hospital | Pregnant Women | Rosado (2023) |
| PRNT | 20.5 % | | | 49 | 239 | 239 | | | Jun 2011 - Mar 2012 | Other | Children | Zambrano (2021) |
| PRNT | 30 % | | | 24 | 85 | 85 | TRUE | University Hospital Professor Edgard Santos (UHPES) in Salvador de Bahia | Feb 2016 - May 2016 | Hospital | Other | Moreira-Soto (2020) |
| PRNT | 40.54 % | | | 15 | 37 | 262 | TRUE | Recife | Jan 2016 - Dec 2016 | Population | Pregnant Women | Castanha (2019) |
| PRNT | 42.5 % | | | 40 | 94 | 94 | | Maranhao state | 2016 - 2018 | Hospital | Persons Under Investigation | Ribeiro (2020) |
| PRNT | 55.8 % | | | 324 | 581 | 581 | | Pernambuco State | Dec 2015 - Jun 2017 | Hospital | Pregnant Women | Ximenes (2019) |
| PRNT | 61.2 % | | | 287 | 469 | 469 | | Salvador (Hospital Geral Roberto Santos) | Oct 2015 | Population | Pregnant Women | Nery (2021) |
| PRNT | 62.5 % | | Other, Time | 187 | 299 | 299 | | Salvador, Bahia | 25 Nov 2015 - 28 May 2016 | Mixed | Mixed Groups | Netto (2017) |
| PRNT | 65.7 % | 95% CI: 61-70.1 | Age, Sex | 272 | 414 | 425 | | Pernambuco State | Apr 2017 - Dec 2017 | Household | Mixed Groups | Magalhaes (2021) |
| PRNT | 76 % | | | 64 | 84 | 84 | | Salvador, Bahia | 01 Jan 2015 - 31 Aug 2015 | Hospital | Other | Styczynski (2017) |

continued on next page

continued from previous page

| Seroprev- alence type | Parameter value | Uncert- ainty | Disag- gregation | Num- erator | Denom- inator | Sample size | PRNT on ELISA + tests | Location | Dates | Population sample type | Population group | Reference |
|---|---|---|---|---|---|---|---|---|---|---|---|---|
| PRNT | 84.8 % | | | 39 | 46 | 70 | | Bahia | Mar 2015 - Feb 2016 | Hospital | Children | Venturi (2019) |
| PRNT | 88 % | | | 36 | 41 | 41 | | Salvador, Bahia | 01 Jan 2015 - 31 Aug 2015 | Hospital | Other | Styczynski (2017) |
| PRNT | 90 % | | | 81 | 90 | 90 | | Bahia | Mar 2015 - Feb 2016 | Hospital | Pregnant Women | Venturi (2019) |
| **Burkina Faso** | | | | | | | | | | | | |
| IgG | 45.7 % | 95% CI: 41.39-50.08 | | 229 | 501 | 501 | | Ouagadougou, Bobo-Dioulasso | 2020 - 2020 | Population | Blood Donors | Tinto (2022) |
| PRNT | 22.75 % | 95% CI: 19.29-26.62 | | 114 | 229 | 501 | TRUE | Ouagadougou, Bobo-Dioulasso | 2020 - 2020 | Population | Blood Donors | Tinto (2022) |
| **Cabo Verde** | | | | | | | | | | | | |
| IgG | 10.9 % | | | 47 | 431 | 431 | | Santiago Island: Praia | 24 Aug 2014 - 04 Nov 2014 | Population | General Population | Ward (2022) |
| IgG | | | | 311 | 1226 | 1226 | | | Oct 2015 - 2016 | Population | Persons Under Investigation | Faye (2020) |
| IgM | | | | 15 | 64 | 64 | | | 05 Oct 2015 - 29 May 2016 | Population | Persons Under Investigation | Lourenco (2018) |
| PRNT | | | | 311 | 311 | 1226 | TRUE | | Oct 2015 - 2016 | Population | Persons Under Investigation | Faye (2020) |
| **Cambodia** | | | | | | | | | | | | |
| PRNT | 4 | | | 2 | 50 | 770 | | Kampong Speu | Jul 2018 - Aug 2018 | Other | Children | Odio (2024) |
| **Cameroon** | | | | | | | | | | | | |
| PRNT | 2 % | | | 3 | 150 | 150 | | Maroua | Aug 2015 - Oct 2015 | Other | Blood Donors | Gake (2017) |
| PRNT | 2 | | | 3 | 149 | 149 | | Ngaoundere | Aug 2015 - Oct 2015 | Other | Blood Donors | Gake (2017) |
| PRNT | 3.3 | | | 9 | 272 | 272 | | Yaounde | Aug 2015 - Oct 2015 | Other | Blood Donors | Gake (2017) |
| PRNT | 4.8 % | | | 9 | 186 | 186 | | Garoua | Aug 2015 - Oct 2015 | Other | Blood Donors | Gake (2017) |
| PRNT | 7.6 | | | 12 | 157 | 157 | | Bertoua | Aug 2015 - Oct 2015 | Other | Blood Donors | Gake (2017) |
| PRNT | 10 | | | 17 | 170 | 170 | | Douala | Aug 2015 - Oct 2015 | Other | Blood Donors | Gake (2017) |
| **China** | | | | | | | | | | | | |
| IgG | 9.5 % | | | 26 | 273 | 273 | | Nanning City, Guanxi | Mar 2019 - Mar 2019 | Population | General Population | Zhou (2020) |
| IgM | 0 % | | | 0 | 150 | 150 | | Affiliated Hospital of Jining Medical University | Oct 2013 - Jun 2017 | Hospital | Persons Under Investigation | Hao (2019) |
| IgM | 1.8 % | | | 5 | 273 | 273 | | Nanning City, Guanxi | Mar 2019 - Mar 2019 | Population | General Population | Zhou (2020) |
| PRNT | 65.4 % | | | 17 | 26 | 273 | TRUE | Nanning City, Guanxi | Mar 2019 - Mar 2019 | Population | General Population | Zhou (2020) |
| **Colombia** | | | | | | | | | | | | |
| IgG | 0 - 5.35 % | 95% CI: 0-16.74 | Time | | 390 | 390 | | Barranquilla | 06 May 2015 - 31 Aug 2015 | Population | General Population | Bayona-Pacheco (2019) |
| IgG | 64.9 % | | Method | 213 | 328 | 328 | | Santander | Unspecified | School | Mixed Groups | Cardenas (2020) |
| IgG | 86.8 % | | | 99 | 114 | 115 | | Risaralda | 22 Nov 2017 - 05 Jun 2019 | Hospital | Pregnant Women | Cardona-Ospina (2022) |
| IgG | 89 % | | | 80 | 90 | 90 | | Cereté: Córdoba state | 01 May 2016 - 31 May 2016 | Hospital | Pregnant Women | Marbán-Castro (2020) |

continued on next page

continued from previous page

| Seroprev- alence type | Parameter value | Uncert- ainty | Disag- gregation | Num- erator | Denom- inator | Sample size | PRNT on ELISA + tests | Location | Dates | Population sample type | Population group | Reference |
|---|---|---|---|---|---|---|---|---|---|---|---|---|
| IgG | 100 % | | | 29 | 29 | 29 | | Cucuta | 29 Jun 2015 - 30 Jul 2016 | Hospital | Other | Anaya (2017) |
| IgM | 0.8 % | | | 9 | 1080 | 1108 | | | Feb 2017 - Mar 2019 | Population | Children | Tannis (2024) |
| IgM | 1 % | | | 1 | 90 | 90 | | Cereté: Córdoba state | 01 May 2016 - 31 May 2016 | Hospital | Pregnant Women | Marbán-Castro (2020) |
| IgM | 4.3 % | | | 1519 | 66 | 1519 | | | Feb 2017 - Mar 2019 | Population | Pregnant Women | Tannis (2024) |
| IgM | 7 % | | | 22 | 301 | 301 | | | 01 Dec 2015 - 31 Dec 2019 | Hospital | Children | Castro-Trujillo (2024) |
| NS1 BOB ELISA | 1 % | | | | 703 | 703 | | | 2013 - 2014 | Population | General Population | Nascimento (2019) |
| NS1 BOB ELISA | 32 % | | | | 703 | 703 | | | 2017 - 2018 | Population | General Population | Nascimento (2019) |
| PRNT | 2.63 % | | | 3 | 114 | 115 | TRUE | Risaralda | 22 Nov 2016 - 05 Jun 2019 | Other | Pregnant Women | Cardona-Ospina (2022) |
| PRNT | 3 % | | | | 703 | 703 | TRUE | | 2013 - 2014 | Population | General Population | Nascimento (2019) |
| PRNT | 16 % | | | | 703 | 703 | TRUE | | 2017 - 2018 | Population | General Population | Nascimento (2019) |
| PRNT | 47.6 % | | | 339 | 712 | 712 | | | Jun 2011 - Mar 2012 | Other | Children | Zambrano (2021) |
| **Czech Republic** | | | | | | | | | | | | |
| IgG | 42.1 % | | | 8 | 19 | 19 | | University Hospital Bulovka, Prague | Jan 2004 - Dec 2019 | Travel | Persons Under Investigation | Trojanek (2023) |
| IgM | 63.1 % | | | 12 | 19 | 19 | | University Hospital Bulovka, Prague | Jan 2004 - Dec 2019 | Travel | Persons Under Investigation | Trojanek (2023) |
| PRNT | 84.2 % | | | 16 | 19 | 19 | | University Hospital Bulovka, Prague | Jan 2004 - Dec 2019 | Travel | Persons Under Investigation | Trojanek (2023) |
| **Dominican Republic** | | | | | | | | | | | | |
| IgM | 3.2 % | | | 6 | 189 | 189 | | | 2016 | Travel | Other | Voss (2020) |
| PRNT | 3.2 % | | | 6 | 6 | 189 | TRUE | | 2016 | Travel | Other | Voss (2020) |
| **DRC** | | | | | | | | | | | | |
| IgG | 3.5 % | | | 34 | 978 | 978 | | | 2013 - 2014 | Population | Children | Wilcox (2018) |
| PRNT | 0.1 % | 95% CI: 0-0.5 | | 1 | 31 | | TRUE | | 2013 - 2014 | Population | Children | Wilcox (2018) |
| **Ecuador** | | | | | | | | | | | | |
| NS1 BOB ELISA | 29 % | 95% CI: 25-33 | Age, Region, Sex | | 1126 | 1126 | | Borbón, Maldonado, Timbiré, Santa Maria, Santo Domingo, Colon Eloy | Aug 2019 - Oct 2019 | Community | General Population | Andrade (2024) |
| NS1 BOB ELISA | 42 % | 95% CI: 39-45 | Age, Other, Region, Sex | | 1192 | 1192 | | Borbón, Maldonado, Timbiré, Santa Maria, Santo Domingo, Colon Eloy | Jul 2018 - Oct 2018 | Community | General Population | Andrade (2024) |
| **El Salvador** | | | | | | | | | | | | |
| IgM | 22.7 % | | | 45 | 198 | 198 | | Sonsonate | Mar 2022 - Sep 2022 | Population | Pregnant Women | Lynn (2024) |
| **Ethiopia** | | | | | | | | | | | | |

continued on next page

continued from previous page

| Seroprev- alence type | Parameter value | Uncert- ainty | Disag- gregation | Num- erator | Denom- inator | Sample size | PRNT on ELISA + tests | Location | Dates | Population sample type | Population group | Reference |
|---|---|---|---|---|---|---|---|---|---|---|---|---|
| IgG | | | | 65 | 1645 | 1645 | | | May 2014 - Jul 2014 | Community | General Population | Tsegaye (2018) |
| IgG | 27.3 % | 95% CI: 20.7-35.1 | | 41 | 150 | 150 | | Itang special district, Lare district | Oct 2018 - Jun 2019 | Community | General Population | Asebe (2021) |
| PRNT | 0.4 % | | Region | 7 | 65 | 1645 | TRUE | | May 2014 - Jul 2014 | Community | General Population | Tsegaye (2018) |
| **Federated States of Micronesia** | | | | | | | | | | | | |
| IgM | 74 % | | Sex | 414 | 557 | 557 | | Yap Island | 01 Apr 2007 - 31 Jul 2007 | Household | General Population | Duffy (2009) |
| **Fiji** | | | | | | | | | | | | |
| MIA | 6.7 % | 95% CI: 4.5-9.4 | | 30 | 451 | 778 | | Central division | Sep 2013 - Nov 2013 | Population | Persons Under Investigation | Kama (2019) |
| MIA | 9 % | 95% CI: 5.8-13 | | 24 | 268 | 778 | | Western division | Sep 2013 - Nov 2013 | Population | Persons Under Investigation | Kama (2019) |
| MIA | 11.9 % | 95% CI: 4.9-22.9 | | 7 | 59 | 778 | | Northern division | Sep 2013 - Nov 2013 | Population | Persons Under Investigation | Kama (2019) |
| MIA | 6.1 - 25.2 % | | Age, Sex, Time | | | | | | Sep 2013 - Nov 2015 | Population | Persons Under Investigation | Kama (2019) |
| MIA | 21.9 % | 95% CI: 17.9-26.8 | | 73 | 333 | 333 | | Central division | Oct 2015 - Nov 2015 | Population | Persons Under Investigation | Kama (2019) |
| **France** | | | | | | | | | | | | |
| IgG | 0.8 % | | Level of Exposure | 5 | 593 | 593 | | French Polynesia | Jul 2011 - Oct 2013 | Population | Mixed Groups | Aubry (2015) |
| IgG | 22 % | 95% CI: 16-28 | | 154 | 700 | 700 | | Society Islands, French Polynesia | Sep 2015 - Nov 2015 | Population | General Population | Aubry (2017) |
| IgG | 26 % | | | 25 | 98 | 98 | | Papeete, Tahiti, French Polynesia | Nov 2013 - Feb 2014 | Hospital | General Population | Cao-Lormeau (2016) |
| IgG | | | | 51 | 179 | 179 | | Guadeloupe | Apr 2016 - Mar 2017 | Population | Pregnant Women | Prisant (2019) |
| IgG | 37 % | 95% CI: 26-47 | | 18 | 49 | 49 | | Society Islands, French Polynesia | Feb 2014 - Mar 2014 | Population | General Population | Aubry (2017) |
| IgG | 45 % | 95% CI: 38-52 | | 22 | 49 | 49 | | Tuamotu Islands, French Polynesia | Feb 2014 - Mar 2014 | Population | General Population | Aubry (2017) |
| IgG | 57 % | 95% CI: 47-68 | | 28 | 49 | 49 | | Marquesas Islands, French Polynesia | Feb 2014 - Mar 2014 | Population | General Population | Aubry (2017) |
| IgG | 59 % | 95% CI: 39-80 | | 29 | 49 | 49 | | Austral–Gambier Islands, French Polynesia | Feb 2014 - Mar 2014 | Population | General Population | Aubry (2017) |
| IgG | 66 % | 95% CI: 60-71 | | 312 | 476 | 476 | | Society Islands, French Polynesia | May 2014 - Jun 2014 | School | Children | Aubry (2017) |
| IgM | 17 % | | | 17 | 98 | 98 | | Papeete, Tahiti, French Polynesia | Nov 2013 - Feb 2014 | Hospital | General Population | Cao-Lormeau (2016) |
| IgM | 23.3 % | 95% CI: 0-62 | Region | | 2697 | 2697 | | 22 munici- palities of French Guiana | Jun 2017 - Oct 2017 | Population | General Population | Flamand (2019) |
| IgM | 26.1 % | | | 76 | 291 | 291 | | French Guiana | 01 Jan 2015 - 15 Jul 2016 | Mixed | Children | Pomar (2020) |
| MIA | 0 - 45.6 % | | Region | | 2697 | 2697 | | French Guiana | Jun 2017 - Oct 2017 | Population | General Population | Bailly (2021) |

continued on next page

continued from previous page

| Seroprev- alence type | Parameter value | Uncert- ainty | Disag- gregation | Num- erator | Denom- inator | Sample size | PRNT on ELISA + tests | Location | Dates | Population sample type | Population group | Reference |
|---|---|---|---|---|---|---|---|---|---|---|---|---|
| PRNT | 56 % | | | 54 | 98 | 98 | | Papeete, Tahiti, French Polynesia | Nov 2013 - Feb 2014 | Hospital | General Population | Cao-Lormeau (2016) |
| PRNT | 76 % | | | 78 | 102 | 102 | | Tahiti, Mo'orea, French Polynesia | 01 Jun 2013 - 31 Aug 2014 | Hospital | Pregnant Women | Subissi (2018) |
| PRNT | 95 % | | | 20 | 21 | 21 | | Tahiti, Mo'orea, French Polynesia | 01 Jun 2013 - 31 Aug 2014 | Hospital | Pregnant Women | Subissi (2018) |
| **Gabon** | | | | | | | | | | | | |
| IgG | 29.1 - 55 % | | Age, Sex, Time | | 462 | 462 | | Lambarene | Nov 2014 - Jan 2017 | Population | General Population | Ushijima (2021) |
| **Germany** | | | | | | | | | | | | |
| IgG | 1.2 % | | | 1 | 81 | 81 | | Germany travelling to Asia | Jan 2017 - Jan 2018 | Travel | Persons Under Investigation | Dammermann (2023) |
| IgM | 1.2 % | | | 1 | 81 | 81 | | Germany travelling to Asia | Jan 2017 - Jan 2018 | Travel | Persons Under Investigation | Dammermann (2023) |
| **Ghana** | | | | | | | | | | | | |
| IgG | | | Age, Sex | 3 | 160 | 160 | | Greater Accra Regional Hospital | Dec 2016 - Nov 2017 | Hospital | | Ankrah (2019) |
| IgM | | | Age, Sex | 30 | 160 | 160 | | Greater Accra Regional Hospital | Dec 2016 - Nov 2017 | Hospital | | Ankrah (2019) |
| **Guatemala** | | | | | | | | | | | | |
| NS1 BOB ELISA | 10 % | | | 20 | 196 | 196 | | Coastal lowlands of SW Guatemala | Oct 2015 - Nov 2015 | Population | Children | Lamb (2022) |
| NS1 BOB ELISA | 37 % | | | 69 | 186 | 186 | | Coastal lowlands of SW Guatemala | Jan 2016 - Feb 2016 | Population | Children | Lamb (2022) |
| PRNT | 0 - 14 % | | Method | | 196 | 196 | | Coastal lowlands of SW Guatemala | Oct 2015 - Nov 2015 | Population | Children | Lamb (2022) |
| PRNT | 19 - 40 % | | Method | | 186 | 186 | | Coastal lowlands of SW Guatemala | Jan 2016 - Feb 2016 | Population | Children | Lamb (2022) |
| **Guinea** | | | | | | | | | | | | |
| IgM | 14.7 % | 95% CI: 8.8-22.4 | Time | 17 | 116 | 116 | | Faranah Prefecture Hospital | May 2018 - Jul 2021 | Hospital | Persons Under Investigation | Bayandin (2023) |
| **Guyana** | | | | | | | | | | | | |
| IgG | 9.82 % | | | | | | | Cayenne, Kourou, Saint Laurent | Jan 2016 - Dec 2016 | Hospital | Pregnant Women | Hallet (2020) |
| IgG | | | Other | 837 | 6654 | 6654 | | Cayenne, Kourou, Saint Laurent | Jan 2016 - Dec 2016 | Hospital | Pregnant Women | Hallet (2020) |
| IgG | 16.5 % | | | | | | | Cayenne, Kourou, Saint Laurent | Jan 2016 - Dec 2016 | Hospital | Pregnant Women | Hallet (2020) |
| **Honduras** | | | | | | | | | | | | |
| IgM | 1.7 - 66.3 % | | Method | | 60 | 60 | | Tegucigalpa | Jul 2016 - Dec 2016 | Hospital | Children | Alger (2024) |

continued on next page

continued from previous page

| Seroprev- alence type | Parameter value | Uncert- ainty | Disag- gregation | Num- erator | Denom- inator | Sample size | PRNT on ELISA + tests | Location | Dates | Population sample type | Population group | Reference |
|---|---|---|---|---|---|---|---|---|---|---|---|---|
| NS1 BOB ELISA | 1 % | | | | 223 | 223 | | | 2013 - 2014 | Population | General Population | Nascimento (2019) |
| NS1 BOB ELISA | 57 % | | | | 223 | 223 | | | 2017 - 2018 | Population | General Population | Nascimento (2019) |
| PRNT | 2 % | | | | 223 | 223 | TRUE | | 2013 - 2014 | Population | General Population | Nascimento (2019) |
| PRNT | 23 % | | | | 223 | 223 | TRUE | | 2017 - 2018 | Population | General Population | Nascimento (2019) |
| PRNT | 79.3 % | | | 180 | 227 | 227 | | | Jun 2011 - Mar 2012 | Other | Children | Zambrano (2021) |
| **Hungary** | | | | | | | | | | | | |
| IFA | | | | 1 | 219 | 219 | | | 2017 - 2017 | Travel | General Population | Nagy (2019) |
| IFA | | | | 1 | 188 | 188 | | | 2018 - 2018 | Travel | General Population | Nagy (2019) |
| IFA | | | | 5 | 196 | 196 | | | 2016 - 2016 | Travel | General Population | Nagy (2019) |
| **India** | | | | | | | | | | | | |
| IgG | 6.66 % | | | 5 | 75 | 75 | | Bhopal region | Unspecified | Unspecified | Blood Donors | Nema (2024) |
| IgG | 15.38 % | | | 10 | 75 | 75 | | Bhopal region | Unspecified | Unspecified | Blood Donors | Nema (2024) |
| PRNT | | | | 4 | 29 | 29 | | | May 2021 - Oct 2021 | Unspecified | Persons Under Investigation | Yadav (2022) |
| **Indonesia** | | | | | | | | | | | | |
| IgM | 0 | | | 0 | 45 | 45 | | | Oct 2016 - Apr 2017 | Population | Children | Putri (2020) |
| IgM | 0 | | | 0 | 113 | 113 | | | Oct 2016 - Apr 2017 | Population | Children | Putri (2020) |
| PRNT | 2 | | | 1 | 46 | 46 | | | Oct 2016 - Apr 2017 | Population | Children | Putri (2020) |
| PRNT | 2 | | | 2 | 110 | 110 | | | Oct 2016 - Apr 2017 | Population | Children | Putri (2020) |
| PRNT | 9.1 % | 95% CI: 3.95-11.01 | Other, Region | 60 | 662 | 662 | | Aceh, North Sumatra, West Sumatra, Jambi, Lampung, Banten, DKI Jakarta, West Java, Central Java, East Java, Bali, East Kalimantan, South Sulawesi, Southeast Sulawesi | Oct 2014 - Nov 2014 | Population | Children | Tedjo Sasmono (2018) |
| PRNT | 13 % | | Sex | 9 | 71 | 71 | | Dasan Geres and Kelayu, Lombok | Unspecified | Community | Other | Olson (1983) |
| PRNT | 6 - 100 | | Other | | 116 | 116 | | Aceh Jaya | 2017 - 2017 | Population | General Population | Harapan (2022) |
| **Iran** | | | | | | | | | | | | |
| IgG | 0 % | | | 0 | 494 | 494 | | Hormozgan province | Sep 2016 - Jun 2017 | Mixed | Mixed Groups | Ziyaeyan (2018) |
| **Italy** | | | | | | | | | | | | |
| IgM | 1.3 % | | | 2 | 156 | 156 | | Bari | Mar 2015 - Jun 2017 | Travel | Persons Under Investigation | Loconsole (2018) |
| IgM | 16.7 % | | | 5 | 30 | 30 | | | Nov 2015 - Nov 2022 | Travel | Persons Under Investigation | Merakou (2023) |
| IgM | 21.4 % | | | 104 | 487 | 497 | | | Nov 2015 - Nov 2022 | Travel | Persons Under Investigation | Merakou (2023) |
| IgM | 57.1 % | | | 4 | 7 | 7 | | | Nov 2015 - Nov 2022 | Travel | Persons Under Investigation | Merakou (2023) |
| IgM | 77.3 % | | | 34 | 44 | 44 | | | Nov 2015 - Nov 2022 | Travel | Persons Under Investigation | Merakou (2023) |

continued on next page

continued from previous page

| Seroprev- alence type | Parameter value | Uncert- ainty | Disag- gregation | Num- erator | Denom- inator | Sample size | PRNT on ELISA + tests | Location | Dates | Population sample type | Population group | Reference |
|---|---|---|---|---|---|---|---|---|---|---|---|---|
| PRNT | 16 % | | | 95 | 594 | 594 | | | Nov 2015 - Nov 2022 | Travel | Persons Under Investigation | Merakou (2023) |
| PRNT | 86 % | | | 38 | 44 | 44 | | | Nov 2015 - Nov 2022 | Travel | Persons Under Investigation | Merakou (2023) |
| **Kenya** | | | | | | | | | | | | |
| HAI/HI | 1.3 % | | Age, Other | 14 | 1042 | 1042 | | Kitui District | Nov 1966 - Apr 1968 | Population | General Population | Geser (1970) |
| HAI/HI | 3.3 % | | Age, Other | 27 | 822 | 822 | | Central Nyanza | Nov 1966 - Apr 1968 | Population | General Population | Geser (1970) |
| HAI/HI | 52 % | | Age, Other | 434 | 834 | 834 | | Malindi District | Nov 1966 - Apr 1968 | Population | General Population | Geser (1970) |
| IgG | 0 % | | | 0 | 135 | 135 | | Eldoret | Jan 2013 - Feb 2013 | Population | Other | Kisuya (2019) |
| IgG | 0 % | | | 0 | 96 | 96 | | Nairobi | Aug 2013 - Sep 2013 | Population | Other | Kisuya (2019) |
| IgG | 0.9 % | | | 5 | 577 | 577 | | Eldoret, Kisumu, Nairobi | Mar 2009 - Jan 2014 | Population | Mixed Groups | Kisuya (2019) |
| IgG | 1.4 % | | | 3 | 212 | 212 | | Nairobi | Mar 2009 - Nov 2012 | Population | Other | Kisuya (2019) |
| IgG | 1.5 % | | | 2 | 134 | 134 | | Kisumu | Dec 2013 - Jan 2014 | Population | Other | Kisuya (2019) |
| IgM | 7.2 % | | | 166 | 2293 | 1916 | | Mombasa | Oct 2017 - Jul 2019 | Hospital | Pregnant Women | Osoro (2022) |
| PRNT | 0 % | | | 0 | 25 | 25 | | Prigan, Turkana | Aug 2017 - Aug 2017 | Population | General Population | Chepkorir (2019) |
| PRNT | 0 % | | | 0 | 61 | 61 | | Kare Edome, Turkana | Aug 2017 - Aug 2017 | Population | General Population | Chepkorir (2019) |
| PRNT | 0 % | | | 0 | 69 | 69 | | Nayanaesanyati, Turkana | Aug 2017 - Aug 2017 | Population | General Population | Chepkorir (2019) |
| PRNT | 0 % | | | 0 | 56 | 56 | | Old Gunner, Turkana | Aug 2017 - Aug 2017 | Population | General Population | Chepkorir (2019) |
| PRNT | 0 % | | | 0 | 139 | 139 | | Lowarengak, Turkana | Aug 2017 - Aug 2017 | Population | General Population | Chepkorir (2019) |
| PRNT | | | | 0 | 11 | 11 | TRUE | Eldoret, Kisumu, Nairobi | Mar 2009 - Jan 2014 | Population | Persons Under Investigation | Kisuya (2019) |
| PRNT | 0.24 % | | Age, Region | 1 | 413 | 413 | | Turkana | Aug 2017 - Aug 2017 | Population | General Population | Chepkorir (2019) |
| PRNT | 1.59 % | | | 1 | 63 | 63 | | Elelea, Turkana | Aug 2017 - Aug 2017 | Population | General Population | Chepkorir (2019) |
| PRNT | 1.67 % | | | 1 | 60 | 60 | | Longarkau, West Pokot | Feb 2016 - Feb 2016 | Population | General Population | Chepkorir (2019) |
| PRNT | | | | 3 | 144 | 1916 | TRUE | Mombasa | Oct 2017 - Jul 2019 | Hospital | Pregnant Women | Osoro (2022) |
| PRNT | 3.85 % | | | 2 | 52 | 52 | | Pkotong, West Pokot | Feb 2016 - Feb 2016 | Population | General Population | Chepkorir (2019) |
| PRNT | 4.11 % | | | 3 | 73 | 73 | | Shaba, West Pokot | Feb 2016 - Feb 2016 | Population | General Population | Chepkorir (2019) |
| PRNT | 7.11 % | | Age, Region | 33 | 464 | 464 | | West Pokot | Feb 2016 - Feb 2016 | Population | General Population | Chepkorir (2019) |
| PRNT | 8 % | | | 8 | 100 | 100 | | Kokipei, West Pokot | Feb 2016 - Feb 2016 | Population | General Population | Chepkorir (2019) |
| PRNT | 9.09 % | | | 6 | 66 | 66 | | Sangakai, West Pokot | Feb 2016 - Feb 2016 | Population | General Population | Chepkorir (2019) |
| PRNT | 11.5 % | | | 13 | 113 | 113 | | Kanyerus, West Pokot | Feb 2016 - Feb 2016 | Population | General Population | Chepkorir (2019) |
| **Laos** | | | | | | | | | | | | |
| IgG | 17.3 % | | | 62 | 359 | 359 | | Vientiane capital | 2003 - 2004 | Population | Blood Donors | Pastorino (2019) |
| IgG | 27.9 % | | | 192 | 687 | 687 | | Vientiane capital | 2015 - 2015 | Population | Blood Donors | Pastorino (2019) |
| PRNT | 3.9 - 10.1 % | 95% CI: | Age, Sex, Time | | | | TRUE | Vientiane capital | 2003 - 2015 | Population | Blood Donors | Pastorino (2019) |
| **Madagascar** | | | | | | | | | | | | |

continued on next page

continued from previous page

| Seroprev- alence type | Parameter value | Uncert- ainty | Disag- gregation | Num- erator | Denom- inator | Sample size | PRNT on ELISA + tests | Location | Dates | Population sample type | Population group | Reference |
|---|---|---|---|---|---|---|---|---|---|---|---|---|
| IgG | 4.9 % | | Age, Region, Sex, Time | | 1036 | 1036 | | | Jan 2018 - Jun 2021 | Hospital | Other | Rakotomalala (2023) |
| **Malaysia** | | | | | | | | | | | | |
| IgG | 0 % | | | 0 | 22 | 22 | | Paya Pelong | Unspecified | Community | General Population | Khor (2020) |
| IgG | | | | 15 | 418 | 418 | | Sabah | 2018 - 2018 | Hospital | Blood Donors | Tun (2022) |
| IgG | 4 % | | | 4 | 101 | 101 | | Sangwai | Unspecified | Community | General Population | Khor (2020) |
| IgG | 4.5 % | | | 3 | 67 | 67 | | Jekjok | Unspecified | Community | General Population | Khor (2020) |
| IgG | 5.3 % | | | 1 | 19 | 19 | | Pasir Intan | Unspecified | Community | General Population | Khor (2020) |
| IgG | 5.9 % | | | 1 | 17 | 17 | | Bukit Payung | Unspecified | Community | General Population | Khor (2020) |
| IgG | 6.9 % | | | 6 | 87 | 87 | | Tumboh Hangat | Unspecified | Community | General Population | Khor (2020) |
| IgG | 7.7 % | | | 3 | 39 | 39 | | Semanggar | Unspecified | Community | General Population | Khor (2020) |
| IgG | | | | 36 | 400 | 400 | | Sabah | 2017 - 2017 | Hospital | Persons Under Investigation | Tun (2022) |
| IgG | 9.4 % | | | 12 | 90 | 90 | | Paya Sendayan | Unspecified | Community | General Population | Khor (2020) |
| IgG | | | | 58 | 498 | 498 | | | Unspecified | Community | General Population | Khor (2020) |
| IgG | 13.2 % | | Region, Sex | 115 | 872 | 872 | | | Unspecified | Community | General Population | Khor (2020) |
| IgG | | | | 57 | 374 | 374 | | | Unspecified | Community | General Population | Khor (2020) |
| IgG | 17.3 % | | | 14 | 81 | 81 | | Lubuk Legong | Unspecified | Community | General Population | Khor (2020) |
| IgG | 17.4 % | | | 8 | 46 | 46 | | Ulu Kelaka | Unspecified | Community | General Population | Khor (2020) |
| IgG | 21.3 % | | | 10 | 47 | 47 | | Paya Lebar | Unspecified | Community | General Population | Khor (2020) |
| IgG | 22 % | | | 20 | 91 | 91 | | Sungai Perah | Unspecified | Community | General Population | Khor (2020) |
| IgG | 22.5 % | | | 8 | 45 | 45 | | Donglai Baru | Unspecified | Community | General Population | Khor (2020) |
| IgG | 22.5 % | | | 27 | 120 | 120 | | Dusun Kubur | Unspecified | Community | General Population | Khor (2020) |
| IgG | 41.7 % | | | 244 | 585 | 585 | | "Peninsula" | Unspecified | Population | General Population | Khor (2024) |
| IgM | | | | 16 | 418 | 418 | | Sabah | 2018 - 2018 | Hospital | Blood Donors | Tun (2022) |
| IgM | | | | 38 | 400 | 400 | | Sabah | 2017 - 2017 | Hospital | Persons Under Investigation | Tun (2022) |
| NS1 BOB ELISA | 7.6 % | 95% CI: 6.1-9.3 | Age, Other, Sex | 82 | 1085 | 1085 | | Kuala Lumpur | 2012 - 2017 | Mixed | Mixed Groups | Sam (2019) |
| PRNT | 0.5 % | 95% CI: 0.13-1.8 | | 2 | 38 | 38 | TRUE | Sabah | 2017 - 2017 | Hospital | Persons Under Investigation | Tun (2022) |
| PRNT | 1.4 % | 95% CI: 0.65-3 | | 6 | 16 | 26 | TRUE | Sabah | 2018 - 2018 | Hospital | Blood Donors | Tun (2022) |
| PRNT | 1.4 % | 95% CI: 0.65-3 | | 6 | 16 | 26 | TRUE | Sabah | 2018 - 2018 | Hospital | Blood Donors | Tun (2022) |
| PRNT | 3.3 % | 95% CI: 2.4-4.6 | | 36 | 82 | 1085 | TRUE | Kuala Lumpur | 2012 - 2017 | Mixed | Mixed Groups | Sam (2019) |
| PRNT | 6 % | 95% CI: 4-8 | | 24 | 38 | 38 | TRUE | Sabah | 2017 - 2017 | Hospital | Persons Under Investigation | Tun (2022) |
| **Mali** | | | | | | | | | | | | |
| IgG | 6.2 % | | | 4 | 65 | 65 | | Niono | Oct 2016 - Nov 2016 | Population | General Population | Diarra (2020) |
| IgG | 8.5 % | | | 11 | 129 | 129 | | Bamako | Oct 2016 - Nov 2016 | Population | General Population | Diarra (2020) |
| IgG | 10.5 % | | | 67 | 637 | 637 | | Bamako | 2013 - 2013 | Population | General Population | Diarra (2020) |
| IgG | 16.2 % | | | 22 | 136 | 136 | | Kadiolo | Oct 2016 - Nov 2016 | Population | General Population | Diarra (2020) |

continued on next page

continued from previous page

| Seroprev- alence type | Parameter value | Uncert- ainty | Disag- gregation | Num- erator | Denom- inator | Sample size | PRNT on ELISA + tests | Location | Dates | Population sample type | Population group | Reference |
|---|---|---|---|---|---|---|---|---|---|---|---|---|
| IgG | 17.3 % | | | 22 | 127 | 127 | | Bougouni | Oct 2016 - Nov 2016 | Population | General Population | Diarra (2020) |
| IgG | 17.5 % | | | 7 | 40 | 40 | | Kita | Oct 2016 - Nov 2016 | Population | General Population | Diarra (2020) |
| IgG | 19 % | 95% CI: 13.6-24.4 | | 38 | 200 | 200 | | Soromba, Sibirila, Bougouni, Sikasso | Feb 2015 - Feb 2015 | Population | General Population | Bane (2024) |
| IgG | 24.5 % | 95% CI: 18.5-30.5 | | 49 | 200 | 200 | | Banzana, Sibirila, Bougouni, Sikasso | Feb 2015 - Feb 2015 | Population | General Population | Bane (2024) |
| IgG | 34 % | 95% CI: 27.4-40.6 | | 68 | 200 | 200 | | Bamba, Sibirila, Bougouni, Sikasso | Feb 2015 - Feb 2015 | Population | General Population | Bane (2024) |
| IgG | 35.8 % | | | 39 | 109 | 109 | | Diema | Oct 2016 - Nov 2016 | Population | General Population | Diarra (2020) |
| IgG | 43.3 % | | | 81 | 187 | 187 | | Bandiagara | Oct 2016 - Nov 2016 | Population | General Population | Diarra (2020) |
| PRNT | 0.3 % | 95% CI: 0.01-1.9 | Age, Sex | | | | | Bamako | Aug 2007 - Oct 2007 | | | Marchi (2020) |
| PRNT | 0.4 % | 95% CI: 0.01-2.42 | Age, Sex | | | | | Bamako | Nov 2011 - Apr 2012 | | | Marchi (2020) |
| PRNT | 3.1 % | | | 2 | 8 | 65 | TRUE | Niono | Oct 2016 - Nov 2016 | Population | General Population | Diarra (2020) |
| PRNT | 5.4 % | | | 7 | 11 | 129 | TRUE | Bamako | Oct 2016 - Nov 2016 | Population | General Population | Diarra (2020) |
| PRNT | 7.4 % | | | 47 | 85 | 637 | TRUE | Bamako | 2013 - 2013 | Population | General Population | Diarra (2020) |
| PRNT | 10.3 % | | | 14 | 23 | 136 | TRUE | Kadiolo | Oct 2016 - Nov 2016 | Population | General Population | Diarra (2020) |
| PRNT | 11.8 % | | | 15 | 25 | 127 | TRUE | Bougouni | Oct 2016 - Nov 2016 | Population | General Population | Diarra (2020) |
| PRNT | 15 % | | | 6 | 12 | 40 | TRUE | Kita | Oct 2016 - Nov 2016 | Population | General Population | Diarra (2020) |
| PRNT | 15.5 % | | | 29 | 105 | 187 | TRUE | Bandiagara | Oct 2016 - Nov 2016 | Population | General Population | Diarra (2020) |
| PRNT | 20.2 % | | | 22 | 53 | 109 | TRUE | Diema | Oct 2016 - Nov 2016 | Population | General Population | Diarra (2020) |
| **Mexico** | | | | | | | | | | | | |
| IgG | 5.5 % | | | | 126 | 126 | | Oaxaca | Dec 2017 - Feb 2018 | Hospital | Children | Porras-Garcia (2024) |
| IgG | 28.6 % | | | 36 | 126 | 126 | | Oaxaca | Dec 2017 - Feb 2018 | Hospital | Pregnant Women | Porras-Garcia (2024) |
| IgG | 2.4 - 27.1 | | Age | 286 | 768 | 768 | | Merida | 2020 - 2020 | Hospital | Persons Under Investigation | Earnest (2024) |
| IgG | 62 % | | Age | 85 | 136 | 136 | | Chiapas | Feb 2019 - Aug 2019 | Hospital | Pregnant Women | Eligio-García (2020) |
| IgM | 8 % | | Other, Symptoms | 9 | 113 | 113 | | Monterrey City | 01 Aug 2017 - 30 Jun 2018 | Hospital | Persons Under Investigation | Gongora-Rivera (2020) |
| IgM | 12 % | | Other, Symptoms | 3 | 26 | 26 | | Monterrey City | 01 Aug 2017 - 30 Jun 2018 | Hospital | Persons Under Investigation | Gongora-Rivera (2020) |
| IgM | 31 % | | | 39 | 126 | 126 | | Oaxaca | Dec 2017 - Feb 2018 | Hospital | Pregnant Women | Porras-Garcia (2024) |
| NS1 BOB ELISA | 0 % | | | | 266 | 266 | | | 2013 - 2014 | Population | General Population | Nascimento (2019) |
| NS1 BOB ELISA | 31 % | | | | 266 | 266 | | | 2017 - 2018 | Population | General Population | Nascimento (2019) |
| PRNT | | | | 2 | 263 | 48 | | Guerrero | May 2019 - Nov 2019 | Hospital | Other | Nunez-Avellaneda (2021) |
| PRNT | | | | 6 | 263 | 60 | | Guerrero | May 2019 - Nov 2019 | Hospital | Other | Nunez-Avellaneda (2021) |
| PRNT | 3 % | | | | 266 | 266 | TRUE | | 2013 - 2014 | Population | General Population | Nascimento (2019) |

continued on next page

continued from previous page

| Seroprev- alence type | Parameter value | Uncert- ainty | Disag- gregation | Num- erator | Denom- inator | Sample size | PRNT on ELISA + tests | Location | Dates | Population sample type | Population group | Reference |
|---|---|---|---|---|---|---|---|---|---|---|---|---|
| PRNT | 7 % | | | | 266 | 266 | TRUE | | 2017 - 2018 | Population | General Population | Nascimento (2019) |
| PRNT | 38.3 % | | | 103 | 269 | 269 | | | Jun 2011 - Mar 2012 | Other | Children | Zambrano (2021) |
| PRNT | 62.1 % | | | 74 | 119 | 119 | | Chiapas State | 30 Nov 2015 - 18 Dec 2015 | Population | General Population | Guerbois (2016) |
| PRNT | | | | 9 | 13 | 13 | | Oaxaca | Dec 2017 - Feb 2018 | Hospital | Children | Porras-Garcia (2024) |
| **Mozambique** | | | | | | | | | | | | |
| IgM | 2.5 % | 95% CI: 8.5-17.4 | | 5 | 193 | 193 | | South Region | 2009 - 2015 | Population | Other | Chelene (2019) |
| IgM | 4.7 % | 95% CI: 3.9-9 | | 17 | 356 | 356 | | Central Region | 2009 - 2015 | Population | Other | Chelene (2019) |
| IgM | 4.9 % | 95% CI: 3.5-6.6 | Age, Sex, Time | 42 | 850 | 850 | | | 2009 - 2015 | Population | Other | Chelene (2019) |
| IgM | 6.6 % | 95% CI: 10.2-17 | | 20 | 301 | 301 | | North Region | 2009 - 2015 | Population | Other | Chelene (2019) |
| **Myanmar** | | | | | | | | | | | | |
| IgM | 31.3 % | | | 89 | 284 | 284 | | Yangon | 2018 - 2018 | Population | General Population | Tun (2020a) |
| **Netherlands** | | | | | | | | | | | | |
| IgG | 0.6 % | 95% CI: 0.1-2.1 | | 2 | 326 | 326 | | | Mar 2014 - Oct 2017 | Travel | General Population | Overbosch (2023) |
| IgG | 14 % | | | 18 | 129 | 129 | | | Mar 2014 - Oct 2017 | Travel | General Population | Overbosch (2023) |
| **New Zealand** | | | | | | | | | | | | |
| IgG | 58.3 % | | Age, Sex | 35 | 60 | 60 | | Aitutaki | Jan 2017 - Apr 2017 | Population | General Population | Saretzki (2024) |
| IgG | 67.8 % | | Age, Sex | 141 | 208 | 208 | | Rarotonga, Cook Islands | Jan 2017 - Apr 2017 | Population | General Population | Saretzki (2024) |
| **Nicaragua** | | | | | | | | | | | | |
| IgG | 66.4 % | | | 89 | 134 | 134 | | León | 05 Feb 2017 | Hospital | Pregnant Women | Zepeda (2023) |
| IgG | 80.4 % | | | 82 | 102 | 102 | | León | 05 Feb 2017 | Hospital | Pregnant Women | Zepeda (2023) |
| IgG | 94 % | | | 176 | 187 | 187 | | León | Feb 2017 - Jul 2017 | Hospital | Pregnant Women | Collins (2020) |
| IgM | | | Age, Sex, Symptoms | 23 | 142 | 142 | | Managua | 31 Aug 2016 - 21 Oct 2016 | Household | Household Contacts Of Survivors | Burger-Calderon (2018) |
| NS1 BOB ELISA | 36.1 % | 95% CI: 34.5-37.8 | Age, Other, Region, Sex | | | | | District II, Managua | Feb 2017 - Jul 2017 | Household | Children | Zambrana (2018) |
| NS1 BOB ELISA | 45.8 % | 95% CI: 43.4-48.3 | Age, Sex | | | | | District II, Managua | Feb 2017 - Jul 2017 | Household | General Population | Zambrana (2018) |
| NS1 BOB ELISA | 56.4 % | 95% CI: 53.1-59.6 | Age, Other, Region, Sex | | | | | District II, Managua | Feb 2017 - Jul 2017 | Household | Other | Zambrana (2018) |
| PRNT | 61 % | | | 107 | 176 | 176 | TRUE | León | Feb 2017 - Jul 2017 | Hospital | Pregnant Women | Collins (2020) |
| **Nigeria** | | | | | | | | | | | | |
| HAI/HI | 56 % | | Age | 150 | 267 | | | Kainji Lake | 1980 - 1980 | | | Adekolu-John (1983) |
| IgG | 0.4 % | | Symptoms | 4 | 1006 | 1006 | | Jos | 01 Apr 2019 - 31 Jan 2022 | Hospital | Pregnant Women | Ogwuche (2023) |
| IgG | 3.6 % | | Sex, Symptoms | 17 | 468 | 468 | | Plateau, Nasarawa | Jan 2016 - Dec 2016 | Community | General Population | Mathe (2018) |
| IgG | 12 % | 95% CI: 9.9-14.4 | | 12 | 100 | 100 | | Cross River State | 17 Mar 2017 - 30 May 2017 | Population | Persons Under Investigation | Otu (2020) |
| IgG | 18.3 % | | | 55 | 300 | 300 | | Baru-Dokp Teaching Hospital, Kaduna, Kaduna State | Dec 2020 - Nov 2021 | Hospital | Mixed Groups | Mac (2023a) |

continued on next page

continued from previous page

| Seroprev- alence type | Parameter value | Uncert- ainty | Disag- gregation | Num- erator | Denom- inator | Sample size | PRNT on ELISA + tests | Location | Dates | Population sample type | Population group | Reference |
|---|---|---|---|---|---|---|---|---|---|---|---|---|
| IgG | 18.6 % | | Age, Other, Region, Sex | 82 | 419 | 871 | | Nasarawa | Dec 2020 - Nov 2021 | Hospital | Mixed Groups | Peter Asaga (2023) |
| IgG | 18.7 % | | Age, Other, Region, Sex | 56 | 300 | 871 | | Kaduna | Dec 2020 - Nov 2021 | Hospital | Mixed Groups | Peter Asaga (2023) |
| IgG | 18.9 % | | | 79 | 419 | 419 | | Federal Medical Centre, Keffi, Nasarawa State. | Dec 2020 - Nov 2021 | Hospital | Mixed Groups | Mac (2023a) |
| IgG | 19.2 % | | Age, Other, Region, Sex | 167 | 871 | 871 | | Abia State University Teaching Hospital, Aba, Abia state, Federal Medical Centre, Keffi, Nasarawa State, Baru-Dokp Teaching Hospital, Kaduna, Kaduna State | Dec 2020 - Nov 2021 | Hospital | Mixed Groups | Mac (2023a) |
| IgG | 19.2 % | 95% CI: 0.16-0.21 | Age, Other, Region, Sex, Symptoms | 167 | 871 | 871 | | 3 states in Nigeria Nigeria, Abia State in Southern Nigeria, and Kaduna | Dec 2020 - Nov 2021 | Hospital | Mixed Groups | Mac (2023a) |
| State in Northern Nigeria | Dec 2020 - Nov 2021 | Hospital | General Population | Mac (2023b) | 871 | 871 | | 3 states in Nigeria Nigeria, Abia State in Southern Nigeria, and Kaduna | Dec 2020 - Nov 2021 | Hospital | Mixed Groups | Mac (2023a) |
| IgG | 21.7 % | | | 33 | 152 | 152 | | Abia State University Teaching Hospital, Aba, Abia state | Dec 2020 - Nov 2021 | Hospital | Mixed Groups | Mac (2023a) |
| IgG | 22.4 % | | Age, Other, Region, Sex | 34 | 152 | 871 | | Abia | Dec 2020 - Nov 2021 | Hospital | Mixed Groups | Peter Asaga (2023) |
| IgG | 0 - 62.7 % | | Age, Other, Region, Sex | | 871 | 871 | | Abia, Kaduna, Nasarawa | Dec 2020 - Nov 2021 | Hospital | Mixed Groups | Peter Asaga (2023) |
| IgG | 85 % | | | 170 | 200 | 200 | | Borno state | Apr 2018 - Aug 2018 | Population | Persons Under Investigation | Baba (2023) |
| IgG and IgM | 20 % | 95% CI: 11.9-30.2 | | 20 | 100 | 100 | | Cross River State | 17 Mar 2017 - 30 May 2017 | Population | Persons Under Investigation | Otu (2020) |
| IgM | 1.4 % | | Symptoms | 14 | 1006 | 1006 | | Jos | 01 Apr 2019 - 31 Jan 2022 | Hospital | Pregnant Women | Ogwuche (2023) |
| IgM | 6.2 % | | Sex, Symptoms | 29 | 468 | 468 | | Plateau, Nasarawa | Jan 2016 - Dec 2016 | Community | General Population | Mathe (2018) |
| IgM | 10 % | 95% CI: 3.8-20.1 | | 10 | 100 | 100 | | Cross River State | 17 Mar 2017 - 30 May 2017 | Population | Persons Under Investigation | Otu (2020) |
| IgM | 12.1 % | | Other | 60 | 496 | 496 | | Adamawa, Bauchi, and Borno states | Apr 2018 - Aug 2018 | Population | Persons Under Investigation | Baba (2023) |

continued on next page

continued from previous page

| Seroprev- alence type | Parameter value | Uncert- ainty | Disag- gregation | Num- erator | Denom- inator | Sample size | PRNT on ELISA + tests | Location | Dates | Population sample type | Population group | Reference |
|---|---|---|---|---|---|---|---|---|---|---|---|---|
| IgM | 14.5 % | | | 29 | 96 | 200 | | Borno state | Apr 2018 - Aug 2018 | Population | Persons Under Investigation | Baba (2023) |
| IgM | 15.5 % | | | 31 | 200 | 200 | | Adamawa state | Apr 2018 - Aug 2018 | Population | Persons Under Investigation | Baba (2023) |
| IgM | 22 % | | | 30 | 137 | 200 | | Borno state | Mar 2018 - Apr 2018 | Hospital | Persons Under Investigation | Oderinde (2020) |
| IgM | 25 % | | | 51 | 200 | 200 | | Borno state | Mar 2018 - Apr 2018 | Hospital | Persons Under Investigation | Oderinde (2020) |
| IgM | 29.4 % | | | 53 | 180 | 180 | | Zaria | Unspecified | Hospital | Pregnant Women | Adekola (2023) |
| PRNT | 3 % | | | 6 | 200 | 200 | | Adamawa state | Apr 2018 - Aug 2018 | Population | Persons Under Investigation | Baba (2023) |
| PRNT | 9.4 % | | | 16 | 170 | 200 | | Borno state | Apr 2018 - Aug 2018 | Population | Persons Under Investigation | Baba (2023) |
| PRNT | 12 % | | | 24 | 200 | 200 | | Borno state | Apr 2018 - Aug 2018 | Population | Persons Under Investigation | Baba (2023) |
| PRNT | 13.1 % | | Other | 65 | 496 | 496 | | Adamawa, Bauchi, and Borno states | Apr 2018 - Aug 2018 | Population | Persons Under Investigation | Baba (2023) |
| PRNT | 30 % | | | 6 | 20 | 20 | | Igbo-Ora | May 1975 - Oct 1975 | Population | Other | Fagbami (1977) |
| PRNT | 36.5 % | | | 35 | 96 | 96 | | Bauchi state | Apr 2018 - Aug 2018 | Population | Persons Under Investigation | Baba (2023) |
| **Pakistan** | | | | | | | | | | | | |
| IgG | 6.48 % | | | 47 | 725 | 725 | | Faisalabad | 2019 - 2019 | Population | | Chen (2024) |
| PRNT | 0.69 % | | | 5 | 47 | 725 | TRUE | Faisalabad | 2019 - 2019 | Population | General Population | Chen (2024) |
| PRNT | 6.48 % | | | 47 | 47 | 725 | TRUE | Faisalabad | 2019 - 2019 | Population | General Population | Chen (2024) |
| **Panama** | | | | | | | | | | | | |
| MIA | 30.3 % | | | 121 | 400 | 400 | | | 2015 - 2016 | Population | Persons Under Investigation | Eskildsen (2020) |
| **Papua New Guinea** | | | | | | | | | | | | |
| IgG | 66 % | | | 87 | 132 | 132 | | Wewak | Apr 2019 - Apr 2019 | Other | Other | Grant (2022b) |
| IgG | 69.7 % | | | 53 | 76 | 76 | | Manus Island | Apr 2019 - Apr 2019 | Other | Other | Grant (2022b) |
| IgM | 9 % | | | 7 | 76 | 76 | | Manus Island | Apr 2019 - Apr 2019 | Other | Other | Grant (2022b) |
| IgM | 9.1 % | | | 12 | 132 | 132 | | Wewak | Apr 2019 - Apr 2019 | Other | Other | Grant (2022b) |
| PRNT | 63.2 % | | | 48 | 55 | 76 | TRUE | Manus Island | Apr 2019 - Apr 2019 | Other | Other | Grant (2022b) |
| PRNT | 65.9 % | | | 87 | 93 | 132 | TRUE | Wewak | Apr 2019 - Apr 2019 | Other | Other | Grant (2022b) |
| **Paraguay** | | | | | | | | | | | | |
| IgG | 31.4 % | | | 49 | 156 | 156 | | Central department and Asuncion | Jan 2018 - May 2018 | Hospital | Other | Rojas (2019) |
| **Peru** | | | | | | | | | | | | |
| IgG | 2.5 % | | | | 200 | 200 | | Chincha Baja | Mar 2019 - May 2019 | Population | General Population | Cachay (2021) |
| IgG | 23.5 % | | | | 200 | 200 | | Pueblo Nuevo | Mar 2019 - May 2019 | Population | General Population | Cachay (2021) |
| **Philippines** | | | | | | | | | | | | |
| Biotinylated- | | | | | | | | | | | | |
| EDIII antigen | | | | | | | | | | | | |
| capture ELISA | 18 % | | | 98 | 547 | 547 | | Balamban and Bogo City in Cebu Province | 2017 - 2017 | Other | Children | Adams (2021) |

continued on next page

continued from previous page

| Seroprev-alence type | Parameter value | Uncert-ainty | Disag- gregation | Num-erator | Denom-inator | Sample size | PRNT on ELISA + tests | Location | Dates | Population sample type | Population group | Reference |
|---|---|---|---|---|---|---|---|---|---|---|---|---|
| IgG | 23 % | | | 118 | 508 | | | | 2016 - 2016 | Hospital | Persons Under Investigation | Biggs (2020) |
| IgG | 31.5 % | | | 314 | 997 | 997 | | | 2016 - 2016 | Population | Persons Under Investigation | Biggs (2021) |
| IgM | 0 - 1 % | | | | 508 | 508 | | | 2016 - 2016 | Hospital | Persons Under Investigation | Biggs (2020) |
| **Republic of the Congo** | | | | | | | | | | | | |
| IgG | 5.7 % | | | | 386 | 386 | | Brazzaville, Pointe-Noire, Ewo, Gamboma, Oyo, Owando | Mar 2011 - Jul 2011 | Other | Blood Donors | Nurtop (2020) |
| PRNT | 1.1 % | | | | | | TRUE | Brazzaville | Mar 2011 - Jul 2011 | Other | Blood Donors | Nurtop (2020) |
| PRNT | 1.8 % | | | | 386 | 386 | TRUE | Brazzaville, Pointe-Noire, Ewo, Gamboma, Oyo, Owando | Mar 2011 - Jul 2011 | Other | Blood Donors | Nurtop (2020) |
| PRNT | 3.2 % | | | | | | TRUE | Pointe-Noir | Mar 2011 - Jul 2011 | Other | Blood Donors | Nurtop (2020) |
| **Rwanda** | | | | | | | | | | | | |
| IgG | 1.4 % | | | 12 | 874 | 874 | | 4 provinces and Kigali | 2015 - 2015 | Population | General Population | Seruyange (2018) |
| **Saudi Arabia** | | | | | | | | | | | | |
| IgG | | | | 0 | 52 | 52 | | Najran city | Nov 2016 - Jul 2017 | Hospital | Other | Alayed (2018) |
| IgG | 1 % | | | 2 | 217 | 217 | | Abha | 2019 - 2020 | Hospital | Pregnant Women | Harish (2023) |
| IgG | | | | 52 | 410 | 410 | | Najran city | Nov 2016 - Jul 2017 | Hospital | Pregnant Women | Alayed (2018) |
| IgM | | | | 0 | 52 | 52 | | Najran city | Nov 2016 - Jul 2017 | Hospital | Other | Alayed (2018) |
| IgM | 1.8 % | | | 4 | 217 | 217 | | Abha | 2019 - 2020 | Hospital | Pregnant Women | Harish (2023) |
| IgM | | | | 24 | 410 | 410 | | Najran city | Nov 2016 - Jul 2017 | Hospital | Pregnant Women | Alayed (2018) |
| **Senegal** | | | | | | | | | | | | |
| IgM | | | | 9 | 13845 | 13845 | | Kedougou | Jul 2009 - Mar 2013 | Hospital | Persons Under Investigation | Sow (2016) |
| PRNT | 13.4 % | 95% CI: 9.65-17.9 | Age, Sex | | | | | Niakhar | Aug 2007 - Oct 2007 | | | Marchi (2020) |
| PRNT | 13.7 % | 95% CI: 9.4-19.14 | Age, Sex | | | | | Niakhar | Nov 2011 - Apr 2012 | | | Marchi (2020) |
| **Solomon Islands** | | | | | | | | | | | | |
| IgG | | | Age | 11 | 1021 | | | | Apr 2018 - Nov 2018 | Community | General Population | Russell (2022) |
| **Spain** | | | | | | | | | | | | |
| IgG | 52.7 % | | | 214 | 406 | 406 | | Barcelona | Jan 2016 - Apr 2019 | Population | Pregnant Women | Martinez-Arias (2023) |
| IgM | 0.5 % | | | 2 | 406 | 406 | | Barcelona | Jan 2016 - Apr 2019 | Population | Pregnant Women | Martinez-Arias (2023) |
| IgM | 7.8 % | | | 47 | 602 | 602 | | Madrid | Jan 2016 - Jan 2017 | Hospital | Persons Under Investigation | Crespillo-Andujar (2020) |
| PRNT | 4.8 % | | | 10 | 209 | 209 | TRUE | Barcelona | Jan 2016 - Apr 2019 | Population | Pregnant Women | Martinez-Arias (2023) |
| PRNT | 84 % | | | 126 | 150 | 150 | | Hospital Universitari Vall d'Hebron Barcelona, Catalonia | May 2016 - Dec 2021 | Mixed | Pregnant Women | Romaní (2022) |

continued on next page

continued from previous page

| Seroprev- alence type | Parameter value | Uncert- ainty | Disag- gregation | Num- erator | Denom- inator | Sample size | PRNT on ELISA + tests | Location | Dates | Population sample type | Population group | Reference |
|---|---|---|---|---|---|---|---|---|---|---|---|---|
| **Sri Lanka** | | | | | | | | | | | | |
| IgG | 34.5 % | | | 102 | 295 | | | Kandy | 2017 - 2019 | Hospital | Persons Under Investigation | Tun (2023) |
| IgG | 42.6 % | | | 128 | 300 | | | Negombo | 2018 - 2019 | Hospital | Persons Under Investigation | Tun (2023) |
| IgM | 1.4 % | | | 4 | 295 | | | Kandy | 2017 - 2019 | Hospital | Persons Under Investigation | Tun (2023) |
| IgM | 9 % | | | 17 | 300 | | | Negombo | 2018 - 2019 | Hospital | Persons Under Investigation | Tun (2023) |
| **Sudan** | | | | | | | | | | | | |
| IgG | 62.7 % | 95% CI: 59.4-66.1 | Age, Region, Sex | 530 | 845 | 845 | | | 2012 - 2012 | Community | Other | Soghaier (2018) |
| PRNT | | | | 1 | 530 | 530 | TRUE | | 2012 - 2012 | Community | Other | Soghaier (2018) |
| PRNT | 3.2 % | | | 3 | 92 | 92 | | | Dec 2012 - Jan 2013 | Population | Other | Adam (2024) |
| PRNT | 21.7 % | | | 23 | 106 | 106 | | | Dec 2012 - Jan 2013 | Population | Other | Adam (2024) |
| **Suriname** | | | | | | | | | | | | |
| IgG | 49.7 % | 95% CI: 41.9-57.5 | Age, Other, Sex, Symptoms | 77 | 155 | 155 | | Kwamalasamutu | Jan 2017 - Feb 2017 | Household | General Population | Langerak (2019) |
| IgG | 68.4 % | 95% CI: 63.8-72.6 | Age, Other, Sex, Symptoms | 290 | 424 | 424 | | Paramaribo | Jan 2017 - Feb 2017 | Hospital | Persons Under Investigation | Langerak (2019) |
| IgG | 83.2 % | 95% CI: 77.3-87.9 | Age, Other, Sex, Symptoms | 159 | 191 | 191 | | Laduani | Jan 2017 - Feb 2017 | Hospital | Persons Under Investigation | Langerak (2019) |
| PRNT | 24.5 % | 95% CI: 18.4-31.9 | Age, Other, Sex, Symptoms | 38 | 155 | 155 | | Kwamalasamutu | Jan 2017 - Feb 2017 | Household | General Population | Langerak (2019) |
| PRNT | 36.7 % | 95% CI: 30.1-43.7 | Age, Other, Sex, Symptoms | 70 | 191 | 191 | | Laduani | Jan 2017 - Feb 2017 | Hospital | Persons Under Investigation | Langerak (2019) |
| PRNT | 38.2 % | 95% CI: 33.7-42.9 | Age, Other, Sex, Symptoms | 162 | 424 | 424 | | Paramaribo | Jan 2017 - Feb 2017 | Hospital | Persons Under Investigation | Langerak (2019) |
| **Sweden** | | | | | | | | | | | | |
| IgG | 0 % | | | 0 | 215 | 215 | | Gothenburg | 2015 - 2015 | Population | General Population | Seruyange (2018) |
| **Taiwan** | | | | | | | | | | | | |
| IgG | 4.2 % | | | 9 | 212 | 212 | | Tainan City | 2015 - 2015 | Travel | General Population | Chien (2019) |
| IgG | 38.8 % | | Age, Other, Sex | | 600 | 600 | | | Jun 2017 - Aug 2017 | Travel | Other | Perng (2019) |
| IgM | 0 % | | | 0 | 212 | 212 | | Tainan City | 2015 - 2015 | Travel | General Population | Chien (2019) |
| IgM | 3 % | | Age, Other, Sex | 18 | 600 | 600 | | | Jun 2017 - Aug 2017 | Travel | Other | Perng (2019) |
| PRNT | 11.11 % | | | 1 | 9 | 212 | TRUE | Tainan City | 2015 - 2015 | Travel | General Population | Chien (2019) |
| **Tanzania** | | | | | | | | | | | | |
| IgG | 2.5 % | | | | | | | Kondoa | Apr 2018 - Nov 2018 | Population | Mixed Groups | Mwanyika (2021) |
| IgG | 3.9 % | | | | | | | Buhigwe | Apr 2018 - Nov 2018 | Population | Mixed Groups | Mwanyika (2021) |
| IgG | 5 % | | | | | | | Kinondoni | Apr 2018 - Nov 2018 | Population | Mixed Groups | Mwanyika (2021) |
| IgG | 5.8 % | | | | | | | Kilindi | Apr 2018 - Nov 2018 | Population | Mixed Groups | Mwanyika (2021) |
| IgG | 5.9 % | | | 59 | 989 | 1818 | | Buhigwe, Kalambo, Kilindi, Kinondoni, Kondoa, Kyela, Mvomero and Ukerewe districts | Apr 2018 - Nov 2018 | Population | Mixed Groups | Mwanyika (2021) |

continued on next page

continued from previous page

| Seroprev- alence type | Parameter value | Uncert- ainty | Disag- gregation | Num- erator | Denom- inator | Sample size | PRNT on ELISA + tests | Location | Dates | Population sample type | Population group | Reference |
|---|---|---|---|---|---|---|---|---|---|---|---|---|
| IgG | 6.8 % | | | | 1818 | 1818 | | Buhigwe, Kalambo, Kilindi, Kinondoni, Kondoa, Kyela, Mvomero and Ukerewe districts | Apr 2018 - Nov 2018 | Population | Mixed Groups | Mwanyika (2021) |
| IgG | 6.1 - 7.6 % | | Age | | 1818 | 1818 | | Buhigwe, Kalambo, Kilindi, Kinondoni, Kondoa, Kyela, Mvomero and Ukerewe districts | Apr 2018 - Nov 2018 | Population | Mixed Groups | Mwanyika (2021) |
| IgG | 7.5 % | | | 62 | 829 | 1818 | | Buhigwe, Kalambo, Kilindi, Kinondoni, Kondoa, Kyela, Mvomero and Ukerewe districts | Apr 2018 - Nov 2018 | Population | Mixed Groups | Mwanyika (2021) |
| IgG | 7.7 % | | | | | | | Kalambo | Apr 2018 - Nov 2018 | Population | Mixed Groups | Mwanyika (2021) |
| IgG | 8.7 % | | | | | | | Kyela | Apr 2018 - Nov 2018 | Population | Mixed Groups | Mwanyika (2021) |
| IgG | 10.6 % | | | | | | | Mvomero | Apr 2018 - Nov 2018 | Population | Mixed Groups | Mwanyika (2021) |
| IgG | 10.6 % | | | | | | | Ukerewe | Apr 2018 - Nov 2018 | Population | Mixed Groups | Mwanyika (2021) |
| **Thailand** | | | | | | | | | | | | |
| IgG | 11 % | | | 8 | 73 | 73 | | Samut Songkhram | Apr 2017 - Apr 2018 | Population | General Population | Sirinam (2022) |
| IgG | 13.4 % | | | 13 | 97 | 97 | | Samut Songkhram | Apr 2017 - Apr 2018 | Population | General Population | Sirinam (2022) |
| IgG | 15.1 % | | Age, Time | 53 | 350 | 350 | | Samut Songkhram | Apr 2017 - Apr 2018 | Population | General Population | Sirinam (2022) |
| IgG | 17.8 % | | | 32 | 180 | 180 | | Samut Songkhram | Apr 2017 - Apr 2018 | Population | General Population | Sirinam (2022) |
| IgG | 16.5 - 27.4 % | 95% CI: 13-33.4 | Other, Sex, Time | | 1648 | 1648 | | | Dec 1997 - Dec 2017 | Population | General Population | Harapan (2022) |
| IgG | 29 % | | | 50 | 174 | 174 | | | 1997 - 2015 | Other | Pregnant Women | Ngo-Giang-Huong (2021) |
| IgG | 30.4 % | | | 290 | 955 | 955 | | Ratchaburi, Bangkok | 2006 - 2009 | School | Children | Sriburin (2021) |
| IgG | 30.77 % | | Age, Other, Time | 200 | 650 | 650 | | Siriraj Hospital | May 2019 - Oct 2019 | Hospital | Pregnant Women | Phatihattakorn (2021) |
| IgG | 83.6 % | | Age, Sex | 51 | 61 | 61 | | Nakhon Ratchasima | Apr 2016 - Jun 2016 | Population | General Population | Hattakam (2021) |
| IgM | 0 % | | | 0 | 174 | 174 | | | 1997 - 2015 | Other | Pregnant Women | Ngo-Giang-Huong (2021) |
| PRNT | 2.8 % | | | | 254 | 254 | | | Dec 2017 - Feb 2020 | Population | General Population | Kitro (2024) |
| PRNT | 8.14 % | | | 21 | 258 | 258 | TRUE | Siriraj Hospital | May 2019 - Oct 2019 | Hospital | Pregnant Women | Phatihattakorn (2021) |
| PRNT | 12.8 % | 95% CI: 9.7-16.5 | | | | | | Narathiwat | Jul 2018 - Mar 2019 | Household | Pregnant Women | Densathaporn (2020) |
| PRNT | 17 % | | | | 147 | 147 | | | 2011 - 2012 | Population | General Population | Yamanaka (2021) |
| PRNT | 21.6 % | | | | 51 | 51 | | Phuket | 2011 - 2012 | Population | General Population | Yamanaka (2021) |
| PRNT | 23.5 % | | | | 34 | 34 | | Nakhon Sawan | 2011 - 2012 | Population | General Population | Yamanaka (2021) |

continued on next page

continued from previous page

| Seroprev- alence type | Parameter value | Uncert- ainty | Disag- gregation | Num- erator | Denom- inator | Sample size | PRNT on ELISA + tests | Location | Dates | Population sample type | Population group | Reference |
|---|---|---|---|---|---|---|---|---|---|---|---|---|
| PRNT | 24.3 % | 95% CI: 20.1-28.8 | | | | | | Surat Thani | Jul 2018 - Mar 2019 | Household | Pregnant Women | Densathaporn (2020) |
| PRNT | 29.7 % | 95% CI: 23.3-26 | | 385 | 499 | 499 | | Narathiwat | Mar 2018 - Sep 2018 | Household | Mixed Groups | Densathaporn (2020) |
| PRNT | 43.7 % | 95% CI: 35.9-51.6 | | 377 | 504 | 504 | | Surat Thani | Mar 2018 - Sep 2018 | Household | Mixed Groups | Densathaporn (2020) |
| PRNT | 44.3 % | | Age, Sex | 27 | 61 | 61 | | Nakhon Ratchasima | Apr 2016 - Jun 2016 | Population | General Population | Hattakam (2021) |
| Western blot | | | | 2 | 21 | 21 | | | Unspecified | Hospital | Persons Under Investigation | Wikan (2016) |
| **The Gambia** | | | | | | | | | | | | |
| PRNT | 3.7 % | 95% CI: 1.87-6.55 | Age, Sex | | | | | Basse Santa Su | Aug 2007 - Oct 2007 | | | Marchi (2020) |
| PRNT | 7.1 % | 95% CI: 4.03-11.45 | Age, Sex | | | | | Basse Santa Su | Nov 2011 - Apr 2012 | | | Marchi (2020) |
| **Uganda** | | | | | | | | | | | | |
| HAI/HI | 6 % | | Region | 8 | 132 | 132 | | Karamoja District | May 1984 - May 1984 | Community | General Population | Rodhain (1989) |
| IgM | | | | 5 | 384 | 384 | | Nkonkonjeru | Feb 2014 - Oct 2017 | Hospital | Other | Kayiwa (2018) |
| PRNT | | | | 3 | 5 | 384 | TRUE | Nkonkonjeru | Feb 2014 - Oct 2017 | Hospital | Other | Kayiwa (2018) |
| **United Kingdom** | | | | | | | | | | | | |
| IgM | 19 % | | | 31 | 161 | 161 | | | 01 Jan 2016 - 31 Dec 2017 | Travel | Persons Under Investigation | Petridou (2019) |
| **United States** | | | | | | | | | | | | |
| IgG | | | | 4 | 18 | 18 | | Hawaii | 2009 - 2012 | Other | Persons Under Investigation | Kumar (2016) |
| IgG | 31.4 % | 95% CI: 30-32.9 | Age, Occupation, Other, Sex | 1268 | 4035 | 4090 | | Ponce, Puerto Rico | Apr 2018 - May 2019 | Household | General Population | Adams (2022) |
| IgM | 2.18 % | | | 277 | 12374 | 12374 | | Hidalgo County, Texas | 2016 - 2018 | Hospital | Mixed Groups | Hinojosa (2020) |
| IgM | 5 % | 95% CI: 2.5-10 | | 8 | 141 | 141 | | | Jan 2016 - Aug 2016 | Travel | Pregnant Women | Rao (2017) |
| IgM | | | | 1 | 18 | 18 | | Hawaii | 2009 - 2012 | Other | Persons Under Investigation | Kumar (2016) |
| IgM | 7 % | | | 22 | 295 | 295 | | New York | 01 Jan 2016 - 30 Jun 2017 | Hospital | Persons Under Investigation | Conners (2018) |
| IgM | 7 % | | | 36 | 547 | 547 | | Dallas, Texas | 14 Mar 2016 - 01 Oct 2016 | Travel | Pregnant Women | Adhikari (2017) |
| IgM | 12.5 % | | | | 713 | 713 | | Ponce, Puerto Rico | 2018 - 2019 | Community | Children | Adams (2023) |
| IgM | 13 % | | | 16 | 141 | 141 | | | 2016 - 2017 | Hospital | Children | Lindsey (2020) |
| IgM | 16 % | | Other | 5 | 31 | 31 | | Congenital Zika Program at Children's National (CZPCN) in Washington DC | Jan 2016 - Jun 2018 | Hospital | Children | Mulkey (2021) |
| IgM | | | | 79 | 367 | 367 | | San Juan, Puerto Rico | 16 Sep 2016 - 27 Oct 2016 | Contact | Other | Lozier (2018) |
| IgM | 22 % | | | 22 | 329 | 329 | | New York City | Jan 2016 - Dec 2017 | Hospital | Persons Under Investigation | Lee (2020) |
| IgM | 30.9 % | 95% CI: 26.3-35.9 | | 112 | 362 | 362 | | Puerto Rico: Ponce, San Juan, Guayama | May 2016 - Jul 2017 | Household | Household Contacts Of Survivors | Rosenberg (2019) |

continued on next page

continued from previous page

| Seroprev- alence type | Parameter value | Uncert- ainty | Disag- gregation | Num- erator | Denom- inator | Sample size | PRNT on ELISA + tests | Location | Dates | Population sample type | Population group | Reference |
|---|---|---|---|---|---|---|---|---|---|---|---|---|
| IgM | | | | 110 | 339 | 339 | | Puerto Rico | Apr 2016 - Dec 2016 | Population | Persons Under Investigation | Williamson (2020) |
| IgM | 37 % | 95% CI: 29-44 | | 56 | 153 | 153 | | | Unspecified | Population | Persons Under Investigation | Hills (2021) |
| IgM | 44 % | | Other | 27 | 61 | 61 | | Congenital Zika Program at Children's National (CZPCN) in Washington DC | Jan 2016 - Jun 2018 | Hospital | Pregnant Women | Mulkey (2021) |
| IgM | 63 % | | | 19 | 30 | 30 | | Miami-Dade County, Florida | Unspecified | Other | Persons Under Investigation | Griffin (2019) |
| NS1 BOB ELISA | 1 % | | | | 91 | 91 | | Puerto Rico | 2013 - 2014 | Population | General Population | Nascimento (2019) |
| NS1 BOB ELISA | 20 % | | | | 91 | 91 | | Puerto Rico | 2017 - 2018 | Population | General Population | Nascimento (2019) |
| PRNT | 0 % | | | 0 | 5 | 5 | TRUE | Congenital Zika Program at Children's National (CZPCN) in Washington DC | Jan 2016 - Jun 2018 | Hospital | Children | Mulkey (2021) |
| PRNT | 0.2 % | 95% CI: 0.01-1.1 | | | 494 | 494 | | Puerto Rico | 01 Jan 2015 - 01 Jun 2015 | Other | Other | Pollett (2022) |
| PRNT | | | | 1 | 141 | 141 | | | Jan 2016 - Aug 2016 | Travel | Pregnant Women | Rao (2017) |
| PRNT | 1 % | | | | 91 | 91 | TRUE | Puerto Rico | 2013 - 2014 | Population | General Population | Nascimento (2019) |
| PRNT | | | | 16 | 969 | 183 | | Veterans Health Administration in US and Caribbean | 01 Dec 2015 - 31 Oct 2016 | Hospital | Other | Schirmer (2018) |
| PRNT | 5 % | | | | 91 | 91 | TRUE | Puerto Rico | 2017 - 2018 | Population | General Population | Nascimento (2019) |
| PRNT | 5.3 % | | | 29 | 36 | 547 | TRUE | Dallas, Texas | 14 Mar 2016 - 01 Oct 2016 | Travel | Pregnant Women | Adhikari (2017) |
| PRNT | 8 % | | | 2 | 26 | 26 | TRUE | Congenital Zika Program at Children's National (CZPCN) in Washington DC | Jan 2016 - Jun 2018 | Hospital | Pregnant Women | Mulkey (2021) |
| PRNT | 18.8 % | | | 44 | 234 | 234 | TRUE | Hidalgo County, Texas | 2016 - 2018 | Hospital | Mixed Groups | Hinojosa (2020) |
| PRNT | 25 % | | | 23 | 92 | 92 | | Puerto Rico | Jun 2011 - Mar 2012 | Other | Children | Zambrano (2021) |
| PRNT | 33.2 % | | | 237 | 713 | 713 | | Ponce, Puerto Rico | 2018 - 2019 | Community | Children | Adams (2023) |
| **Vanuatu** | | | | | | | | | | | | |
| IgG | 51.8 % | | Age, Sex | 102 | 197 | 197 | | Espiritu Santo | Aug 2016 - Jan 2017 | Population | General Population | Saretzki (2024) |
| **Vietnam** | | | | | | | | | | | | |
| IgG | 2.3 % | | | | 176 | 176 | | Ho Chi Minh City | Jan 2008 - Dec 2008 | Hospital | Pregnant Women | Chui (2023) |
| IgG | | | | 1 | 21 | 21 | | Ho Chi Minh City | Mar 2016 - Nov 2017 | Hospital | Children | Grant (2021) |

continued on next page

continued from previous page

| Seroprev- alence type | Parameter value | Uncert- ainty | Disag- gregation | Num- erator | Denom- inator | Sample size | PRNT on ELISA + tests | Location | Dates | Population sample type | Population group | Reference |
|---|---|---|---|---|---|---|---|---|---|---|---|---|
| IgG | | | | 18 | 20 | 20 | | Ho Chi Minh City | Mar 2016 - Nov 2017 | Hospital | Pregnant Women | Grant (2021) |
| IgM | 1 % | | | 21 | 2013 | 2013 | | Nha Trang | jul 2017 - Sep 2018 | Hospital | Children | Tun (2020b) |
| IgM | 10.3 % | | Age, Other | 83 | 801 | 801 | | Krong Buk | Jan 2017 - Jul 2018 | Community | Mixed Groups | Nguyen (2020) |
| PRNT | 0.5 % | | | 11 | 21 | 2013 | TRUE | Nha Trang | Jul 2017 - Sep 2018 | Hospital | Children | Tun (2020b) |
| PRNT | 1.1 % | | Age, Other | 9 | 83 | 801 | TRUE | Krong Buk | Jan 2017 - Jul 2018 | Community | Mixed Groups | Nguyen (2020) |
| **Zambia** | | | | | | | | | | | | |
| IgG | 10.8 | 95% CI: 6.9-15.7 | Age, Occupation, Other, Sex | 23 | 214 | 214 | | Lukanga swamps, Central Province of Zambia | Oct 2016 - Nov 2016 | Community | Other | Chisenga (2020) |

Table B.10: Overview of extracted seroprevalence parameters.

##### B.7.5 Risk factors

| Risk factor name | Statistically significant | Adjusted | Sample size | Location | Dates | Population sample type | Population group | Reference |
|---|---|---|---|---|---|---|---|---|
| **Infection** | | | | | | | | |
| Age | Significant | Not Adjusted | | Guadeloupe, Martinique (France) | 06 Jan 2016 | Hospital | Persons Under Investigation | Lannuzel (2019) |
| Age | Significant | Unspecified | | Managua (Nicaragua) | 2016 - 2016 | Population | General Population | Counotte (2019) |
| Age, Other | Significant | Adjusted | 362 | Puerto Rico: Ponce, San Juan, Guayama (United States) | May 2016 - Jul 2017 | Household | Household Contacts Of Survivors | Rosenberg (2019) |
| Age, Other | Significant | Adjusted | 805 | Narathiwat and Surat Thani (Thailand) | Mar 2018 - Mar 2019 | Household | Mixed Groups | Densathaporn (2020) |
| Age, Other | Significant | Not Adjusted | 1717 | Thailand | Jan 2016 - Dec 2017 | Population | General Population | Ruchusatsawat (2019) |
| Age, Other, Sex | Significant | Adjusted | 4090 | Ponce, Puerto Rico (United States) | Apr 2018 - May 2019 | Household | General Population | Adams (2022) |
| Age, Other, Sex | Significant | Not Adjusted | 467 | Portoviejo, Manabi (Ecuador) | 01 Jan 2016 - 31 Aug 2016 | Hospital | General Population | Fors (2018) |
| Age, Other, Sex | Significant | Unspecified | 106033 | Colombia | Aug 2015 - Jun 2017 | Population | Persons Under Investigation | Charniga (2021a) |
| Age, Prior immunity to arboviruses, Sex | Significant | Not Adjusted | 3296 | Managua (Nicaragua) | 01 Jan 2016 - 28 Feb 2017 | Community | Children | Gordon (2019) |
| Age, Sex | Significant | Adjusted | 1936 | Girardot (Colombia) | 19 Oct 2015 - 22 Jan 2016 | Hospital | Persons Under Investigation | Rojas (2016) |
| Age, Sex | Significant | Adjusted | 928 | San Andrés (Colombia) | 06 Sep 2015 - 30 Jan 2016 | Hospital | Persons Under Investigation | Rojas (2016) |
| Age, Sex | Significant | Adjusted | 3296 | Managua (Nicaragua) | 01 Jan 2016 - 28 Feb 2017 | Community | Children | Gordon (2019) |
| Infection during pregnancy, Prior immunity to arboviruses | Significant | Adjusted | 225 | León (Nicaragua) | Jan 2016 - Aug 2017 | Hospital | Persons Under Investigation | Bowman (2021) |
| Other | Significant | | | Fiji | Sep 2013 - Nov 2015 | Population | Persons Under Investigation | Kama (2019) |
| Other | Significant | Adjusted | 367 | San Juan, Puerto Rico (United States) | 16 Sep 2016 - 27 Oct 2016 | Contact | Other | Lozier (2018) |
| Other | Significant | Adjusted | | Multi-country: Americas (n = 19), Europe (n = 8), Asia (n = 1), Oceania (n = 4) | 2015 - 2016 | Unspecified | Unspecified | Nah (2016) |
| Other | Significant | Adjusted | 142 | Managua (Nicaragua) | 31 Aug 2016 - 21 Oct 2016 | Household | Household Contacts Of Survivors | Burger-Calderon (2018) |
| Other | Significant | Adjusted | | Colombia | Sep 2015 - Jul 2016 | Population | General Population | Kellemen (2021) |

continued on next page

continued from previous page

| Risk factor name | Statistically significant | Adjusted | Sample size | Location | Dates | Population sample type | Population group | Reference |
|---|---|---|---|---|---|---|---|---|
| Other | Significant | Adjusted | | Rio de Janeiro (Brazil) | 2015 - 2016 | Population | General Population | Queiroz (2021) |
| Other | Significant | Adjusted | | Unspecified | 2016 | | | Weinstein (2020) |
| Other | Significant | Adjusted | 602 | Madrid (Spain) | Jan 2016 - Jan 2017 | Hospital | Persons Under Investigation | Crespillo-Andujar (2020) |
| Other | Significant | Adjusted | 7029 | Barranquilla (Colombia) | 2014 - 2016 | Population | General Population | Mchale (2019) |
| Other | Significant | Adjusted | 12823 | Mexico | Jan 2012 - Dec 2020 | Population | Persons Under Investigation | Bukhari (2023) |
| Other | Significant | Adjusted | 3493 | Cayenne, Kourou, Saint Laurent (Guyana) | Jan 2016 - Dec 2016 | Hospital | Pregnant Women | Hallet (2020) |
| Other | Significant | Adjusted | 739 | St. Croix, St. John, St. Thomas (Virgin Islands (U.S.)) | 03 Jan 2016 - 24 Jan 2018 | Population | Persons Under Investigation | Browne (2022) |
| Other | Significant | Adjusted | 1031 | St. Thomas (Virgin Islands (U.S.)) | 03 Jan 2016 - 24 Jan 2018 | Population | Persons Under Investigation | Browne (2022) |
| Other | Significant | Adjusted | 1031 | St. Croix (Virgin Islands (U.S.)) | 03 Jan 2016 - 24 Jan 2018 | Population | Persons Under Investigation | Browne (2022) |
| Other | Significant | Adjusted | 1031 | St. John (Virgin Islands (U.S.)) | 03 Jan 2016 - 24 Jan 2018 | Population | Persons Under Investigation | Browne (2022) |
| Other | Significant | Adjusted | 865 | Unspecified | Jan 2016 - Jul 2019 | Other | Pregnant Women | Vouga (2021) |
| Other | Significant | Not Adjusted | 362 | Puerto Rico: Ponce, San Juan, Guayama (United States) | May 2016 - Jul 2017 | Household | Household Contacts Of Survivors | Rosenberg (2019) |
| Other | Significant | Not Adjusted | 367 | San Juan, Puerto Rico (United States) | 16 Sep 2016 - 27 Oct 2016 | Contact | Other | Lozier (2018) |
| Other | Significant | Not Adjusted | 4533 | Multi-country: Americas (n = 21) | 15 Aug 2015 - 11 Jun 2016 | Population | Persons Under Investigation | Ogden (2017) |
| Other | Significant | Not Adjusted | | Manaus (Brazil) | Unspecified | Population | General Population | Giovanetti (2020) |
| Other | Significant | Not Adjusted | 382 | Goiania (Brazil) | 01 Jan 2016 - 31 Dec 2017 | Population | Pregnant Women | Rosado (2022) |
| Other | Significant | Not Adjusted | 144 | Recife (Brazil) | Dec 2015 - Apr 2017 | Community | Pregnant Women | Lobkowicz (2021) |
| Other | Significant | Not Adjusted | 3493 | Cayenne, Kourou, Saint Laurent (Guyana) | Jan 2016 - Dec 2016 | Hospital | Pregnant Women | Hallet (2020) |
| Other | Significant | Not Adjusted | 189 | Dominican Republic | 2016 | Travel | Other | Voss (2020) |

continued on next page

continued from previous page

| Risk factor name | Statistically significant | Adjusted | Sample size | Location | Dates | Population sample type | Population group | Reference |
|---|---|---|---|---|---|---|---|---|
| Other | Significant | Not Adjusted | 751 | Bangkok, Samut Prakan, Samut Sakhon, Ratchaburi, Chon Buri (Thailand) | Mar 2020 - Mar 2023 | Hospital | Persons Under Investigation | Khongwichit (2023) |
| Other | Significant | Unspecified | | Rio de Janeiro (Brazil) | Feb 2015 - May 2016 | Population | Persons Under Investigation | Fuller (2017) |
| Other | Significant | Unspecified | 270 | Miami-Dade County, Florida (United States) | Jul 2016 - Dec 2016 | Community | General Population | Ajelli (2017) |
| Other | Significant | Unspecified | 1289 | Fortaleza (Brazil) | Feb 2018 - Dec 2018 | Community | Other | Frota (2023) |
| Other, Sex | Significant | Unspecified | 21468 | Puerto Rico (United States) | 03 Apr 2016 - 12 Aug 2016 | Community | Other | Chevalier (2017) |
| Age | Not Significant | Not Adjusted | 3493 | Cayenne, Kourou, Saint Laurent (Guyana) | Jan 2016 - Dec 2016 | Hospital | Pregnant Women | Hallet (2020) |
| Age | Not Significant | Unspecified | 21468 | Puerto Rico (United States) | 03 Apr 2016 - 12 Aug 2016 | Community | Other | Chevalier (2017) |
| Age, Comorbidity, Other, Sex | Not Significant | Adjusted | 367 | San Juan, Puerto Rico (United States) | 16 Sep 2016 - 27 Oct 2016 | Contact | Other | Lozier (2018) |
| Age, Comorbidity, Other, Sex | Not Significant | Not Adjusted | 367 | San Juan, Puerto Rico (United States) | 16 Sep 2016 - 27 Oct 2016 | Contact | Other | Lozier (2018) |
| Age, Household contact, Other, Sex | Not Significant | Adjusted | 142 | Managua (Nicaragua) | 31 Aug 2016 - 21 Oct 2016 | Household | Household Contacts Of Survivors | Burger-Calderon (2018) |
| Age, Occupation, Other | Not Significant | Adjusted | | Buhigwe (Tanzania) | Apr 2018 - Nov 2018 | Population | Mixed Groups | Mwanyika (2021) |
| Age, Occupation, Other | Not Significant | Not Adjusted | | Buhigwe (Tanzania) | Apr 2018 - Nov 2018 | Population | Mixed Groups | Mwanyika (2021) |
| Age, Occupation, Other, Sex | Not Significant | Not Adjusted | 362 | Puerto Rico: Ponce, San Juan, Guayama (United States) | May 2016 - Jul 2017 | Household | Household Contacts Of Survivors | Rosenberg (2019) |
| Age, Other | Not Significant | Adjusted | 3493 | Cayenne, Kourou, Saint Laurent (Guyana) | Jan 2016 - Dec 2016 | Hospital | Pregnant Women | Hallet (2020) |
| Age, Other | Not Significant | Not Adjusted | 547 | Dallas, Texas (United States) | 14 Mar 2016 - 01 Oct 2016 | Travel | Pregnant Women | Adhikari (2017) |
| Age, Other | Not Significant | Not Adjusted | 135 | Diamantina (Brazil) | 2018 - 2019 | Population | Pregnant Women | Santos (2023) |
| Age, Other, Sex | Not Significant | Adjusted | 225 | León (Nicaragua) | Jan 2016 - Aug 2017 | Hospital | Persons Under Investigation | Bowman (2021) |

continued on next page

continued from previous page

| Risk factor name | Statistically significant | Adjusted | Sample size | Location | Dates | Population sample type | Population group | Reference |
|---|---|---|---|---|---|---|---|---|
| Age, Other, Sex | Not Significant | Unspecified | 751 | Bangkok, Samut Prakan, Samut Sakhon, Ratchaburi, Chon Buri (Thailand) | Mar 2020 - Mar 2023 | Hospital | Persons Under Investigation | Khongwichit (2023) |
| Age, Prior immunity to arboviruses | Not Significant | Adjusted | 2749 | Rio de Janeiro (Brazil) | Jul 2018 - Oct 2018 | Population | General Population | Perisse (2020) |
| Age, Sex | Not Significant | Unspecified | 106033 | Colombia | Aug 2015 - Jun 2017 | Population | Persons Under Investigation | Charniga (2021a) |
| Household contact, Other | Not Significant | Adjusted | 805 | Narathiwat and Surat Thani (Thailand) | Mar 2018 - Mar 2019 | Household | Mixed Groups | Densathaporn (2020) |
| Other | Not Significant | | | Fiji | Sep 2013 - Nov 2015 | Population | Persons Under Investigation | Kama (2019) |
| Other | Not Significant | Adjusted | 362 | Puerto Rico: Ponce, San Juan, Guayama (United States) | May 2016 - Jul 2017 | Household | Household Contacts Of Survivors | Rosenberg (2019) |
| Other | Not Significant | Adjusted | | Multi-country: Americas (n = 19), Europe (n = 8), Asia (n = 1), Oceania (n = 4) | 2015 - 2016 | Unspecified | Unspecified | Nah (2016) |
| Other | Not Significant | Adjusted | | Colombia | Sep 2015 - Jul 2016 | Population | General Population | Kellemen (2021) |
| Other | Not Significant | Adjusted | | Rio de Janeiro (Brazil) | 2015 - 2016 | Population | General Population | Queiroz (2021) |
| Other | Not Significant | Adjusted | | Centre Hospitalier de l'Ouest Guyanais (CHOG, Saint-Laurent-du-Maroni, in French Guiana) (France) | Jan 2015 - Jul 2016 | Hospital | Pregnant Women | Pomar (2021) |
| Other | Not Significant | Adjusted | 12823 | Mexico | Jan 2012 - Dec 2020 | Population | Persons Under Investigation | Bukhari (2023) |
| Other | Not Significant | Adjusted | 4090 | Ponce, Puerto Rico (United States) | Apr 2018 - May 2019 | Household | General Population | Adams (2022) |
| Other | Not Significant | Adjusted | 1031 | St. Thomas (Virgin Islands (U.S.)) | 03 Jan 2016 - 24 Jan 2018 | Population | Persons Under Investigation | Browne (2022) |
| Other | Not Significant | Adjusted | 1031 | St. Croix (Virgin Islands (U.S.)) | 03 Jan 2016 - 24 Jan 2018 | Population | Persons Under Investigation | Browne (2022) |
| Other | Not Significant | Adjusted | 1031 | St. John (Virgin Islands (U.S.)) | 03 Jan 2016 - 24 Jan 2018 | Population | Persons Under Investigation | Browne (2022) |

continued on next page

continued from previous page

| Risk factor name | Statistically significant | Adjusted | Sample size | Location | Dates | Population sample type | Population group | Reference |
|---|---|---|---|---|---|---|---|---|
| Other | Not Significant | Not Adjusted | 2659 | Managua (Nicaragua) | 01 Jan 2016 - 28 Feb 2017 | Community | Children | Gordon (2019) |
| Other | Not Significant | Not Adjusted | | Centre Hospitalier de l'Ouest Guyanais (CHOG, Saint-Laurent-du-Maroni, in French Guiana) (France) | Jan 2015 - Jul 2016 | Hospital | Pregnant Women | Pomar (2021) |
| Other | Not Significant | Not Adjusted | 189 | Dominican Republic | 2016 | Travel | Other | Voss (2020) |
| Other | Not Significant | Not Adjusted | | Guadeloupe, Martinique (France) | 06 Jan 2016 | Hospital | Persons Under Investigation | Lannuzel (2019) |
| Other, Prior immunity to arboviruses | Not Significant | Adjusted | 2659 | Managua (Nicaragua) | 01 Jan 2016 - 28 Feb 2017 | Community | Children | Gordon (2019) |
| Other, Sex | Not Significant | Adjusted | 7029 | Barranquilla (Colombia) | 2014 - 2016 | Population | General Population | Mchale (2019) |
| Other, Sex | Not Significant | Not Adjusted | 1717 | Thailand | Jan 2016 - Dec 2017 | Population | General Population | Ruchusatsawat (2019) |
| Other | Unspecified | Unspecified | 10595 | Australia | Jul 2013 - Jun 2014 | Other | Blood Donors | Coghlan (2018) |
| Sex | Unspecified | Unspecified | | United States | 2016 - 2017 | Mixed | Mixed Groups | Major (2021) |
| **Serology** | | | | | | | | |
| Age | Significant | Adjusted | 845 | Sudan | 2012 - 2012 | Community | Other | Soghaier (2018) |
| Age | Significant | Not Adjusted | | District II, Managua (Nicaragua) | Feb 2017 - Jul 2017 | Household | Other | Zambrana (2018) |
| Age | Significant | Not Adjusted | 1430 | Mali | 2013 - Nov 2016 | Population | General Population | Diarra (2020) |
| Age | Significant | Unspecified | 801 | Krong Buk (Vietnam) | Jan 2017 - Jul 2018 | Community | Mixed Groups | Nguyen (2020) |
| Age, Comorbidity, Other | Significant | Not Adjusted | 871 | Abia State University Teaching Hospital, Aba, Abia state, Federal Medical Centre, Keffi, Nasarawa State, Baru-Dokp Teaching Hospital, Kaduna, Kaduna State (Nigeria) | Dec 2020 - Nov 2021 | Hospital | Mixed Groups | Mac (2023a) |
| Age, Other | Significant | Adjusted | 1295 | Thailand | Dec 1997 - Dec 2017 | Population | Pregnant Women | Harapan (2022) |
| Age, Other | Significant | Adjusted | 469 | Salvador (Hospital Geral Roberto Santos) | Oct 2015 | Population | Pregnant Women | Nery (2021) |
| Age, Other | Significant | Adjusted | | Coastal lowlands of SW Guatemala (Guatemala) | 2015 - 2016 | Population | Children | Lamb (2022) |

continued on next page

continued from previous page

| Risk factor name | Statistically significant | Adjusted | Sample size | Location | Dates | Population sample type | Population group | Reference |
|---|---|---|---|---|---|---|---|---|
| Age, Other | Significant | Adjusted | 1192 | Borbón, Maldonado, Timbiré, Santa Maria, Santo Domingo, Colon Eloy (Ecuador) | Jul 2018 - Oct 2018 | Community | General Population | Andrade (2024) |
| Age, Other | Significant | Not Adjusted | 501 | Ouagadougou, Bobo-Dioulasso (Burkina Faso) | 2020 - 2020 | Population | Blood Donors | Tinto (2022) |
| Age, Other | Significant | Not Adjusted | 1295 | Thailand | Dec 1997 - Dec 2017 | Population | Pregnant Women | Harapan (2022) |
| Age, Other | Significant | Not Adjusted | 469 | Salvador (Hospital Geral Roberto Santos) | Oct 2015 | Population | Pregnant Women | Nery (2021) |
| Age, Other | Significant | Not Adjusted | 455 | Unspecified | Mar 2014 - Oct 2017 | Travel | General Population | Overbosch (2023) |
| Age, Other | Significant | Not Adjusted | 585 | "Peninsula" (Malaysia) | Unspecified | Population | General Population | Khor (2024) |
| Age, Other | Significant | Not Adjusted | 1192 | Borbón, Maldonado, Timbiré, Santa Maria, Santo Domingo, Colon Eloy (Ecuador) | Jul 2018 - Oct 2018 | Community | General Population | Andrade (2024) |
| Age, Other | Significant | Unspecified | | Malaysia | Unspecified | Community | General Population | Khor (2020) |
| Age, Other, Prior immunity to arboviruses | Significant | Adjusted | | Recife (Brazil) | Aug 2018 - Feb 2019 | Household | General Population | Braga (2023) |
| Age, Other, Prior immunity to arboviruses | Significant | Adjusted | 1126 | Borbón, Maldonado, Timbiré, Santa Maria, Santo Domingo, Colon Eloy (Ecuador) | Aug 2019 - Oct 2019 | Community | General Population | Andrade (2024) |
| Age, Other, Prior immunity to arboviruses | Significant | Not Adjusted | | Recife (Brazil) | Aug 2018 - Feb 2019 | Household | General Population | Braga (2023) |
| Age, Other, Sex | Significant | Not Adjusted | | District II, Managua (Nicaragua) | Feb 2017 - Jul 2017 | Household | Children | Zambrana (2018) |
| Age, Sex | Significant | Adjusted | | District II, Managua (Nicaragua) | Feb 2017 - Jul 2017 | Household | Other | Zambrana (2018) |
| Age, Sex | Significant | Adjusted | 1085 | Kuala Lumpur (Malaysia) | 2012 - 2017 | Mixed | Mixed Groups | Sam (2019) |
| Household contact, Other | Significant | Adjusted | 425 | Pernambuco State (Brazil) | Apr 2017 - Dec 2017 | Household | Mixed Groups | Magalhaes (2021) |
| Household contact, Other | Significant | Not Adjusted | 425 | Pernambuco State (Brazil) | Apr 2017 - Dec 2017 | Household | Mixed Groups | Magalhaes (2021) |
| Infection | Significant | Unspecified | 328 | Santander (Colombia) | Unspecified | School | Mixed Groups | Cardenas (2020) |
| Occupation, Other | Significant | Adjusted | 1084 | Maroua, Garoua, Ngaoundere, Douala, Yaounde, Bertoua (Cameroon) | Aug 2015 - Oct 2015 | Other | Blood Donors | Gake (2017) |

continued on next page

continued from previous page

| Risk factor name | Statistically significant | Adjusted | Sample size | Location | Dates | Population sample type | Population group | Reference |
|---|---|---|---|---|---|---|---|---|
| Occupation, Other, Sex | Significant | Adjusted | 150 | Itang special district, Lare district (Ethiopia) | Oct 2018 - Jun 2019 | Community | General Population | Asebe (2021) |
| Other | Significant | Adjusted | 100 | Cross River State (Nigeria) | 17 Mar 2017 - 30 May 2017 | Population | Persons Under Investigation | Otu (2020) |
| Other | Significant | Adjusted | 359 | Salvador, Bahia (Brazil) | Aug 2015 - Jul 2016 | Population | Persons Under Investigation | Ticona (2021) |
| Other | Significant | Adjusted | 94 | Maranhao state (Brazil) | 2016 - 2018 | Hospital | Persons Under Investigation | Ribeiro (2020) |
| Other | Significant | Adjusted | 33 | West Pokot (Kenya) | Feb 2016 - Feb 2016 | Population | General Population | Chepkorir (2019) |
| Other | Significant | Adjusted | 2697 | French Guiana (France) | Jun 2017 - Oct 2017 | Population | General Population | Bailly (2021) |
| Other | Significant | Adjusted | 496 | Adamawa, Bauchi, and Borno states (Nigeria) | Apr 2018 - Aug 2018 | Population | Persons Under Investigation | Baba (2023) |
| Other | Significant | Adjusted | 871 | Abia, Kaduna, Nasarawa (Nigeria) | Dec 2020 - Nov 2021 | Hospital | Mixed Groups | Peter Asaga (2023) |
| Other | Significant | Not Adjusted | 642 | Salvador, Bahia (Brazil) | Oct 2015 - Oct 2015 | Population | Other | Rodriguez-Barraquer (2019) |
| Other | Significant | Not Adjusted | 410 | Saudi Arabia | Nov 2016 - Jul 2017 | Hospital | Pregnant Women | Alayed (2018) |
| Other | Significant | Not Adjusted | 2697 | 22 municipalities of French Guiana (France) | Jun 2017 - Oct 2017 | Population | General Population | Flamand (2019) |
| Other | Significant | Not Adjusted | 359 | Salvador, Bahia (Brazil) | Aug 2015 - Jul 2016 | Population | Persons Under Investigation | Ticona (2021) |
| Other | Significant | Not Adjusted | 469 | Salvador (Hospital Geral Roberto Santos) | Oct 2015 | Population | Pregnant Women | Nery (2021) |
| Other | Significant | Not Adjusted | | Salvador, Bahia (Brazil) | 01 Jan 2015 - 31 Jan 2017 | Hospital | Pregnant Women | Aromolaran (2022) |
| Other | Significant | Not Adjusted | 496 | Adamawa, Bauchi, and Borno states (Nigeria) | Apr 2018 - Aug 2018 | Population | Persons Under Investigation | Baba (2023) |
| Other | Significant | Unspecified | 1289 | Fortaleza (Brazil) | Feb 2018 - Dec 2018 | Community | Other | Frota (2023) |
| Other, Prior immunity to arboviruses | Significant | Not Adjusted | 431 | Santiago Island: Praia (Cabo Verde) | 24 Aug 2014 - 04 Nov 2014 | Population | General Population | Ward (2022) |
| Other, Sex | Significant | Adjusted | | District II, Managua (Nicaragua) | Feb 2017 - Jul 2017 | Household | Children | Zambrana (2018) |
| Other, Sex | Significant | Not Adjusted | 468 | Plateau, Nasarawa (Nigeria) | Jan 2016 - Dec 2016 | Community | General Population | Mathe (2018) |
| Other, Sex | Significant | Unspecified | 557 | Yap Island (Federated States of Micronesia) | 01 Apr 2007 - 31 Jul 2007 | Household | General Population | Duffy (2009) |
| Prior immunity to arboviruses | Significant | Adjusted | 642 | Salvador, Bahia (Brazil) | Oct 2015 - Oct 2015 | Population | Other | Rodriguez-Barraquer (2019) |

continued on next page

continued from previous page

| Risk factor name | Statistically significant | Adjusted | Sample size | Location | Dates | Population sample type | Population group | Reference |
|---|---|---|---|---|---|---|---|---|
| Prior immunity to arboviruses | Significant | Not Adjusted | 642 | Salvador, Bahia (Brazil) | Oct 2015 - Oct 2015 | Population | Other | Rodriguez-Barraquer (2019) |
| Sex | Significant | Unspecified | 153 | United States | Unspecified | Population | Persons Under Investigation | Hills (2021) |
| Age | Not Significant | Adjusted | 642 | Salvador, Bahia (Brazil) | Oct 2015 - Oct 2015 | Population | Other | Rodriguez-Barraquer (2019) |
| Age, Infection during pregnancy, Other, Sex | Not Significant | Adjusted | 871 | Abia, Kaduna, Nasarawa (Nigeria) | Dec 2020 - Nov 2021 | Hospital | Mixed Groups | Peter Asaga (2023) |
| Age, Occupation, Other, Prior immunity to arboviruses, Sex | Not Significant | Adjusted | 33 | West Pokot (Kenya) | Feb 2016 - Feb 2016 | Population | General Population | Chepkorir (2019) |
| Age, Other | Not Significant | Adjusted | | District II, Managua (Nicaragua) | Feb 2017 - Jul 2017 | Household | Children | Zambrana (2018) |
| Age, Other | Not Significant | Adjusted | | District II, Managua (Nicaragua) | Feb 2017 - Jul 2017 | Household | Other | Zambrana (2018) |
| Age, Other | Not Significant | Adjusted | 1295 | Thailand | Dec 1997 - Dec 2017 | Population | Pregnant Women | Harapan (2022) |
| Age, Other | Not Significant | Adjusted | | Recife (Brazil) | Aug 2018 - Feb 2019 | Household | General Population | Braga (2023) |
| Age, Other | Not Significant | Adjusted | 150 | Itang special district, Lare district (Ethiopia) | Oct 2018 - Jun 2019 | Community | General Population | Asebe (2021) |
| Age, Other | Not Significant | Adjusted | 1192 | Borbón, Maldonado, Timbiré, Santa Maria, Santo Domingo, Colon Eloy (Ecuador) | Jul 2018 - Oct 2018 | Community | General Population | Andrade (2024) |
| Age, Other | Not Significant | Not Adjusted | 1295 | Thailand | Dec 1997 - Dec 2017 | Population | Pregnant Women | Harapan (2022) |
| Age, Other | Not Significant | Unspecified | 557 | Yap Island (Federated States of Micronesia) | 01 Apr 2007 - 31 Jul 2007 | Household | General Population | Duffy (2009) |
| Age, Other | Not Significant | Unspecified | 153 | United States | Unspecified | Population | Persons Under Investigation | Hills (2021) |
| Age, Other | Not Significant | Unspecified | 650 | Siriraj Hospital (Thailand) | May 2019 - Oct 2019 | Hospital | Pregnant Women | Phatihattakorn (2021) |
| Age, Other, Prior immunity to arboviruses, Sex | Not Significant | Not Adjusted | | Recife (Brazil) | Aug 2018 - Feb 2019 | Household | General Population | Braga (2023) |
| Age, Other, Sex | Not Significant | Adjusted | 845 | Sudan | 2012 - 2012 | Community | Other | Soghaier (2018) |
| Age, Other, Sex | Not Significant | Adjusted | 100 | Cross River State (Nigeria) | 17 Mar 2017 - 30 May 2017 | Population | Persons Under Investigation | Otu (2020) |
| Age, Other, Sex | Not Significant | Adjusted | 2697 | French Guiana (France) | Jun 2017 - Oct 2017 | Population | General Population | Bailly (2021) |
| Age, Other, Sex | Not Significant | Adjusted | 496 | Adamawa, Bauchi, and Borno states (Nigeria) | Apr 2018 - Aug 2018 | Population | Persons Under Investigation | Baba (2023) |

continued on next page

continued from previous page

| Risk factor name | Statistically significant | Adjusted | Sample size | Location | Dates | Population sample type | Population group | Reference |
|---|---|---|---|---|---|---|---|---|
| Age, Other, Sex | Not Significant | Adjusted | 1126 | Borbón, Maldonado, Timbiré, Santa Maria, Santo Domingo, Colon Eloy (Ecuador) | Aug 2019 - Oct 2019 | Community | General Population | Andrade (2024) |
| Age, Other, Sex | Not Significant | Not Adjusted | | District II, Managua (Nicaragua) | Feb 2017 - Jul 2017 | Household | Other | Zambrana (2018) |
| Age, Other, Sex | Not Significant | Not Adjusted | 642 | Salvador, Bahia (Brazil) | Oct 2015 - Oct 2015 | Population | Other | Rodriguez-Barraquer (2019) |
| Age, Other, Sex | Not Significant | Not Adjusted | 2697 | 22 municipalities of French Guiana (France) | Jun 2017 - Oct 2017 | Population | General Population | Flamand (2019) |
| Age, Other, Sex | Not Significant | Not Adjusted | 431 | Santiago Island: Praia (Cabo Verde) | 24 Aug 2014 - 04 Nov 2014 | Population | General Population | Ward (2022) |
| Age, Other, Sex | Not Significant | Not Adjusted | 496 | Adamawa, Bauchi, and Borno states (Nigeria) | Apr 2018 - Aug 2018 | Population | Persons Under Investigation | Baba (2023) |
| Age, Other, Sex | Not Significant | Not Adjusted | 1192 | Borbón, Maldonado, Timbiré, Santa Maria, Santo Domingo, Colon Eloy (Ecuador) | Jul 2018 - Oct 2018 | Community | General Population | Andrade (2024) |
| Age, Other, Sex | Not Significant | Unspecified | 328 | Santander (Colombia) | Unspecified | School | Mixed Groups | Cardenas (2020) |
| Age, Other, Sex | Not Significant | Unspecified | 850 | Mozambique | 2009 - 2015 | Population | Other | Chelene (2019) |
| Age, Prior immunity to arboviruses | Not Significant | Not Adjusted | 642 | Salvador, Bahia (Brazil) | Oct 2015 - Oct 2015 | Population | Other | Rodriguez-Barraquer (2019) |
| Age, Sex | Not Significant | Unspecified | 814 | Beni, Santa Cruz, Tarija, Cochabamba, La Paz (Bolivia) | Dec 2016 - Apr 2017 | Population | General Population | Villarroel (2018) |
| Comorbidity, Prior immunity to arboviruses, Sex | Not Significant | Not Adjusted | 468 | Plateau, Nasarawa (Nigeria) | Jan 2016 - Dec 2016 | Community | General Population | Mathe (2018) |
| Occupation, Other | Not Significant | Not Adjusted | 254 | Thailand | Dec 2017 - Feb 2020 | Population | General Population | Kitro (2024) |
| Other | Not Significant | Adjusted | 1085 | Kuala Lumpur (Malaysia) | 2012 - 2017 | Mixed | Mixed Groups | Sam (2019) |
| Other | Not Significant | Adjusted | 359 | Salvador, Bahia (Brazil) | Aug 2015 - Jul 2016 | Population | Persons Under Investigation | Ticona (2021) |
| Other | Not Significant | Not Adjusted | | District II, Managua (Nicaragua) | Feb 2017 - Jul 2017 | Household | Children | Zambrana (2018) |
| Other | Not Significant | Not Adjusted | 359 | Salvador, Bahia (Brazil) | Aug 2015 - Jul 2016 | Population | Persons Under Investigation | Ticona (2021) |
| Other | Not Significant | Not Adjusted | 469 | Salvador (Hospital Geral Roberto Santos) | Oct 2015 | Population | Pregnant Women | Nery (2021) |

continued on next page

continued from previous page

| Risk factor name | Statistically significant | Adjusted | Sample size | Location | Dates | Population sample type | Population group | Reference |
|---|---|---|---|---|---|---|---|---|
| Other, Sex | Not Significant | Not Adjusted | 871 | Abia State University Teaching Hospital, Aba, Abia state, Federal Medical Centre, Keffi, Nasarawa State, Baru-Dokp Teaching Hospital, Kaduna, Kaduna State (Nigeria) | Dec 2020 - Nov 2021 | Hospital | Mixed Groups | Mac (2023a) |
| Other, Sex | Not Significant | Not Adjusted | | Coastal lowlands of SW Guatemala (Guatemala) | 2015 - 2016 | Population | Children | Lamb (2022) |
| Other, Sex | Not Significant | Not Adjusted | 455 | Unspecified | Mar 2014 - Oct 2017 | Travel | General Population | Overbosch (2023) |
| Other, Sex | Not Significant | Unspecified | 801 | Krong Buk (Vietnam) | Jan 2017 - Jul 2018 | Community | Mixed Groups | Nguyen (2020) |
| Sex | Not Significant | Not Adjusted | | Vientiane capital (Laos) | 2003 - 2015 | Population | Blood Donors | Pastorino (2019) |
| Sex | Not Significant | Not Adjusted | 501 | Ouagadougou, Bobo-Dioulasso (Burkina Faso) | 2020 - 2020 | Population | Blood Donors | Tinto (2022) |
| Sex | Not Significant | Not Adjusted | 1430 | Mali | 2013 - Nov 2016 | Population | General Population | Diarra (2020) |
| Sex | Not Significant | Not Adjusted | 585 | "Peninsula" (Malaysia) | Unspecified | Population | General Population | Khor (2024) |
| **Guillain Barre syndrome** | | | | | | | | |
| Infection | Significant | Not Adjusted | 281 | 9 unspecified states (Mexico) | 01 Jul 2016 - 30 Jun 2018 | Hospital | Persons Under Investigation | Grijalva (2020) |
| Infection, Other | Significant | Adjusted | 40 | Barranquilla (Colombia) | 01 Oct 2015 - 02 Apr 2016 | Hospital | Other | Salinas (2017) |
| Infection, Other | Significant | Not Adjusted | 41 | Salvador, Bahia (Brazil) | 01 Jan 2015 - 31 Aug 2015 | Hospital | Other | Styczynski (2017) |
| Occupation, Other | Significant | Not Adjusted | 29 | Cucuta (Colombia) | 29 Jun 2015 - 30 Jul 2016 | Hospital | Other | Anaya (2017) |
| Other | Significant | Unspecified | 191 | Monterrey City (Mexico) | 01 Aug 2017 - 30 Jun 2018 | Hospital | Persons Under Investigation | Gongora-Rivera (2020) |
| Comorbidity, Occupation, Other, Sex | Not Significant | Not Adjusted | 29 | Cucuta (Colombia) | 29 Jun 2015 - 30 Jul 2016 | Hospital | Other | Anaya (2017) |
| Comorbidity, Other, Sex | Not Significant | Not Adjusted | 41 | Salvador, Bahia (Brazil) | 01 Jan 2015 - 31 Aug 2015 | Hospital | Other | Styczynski (2017) |
| Infection | Not Significant | Unspecified | 191 | Monterrey City (Mexico) | 01 Aug 2017 - 30 Jun 2018 | Hospital | Persons Under Investigation | Gongora-Rivera (2020) |
| Other | Not Significant | Not Adjusted | | Bangladesh | 2011 - 2015 | Population | Mixed Groups | Geurtsvankessel (2018) |
| **Hospitalisation** | | | | | | | | |

continued on next page

continued from previous page

| Risk factor name | Statistically significant | Adjusted | Sample size | Location | Dates | Population sample type | Population group | Reference |
|---|---|---|---|---|---|---|---|---|
| Age, Comorbidity, Other | Significant | Not Adjusted | 94 | Veterans Health Administration in US and Caribbean | 01 Dec 2015 - 31 Oct 2016 | Hospital | Other | Schirmer (2018) |
| **Low birthweight** | | | | | | | | |
| Infection, Infection during pregnancy | Significant | Adjusted | 291 | French Guiana (France) | 01 Jan 2015 - 15 Jul 2016 | Mixed | Children | Pomar (2020) |
| Infection during pregnancy | Not Significant | Not Adjusted | 745 | Manaus (Brazil) | 2015 - 2016 | Community | Pregnant Women | De Fatima Redivo (2020) |
| **Microcephaly** | | | | | | | | |
| Infection during pregnancy | Significant | Not Adjusted | | French Polynesia (France) | Sep 2013 - Jul 2015 | Population | Pregnant Women | Cauchemez (2016) |
| Infection during pregnancy | Significant | Not Adjusted | 109 | Tangará da Serra (Brazil) | 01 Jan 2016 - 31 Dec 2016 | Other | Children | Herrero Da Silva (2022) |
| Infection during pregnancy | Significant | Not Adjusted | 745 | Manaus (Brazil) | 2015 - 2016 | Community | Pregnant Women | De Fatima Redivo (2020) |
| Infection during pregnancy | Significant | Unspecified | 108 | Maranhao state (Brazil) | Mar 2015 - Sep 2018 | Contact | Pregnant Women | Mendes (2020) |
| Infection during pregnancy, Occupation | Significant | Adjusted | 64 | Aracaju city in Sergipe (Brazil) | 01 Sep 2015 - 05 Jan 2016 | Hospital | Children | Santa Rita (2017) |
| Infection during pregnancy, Other | Significant | Adjusted | 3603823 | Brazil | 01 Jan 2015 - 23 May 2017 | Population | Children | Brady (2019) |
| Infection, Other | Significant | Adjusted | 191 | Paraíba (Brazil) | Jan 2015 - Dec 2016 | Community | Children | Bezerra (2023) |
| Infection, Other, Sex | Significant | Not Adjusted | 264 | Recife (Brazil) | 15 Jan 2016 - 30 Nov 2016 | Hospital | Children | De Araujo (2018) |
| Other | Significant | Adjusted | 163 | Rio de Janeiro (Brazil) | Feb 2016 - Sep 2017 | Other | Children | Power (2020) |
| Other | Significant | Not Adjusted | 555 | Martinique, Guadeloupe, French Guiana (France) | Mar 2016 - 27 Apr 2017 | Population | Pregnant Women | Hoen (2018) |
| Other | Significant | Not Adjusted | 163 | Rio de Janeiro (Brazil) | Feb 2016 - Sep 2017 | Other | Children | Power (2020) |
| Infection | Not Significant | Not Adjusted | 98 | Rio de Janeiro (Brazil) | 2016 - 2016 | Other | Children | De Melo Espindola (2021) |
| Infection during pregnancy | Not Significant | Not Adjusted | 334 | Dallas, Texas (United States) | 14 Mar 2016 - 01 Oct 2016 | Hospital | Children | Adhikari (2017) |
| Infection during pregnancy | Not Significant | Unspecified | | Unspecified | Unspecified | | | Pomar (2017) |
| Infection during pregnancy | Not Significant | Unspecified | 18 | Hawaii (United States) | 2009 - 2012 | Other | Persons Under Investigation | Kumar (2016) |
| Occupation, Other | Not Significant | Not Adjusted | 163 | Rio de Janeiro (Brazil) | Feb 2016 - Sep 2017 | Other | Children | Power (2020) |
| Other | Not Significant | Adjusted | 3603828 | Brazil | 01 Jan 2015 - 23 May 2017 | Population | Children | Brady (2019) |
| Other | Not Significant | Adjusted | 163 | Rio de Janeiro (Brazil) | Feb 2016 - Sep 2017 | Other | Children | Power (2020) |
| Other | Not Significant | Not Adjusted | 31 | Goiania (Brazil) | 01 Jan 2016 - 31 Dec 2017 | Population | Pregnant Women | Rosado (2022) |
| **Miscarriage/stillbirth** | | | | | | | | |

continued on next page

continued from previous page

| Risk factor name | Statistically significant | Adjusted | Sample size | Location | Dates | Population sample type | Population group | Reference |
|---|---|---|---|---|---|---|---|---|
| Infection during pregnancy | Significant | Adjusted | 511 | Ribeirão Preto (Brazil) | 2015 - 2016 | Population | Children | Coutinho (2021) |
| Infection during pregnancy | Significant | Adjusted | | Centre Hospitalier de l'Ouest Guyanais (CHOG, Saint-Laurent-du-Maroni, in French Guiana) (France) | Jan 2015 - Jul 2016 | Hospital | Pregnant Women | Pomar (2021) |
| Infection during pregnancy | Significant | Not Adjusted | 511 | Ribeirão Preto (Brazil) | 2015 - 2016 | Population | Children | Coutinho (2021) |
| Infection during pregnancy | Significant | Not Adjusted | | Centre Hospitalier de l'Ouest Guyanais (CHOG, Saint-Laurent-du-Maroni, in French Guiana) (France) | Jan 2015 - Jul 2016 | Hospital | Pregnant Women | Pomar (2021) |
| Infection, Infection during pregnancy | Significant | Not Adjusted | 625 | French Guiana (France) | 01 Jan 2015 - 15 Jul 2016 | Mixed | Children | Pomar (2020) |
| Infection during pregnancy | Not Significant | Not Adjusted | | Centre Hospitalier de l'Ouest Guyanais (CHOG, Saint-Laurent-du-Maroni, in French Guiana) (France) | Jan 2015 - Jul 2016 | Hospital | Pregnant Women | Pomar (2021) |
| Infection during pregnancy | Not Significant | Not Adjusted | 762 | Manaus (Brazil) | 2015 - 2016 | Community | Pregnant Women | De Fatima Redivo (2020) |
| **Other** | | | | | | | | |
| Age, Other, Prior immunity to arboviruses | Significant | Not Adjusted | 3893 | Managua (Nicaragua) | 01 Jan 2016 - 28 Feb 2017 | Community | Children | Gordon (2019) |
| Age, Other, Prior immunity to arboviruses, Sex | Significant | Adjusted | 3893 | Managua (Nicaragua) | 01 Jan 2016 - 28 Feb 2017 | Community | Children | Gordon (2019) |
| Infection | Significant | Unspecified | 59 | United States | 13 Jul 2016 - 19 Sep 2017 | Hospital | Persons Under Investigation | El Sahly (2019) |
| Infection during pregnancy | Significant | Not Adjusted | 109 | Tangará da Serra (Brazil) | 01 Jan 2016 - 31 Dec 2016 | Other | Children | Herrero Da Silva (2022) |
| Infection during pregnancy | Significant | Not Adjusted | 102 | Colombia | Oct 2015 - Jan 2017 | Hospital | Pregnant Women | Gutierrez-Sanchez (2022) |
| Infection, Infection during pregnancy | Significant | Adjusted | 625 | French Guiana (France) | 01 Jan 2015 - 15 Jul 2016 | Mixed | Children | Pomar (2020) |
| Other | Significant | Adjusted | 163 | Rio de Janeiro (Brazil) | Feb 2016 - Sep 2017 | Other | Children | Power (2020) |
| Other | Significant | Adjusted | 39331 | Rio de Janeiro (Brazil) | 2015 - 2016 | Population | General Population | Raymundo (2021) |
| Other | Significant | Adjusted | | Brazil | 01 Aug 2015 - 31 Dec 2019 | Population | Other | Pescarini (2022) |

continued on next page

continued from previous page

| Risk factor name | Statistically significant | Adjusted | Sample size | Location | Dates | Population sample type | Population group | Reference |
|---|---|---|---|---|---|---|---|---|
| Other | Significant | Adjusted | 200 | Chincha Baja and Pueblo Nuevo (Peru) | Mar 2019 - May 2019 | Population | General Population | Cachay (2021) |
| Other | Significant | Not Adjusted | 523 | Brazil | 19 Nov 2015 - 27 Feb 2016 | Population | Pregnant Women | França (2016) |
| Other | Significant | Not Adjusted | 602 | Brazil | 19 Nov 2015 - 27 Feb 2016 | Population | Pregnant Women | França (2016) |
| Other | Significant | Not Adjusted | | Queensland (Australia) | Apr 2016 | | | Hugo (2019) |
| Other | Significant | Not Adjusted | | Jundiai, Parana (Brazil) | 2015 - 2016 | Hospital | Mixed Groups | De Moraes (2022) |
| Age, Other, Sex | Not Significant | Adjusted | 3893 | Managua (Nicaragua) | 01 Jan 2016 - 28 Feb 2017 | Community | Children | Gordon (2019) |
| Age, Sex | Not Significant | Adjusted | | Arauca, Armenia, Barranquilla, Bucaramanga, Cali, Cartagena, Cúcuta, Florencia, Ibagué, Inírida, Leticia, Medellín, Mitú, Mocoa, Montería, Neiva, Pereira, Popayán, Puerto Carreño, Quibdó, Riohacha, San Andrés, San José del Guaviare, Santa Marta, Sincelejo, Valledupar, Villavicencio, Yopal (Colombia) | 2015 - 2017 | Population | General Population | Charniga (2022) |
| Infection | Not Significant | Unspecified | 59 | United States | 13 Jul 2016 - 19 Sep 2017 | Hospital | Persons Under Investigation | El Sahly (2019) |
| Infection during pregnancy | Not Significant | Adjusted | 83 | Jundiai Hospital Medical school (Brazil) | 01 Mar 2016 - 30 Jun 2019 | Hospital | Pregnant Women | Gazeta (2021) |
| Infection during pregnancy | Not Significant | Not Adjusted | 745 | Manaus (Brazil) | 2015 - 2016 | Community | Pregnant Women | De Fatima Redivo (2020) |
| Other | Not Significant | Adjusted | 163 | Rio de Janeiro (Brazil) | Feb 2016 - Sep 2017 | Other | Children | Power (2020) |
| Other | Not Significant | Not Adjusted | | Jundiai, Parana (Brazil) | 2015 - 2016 | Hospital | Mixed Groups | De Moraes (2022) |
| Sex | Not Significant | Not Adjusted | 3893 | Managua (Nicaragua) | 01 Jan 2016 - 28 Feb 2017 | Community | Children | Gordon (2019) |

continued on next page

continued from previous page

| Risk factor name | Statistically significant | Adjusted | Sample size | Location | Dates | Population sample type | Population group | Reference |
|---|---|---|---|---|---|---|---|---|
| Other | Unspecified | Not Adjusted | 5108 | Belo Horizonte, Salvador, Laranjeiras, Fortaleza, Recife, Cuiaba and Campo Grande, Porto Velho (Brazil) | Jul 2016 - Jun 2019 | Other | Other | Botosso (2023) |
| **Other neurological symptoms in general population** | | | | | | | | |
| Age, Sex | Significant | Unspecified | 418 | Colombia | Jul 2015 - Oct 2017 | Population | Other | Charniga (2021a) |
| Comorbidity, Hospitalisation | Significant | Not Adjusted | 46 | Veterans Health Administration in US and Caribbean | 01 Dec 2015 - 31 Oct 2016 | Hospital | Other | Schirmer (2018) |
| Infection | Significant | Not Adjusted | | Guadeloupe, Martinique (France) | 06 Jan 2016 | Hospital | Persons Under Investigation | Lannuzel (2019) |
| Infection during pregnancy | Significant | Adjusted | | Centre Hospitalier de l'Ouest Guyanais (CHOG, Saint-Laurent-du-Maroni, in French Guiana) (France) | Jan 2015 - Jul 2016 | Hospital | Pregnant Women | Pomar (2021) |
| Infection during pregnancy | Significant | Not Adjusted | 129 | León (Nicaragua) | Feb 2017 - Jul 2017 | Other | Children | Stringer (2021) |
| Infection during pregnancy | Significant | Not Adjusted | | Centre Hospitalier de l'Ouest Guyanais (CHOG, Saint-Laurent-du-Maroni, in French Guiana) (France) | Jan 2015 - Jul 2016 | Hospital | Pregnant Women | Pomar (2021) |
| Infection during pregnancy, Other | Significant | Adjusted | 129 | León (Nicaragua) | Feb 2017 - Jul 2017 | Other | Children | Stringer (2021) |
| Age, Other | Not Significant | Not Adjusted | 129 | León (Nicaragua) | Feb 2017 - Jul 2017 | Other | Children | Stringer (2021) |
| Age, Other, Sex | Not Significant | Adjusted | 129 | León (Nicaragua) | Feb 2017 - Jul 2017 | Other | Children | Stringer (2021) |

continued on next page

continued from previous page

| Risk factor name | Statistically significant | Adjusted | Sample size | Location | Dates | Population sample type | Population group | Reference |
|---|---|---|---|---|---|---|---|---|
| Age, Sex | Not Significant | Adjusted | | Arauca, Armenia, Barranquilla, Bucaramanga, Cali, Cartagena, Cúcuta, Florencia, Ibagué, Inírida, Leticia, Medellín, Mitú, Mocoa, Montería, Neiva, Pereira, Popayán, Puerto Carreño, Quibdó, Riohacha, San Andrés, San José del Guaviare, Santa Marta, Sincelejo, Valledupar, Villavicencio, Yopal (Colombia) | 2015 - 2017 | Population | General Population | Charniga (2022) |
| Age, Sex | Not Significant | Unspecified | 418 | Colombia | Jul 2015 - Oct 2017 | Population | Other | Charniga (2021a) |
| Infection | Not Significant | Not Adjusted | 98 | Rio de Janeiro (Brazil) | 2016 - 2016 | Other | Children | De Melo Espindola (2021) |
| Infection during pregnancy | Not Significant | Adjusted | 152 | Tegucigalpa (Honduras) | Jul 2016 - Dec 2016 | Hospital | Children | Alger (2024) |
| Infection during pregnancy | Not Significant | Not Adjusted | | Centre Hospitalier de l'Ouest Guyanais (CHOG, Saint-Laurent-du-Maroni, in French Guiana) (France) | Jan 2015 - Jul 2016 | Hospital | Pregnant Women | Pomar (2021) |
| Other | Not Significant | Adjusted | | Centre Hospitalier de l'Ouest Guyanais (CHOG, Saint-Laurent-du-Maroni, in French Guiana) (France) | Jan 2015 - Jul 2016 | Hospital | Pregnant Women | Pomar (2021) |
| Other | Not Significant | Not Adjusted | | Centre Hospitalier de l'Ouest Guyanais (CHOG, Saint-Laurent-du-Maroni, in French Guiana) (France) | Jan 2015 - Jul 2016 | Hospital | Pregnant Women | Pomar (2021) |

continued on next page

continued from previous page

| Risk factor name | Statistically significant | Adjusted | Sample size | Location | Dates | Population sample type | Population group | Reference |
|---|---|---|---|---|---|---|---|---|
| Other | Not Significant | Not Adjusted | | Guadeloupe, Martinique (France) | 06 Jan 2016 | Hospital | Persons Under Investigation | Lannuzel (2019) |
| **Premature birth** | | | | | | | | |
| Infection during pregnancy | Significant | Not Adjusted | 745 | Manaus (Brazil) | 2015 - 2016 | Community | Pregnant Women | De Fatima Redivo (2020) |
| Infection | Not Significant | Not Adjusted | 98 | Rio de Janeiro (Brazil) | 2016 - 2016 | Other | Children | De Melo Espindola (2021) |
| Infection | Not Significant | Not Adjusted | 250 | New York (United States) | 2016 - 2016 | Population | Pregnant Women | Cooper (2019) |
| **Severe disease (in general population)** | | | | | | | | |
| Age, Other, Sex | Significant | Not Adjusted | 10319 | Mexico | Jan 2012 - Mar 2020 | Population | Persons Under Investigation | Ananth (2020) |
| Prior immunity to arboviruses | Not Significant | Not Adjusted | 997 | Philippines | 2016 - 2016 | Population | Persons Under Investigation | Biggs (2021) |
| **Symptomatic infection** | | | | | | | | |
| Age, Comorbidity, Sex | Significant | Adjusted | 114 | San Juan, Puerto Rico (United States) | 16 Sep 2016 - 27 Oct 2016 | Contact | Other | Lozier (2018) |
| Age, Comorbidity, Sex | Significant | Not Adjusted | 114 | San Juan, Puerto Rico (United States) | 16 Sep 2016 - 27 Oct 2016 | Contact | Other | Lozier (2018) |
| Other | Not Significant | Adjusted | 114 | San Juan, Puerto Rico (United States) | 16 Sep 2016 - 27 Oct 2016 | Contact | Other | Lozier (2018) |
| Other | Not Significant | Not Adjusted | 114 | San Juan, Puerto Rico (United States) | 16 Sep 2016 - 27 Oct 2016 | Contact | Other | Lozier (2018) |
| **Zika congenital syndrome/other birth defects** | | | | | | | | |
| Infection during pregnancy | Significant | | 50 | Rio de Janeiro and Manaus (Brazil) | Sep 2015 - Jun 2016 | Population | Pregnant Women | Damasceno (2020) |
| Infection during pregnancy | Significant | Adjusted | 21 | Tahiti, Mo'orea, French Polynesia (France) | 01 Jun 2013 - 31 Aug 2014 | Hospital | Children | Subissi (2018) |
| Infection during pregnancy | Significant | Adjusted | 219 | Rio de Janeiro (Brazil) | Jan 2015 - Aug 2016 | Hospital | Pregnant Women | João (2018) |
| Infection during pregnancy | Significant | Adjusted | 511 | Ribeirão Preto (Brazil) | 2015 - 2016 | Population | Children | Coutinho (2021) |
| Infection during pregnancy | Significant | Adjusted | 291 | French Guiana (France) | 01 Jan 2015 - 15 Jul 2016 | Mixed | Children | Pomar (2020) |
| Infection during pregnancy | Significant | Adjusted | 129 | western French Guiana (France) | Jan 2016 - Sep 2016 | Hospital | Mixed Groups | Hcini (2021) |
| Infection during pregnancy | Significant | Adjusted | 83 | Jundiai Hospital Medical school (Brazil) | 01 Mar 2016 - 30 Jun 2019 | Hospital | Pregnant Women | Gazeta (2021) |
| Infection during pregnancy | Significant | Not Adjusted | 511 | Ribeirão Preto (Brazil) | 2015 - 2016 | Population | Children | Coutinho (2021) |

continued on next page

continued from previous page

| Risk factor name | Statistically significant | Adjusted | Sample size | Location | Dates | Population sample type | Population group | Reference |
|---|---|---|---|---|---|---|---|---|
| Infection during pregnancy | Significant | Not Adjusted | 557 | Jundiai, São Paulo (Brazil) | 01 Mar 2016 - 23 Aug 2017 | Hospital | Children | Clemente (2020) |
| Infection during pregnancy | Significant | Not Adjusted | 129 | western French Guiana (France) | Jan 2016 - Sep 2016 | Hospital | Mixed Groups | Hcini (2021) |
| Infection during pregnancy | Significant | Not Adjusted | 42 | Rio de Janeiro (Brazil) | Apr 2015 - Oct 2017 | Hospital | Children | Lima (2019) |
| Infection during pregnancy | Significant | Not Adjusted | 94 | Rio de Janeiro (Brazil) | 01 Mar 2016 - 30 Jun 2017 | Hospital | Pregnant Women | Pool (2019) |
| Infection during pregnancy | Significant | Unspecified | 108 | Maranhao state (Brazil) | Mar 2015 - Sep 2018 | Contact | Pregnant Women | Mendes (2020) |
| Infection during pregnancy, Other | Significant | Adjusted | 3603828 | Brazil | 01 Jan 2015 - 23 May 2017 | Population | Children | Brady (2019) |
| Infection during pregnancy, Other | Significant | Not Adjusted | 865 | Unspecified | Jan 2016 - Jul 2019 | Other | Pregnant Women | Vouga (2021) |
| Infection, Infection during pregnancy, Other | Significant | Adjusted | 174 | Ceará (Brazil) | Oct 2015 - Jun 2017 | Population | Children | Rocha (2019) |
| Infection, Infection during pregnancy, Other | Significant | Unspecified | 157 | Paraíba (Brazil) | Unspecified | Other | Persons Under Investigation | Krow-Lucal (2018) |
| Other | Significant | Adjusted | 143 | Rio de Janeiro (Brazil) | Dec 2015 | Other | Children | Azamor (2021) |
| Other | Significant | Not Adjusted | 555 | Martinique, Guadeloupe, French Guiana (France) | Mar 2016 - 27 Apr 2017 | Population | Pregnant Women | Hoen (2018) |
| Other | Significant | Not Adjusted | 120 | Mato Grosso do Sul (Brazil) | Nov 2017 - Oct 2018 | Population | Persons Under Investigation | De Sousa (2020) |
| Other | Significant | Not Adjusted | | Salvador, Bahia (Brazil) | 01 Jan 2015 - 31 Jan 2017 | Hospital | Mixed Groups | Aromolaran (2022) |
| Other, Sex | Significant | Adjusted | 11366686 | Brazil | 01 Jan 2015 - 31 Dec 2018 | Population | Children | Paixão (2022) |
| Age | Not Significant | Adjusted | 11366686 | Brazil | 01 Jan 2015 - 31 Dec 2018 | Population | Children | Paixão (2022) |
| Age, Other | Not Significant | Not Adjusted | 42 | Rio de Janeiro (Brazil) | Apr 2015 - Oct 2017 | Hospital | Children | Lima (2019) |
| Comorbidity, Other | Not Significant | Not Adjusted | 120 | Mato Grosso do Sul (Brazil) | Nov 2017 - Oct 2018 | Population | Persons Under Investigation | De Sousa (2020) |
| Infection during pregnancy | Not Significant | Adjusted | 174 | Thailand | 1997 - 2015 | Other | Pregnant Women | Ngo-Giang-Huong (2021) |
| Infection during pregnancy | Not Significant | Adjusted | 376 | Pernambuco (Recife) (Brazil) | Dec 2015 - Jun 2017 | Hospital | Pregnant Women | Ximenes (2021) |
| Infection during pregnancy | Not Significant | Not Adjusted | 1236 | Mombasa (Kenya) | Oct 2017 - Jul 2019 | Hospital | Children | Osoro (2022) |
| Infection during pregnancy | Not Significant | Not Adjusted | 102 | Colombia | Oct 2015 - Jan 2017 | Hospital | Pregnant Women | Gutierrez-Sanchez (2022) |
| Infection during pregnancy | Not Significant | Not Adjusted | 557 | Jundiai, São Paulo (Brazil) | 01 Mar 2016 - 23 Aug 2017 | Hospital | Children | Clemente (2020) |
| Infection during pregnancy | Not Significant | Not Adjusted | 70 | New York (United States) | Mar 2016 - Apr 2016 | Hospital | Pregnant Women | Merriam (2020) |

continued on next page

continued from previous page

| Risk factor name | Statistically significant | Adjusted | Sample size | Location | Dates | Population sample type | Population group | Reference |
|---|---|---|---|---|---|---|---|---|
| Infection during pregnancy, Other, Sex | Not Significant | Not Adjusted | 80 | Santo Domingo, Santiago (Dominican Republic) | 2016 - 2019 | Hospital | Children | Pimintel (2021) |
| Infection, Infection during pregnancy, Other | Not Significant | Unspecified | 157 | Paraíba (Brazil) | Unspecified | Other | Persons Under Investigation | Krow-Lucal (2018) |
| Infection, Infection during pregnancy, Other | Not Significant | Unspecified | 194 | León (Nicaragua) | Feb 2017 - Jul 2017 | Hospital | Children | Collins (2020) |
| Other | Not Significant | Adjusted | 143 | Rio de Janeiro (Brazil) | Dec 2015 | Other | Children | Azamor (2021) |
| Other | Not Significant | Not Adjusted | 143 | Rio de Janeiro (Brazil) | Dec 2015 | Other | Children | Azamor (2021) |
| **ZIKV detection** | | | | | | | | |
| Age, Occupation, Other | Not Significant | Not Adjusted | 315 | University Hospital of the Faculty of Medicine of Jundiaí, Jundiaí, São-Paulo (Brazil) | Mar 2016 - Aug 2017 | Hospital | Pregnant Women | Pires (2021) |
| Other | Not Significant | Adjusted | 315 | University Hospital of the Faculty of Medicine of Jundiaí, Jundiaí, São-Paulo (Brazil) | Mar 2016 - Aug 2017 | Hospital | Pregnant Women | Pires (2021) |

Table B.11: Overview of extracted risk factors. Levels of statistical significance may be different between studies.

##### B.7.6 Transmission

| Parameter value | Uncertainty | Disaggr-egation | Method for R estimation | Sample size | Location | Dates | Population sample type | Population group | Reference |
|---|---|---|---|---|---|---|---|---|---|
| **Attack Rate** | | | | | | | | | |
| 0 | 95% CrI: 0-0 | | | | Federal District (Mexico) | 28 Nov 2015 - 05 Nov 2018 | Population | General Population | Moore (2020) |
| 0.007 | 95% CI: 595-772 | Method | | 11247972 | Rio Grande (Brazil) | Oct 2015 - May 2016 | Population | Persons Under Investigation | Zhao (2019) |
| 0.016 | 95% CrI: 0.01-0.025 | | | | State of Paraná (Brazil) | Apr 2015 - 31 Dec 2018 | Population | General Population | Moore (2020) |
| 0.03 | 95% CrI: 0-0.1 | | | | Quibdó (Colombia) | 2015 - 2017 | Population | General Population | Charniga (2022) |
| 0.034 | 95% CI: 2040-2578 | Method | | 6610681 | Goiania City (Brazil) | Oct 2015 - May 2016 | Population | Persons Under Investigation | Zhao (2019) |
| 0.036 | 95% CI: 3610-4382 | Method | | 11163018 | Panara (Brazil) | Oct 2015 - May 2016 | Population | Persons Under Investigation | Zhao (2019) |
| 0.05 % | 95% CI: 0.01-0.29 | | | 1962 | Campinas (Brazil) | Mar 2015 - Mar 2015 | Population | Blood Donors | Benites (2019) |
| 0.054 | 95% CI: 1671-2364 | Method | | 3929911 | Espirito Santo (Brazil) | Oct 2015 - May 2016 | Population | Persons Under Investigation | Zhao (2019) |
| 0.084 | 95% CrI: 0.067-0.096 | | | | Peru | 01 Jan 2016 - 15 Sep 2018 | Population | General Population | Moore (2020) |
| 0.09 | 95% CrI: 0.02-0.19 | | | | Costa Rica | 01 Jan 2016 - 31 Dec 2017 | Population | General Population | Moore (2020) |
| 0.097 | 95% CI: 772-925 | Method | | 803513 | Acre (Brazil) | Oct 2015 - May 2016 | Population | Persons Under Investigation | Zhao (2019) |
| 0.106 | 95% CI: 9320-10721 | Method | | 9345173 | Pernambuco State (Brazil) | Oct 2015 - May 2016 | Population | Persons Under Investigation | Zhao (2019) |
| 0.12 | 95% CrI: 0.06-0.22 | | | | Panama | 22 Nov 2015 - 10 Nov 2018 | Population | General Population | Moore (2020) |
| 0.16 | 95% CrI: 0.07-0.29 | | | | Bolivia | 01 Jan 2016 - 30 Jun 2018 | Population | General Population | Moore (2020) |
| 0.17 % | 95% CI: 0.06-0.5 | | | 1775 | Campinas (Brazil) | Mar 2016 - Mar 2016 | Population | Blood Donors | Benites (2019) |
| 0.19 | 95% CrI: 0.15-0.23 | | | | Colombia | 09 Aug 2015 - 31 Dec 2017 | Population | General Population | Moore (2020) |
| 0.2 | 95% CrI: 0.15-0.25 | | | | Mexico | 28 Nov 2015 - 05 Nov 2018 | Population | General Population | Moore (2020) |
| 0.23 | 95% CrI: 0.07-0.49 | | | | Belize | 01 Jan 2016 - 31 Dec 2017 | Population | General Population | Moore (2020) |
| 0.23 | 95% CrI: 0.16-0.29 | | | | Guatemala | 29 Nov 2015 - 20 Oct 2018 | Population | General Population | Moore (2020) |
| 0.25 | 95% CrI: 0.19-0.31 | | | | Brazil | Apr 2015 - 31 Dec 2018 | Population | General Population | Moore (2020) |
| 0.25 | 95% CrI: 0.18-0.33 | | | | Dominican Republic | 01 Jan 2016 - 25 Mar 2018 | Population | General Population | Moore (2020) |
| 0.28 | 95% CrI: 0.16-0.4 | | | | El Salvador | 22 Nov 2015 - 16 Dec 2017 | Population | General Population | Moore (2020) |
| 0.32 | 95% CrI: 0.29-0.35 | | | | Puerto Rico (United States) | 01 Jan 2007 - 07 Jan 2017 | Population | General Population | Moore (2020) |
| 0.33 | 95% CrI: 0.21-0.46 | | | | Nicaragua | 27 Jan 2016 - 31 Dec 2017 | Population | General Population | Moore (2020) |
| 0.36 | 95% CrI: 0.22-0.49 | | | | Honduras | 13 Dec 2015 - 04 Jan 2018 | Population | General Population | Moore (2020) |
| 0.361 | 95% CrI: 0.214-0.514 | | | | Ecuador | 20 Dec 2015 - 23 Oct 2018 | Population | General Population | Moore (2020) |
| 0.365 | 95% CI: 50249-59773 | Method | | 15203934 | Bahia (Brazil) | Oct 2015 - May 2016 | Population | Persons Under Investigation | Zhao (2019) |

continued on next page

continued from previous page

| Parameter value | Uncertainty | Disaggr-egation | Method for R estimation | Sample size | Location | Dates | Population sample type | Population group | Reference |
|---|---|---|---|---|---|---|---|---|---|
| 0.38 | 95% CrI: 0.17-0.92 | Region | | | Arauca, Armenia, Barranquilla, Bucaramanga, Cali, Cartagena, Cúcuta, Florencia, Ibagué, Inírida, Leticia, Medellín, Mitú, Mocoa, Montería, Neiva, Pereira, Popayán, Puerto Carreño, Quibdó, Riohacha, San Andrés, San José del Guaviare, Santa Marta, Sincelejo, Valledupar, Villavicencio, Yopal (Colombia) | 2015 - 2017 | Population | General Population | Charniga (2022) |
| 0.514 | 95% CrI: 0.388-0.631 | | | | Bahia (Brazil) | Apr 2015 - 31 Dec 2018 | Population | General Population | Moore (2020) |
| 0.606 | 95% CI: 12901-19791 | Method | | 3265486 | Mato Grosso (Brazil) | Oct 2015 - May 2016 | Population | Persons Under Investigation | Zhao (2019) |
| 0.766 | 95% CrI: 0.569-0.942 | | | | State of Sergipe (Brazil) | Apr 2015 - 31 Dec 2018 | Population | General Population | Moore (2020) |
| 0.793 | 95% CrI: 0.524-0.963 | | | | State of Yucatán (Mexico) | 28 Nov 2015 - 05 Nov 2018 | Population | General Population | Moore (2020) |
| 0.8 | 95% CrI: 0.56-0.99 | | | | San Andrés (Colombia) | 2015 - 2017 | Population | General Population | Charniga (2022) |
| 1.213 % | | Age, Sex | | 928 | San Andrés (Colombia) | 06 Sep 2015 - 30 Jan 2016 | Hospital | Persons Under Investigation | Rojas (2016) |
| 1.843 % | | Age, Sex | | 1936 | Girardot (Colombia) | 19 Oct 2015 - 22 Jan 2016 | Hospital | Persons Under Investigation | Rojas (2016) |
| 2.7 per 10k | | Age | | | Dominican Republic | Jan 2016 - Dec 2016 | Population | General Population | Bowman (2018) |
| 5.2 per 10k | | | | 5235 | Dominican Republic | Jan 2016 - Dec 2016 | Population | General Population | Bowman (2018) |
| 5.5 % | 95% CI: 3-10.4 | | | 997 | Philippines | 2016 - 2016 | Population | General Population | Biggs (2021) |
| 7.6 per 10k | | Age | | | Dominican Republic | Jan 2016 - Dec 2016 | Population | General Population | Bowman (2018) |
| 12 % | | | | 34 | China | Feb 2016 - Feb 2016 | Travel | Other | Sun (2017) |
| 14.4 % | 95% CI: 5.6-27.4 | | | | Wynwood Neighborhood, Miami-Dade County, Florida (United States) | 01 Jun 2016 - 05 Aug 2016 | Population | General Population | Marini (2017) |
| 14.6 per 10k | | Age, Region, Sex | | 185 | Yap Island (Federated States of Micronesia) | 01 Apr 2007 - 31 Jul 2007 | Hospital | Persons Under Investigation | Duffy (2009) |
| 20 per 100k | | Age, Method, Other, Region, Sex | | 48747574 | Colombia | 10 Oct 2015 - 24 Jun 2017 | Population | General Population | Méndez (2017) |
| 20.8 % | 95% CI: 1.1-50.3 | | | | Colombia | Aug 2015 - May 2016 | Population | General Population | He (2017) |
| 32.4 % | 95% CI: 2.5-94.2 | | | | Bahia (Brazil) | Feb 2015 - Feb 2016 | Population | General Population | He (2017) |
| 50 % | 95% CrI: 43-58 | | | | Martinique (France) | 2015 - 2016 | Mixed | Mixed Groups | Cousien (2019) |
| % | | | | 1110 | Managua (Nicaragua) | Jan 2016 - Feb 2017 | Hospital | Persons Under Investigation | Burger-Calderon (2020) |
| 51.1 | Other: 42.1-61.1 | Method | | 7580 | Santiago, Fogo, Maio, Boavista (Cabo Verde) | 05 Oct 2015 - 29 May 2016 | Population | Children | Lourenco (2018) |

continued on next page

continued from previous page

| Parameter value | Uncertainty | Disaggr-egation | Method for R estimation | Sample size | Location | Dates | Population sample type | Population group | Reference |
|---|---|---|---|---|---|---|---|---|---|
| 62.5 % | 95% CI: 59.2-82.5 | | | | "Other four archipelagos", French Polynesia (France) | Aug 2013 - May 2014 | Population | General Population | He (2017) |
| 65 % | 95% CI: 57-72.9 | | | | Feira de Santana (Brazil) | 01 Feb 2015 - 30 Apr 2017 | Population | General Population | Lourenco (2017) |
| 70.1 % | 95% CI: 66.3-92.5 | | | | Sous-le-vent Islands, French Polynesia (France) | Aug 2013 - May 2014 | Population | General Population | He (2017) |
| 71.3 % | 95% CI: 67.4-94.1 | | | | Tahiti, French Polynesia (France) | Aug 2013 - May 2014 | Population | General Population | He (2017) |
| 73 % | 95% CI: 70-76 | Region | | 1453 | Salvador, Bahia (Brazil) | 2015 - 2015 | Population | General Population | Rodriguez-Barraquer (2019) |
| 78 % | 95% CI: 63.5-86.3 | Region | | | French Polynesia (France) | Aug 2013 - May 2014 | Population | General Population | He (2017) |
| 94 % | | | Compartmental Model | | French Polynesia (France) | 11 Oct 2013 - 28 Mar 2014 | Population | Persons Under Investigation | Kucharski (2016) |
| 552 per 100k | | Age | | | Cucuta (Colombia) | 29 Jun 2015 - 30 Jul 2016 | Hospital | Persons Under Investigation | Anaya (2017) |
| 1306 per 100k | | Age | | | Cucuta (Colombia) | 29 Jun 2015 - 30 Jul 2016 | Hospital | Persons Under Investigation | Anaya (2017) |
| 1 - 43 % | | Region | | | Multi-country: Central and South America (n = 8) | Jan 2014 - 01 Feb 2016 | Hospital | Persons Under Investigation | Zhang (2017) |
| 5 - 49 % | | Region | | | Multi-country: Central and South America (n = 8) | 01 Feb 2016 - 28 Feb 2017 | Hospital | Persons Under Investigation | Zhang (2017) |
| 0.897 - 21.16 per 10k | 95% CI: 0.897-21.548 | Region | | | All municipalities (Dominican Republic) | 2014 - 2016 | Population | General Population | Kingston (2022) |
| 0.219 - 0.285 | | Method | | | Northeastern (Brazil) | Jan 2015 - Nov 2016 | Population | Persons Under Investigation | He (2020) |
| 46 - 53 % | | Method | | | Fiji | May 2013 - Nov 2017 | Population | Persons Under Investigation | Henderson (2021) |
| 0 - 0.9 % | | Method, Other | | 481 | Netherlands | Mar 2014 - Oct 2017 | Travel | General Population | Overbosch (2023) |
| **Basic Reproduction Number** | | | | | | | | | |
| 0.237 | | Method, Other | Compartmental Model | | Amazonas (Brazil) | 06 Feb 2021 - 30 Apr 2021 | Population | Persons Under Investigation | Omame (2023a) |
| 0.494 | | | Unspecified | | Colombia | 2015 - 2015 | Unspecified | Unspecified | Zafar (2024) |
| 0.548 | | | Compartmental Model | | Colombia | 2016 - 2016 | Unspecified | Unspecified | Alzahrani (2021) |
| 0.737 | | | Compartmental Model | | Suriname | Unspecified | Population | General Population | Dénes (2019) |
| 0.924 | | | Compartmental Model | | Costa Rica | 2016 - 2018 | Population | General Population | Dénes (2019) |
| 0.98 | 95% CI: 0.93-1.02 | | Compartmental Model | 1841 | Envigado (Colombia) | 01 Jan 2016 - 11 Apr 2016 | Population | Persons Under Investigation | Ospina (2017) |
| 0.98 | | | Compartmental Model | 1841 | Itagüí (Colombia) | 01 Jan 2016 - 11 Apr 2016 | Population | Persons Under Investigation | Ospina (2017) |
| 0.98 | 95% CI: 0.88-1.08 | | Compartmental Model | 1841 | Mutatá (Colombia) | 01 Jan 2016 - 11 Apr 2016 | Population | Persons Under Investigation | Ospina (2017) |
| 0.98 | 95% CI: 0.92-1.03 | | Compartmental Model | 1841 | Rionegro (Colombia) | 01 Jan 2016 - 11 Apr 2016 | Population | Persons Under Investigation | Ospina (2017) |
| 0.98 | | | Compartmental Model | 1841 | Zaragoza (Colombia) | 01 Jan 2016 - 11 Apr 2016 | Population | Persons Under Investigation | Ospina (2017) |
| 1.009 | 95% CI: 0.99-1.02 | | Compartmental Model | 1841 | Caucasia (Colombia) | 01 Jan 2016 - 11 Apr 2016 | Population | Persons Under Investigation | Ospina (2017) |
| 1.03 | 95% CI: 0.97-1.09 | | Compartmental Model | 1841 | Puerto Triunfo (Colombia) | 01 Jan 2016 - 11 Apr 2016 | Population | Persons Under Investigation | Ospina (2017) |

continued on next page

continued from previous page

| Parameter value | Uncertainty | Disaggr-egation | Method for R estimation | Sample size | Location | Dates | Population sample type | Population group | Reference |
|---|---|---|---|---|---|---|---|---|---|
| 1.05 | 95% CI: 1.03-1.08 | | Other | | Guatemala | Unspecified | | | Hsieh (2017) |
| 1.07 | 95% CI: 0.8-1.26 | | Compartmental Model | 1841 | Sopetrán (Colombia) | 01 Jan 2016 - 11 Apr 2016 | Population | Persons Under Investigation | Ospina (2017) |
| 1.08 | 95% CI: 1.02-1.15 | | Compartmental Model | 1841 | Puerto Berrío (Colombia) | 01 Jan 2016 - 11 Apr 2016 | Population | Persons Under Investigation | Ospina (2017) |
| 1.11 | 95% CI: 1.09-1.13 | | Compartmental Model | 1841 | Apartadó (Colombia) | 01 Jan 2016 - 11 Apr 2016 | Population | Persons Under Investigation | Ospina (2017) |
| 1.117 | | | Other | | Tuamotu-Gambier, French Polynesia (France) | 2013 - 2014 | Population | Persons Under Investigation | Prasad (2023) |
| 1.12 | | Region | Compartmental Model | 1841 | Antioquia (Colombia) | 01 Jan 2016 - 11 Apr 2016 | Population | Persons Under Investigation | Ospina (2017) |
| 1.12 | 95% CI: 0.99-1.24 | | Compartmental Model | 1841 | Necoclí (Colombia) | 01 Jan 2016 - 11 Apr 2016 | Population | Persons Under Investigation | Ospina (2017) |
| 1.12 | 95% CrI: 0.8-1.5 | Method, Time | Compartmental Model | | Fiji | 2013 - 2018 | Population | Persons Under Investigation | Henderson (2021) |
| 1.13 | 95% CI: 0.84-1.42 | | Compartmental Model | 1841 | Remedios (Colombia) | 01 Jan 2016 - 11 Apr 2016 | Population | Persons Under Investigation | Ospina (2017) |
| 1.142 | | | Other | | Marquesas Islands, French Polynesia (France) | 2013 - 2014 | Population | Persons Under Investigation | Prasad (2023) |
| 1.15 | 95% CI: 1.12-1.17 | | Compartmental Model | 1841 | Turbo (Colombia) | 01 Jan 2016 - 11 Apr 2016 | Population | Persons Under Investigation | Ospina (2017) |
| 1.151 | | | Other | | Mo'orea, French Polynesia (France) | 2013 - 2014 | Population | Persons Under Investigation | Prasad (2023) |
| 1.172 | | | Other | | Colombia | 2016 - 2016 | Population | Persons Under Investigation | Prasad (2023) |
| 1.18 | 95% CI: 1.06-1.3 | | Compartmental Model | 1841 | Bello (Colombia) | 01 Jan 2016 - 11 Apr 2016 | Population | Persons Under Investigation | Ospina (2017) |
| 1.18 | 95% CI: 1.04-1.41 | | Other | 751 | Porto Velho (Brazil) | Jul 2016 - Jun 2019 | Other | Other | Botosso (2023) |
| 1.185 | | | Other | | Sous-le-vent Islands, French Polynesia (France) | 2013 - 2014 | Population | Persons Under Investigation | Prasad (2023) |
| 1.225 | | | Other | | Australes, French Polynesia (France) | 2013 - 2014 | Population | Persons Under Investigation | Prasad (2023) |
| 1.23 | 95% CI: 1.18-1.28 | | Compartmental Model | 1841 | Carepa (Colombia) | 01 Jan 2016 - 11 Apr 2016 | Population | Persons Under Investigation | Ospina (2017) |
| 1.28 | 95% CI: 1.08-1.69 | | Other | 817 | Belo Horizonte (Brazil) | Jul 2016 - Jun 2016 | Other | Other | Botosso (2023) |
| 1.31 | 95% CI: 1.1-1.52 | | Compartmental Model | 1841 | San Pedro de Urabá (Colombia) | 01 Jan 2016 - 11 Apr 2016 | Population | Persons Under Investigation | Ospina (2017) |
| 1.41 | 95% CI: 1.15-1.74 | | Other | 928 | San Andrés (Colombia) | 06 Sep 2015 - 30 Jan 2016 | Hospital | Persons Under Investigation | Rojas (2016) |
| 1.41 | 95% CI: 1.12-2.08 | | Other | 494 | Salvador, Bahia (Brazil) | Jul 2016 - Jun 2019 | Other | Other | Botosso (2023) |
| 1.43 | 95% CI: 1.21-1.85 | | Other | 345 | Fortaleza (Brazil) | Jul 2016 - Jun 2019 | Other | Other | Botosso (2023) |
| 1.53 | 95% CI: 1.71-1.89 | | Compartmental Model | 1841 | Chigorodó (Colombia) | 01 Jan 2016 - 11 Apr 2016 | Population | Persons Under Investigation | Ospina (2017) |
| 1.54 | 95% CI: 1.35-1.92 | Region | Other | 5108 | Belo Horizonte, Salvador, Laranjeiras, Fortaleza, Recife, Cuiaba and Campo Grande, Porto Velho (Brazil) | Jul 2016 - Jun 2019 | Other | Other | Botosso (2023) |
| 1.58 | 95% CrI: 1.56-1.59 | | Next Generation Matrix | | Managua (Nicaragua) | 2016 - 2016 | Population | General Population | Counotte (2019) |
| 1.59 | 95% CI: 1.28-1.9 | Time | Other | | Guatemala | 2015 - 2016 | | | Hsieh (2017) |
| 1.6 | 95% CrI: 1.5-1.7 | | Next Generation Matrix | | Tahiti, French Polynesia (France) | Oct 2013 - Apr 2014 | Population | Persons Under Investigation | Champagne (2016) |

continued on next page

continued from previous page

| Parameter value | Uncertainty | Disaggr-egation | Method for R estimation | Sample size | Location | Dates | Population sample type | Population group | Reference |
|---|---|---|---|---|---|---|---|---|---|
| 1.6 | 95% CrI: 1.5-1.7 | | Next Generation Matrix | | New Caledonia (France) | 12 Nov 2013 - Aug 2014 | Population | Persons Under Investigation | Champagne (2016) |
| 1.67 | 95% CI: 1.43-3.57 | | Other | 1017 | Cuiaba and Campo Grande (Brazil) | Jul 2016 - Jun 2019 | Other | Other | Botosso (2023) |
| 1.68 | 95% CI: 1.32-2.04 | | Other | | Suriname | 2015 - 2016 | | | Hsieh (2017) |
| 1.72 | 95% CI: 1.69-1.75 | Age | Growth Rate | | Dominican Republic | Jan 2016 - Dec 2016 | Population | General Population | Bowman (2018) |
| 1.727 | | | Next Generation Matrix | | Colombia | 2015 - Dec 2016 | Population | General Population | Valega-Mackenzie (2023) |
| 1.75 | 95% CI: 1.34-2.16 | Time | Other | | Colombia | 2015 - 2016 | | | Hsieh (2017) |
| 1.79 | 95% CI: 1.29-2.3 | | Other | | Colombia | Unspecified | | | Hsieh (2017) |
| 1.79 | Other: 1.68-1.91 | | Other | | Yap Island, French Polynesia (France) | 2007 - 2007 | Population | General Population | Gao (2016) |
| 1.8 | 95% CI: 1.7-1.9 | Region | Other | 1453 | Salvador, Bahia (Brazil) | 2015 - 2015 | Population | General Population | Rodriguez-Barraquer (2019) |
| 1.8 | | | Other | 573 | Dominican Republic | 30 Jan 2016 - 27 Feb 2016 | Population | Persons Under Investigation | Rocklov (2016) |
| 1.8 | 95% CrI: 1.6-2 | | Next Generation Matrix | | Mo'orea, French Polynesia (France) | Oct 2013 - Apr 2014 | Population | Persons Under Investigation | Champagne (2016) |
| 1.8 | 95% CI: 1.78-1.82 | | Growth Rate | 5235 | Dominican Republic | Jan 2016 - Dec 2016 | Population | General Population | Bowman (2018) |
| 1.81 | 95% CrI: 1.74-1.87 | Other, Region | Compartmental Model | | 90 major cities across LAC (Multi-country: Americas (n = 32)) | 2015 - 2017 | Population | General Population | O'Reilly (2018) |
| 1.816 | | | Compartmental Model | | Brazil | 25 Mar 2016 - 14 Apr 2018 | Population | Persons Under Investigation | Zhao (2020) |
| 1.82 | 95% CI: 1.19-2.68 | | Growth Rate | | Rio de Janeiro (Brazil) | 2012 | | | Villela (2017) |
| 1.84 | 95% CI: 1.82-1.87 | Age | Growth Rate | | Dominican Republic | Jan 2016 - Dec 2016 | Population | General Population | Bowman (2018) |
| 1.85 | 95% CI: 1.5-2.2 | | Growth Rate | 7580 | Santiago, Fogo, Maio, Boavista (Cabo Verde) | 05 Oct 2015 - 29 May 2016 | Population | Persons Under Investigation | Lourenco (2018) |
| 1.866 | | | Other | | Costa Rica | 2016 - 2016 | Population | Persons Under Investigation | Prasad (2023) |
| 1.88 | 95% CrI: 1.59-2.22 | Region | Branching Process | 1e+05 | Polynesia and West Indies | Oct 2013 - Oct 2016 | Population | Persons Under Investigation | Riou (2017) |
| 1.89 | 95% CI: 1.21-2.13 | Method | Compartmental Model | 18364 | Colombia | 2015 - 2016 | Population | Pregnant Women | Sasmal (2018) |
| 1.89 | 95% CI: 1.33-3.23 | | Other | 1110 | Recife (Brazil) | Jul 2016 - Jun 2019 | Other | Other | Botosso (2023) |
| 1.9 | | | Other | 49 | Panama | 01 Jan 2016 - 13 Mar 2016 | Population | Persons Under Investigation | Rocklov (2016) |
| 1.947 | | | Other | | Tahiti, French Polynesia (France) | 2013 - 2014 | Population | Persons Under Investigation | Prasad (2023) |
| 2 | 95% CrI: 1.8-2.2 | | Next Generation Matrix | | New Caledonia (France) | 12 Nov 2013 - Aug 2014 | Population | Persons Under Investigation | Champagne (2016) |
| 2.027 | 95% CI: 1.907-2.14 | | | | Australes, French Polynesia (France) | Oct 2013 - Mar 2014 | Population | General Population | Rahman (2019) |
| 2.03 | 95% CrI: 1.63-2.36 | | Other | | Australes, French Polynesia (France) | 01 Dec 2015 - 30 Mar 2017 | Community | Persons Under Investigation | Riou (2018) |
| 2.05 | 95% CrI: 1.71-2.37 | | Other | | Tuamotu, French Polynesia (France) | 20 Oct 2015 - 02 Mar 2017 | Community | Persons Under Investigation | Riou (2018) |
| 2.055 | 95% CI: 0.523-6.3 | | Compartmental Model | | Multi-country: Brazil, Colombia, El Salvador | Apr 2015 - 27 Feb 2016 | Population | General Population | Gao (2016) |

continued on next page

continued from previous page

| Parameter value | Uncertainty | Disaggr-egation | Method for R estimation | Sample size | Location | Dates | Population sample type | Population group | Reference |
|---|---|---|---|---|---|---|---|---|---|
| 2.069 | 95% CI: 1.261-4.316 | | | | Marquesas Islands, French Polynesia (France) | Oct 2013 - Mar 2014 | Population | General Population | Rahman (2019) |
| 2.1 | | | Other | 717 | Guadeloupe (France) | 29 Jan 2016 - 10 Mar 2016 | Population | Persons Under Investigation | Rocklov (2016) |
| 2.1 | | | Other | 129 | Mexico | 01 Jan 2016 - 13 Mar 2016 | Population | Persons Under Investigation | Rocklov (2016) |
| 2.1 | 95% CrI: 1.78-2.47 | | Other | | Mo'orea, French Polynesia (France) | 27 Oct 2015 - 09 Mar 2017 | Community | Persons Under Investigation | Riou (2018) |
| 2.1 | 95% CrI: 1.8-2.43 | | Other | | Sous-le-vent Islands, French Polynesia (France) | 27 Oct 2015 - 23 Mar 2017 | Community | Persons Under Investigation | Riou (2018) |
| 2.1 | 95% CrI: 1.86-2.35 | | Other | | Tahiti, French Polynesia (France) | 13 Oct 2015 - 16 Mar 2017 | Community | Persons Under Investigation | Riou (2018) |
| 2.12 | 95% CrI: 1.82-2.47 | | Other | | Marquesas Islands, French Polynesia (France) | 27 Oct 2015 - 23 Mar 2017 | Community | Persons Under Investigation | Riou (2018) |
| 2.13 | 95% CI: 1.61-2.64 | | Next Generation Matrix | | Brazil | Jan 2015 - Sep 2016 | Population | General Population | Wang (2024a) |
| 2.135 | 95% CI: 2.07-2.2 | | Next Generation Matrix | | Unspecified | 01 Oct 2013 - 30 Apr 2014 | | | Gwalani (2018) |
| 2.2 | 95% CI: 1.54-2.86 | | Compartmental Model | 1841 | Nechí (Colombia) | 01 Jan 2016 - 11 Apr 2016 | Population | Persons Under Investigation | Ospina (2017) |
| 2.2 | 95% CrI: 1.9-2.6 | | Next Generation Matrix | | Yap Island (Federated States of Micronesia) | 01 Apr 2007 - 29 Jul 2007 | Population | Persons Under Investigation | Champagne (2016) |
| 2.205 | 95% CrI: 1.466-3.166 | | Next Generation Matrix | | El Salvador | 2015 - 22 Jun 2016 | Population | Persons Under Investigation | Shutt (2017) |
| 2.227 | | | Next Generation Matrix | | Puerto Rico (United States) | Jan 2016 - Dec 2017 | Population | General Population | Valega-Mackenzie (2023) |
| 2.23 | | Other | Next Generation Matrix | | Colombia | Unspecified | Unspecified | Unspecified | Li (2019) |
| 2.28 | | | Compartmental Model | 3681000 | Puerto Rico (United States) | Dec 2015 - Dec 2016 | Unspecified | General Population | Biswas (2024) |
| 2.33 | 95% CI: 1.97-2.97 | Other, Region | Growth Rate | | Rio de Janeiro (Multi-country: Brazil and Colombia) | 18 Oct 2015 | | | Villela (2017) |
| 2.33 | 95% CI: 1.54-3.85 | | Other | 574 | Laranjeiras (Brazil) | Jul 2016 - Jun 2019 | Other | Other | Botosso (2023) |
| 2.337 | 95% CI: 2.152-2.508 | | | | Tuamotu-Gambier, French Polynesia (France) | Oct 2013 - Mar 2014 | Population | General Population | Rahman (2019) |
| 2.338 | 95% CI: 2.179-2.488 | | | | Tahiti, French Polynesia (France) | Oct 2013 - Mar 2014 | Population | General Population | Rahman (2019) |
| 2.37 | 95% CI: 2.35-2.39 | | Next Generation Matrix | | Costa Rica | 12 Apr 2016 - 24 Aug 2016 | Population | General Population | Luo (2021) |
| 2.4 | 95% CrI: 2-3.2 | | Next Generation Matrix | | Tahiti, French Polynesia (France) | Oct 2013 - Apr 2014 | Population | Persons Under Investigation | Champagne (2016) |
| 2.422 | 95% CrI: 1.653-3.47 | | Next Generation Matrix | | Suriname | 2015 - 22 Jun 2016 | Population | Persons Under Investigation | Shutt (2017) |
| 2.45 | 95% CI: 1.57-3.65 | | Growth Rate | | Rio de Janeiro (Brazil) | 2002 | | | Villela (2017) |
| 2.498 | 95% CI: 2.042-3.148 | | | | Sous-le-vent Islands, French Polynesia (France) | Oct 2013 - Mar 2014 | Population | General Population | Rahman (2019) |
| 2.502 | | | Next Generation Matrix | | Brazil | 06 Feb 2016 - 02 Jun 2016 | Population | Persons Under Investigation | Wang (2017) |
| 2.53 | 95% CI: 1.59-3.45 | | Compartmental Model | | Rio de Janeiro (Brazil) | 2015 - 2016 | Population | General Population | Bastos (2018) |
| 2.6 | 95% CrI: 1.7-5.3 | | Compartmental Model | | Marquesas Islands, French Polynesia (France) | 11 Oct 2013 - 28 Mar 2014 | Population | Persons Under Investigation | Kucharski (2016) |

continued on next page

continued from previous page

| Parameter value | Uncertainty | Disaggr- egation | Method for R estimation | Sample size | Location | Dates | Population sample type | Population group | Reference |
|---|---|---|---|---|---|---|---|---|---|
| 2.6 | 95% CrI: 2.2-3.3 | | Next Generation Matrix | | Mo'orea, French Polynesia (France) | Oct 2013 - Apr 2014 | Population | Persons Under Investigation | Champagne (2016) |
| 2.69 | 95% CrI: 2.31-3.11 | | Growth Rate | | Manaus (Brazil) | Nov 2015 | Population | General Population | Giovanetti (2020) |
| 2.7 | Range: 1-4.3 | | Unspecified | 1946 | Feira de Santana (Brazil) | 01 Feb 2015 - 30 Apr 2017 | Population | General Population | Lourenco (2017) |
| 2.75 | 95% CI: 2.53-2.98 | | | | French Polynesia (France) | 2013 | Unspecified | Unspecified | Sun (2018) |
| 2.75 | 95% CI: 2.53-2.98 | | Compartmental Model | | French Polynesia (France) | 2013 - 2013 | Population | Persons Under Investigation | Perrotta (2022) |
| 3 | 95% CrI: 2.2-6.1 | | Compartmental Model | | Tuamotu-Gambier, French Polynesia (France) | 11 Oct 2013 - 28 Mar 2014 | Population | Persons Under Investigation | Kucharski (2016) |
| 3.1 | 95% CrI: 2.2-4.6 | | Compartmental Model | | Australes, French Polynesia (France) | 11 Oct 2013 - 28 Mar 2014 | Population | Persons Under Investigation | Kucharski (2016) |
| 3.188 | 95% CI: 2.61-3.676 | | | | Mo'orea, French Polynesia (France) | Oct 2013 - Mar 2014 | Population | General Population | Rahman (2019) |
| 3.199 | 95% CI: 3.073-3.32 | | | | Yap Island (Federated States of Micronesia) | Oct 2013 - Mar 2014 | Population | General Population | Rahman (2019) |
| 3.2 | | | Other | 1593 | Colombia | 01 Jan 2016 - 13 Mar 2016 | Population | Persons Under Investigation | Rocklov (2016) |
| 3.2 | 95% CrI: 2.4-4.1 | | Next Generation Matrix | | Yap Island (Federated States of Micronesia) | 01 Apr 2007 - 29 Jul 2007 | Population | Persons Under Investigation | Champagne (2016) |
| 3.26 | Range: 2-5 | | Other | | Colombia | 31 May 2015 - 16 Apr 2016 | Population | Persons Under Investigation | Majumder (2016) |
| 3.5 | 95% CrI: 2.6-5.3 | | Compartmental Model | | Tahiti, French Polynesia (France) | 11 Oct 2013 - 28 Mar 2014 | Population | Persons Under Investigation | Kucharski (2016) |
| 3.62 | 95% CI: 3.48-3.77 | Time | Other | | Singapore | 31 Jul 2016 - 01 Sep 2016 | | Persons Under Investigation | Singapore Zika Study Group (2017) |
| 3.7 | | | Other | 198 | Puerto Rico (United States) | 26 Nov 2015 - 24 Feb 2016 | Population | Persons Under Investigation | Rocklov (2016) |
| 3.8 | 95% CI: 2.4-5.6 | | Other | 359 | Barranquilla (Colombia) | 01 Oct 2015 - Nov 2015 | Hospital | Persons Under Investigation | Towers (2016) |
| 4.1 | 95% CrI: 3.1-5.7 | | Compartmental Model | | Sous-le-vent Islands, French Polynesia (France) | 11 Oct 2013 - 28 Mar 2014 | Population | Persons Under Investigation | Kucharski (2016) |
| 4.61 | 95% CI: 4.11-5.16 | | Other | 1936 | Girardot (Colombia) | 19 Oct 2015 - 22 Jan 2016 | Hospital | Persons Under Investigation | Rojas (2016) |
| 4.8 | | | Other | 5618 | El Salvador | 01 Jan 2016 - 13 Mar 2016 | Population | Persons Under Investigation | Rocklov (2016) |
| 4.8 | 95% CrI: 3.2-8.4 | | Compartmental Model | | Mo'orea, French Polynesia (France) | 11 Oct 2013 - 28 Mar 2014 | Population | Persons Under Investigation | Kucharski (2016) |
| 5 | | | Other | 386 | Guatemala | 16 Nov 2015 - 15 Feb 2016 | Population | Persons Under Investigation | Rocklov (2016) |
| 5.36 | Range: 3-10 | | Other | | Colombia | 22 Aug 2015 - 16 Apr 2016 | Population | Persons Under Investigation | Majumder (2016) |
| 5.7 | 95% CI: 0-11.75 | | Other | | Saint Martin (Multi-country: France and Netherlands) | 2015 - 2016 | | | Hsieh (2017) |
| 6 | 95% CI: 0.06-11.95 | Region | Other | | French Polynesia (France) | 2013 - 2014 | | | Hsieh (2017) |
| 6.89 | 95% CI: 0-16.24 | | Other | | Puerto Rico (United States) | 2015 - 2016 | | | Hsieh (2017) |
| 6.93 | 95% CI: 5.82-8.05 | | Compartmental Model | | Singapore (Singapore) | 07 Aug 2016 - 31 Aug 2016 | Population | General Population | Zhu (2022) |
| 7 | | | Other | 96 | Ecuador | 01 Jan 2016 - 13 Mar 2016 | Population | Persons Under Investigation | Rocklov (2016) |
| 7.2 | | | Other | 102 | Martinique (France) | 21 Dec 2015 - 21 Jan 2016 | Population | Persons Under Investigation | Rocklov (2016) |

continued on next page

continued from previous page

| Parameter value | Uncertainty | Disaggr-egation | Method for R estimation | Sample size | Location | Dates | Population sample type | Population group | Reference |
|---|---|---|---|---|---|---|---|---|---|
| 22.2 | | | Compartmental Model | 1841 | Medellín (Colombia) | 01 Jan 2016 - 11 Apr 2016 | Population | Persons Under Investigation | Ospina (2017) |
| 56.38 | | | Compartmental Model | 1841 | Cáceres (Colombia) | 01 Jan 2016 - 11 Apr 2016 | Population | Persons Under Investigation | Ospina (2017) |
| 1.24 - 1.47 | | Method | Compartmental Model | | Florida (United States) | 19 Jul 2016 - 29 Sep 2016 | Population | Persons Under Investigation | Tuncer (2018) |
| 1.54 - 1.9 | | Method, Time | Other | | Guadeloupe (France) | 24 Jan 2015 - 29 Jan 2017 | Community | Persons Under Investigation | Riou (2018) |
| 1.3 - 1.62 | | Method, Time | Other | | Martinique (France) | 20 Dec 2015 - 05 Feb 2017 | Community | Persons Under Investigation | Riou (2018) |
| 1.34 - 1.57 | | Method, Time | Other | | Saint Martin (France) | 24 Jan 2015 - 19 Feb 2017 | Community | Persons Under Investigation | Riou (2018) |
| 0.4 - 2.73 | | | Compartmental Model | | Wynwood Neighborhood, Miami-Dade County, Florida (United States) | 26 Jan 2016 - 05 Aug 2016 | Population | General Population | Marini (2017) |
| 2.8 - 12.5 | | Other | Growth Rate | 108 | Yap Island (Federated States of Micronesia) | 2007 - 2007 | Unspecified | Unspecified | Nishiura (2016) |
| 1.5 - 3.1 | | Other | Growth Rate | 8581 | French Polynesia (France) | 2013 - 2014 | Unspecified | Unspecified | Nishiura (2016) |
| 1.29 - 1.71 | | Method | Branching Process | 44825 | Centre-West Brazil (Brazil) | 01 Jan 2015 - 19 Nov 2016 | Population | Persons Under Investigation | Faria (2017) |
| 1.58 - 2.48 | | Method | Branching Process | 22373 | North Brazil (Brazil) | 01 Jan 2015 - 19 Nov 2016 | Population | Persons Under Investigation | Faria (2017) |
| 1.75 - 3.12 | | Method, Time | Branching Process | 122779 | Northeast Brazil (Brazil) | 01 Jan 2015 - 19 Nov 2016 | Population | Persons Under Investigation | Faria (2017) |
| 1.98 - 3.85 | | Method | Branching Process | 112689 | South-East Brazil (Brazil) | 01 Jan 2015 - 19 Nov 2016 | Population | Persons Under Investigation | Faria (2017) |
| 1.61 - 2.57 | | Method | Branching Process | 4944 | South Brazil (Brazil) | 01 Jan 2015 - 19 Nov 2016 | Population | Persons Under Investigation | Faria (2017) |
| 2.68 - 4.57 | | Method | Growth Rate | 6117 | Cucuta (Colombia) | 29 Jun 2015 - 30 Jul 2016 | Hospital | Persons Under Investigation | Anaya (2017) |
| **Basic Reproduction Number - Sexual** | | | | | | | | | |
| 0.005 | | | Next Generation Matrix | | Puerto Rico (United States) | Jan 2016 - Dec 2017 | Population | General Population | Valega-Mackenzie (2023) |
| 0.054 | | | Next Generation Matrix | | Brazil | 06 Feb 2016 - 02 Jun 2016 | Population | Persons Under Investigation | Wang (2017) |
| 0.136 | 95% CI: 0.009-0.521 | | Compartmental Model | | Multi-country: Brazil, Colombia, El Salvador | Apr 2015 - 27 Feb 2016 | Population | General Population | Gao (2016) |
| 0.19 | 95% CI: 0.05-0.48 | Region | Ross-Macdonald Formula | | Texas (United States) | Jan 2016 - Jan 2017 | Population | General Population | Fox (2019) |
| 0.2 | | | Compartmental Model | | Brazil | 25 Mar 2016 - 14 Apr 2018 | Population | Persons Under Investigation | Zhao (2020) |
| 0.42 | 95% CI: 0.29-0.64 | Method | Compartmental Model | 18364 | Colombia | 2015 - 2016 | Population | Pregnant Women | Sasmal (2018) |
| 0.624 | | | Next Generation Matrix | | Colombia | 2015 - Dec 2016 | Population | General Population | Valega-Mackenzie (2023) |
| 0.881 | | | Compartmental Model | | Colombia | 2016 - 2016 | Population | General Population | Aranda (2019) |
| 1.318 | | | Compartmental Model | | Brazil | 2015 - 2015 | Population | General Population | Liang (2019) |
| 1.519 | 95% CI: 1.508-1.531 | | Unspecified | 10234 | Costa Rica | Jan 2016 - Dec 2017 | Population | General Population | Sanchez (2019) |
| 1.66 | | | Next Generation Matrix | | Itabuna (Brazil) | 2016 - 2016 | Population | General Population | Hirata (2023) |
| 1.667 | 95% Crl: 0.147-4.85 | | Next Generation Matrix | | Suriname | 2015 - 22 Jun 2016 | Population | Persons Under Investigation | Shutt (2017) |

continued on next page

continued from previous page

| Parameter value | Uncertainty | Disaggr-egation | Method for R estimation | Sample size | Location | Dates | Population sample type | Population group | Reference |
|---|---|---|---|---|---|---|---|---|---|
| 1.8 | | | Next Generation Matrix | | Cuiaba (Brazil) | 2016 - 2016 | Population | General Population | Hirata (2023) |
| 1.87 | | | Next Generation Matrix | | Rio de Janeiro (Brazil) | 2016 - 2016 | Population | General Population | Hirata (2023) |
| 1.99 | 95% CI: 1.97-2.01 | | Next Generation Matrix | | Costa Rica | 12 Apr 2016 - 24 Aug 2016 | Population | General Population | Luo (2021) |
| 2.1 | 95% CI: 1.8-2.5 | | Unspecified | | Salvador, Bahia (Brazil) | Jan 2015 - 28 May 2016 | Population | General Population | Netto (2017) |
| 2.2 | 95% CI: 1.1-3.8 | | Branching Process | 330 | Reynosa (Mexico) | 2017 - 2017 | Population | General Population | Olson (2020) |
| 2.459 | 95% CrI: 0.152-7.402 | | Next Generation Matrix | | El Salvador | 2015 - 22 Jun 2016 | Population | Persons Under Investigation | Shutt (2017) |
| 2.76 | | | Next Generation Matrix | | Goiania (Brazil) | 2016 - 2016 | Population | General Population | Hirata (2023) |
| 4.554 | | | Next Generation Matrix | | El Salvador | Oct 2015 - Apr 2016 | Population | General Population | Kumar (2017) |
| 7.6 | 95% CI: 4.8-14 | Method | Next Generation Matrix | 108 | Yap Main Islands (Federated States of Micronesia) | Apr 2007 - Jul 2007 | Population | Persons Under Investigation | Funk (2016) |
| 0 - 8.31 | | Region | Other | 377525 | Multi-country: Americas (n = 21) | 15 Aug 2015 - 11 Jun 2016 | Population | Persons Under Investigation | Ogden (2017) |
| 0.5 - 2.5 | | | Unspecified | | Multi-country: Brazil, Colombia, French Polynesia | Aug 2013 - Jun 2016 | Population | General Population | He (2017) |
| 1.15 - 2.75 | | Other | Ross-Macdonald Formula | | Manacapuru (Brazil) | Feb 2014 - Feb 2015 | | | Abad-Franch (2017) |
| 0 - 4 | | | Unspecified | | Global | 2015 - 2016 | Mixed | General Population | Sun (2020) |
| **Basic Reproduction Number - Vector-Borne** | | | | | | | | | |
| 1.04 | 95% CI: 1.03-1.05 | | Next Generation Matrix | | Costa Rica | 12 Apr 2016 - 24 Aug 2016 | Population | General Population | Luo (2021) |
| 1.103 | | | Next Generation Matrix | | Colombia | 2015 - Dec 2016 | Population | General Population | Valega-Mackenzie (2023) |
| 1.51 | 95% CI: 1.23-1.87 | Method | Compartmental Model | 18364 | Colombia | 2015 - 2016 | Population | Pregnant Women | Sasmal (2018) |
| 1.713 | | | Compartmental Model | | Brazil | 25 Mar 2016 - 14 Apr 2018 | Population | Persons Under Investigation | Zhao (2020) |
| 1.96 | 95% CI: 0.45-6.227 | | Compartmental Model | | Multi-country: Brazil, Colombia, El Salvador | Apr 2015 - 27 Feb 2016 | Population | General Population | Gao (2016) |
| 2.04 | 95% CI: 1.53-2.55 | | Next Generation Matrix | | Brazil | Jan 2015 - Sep 2016 | Population | General Population | Wang (2024a) |
| 2.222 | | | Next Generation Matrix | | Puerto Rico (United States) | Jan 2016 - Dec 2017 | Population | General Population | Valega-Mackenzie (2023) |
| 2.475 | | | Next Generation Matrix | | Brazil | 06 Feb 2016 - 02 Jun 2016 | Population | Persons Under Investigation | Wang (2017) |
| 2.96 | 95% CrI: 2.58-3.39 | | Ross-Macdonald Formula | | Unspecified | Unspecified | Unspecified | Other | Armstrong (2020) |
| 4.05 | 95% CrI: 3.22-5.17 | | Ross-Macdonald Formula | | Unspecified | Unspecified | Unspecified | Other | Armstrong (2020) |
| 4.274 | 95% CrI: 0.166-12.941 | | Next Generation Matrix | | El Salvador | 2015 - 22 Jun 2016 | Population | Persons Under Investigation | Shutt (2017) |
| 6.093 | 95% CrI: 0.6-15.738 | | Next Generation Matrix | | Suriname | 2015 - 22 Jun 2016 | Population | Persons Under Investigation | Shutt (2017) |
| **Effective Reproduction Number** | | | | | | | | | |
| 0.16 | 95% CI: 0.13-0.19 | | Branching Process | | Florida (United States) | 01 May 2016 - 23 Sep 2016 | Population | General Population | Dinh (2016) |
| 0.24 | 95% CI: 0.07-0.41 | | Compartmental Model | | Singapore | 04 Sep 2016 - 30 Nov 2016 | Population | General Population | Zhu (2022) |

continued on next page

continued from previous page

| Parameter value | Uncertainty | Disaggr-egation | Method for R estimation | Sample size | Location | Dates | Population sample type | Population group | Reference |
|---|---|---|---|---|---|---|---|---|---|
| 0.62 | Range: 0.25-1.06 | | Unspecified | | Feira de Santana (Brazil) | 2017 - 30 Apr 2017 | Population | General Population | Lourenco (2017) |
| 1.22 | 95% CI: 1.19-1.24 | Time | Other | | Singapore | 01 Sep 2016 - 24 Nov 2016 | | Persons Under Investigation | Singapore Zika Study Group (2017) |
| 1.45 | | | Compartmental Model | | Suriname | Unspecified | Population | General Population | Dénes (2019) |
| 1.47 | | | Compartmental Model | | Costa Rica | 2016 - 2018 | Population | General Population | Dénes (2019) |
| 1.54 | 95% CI: 1.43-1.65 | Method | Branching Process | 11247972 | Rio Grande (Brazil) | Oct 2015 - May 2016 | Population | Persons Under Investigation | Zhao (2019) |
| 1.63 | Range: 1-2 | | Other | | Colombia | 31 May 2015 - 16 Apr 2016 | Population | Persons Under Investigation | Majumder (2016) |
| 1.63 | 95% CI: 1.58-1.68 | Method | Branching Process | 15203934 | Bahia (Brazil) | Oct 2015 - May 2016 | Population | Persons Under Investigation | Zhao (2019) |
| 1.67 | 95% CI: 1.56-1.79 | Method | Branching Process | 3265486 | Mato Grosso (Brazil) | Oct 2015 - May 2016 | Population | Persons Under Investigation | Zhao (2019) |
| 1.85 | 95% CI: 1.75-1.95 | Method | Branching Process | 9345173 | Pernambuco State (Brazil) | Oct 2015 - May 2016 | Population | Persons Under Investigation | Zhao (2019) |
| 1.96 | Range: 1-3 | | Other | | Colombia | 22 Aug 2015 - 16 Apr 2016 | Population | Persons Under Investigation | Majumder (2016) |
| 2.13 | 95% CI: 2.07-2.19 | Method | Branching Process | 803513 | Acre (Brazil) | Oct 2015 - May 2016 | Population | Persons Under Investigation | Zhao (2019) |
| 2.2 | | Time | Next Generation Matrix | | Northeastern (Brazil) | Jan 2015 - Nov 2016 | Population | Persons Under Investigation | He (2020) |
| 2.82 | 95% CI: 2.66-2.99 | Method | Branching Process | 11163018 | Panara (Brazil) | Oct 2015 - May 2016 | Population | Persons Under Investigation | Zhao (2019) |
| 3.05 | 95% CI: 2.73-3.41 | Method | Branching Process | 3929911 | Espirito Santo (Brazil) | Oct 2015 - May 2016 | Population | Persons Under Investigation | Zhao (2019) |
| 3.07 | 95% CI: 2.92-3.24 | Method | Branching Process | 6610681 | Goiania City (Brazil) | Oct 2015 - May 2016 | Population | Persons Under Investigation | Zhao (2019) |
| 0.7 - 2.2 | | Time | Branching Process | | Salvador, Bahia (Brazil) | 2015 - 2015 | Population | General Population | Rodriguez-Barraquer (2019) |
| 0.882 - 0.941 | | Method | Compartmental Model | | Brazil | 25 Mar 2016 - 14 Apr 2018 | Population | Persons Under Investigation | Zhao (2020) |
| 0 - 3.1 | | Time | Compartmental Model | 3681000 | Puerto Rico (United States) | Dec 2015 - Dec 2016 | Unspecified | General Population | Biswas (2024) |
| **Effective Reproduction Number - Sexual** | | | | | | | | | |
| 0.05 | | | Compartmental Model | | Brazil | 25 Mar 2016 - 14 Apr 2018 | Population | Persons Under Investigation | Zhao (2020) |
| 3.3 | 95% CI: 1.3-5.3 | | Growth Rate | | Managua (Nicaragua) | Jun 2016 - Jun 2016 | Community | Children | Gordon (2019) |
| 3.4 | 95% CI: 2.4-4.7 | | Growth Rate | | Managua (Nicaragua) | Jun 2016 - Jul 2016 | Community | Children | Gordon (2019) |
| 1.6 - 10.3 | | Method, Time | Other | | Antioquia (Colombia) | 14 Oct 2015 - 10 Apr 2016 | Population | General Population | Chowell (2016) |
| 0.85 - 1.71 | | | Next Generation Matrix | | "north-eastern" (Brazil) | Jan 2015 - Sep 2016 | Unspecified | Unspecified | Yuan (2021) |
| **Effective Reproduction Number - Vector-Borne** | | | | | | | | | |
| 0.857 | | | Compartmental Model | | Brazil | 25 Mar 2016 - 14 Apr 2018 | Population | Persons Under Investigation | Zhao (2020) |
| **Growth Rate (R)** | | | | | | | | | |
| 0.076 per day | 95% CI: 0.066-0.087 | | | 359 | Barranquilla (Colombia) | 01 Oct 2015 - Nov 2015 | Hospital | Persons Under Investigation | Towers (2016) |
| 0.1 | 95% CI: 0.1-0.2 | | | | Managua (Nicaragua) | Jun 2016 - Jul 2016 | Community | Children | Gordon (2019) |
| 0.2 | 95% CI: 0.1-0.4 | | | | Managua (Nicaragua) | Jun 2016 - Jul 2016 | Community | Children | Gordon (2019) |

continued on next page

continued from previous page

| Parameter value | Uncertainty | Disaggr-egation | Method for R estimation | Sample size | Location | Dates | Population sample type | Population group | Reference |
|---|---|---|---|---|---|---|---|---|---|
| 0.823 per week | SD: 0.053 | | Growth Rate | | Rio de Janeiro (Brazil) | 18 Oct 2015 | | | Villela (2017) |
| **Sexual Transmission Contribution** | | | | | | | | | |
| 1.4 % | | | | | "north-eastern" (Brazil) | Jan 2015 - Sep 2016 | Unspecified | Unspecified | Yuan (2021) |
| 3.5 % | | | | | Brazil | Jan 2015 - Sep 2016 | Population | General Population | Wang (2024a) |
| 15.36 % | 95% CI: 12.83-17.14 | Method | Compartmental Model | 18364 | Colombia | 2015 - 2016 | Population | Pregnant Women | Sasmal (2018) |
| 23 % | 95% CI: 1-47 | | | 359 | Barranquilla (Colombia) | 01 Oct 2015 - Nov 2015 | Hospital | Persons Under Investigation | Towers (2016) |
| 32 % | | | | | Suriname | Unspecified | Population | General Population | Dénes (2019) |
| 48.15 % | | | | | Costa Rica | 12 Apr 2016 - 24 Aug 2016 | Population | General Population | Luo (2021) |
| 54 % | | | | | Costa Rica | 2016 - 2018 | Population | General Population | Dénes (2019) |
| **Vector Transmission Contribution** | | | | | | | | | |
| 51.85 % | | | | | Costa Rica | 12 Apr 2016 - 24 Aug 2016 | Population | General Population | Luo (2021) |
| 98.6 % | | | | | "north-eastern" (Brazil) | Jan 2015 - Sep 2016 | Unspecified | Unspecified | Yuan (2021) |

Table B.12: Overview of extracted transmission parameters.

##### B.7.7 Genomic parameters

| Parameter value | Uncertainty | Genome site | Sample size | Dates | Population sample type | Population group | Reference |
|---|---|---|---|---|---|---|---|
| **Evolutionary Rate** | | | | | | | |
| 0.6 s/s/y $10^{-3}$ | 95% CI: 0.3-0.8 | | 40 | 1966 - 2017 | Unspecified | Unspecified | Barzilai (2019) |
| 0.8 s/s/y $10^{-3}$ | 95% CI: 0.5-1.1 | | 40 | 2013 - 2017 | Unspecified | Unspecified | Barzilai (2019) |
| 1.18 s/s/y $10^{-3}$ | | | 238 | Jun 2017 - Jun 2019 | Population | Persons Under Investigation | Da Costa Castilho (2024) |
| 0.98 - 1.06 s/s/y $10^{-3}$ | | near-complete genome | | 1947 - Dec 2015 | | | Faria (2016) |
| 8.61 s/s/y $10^{-4}$ | HPDI 95%: 6.34-11.1 | Whole genome sequence | 88 | Aug 2018 | Unspecified | Unspecified | Liu (2019) |
| 8.65 s/s/y $10^{-4}$ | | Whole genome sequence | 360 | Unspecified | Unspecified | Unspecified | Black (2019) |
| 9.2 s/s/y $10^{-4}$ | 95% CrI: 7.1-12 | Whole genome sequence | 65 | 1966 - Aug 2017 | | | Delatorre (2017) |
| 1.09 s/s/y $10^{-3}$ | 95% CrI: 0.77-1.43 | Whole genome sequence | 59 | 15 Dec 2015 - Mar 2017 | Population | General Population | Giovanetti (2020) |
| 1.12 s/s/y $10^{-3}$ | 95% CrI: 0.97-1.27 | Whole genome sequence | 254 | 1966 - 01 Mar 2016 | Unspecified | Persons Under Investigation | Faria (2017) |
| 1.2 s/s/y $10^{-3}$ | HPDI 95%: 0.951-1.41 | Whole genome sequence | 39 | 2007 - 2016 | Unspecified | Unspecified | Fajardo (2016) |
| 1.55 s/s/y $10^{-3}$ | HPDI 95%: 1.06-2.05 | Whole genome sequence | 49 | Dec 2014 - Oct 2016 | | | Aldunate (2017) |
| 1.57 s/s/y $10^{-3}$ | | Whole genome sequence | 139 | 2016 | | | Boskova (2018) |
| 1.76 s/s/y $10^{-3}$ | | Whole genome sequence | 67 | 2016 | | | Boskova (2018) |
| 5.71 s/s/y $10^{-3}$ | | Whole genome sequence | 269 | 2013 - Aug 2018 | Unspecified | Unspecified | Grubaugh (2019) |
| 6.42 s/s/y $10^{-3}$ | | Whole genome sequence | 23 | Jun 2016 - 11 Oct 2016 | | | Boskova (2018) |
| 0.53 - 4.39 s/s/y $10^{-3}$ | | Whole genome sequence | 58 | 1947 - 2016 | | | Yokoyama (2017) |
| **Mutation Rate** | | | | | | | |
| 0.37 Mutations/site/generation (mu) | HPDI 95%: 0.298-0.448 | NS5, 1st codon | 108 | 1947 - Oct 2017 | Population | Persons Under Investigation | Shao (2020) |
| 0.16 Mutations/site/generation (mu) | HPDI 95%: 0.113-0.211 | NS5, 2nd codon | 108 | 1947 - Oct 2017 | Population | Persons Under Investigation | Shao (2020) |
| 2.471 Mutations/site/generation (mu) | HPDI 95%: 2.382-2.555 | NS5, 3rd codon | 108 | 1947 - Oct 2017 | Population | Persons Under Investigation | Shao (2020) |
| **Substitution Rate** | | | | | | | |
| 12 - 25 Mutations/year | | | 33 | 1947 - 06 Feb 2016 | Unspecified | Unspecified | Logan (2016) |
| 7.95 s/s/y $10^{-4}$ | | Whole genome sequence | 214 | 1966 - 2023 | Mixed | Persons Under Investigation | Khongwichit (2023) |
| 8 s/s/y $10^{-4}$ | HPDI 95%: 6.4-9.7 | Whole genome sequence | 135 | 1947 - 2016 | Unspecified | Unspecified | Ebranati (2019) |
| 1.114 s/s/y $10^{-3}$ | HPDI 95%: 0.993-1.254 | Whole genome sequence | 182 | 01 Mar 2017 | Mixed | Unspecified | Le Hingrat (2019) |

Table B.13: Overview of extracted genomic parameters.

##### B.7.8 Severity

| Parameter value | Uncertainty | Disaggr-egation | Naive or Adjusted | Numerator | Denominator | Population country | Dates | Population sample type | Population group | Reference |
|---|---|---|---|---|---|---|---|---|---|---|
| **Case fatality ratio** | | | | | | | | | | |
| 8.3 % | 95% CI: 7.2-9.6 | | Naive | 171 | 2063 | Brazil | 10 Nov 2015 - 15 Oct 2016 | Hospital | Other | Cunha (2017) |
| 10.5 % | 95% CI: 9.5-11.7 | | Unspecified | 313 | 2974 | Brazil | 10 Nov 2015 - 15 Oct 2016 | Hospital | Other | Cunha (2017) |
| 38 % | | | Naive | 6 | 16 | Rio de Janeiro (Brazil) | Jan 2016 - Oct 2016 | Hospital | Persons Under Investigation | Pereira Jr (2019) |
| 8.4 % | | | Unspecified | 35 | 418 | Colombia | Jul 2015 - Oct 2017 | Population | Persons Under Investigation | Charniga (2021a) |
| 0.04 % | | | Unspecified | | 5161 | Dominican Republic | 2016 - 2016 | Population | General Population | Petrone (2021) |
| % | | | | 2 | 87 | Guadeloupe, Martinique (France) | 06 Jan 2016 | Hospital | Persons Under Investigation | Lannuzel (2019) |
| **Proportion of symptomatic cases** | | | | | | | | | | |
| 27 % | 95% CrI: 15-37 | Method | | | | Yap Island (Federated States of Micronesia) | 2007 - 2007 | Unspecified | Unspecified | Mitchell (2019) |
| 23.2 % | | | | 133 | 573 | French Guiana (France) | 01 Feb 2016 - 01 Jun 2016 | Population | Pregnant Women | Flamand (2017) |
| 44 % | 95% CrI: 26-66 | Method | | | | French Polynesia (France) | 2013 - 2014 | Unspecified | Unspecified | Mitchell (2019) |
| 44 % | 95% CI: 26-69 | | Naive | 8 | 18 | Society Islands, French Polynesia (France) | Feb 2014 - Mar 2014 | Population | General Population | Aubry (2017) |
| 47 % | 95% CI: 40-55 | | Naive | 73 | 154 | Society Islands, French Polynesia (France) | Sep 2015 - Nov 2015 | Population | General Population | Aubry (2017) |
| 55 % | 95% CI: 34-75 | | Naive | 12 | 22 | Tuamotu Islands, French Polynesia (France) | Feb 2014 - Mar 2014 | Population | General Population | Aubry (2017) |
| 57 % | 95% CI: 39-75 | | Naive | 16 | 28 | Marquesas Islands, French Polynesia (France) | Feb 2014 - Mar 2014 | Population | General Population | Aubry (2017) |
| 66 % | 95% CI: 48-83 | | Naive | 19 | 29 | Austral–Gambier Islands, French Polynesia (France) | Feb 2014 - Mar 2014 | Population | General Population | Aubry (2017) |
| 71 % | 95% CI: 66-76 | | Naive | 221 | 312 | Society Islands, French Polynesia (France) | May 2014 - Jun 2014 | School | Children | Aubry (2017) |
| 24 % | | | Naive | 7 | 29 | Congenital Zika Program at Children's National (CZPCN) in Washington DC (United States) | Jan 2016 - Jun 2018 | Hospital | Pregnant Women | Mulkey (2021) |
| 50 % | 95% CrI: 34-92 | Method | | | | Puerto Rico (United States) | 2016 - 2016 | Unspecified | Unspecified | Mitchell (2019) |

Table B.14: Overview of extracted severity parameters.

##### B.7.9 Relative contributions

| Parameter value | Uncertainty | Population country | Dates | Population sample type | Population group | Reference |
|---|---|---|---|---|---|---|
| **Relative contribution - sexual** | | | | | | |
| 1.4 % | | "north-eastern" (Brazil) | Jan 2015 - Sep 2016 | Unspecified | Unspecified | Yuan (2021) |
| 3.5 % | | Brazil | Jan 2015 - Sep 2016 | Population | General Population | Wang (2024a) |
| 15.36 % | 95% CI: 12.83-17.14 | Colombia | 2015 - 2016 | Population | Pregnant Women | Sasmal (2018) |
| 23 % | 95% CI: 1-47 | Barranquilla (Colombia) | 01 Oct 2015 - Nov 2015 | Hospital | Persons Under Investigation | Towers (2016) |
| 48.15 % | | Costa Rica | 12 Apr 2016 - 24 Aug 2016 | Population | General Population | Luo (2021) |
| 54 % | | Costa Rica | 2016 - 2018 | Population | General Population | Dénes (2019) |
| 32 % | | Suriname | Unspecified | Population | General Population | Dénes (2019) |
| **Relative contribution - vector-borne** | | | | | | |
| 98.6 % | | "north-eastern" (Brazil) | Jan 2015 - Sep 2016 | Unspecified | Unspecified | Yuan (2021) |
| 51.85 % | | Costa Rica | 12 Apr 2016 - 24 Aug 2016 | Population | General Population | Luo (2021) |

Table B.15: Overview of extracted relative contribution parameters.

##### B.7.10 Adverse birth outcomes

| Parameter value | Uncertainty | Naive or Adjusted | Numerator | Denom-inator | Population country | Dates | Population sample type | Reference |
|---|---|---|---|---|---|---|---|---|
| **Miscarriage probability** | | | | | | | | |
| | | Naive | 1 | 4 | Veterans Health Administration in US and Caribbean | 01 Dec 2015 - 31 Oct 2016 | Hospital | Schirmer (2018) |
| 0 % | | Unspecified | 0 | 44 | Jundiai, São Paulo (Brazil) | 01 Mar 2016 - 23 Aug 2017 | Hospital | Clemente (2020) |
| 0.83 % | | Naive | 1 | 120 | Mato Grosso do Sul (Brazil) | Nov 2017 - Oct 2018 | Population | De Sousa (2020) |
| % | | Naive | 1 | 92 | Rio de Janeiro (Brazil) | 01 Sep 2015 - 31 May 2016 | Other | Pereira (2018) |
| 4.8 % | | Unspecified | 6 | 134 | Rio de Janeiro state (Brazil) | Sep 2015 - May 2016 | Hospital | Brasil (2016) |
| % | | Naive | 1 | 6 | São Paulo (Brazil) | Jan 2016 - Sep 2016 | Hospital | Tozetto-Mendoza (2019) |
| 1.3 % | | Unspecified | 16 | 1259 | Cali (Colombia) | Oct 2015 - Jul 2016 | Hospital | Cañas (2020) |
| 2 % | | Naive | 75 | 4481 | Colombia | 15 Jun 2015 - 31 Jul 2016 | Population | Ospina (2020) |
| 2.3 % | | Unspecified | 10 | 499 | Villavicencio (Colombia) | Oct 2015 - Jul 2016 | Hospital | Cañas (2020) |
| 3 % | | Naive | 172 | 5673 | Colombia | 15 Jun 2015 - 31 Jul 2016 | Population | Ospina (2020) |
| 4.7 % | | Unspecified | 5 | 115 | Colombia | Feb 2017 - Mar 2019 | Population | Tannis (2024) |
| | | Naive | 10 | 171 | Valle del Cauca (Colombia) | Nov 2015 - Jan 2017 | Population | Calle-Giraldo (2019) |
| 8 % | | Naive | 97 | 1192 | Colombia | 15 Jun 2015 - 31 Jul 2016 | Population | Ospina (2020) |
| 13.1 % | | Unspecified | 26 | 296 | Dominican Republic | Jan 2016 - Apr 2017 | Population | Peña (2019) |
| 0 % | | Unspecified | 0 | 252 | Martinique, Guadeloupe, French Guiana (France) | Mar 2016 - 27 Apr 2017 | Population | Hoen (2018) |
| 0 % | | Unspecified | 0 | 114 | Martinique, Guadeloupe, French Guiana (France) | Mar 2016 - 27 Apr 2017 | Population | Hoen (2018) |
| 2 % | | Unspecified | 11 | 555 | Martinique, Guadeloupe, French Guiana (France) | Mar 2016 - 27 Apr 2017 | Population | Hoen (2018) |
| | | Naive | 11 | 546 | Guadeloupe, Martinique, French Guiana (France) | 02 Mar 2016 - 24 Nov 2016 | Other | Grant (2022a) |
| % | | Unspecified | 11 | 300 | French Guiana (France) | 01 Jan 2015 - 15 Jul 2016 | Mixed | Pomar (2020) |
| 4 % | 95% CI: 2-7 | Naive | 12 | 291 | French Guiana (France) | Jan 2016 - Jul 2016 | Hospital | Pomar (2018) |
| 5.8 % | | Unspecified | 11 | 189 | Martinique, Guadeloupe, French Guiana (France) | Mar 2016 - 27 Apr 2017 | Population | Hoen (2018) |

continued on next page

continued from previous page

| Parameter value | Uncertainty | Naive or Adjusted | Numerator | Denom- inator | Population country | Dates | Population sample type | Reference |
|---|---|---|---|---|---|---|---|---|
| | | Naive | 1 | 4 | Spain | Jan 2016 - Feb 2020 | Travel | Marban-Castro (2021) |
| 0 % | | Unspecified | 0 | 12 | Bueng Kan, Mudkahan provinces (Thailand) | May 2018 - Jan 2020 | Population | Wongsawat (2024) |
| | | Unspecified | 1 | 70 | New York (United States) | Mar 2016 - Apr 2016 | Hospital | Merriam (2020) |
| % | | Unspecified | 47 | 442 | continental United States and Hawaii (United States) | 15 Jan 2016 - 22 Sep 2016 | Population | Honein (2017) |
| **Zika congenital syndrome (microcephaly) probability** | | | | | | | | |
| 0 % | | Unspecified | 0 | 16 | Antigua and Barbuda | May 2015 - Jan 2018 | Population | Morris (2020) |
| 3.13 % | | Unspecified | 1 | 32 | Barbados | May 2015 - Jan 2018 | Population | Morris (2020) |
| 0 % | | Unspecified | 0 | 1 | Belize | May 2015 - Jan 2018 | Population | Morris (2020) |
| 7.41 % | | Unspecified | 14 | 189 | Bolivia | May 2015 - Jan 2018 | Population | Morris (2020) |
| 0 % | | Naive | 0 | 6 | São Paulo (Brazil) | 01 Jan 2016 - Sep 2016 | Hospital | Tozetto-Mendoza (2019) |
| 0.006 % | | Unspecified | | | Tocantins (Brazil) | Jul 2015 - Dec 2016 | Hospital | Rodrigues (2020) |
| 0.49 - 2.1 % | | Unspecified | | | Northeast Brazil (Brazil) | 15 Nov 2015 - 02 Apr 2016 | Other | Alfaro-Murillo (2016) |
| 1.8 % | 95% CI: 0.2-10 | Naive | 1 | 38 | Pernambuco (Recife) (Brazil) | Dec 2015 - Jun 2017 | Hospital | Ximenes (2021) |
| % | | Unspecified | 15 | 780 | Brazil | 2016 - 2016 | Hospital | Martins (2019) |
| 2.2 % | 95% CI: 0.7-6.6 | Naive | 3 | 86 | Pernambuco (Recife) (Brazil) | Dec 2015 - Jun 2017 | Hospital | Ximenes (2021) |
| 2.5 % | 95% CI: 1.2-5.2 | Naive | 7 | 278 | Pernambuco (Recife) (Brazil) | Dec 2015 - Jun 2017 | Hospital | Ximenes (2021) |
| 3.4 % | | Unspecified | 4 | 117 | Rio de Janeiro state (Brazil) | 01 Jan 2016 - 31 Jul 2016 | Hospital | Brasil (2016) |
| 3.6 % | | Naive | 3 | 83 | Jundiai Hospital Medical school (Brazil) | 01 Mar 2016 - 30 Jun 2019 | Hospital | Gazeta (2021) |
| 3.7 % | | Unspecified | 8 | 216 | Rio de Janeiro (Brazil) | Dec 2015 - Dec 2016 | Hospital | Brasil (2020) |
| 3.8 % | 95% CI: 1.8-7.8 | Naive | 7 | 154 | Pernambuco (Recife) (Brazil) | Dec 2015 - Jun 2017 | Hospital | Ximenes (2021) |
| % | | Unspecified | 2 | 48 | São Paulo state (Brazil) | 01 Mar 2016 - 23 Aug 2017 | Hospital | Sanchez Clemente (2020) |
| 4.5 % | | Unspecified | 2 | 44 | Jundiai, São Paulo (Brazil) | 01 Mar 2016 - 23 Aug 2017 | Hospital | Clemente (2020) |
| 4.8 % | | Unspecified | 5 | 109 | Tangará da Serra (Brazil) | 01 Jan 2016 - 31 Dec 2016 | Other | Herrero Da Silva (2022) |

continued on next page

continued from previous page

| Parameter value | Uncertainty | Naive or Adjusted | Numerator | Denom- inator | Population country | Dates | Population sample type | Reference |
|---|---|---|---|---|---|---|---|---|
| 5 % | | Naive | 2 | 43 | Private hospital Salvador, Bahia (Brazil) | 01 Jan 2015 - 31 Jan 2017 | Hospital | Aromolaran (2022) |
| 5.2 % | | Unspecified | 4 | 77 | Manaus, Amazonas (Brazil) | Mar 2016 - Jun 2018 | Hospital | Abtibol-Bernardino (2022) |
| 7 % | 95% CI: 1.5-19.1 | Unspecified | 3 | 43 | Central-West (Brazil) | Jan 2017 - Apr 2019 | Hospital | Rosado (2023) |
| 7.1 % | | Unspecified | 7 | 98 | Rio de Janeiro (Brazil) | 2016 - 2016 | Other | De Melo Espindola (2021) |
| % | | Naive | 7 | 92 | Rio de Janeiro (Brazil) | 01 Sep 2015 - 31 May 2016 | Other | Pereira (2018) |
| 8.7 % | 95% CrI: 7.13-11.39 | Unspecified | | | Brazil | 11 Aug 2015 - 31 Dec 2018 | Population | Moore (2020) |
| % | | Naive | 4 | 40 | Mato Grosso do Sul (Brazil) | Nov 2017 - Oct 2018 | Population | De Sousa (2020) |
| 11.33 % | | Unspecified | 2952 | 26066 | Brazil | May 2015 - Jan 2018 | Population | Morris (2020) |
| % | | Naive | 6 | 51 | Mato Grosso do Sul (Brazil) | Nov 2017 - Oct 2018 | Population | De Sousa (2020) |
| % | | Unspecified | 76 | 602 | Brazil | 19 Nov 2015 - 27 Feb 2016 | Population | França (2016) |
| % | | Naive | 23 | 120 | Mato Grosso do Sul (Brazil) | Nov 2017 - Oct 2018 | Population | De Sousa (2020) |
| 24.7 % | | Naive | 53 | 219 | Rio de Janeiro (Brazil) | Sep 2015 - Jun 2017 | Hospital | Cranston (2020) |
| 33 % | | Naive | 42 | 127 | Public hospital Salvador, Bahia (Brazil) | 01 Jan 2015 - 31 Jan 2017 | Hospital | Aromolaran (2022) |
| 38.5 % | | Unspecified | | 104 | Salvador, Bahia (Brazil) | Nov 2015 - Feb 2016 | Hospital | Robbiani (2019) |
| % | | Naive | 12 | 28 | Mato Grosso do Sul (Brazil) | Nov 2017 - Oct 2018 | Population | De Sousa (2020) |
| 49 % | | Naive | 54 | 110 | Rio de Janeiro (Brazil) | 01 Mar 2016 - 30 Jun 2017 | Hospital | Pool (2019) |
| 70.7 % | | Naive | 53 | 75 | Campina Grande, Paraíba (Brazil) | Mar 2016 - Dec 2018 | Hospital | Sampaio (2021) |
| % | | Unspecified | 0 | 30 | Colombia | Oct 2015 - Jan 2017 | Hospital | Gutierrez-Sanchez (2022) |
| 0 % | | Naive | 0 | 616 | Colombia | 09 Aug 2015 - 02 Apr 2016 | Other | Pacheco (2020) |
| 0 % | | Unspecified | 0 | 93 | Colombia | Feb 2017 - Mar 2019 | Population | Tannis (2024) |
| 1.24 % | | Unspecified | 248 | 19993 | Colombia | May 2015 - Jan 2018 | Population | Morris (2020) |
| % | | Unspecified | 1 | 50 | Valle del Cauca (Colombia) | Feb 2018 - Apr 2020 | Population | Calle-Giraldo (2019) |
| % | | Unspecified | 3 | 75 | Valle del Cauca (Colombia) | Jan 2017 - Mar 2019 | Population | Calle-Giraldo (2019) |
| 4.5 % | | Naive | 7 | 154 | Valle del Cauca (Colombia) | Nov 2015 - Jan 2017 | Population | Calle-Giraldo (2019) |
| 8.8 % | | Unspecified | 9 | 102 | Colombia | Oct 2015 - Jan 2017 | Hospital | Gutierrez-Sanchez (2022) |

continued on next page

continued from previous page

| Parameter value | Uncertainty | Naive or Adjusted | Numerator | Denom- inator | Population country | Dates | Population sample type | Reference |
|---|---|---|---|---|---|---|---|---|
| % | | Unspecified | 5 | 53 | Colombia | Oct 2015 - Jan 2017 | Hospital | Gutierrez-Sanchez (2022) |
| % | | Unspecified | 3 | 29 | Valle del Cauca (Colombia) | Dec 2016 - Feb 2018 | Population | Calle-Giraldo (2019) |
| % | | Unspecified | 9 | 25 | Colombia | Oct 2015 - Jan 2017 | Hospital | Gutierrez-Sanchez (2022) |
| 9.05 % | | Unspecified | 19 | 210 | Costa Rica | May 2015 - Jan 2018 | Population | Morris (2020) |
| 0 % | | Unspecified | 0 | 30 | Curaçao | May 2015 - Jan 2018 | Population | Morris (2020) |
| 0 % | | Unspecified | 0 | 13 | Dominica | May 2015 - Jan 2018 | Population | Morris (2020) |
| 0 % | | Unspecified | 0 | 296 | Dominican Republic | Jan 2016 - Apr 2017 | Population | Peña (2019) |
| 8.8 % | | Unspecified | 85 | 966 | Dominican Republic | May 2015 - Jan 2018 | Population | Morris (2020) |
| 11 % | | Unspecified | 87 | 800 | Dominican Republic | 2016 | Hospital | Pimintel (2021) |
| 1.54 % | | Unspecified | 14 | 912 | Ecuador | May 2015 - Jan 2018 | Population | Morris (2020) |
| 1.02 % | | Unspecified | 4 | 391 | El Salvador | May 2015 - Jan 2018 | Population | Morris (2020) |
| 0 % | | Unspecified | 0 | 11 | Saint Barthelemy (France) | May 2015 - Jan 2018 | Population | Morris (2020) |
| 0.05 % | | Unspecified | 1 | 2211 | French Guiana (France) | May 2015 - Jan 2018 | Population | Morris (2020) |
| 0.6 % | | Unspecified | 5 | 830 | Martinique (France) | May 2015 - Jan 2018 | Population | Morris (2020) |
| 0.61 % | | Unspecified | 5 | 815 | Guadeloupe (France) | May 2015 - Jan 2018 | Population | Morris (2020) |
| % | | Naive | 3 | 129 | western French Guiana (France) | Jan 2016 - Sep 2016 | Hospital | Hcini (2021) |
| 3.2 % | | Unspecified | 8 | 252 | Martinique, Guadeloupe, French Guiana (France) | Mar 2016 - 27 Apr 2017 | Population | Hoen (2018) |
| 4.4 % | | Unspecified | 5 | 114 | Martinique, Guadeloupe, French Guiana (France) | Mar 2016 - 27 Apr 2017 | Population | Hoen (2018) |
| % | | Naive | 17 | 320 | Guadeloupe, Martinique, French Guiana (France) | 02 Mar 2016 - 24 Nov 2016 | Other | Grant (2022a) |
| 5.8 % | | Unspecified | 32 | 555 | Martinique, Guadeloupe, French Guiana (France) | Mar 2016 - 27 Apr 2017 | Population | Hoen (2018) |
| 10 % | | Naive | 27 | 273 | French Guiana (France) | Jan 2016 - Jul 2016 | Hospital | Pomar (2018) |
| 10.1 % | | Unspecified | 19 | 189 | Martinique, Guadeloupe, French Guiana (France) | Mar 2016 - 27 Apr 2017 | Population | Hoen (2018) |
| 9.9 % | | Unspecified | 140 | 1414 | Guatemala | May 2015 - Jan 2018 | Population | Morris (2020) |

continued on next page

continued from previous page

| Parameter value | Uncertainty | Naive or Adjusted | Numerator | Denom-inator | Population country | Dates | Population sample type | Reference |
|---|---|---|---|---|---|---|---|---|
| 4.55 % | | Unspecified | 1 | 22 | Haiti | May 2015 - Jan 2018 | Population | Morris (2020) |
| 1.17 % | | Unspecified | 8 | 681 | Honduras | May 2015 - Jan 2018 | Population | Morris (2020) |
| 33.3 % | 95% CI: 4.3-77.7 | Unspecified | 2 | 6 | Alonso Suazo Health Center (Honduras) | Jul 2016 - 31 Dec 2016 | Hospital | Alger (2021) |
| 0 % | | Unspecified | 0 | 712 | Jamaica | May 2015 - Jan 2018 | Population | Morris (2020) |
| % | | Naive | 2 | 166 | Mombasa (Kenya) | Oct 2017 - Jul 2019 | Hospital | Osoro (2022) |
| 0.35 % | | Unspecified | 20 | 5667 | Mexico | May 2015 - Jan 2018 | Population | Morris (2020) |
| 0.07 % | 95% CrI: 0.01-0.19 | Unspecified | | | Nicaragua | 27 Jan 2016 - 21 Nov 2016 | Population | Moore (2020) |
| 0.18 % | | Unspecified | 2 | 1117 | Nicaragua | May 2015 - Jan 2018 | Population | Morris (2020) |
| 8.02 % | | Unspecified | 17 | 212 | Panama | May 2015 - Jan 2018 | Population | Morris (2020) |
| 6.45 % | | Unspecified | 2 | 31 | Paraguay | May 2015 - Jan 2018 | Population | Morris (2020) |
| 0 % | | Unspecified | 0 | 279 | Peru | May 2015 - Jan 2018 | Population | Morris (2020) |
| 0 % | | Unspecified | 0 | 10 | Sint Maarten (Dutch part) | May 2015 - Jan 2018 | Population | Morris (2020) |
| 15.8 % | 95% CI: 3.4-39.6 | Naive | 3 | 19 | Spain | Jan 2016 - Feb 2019 | Travel | Soriano-Arandes (2020) |
| % | | Naive | 2 | 12 | Barcelona (Spain) | Jan 2016 - Apr 2019 | Population | Martinez-Arias (2023) |
| | | Unspecified | 2 | 11 | Barcelona (Spain) | May 2016 - Dec 2021 | Other | Romaní (2022) |
| 2.08 % | | Unspecified | 1 | 48 | St. Martin (French part) | May 2015 - Jan 2018 | Population | Morris (2020) |
| 0 % | | Unspecified | 0 | 3 | St. Vincent and the Grenadines | May 2015 - Jan 2018 | Population | Morris (2020) |
| 3.67 % | | Unspecified | 17 | 463 | Trinidad and Tobago | May 2015 - Jan 2018 | Population | Morris (2020) |
| | | Unspecified | 0 | 70 | New York (United States) | Mar 2016 - Apr 2016 | Hospital | Merriam (2020) |
| 1.16 % | | Unspecified | 47 | 4047 | Puerto Rico (United States) | May 2015 - Jan 2018 | Population | Morris (2020) |
| 2.6 % | | Unspecified | 3 | 114 | Ponce, Puerto Rico (United States) | 01 May 2017 - 28 Feb 2020 | Population | Alvarado-Domenech (2022) |
| 4 % | | Unspecified | 18 | 442 | continental United States and Hawaii (United States) | 15 Jan 2016 - 22 Sep 2016 | Population | Honein (2017) |
| 4.5 % | | Unspecified | 18 | 404 | New York City (United States) | Jan 2016 - Dec 2017 | Hospital | Lee (2020) |
| 5.26 % | | Unspecified | 5 | 95 | Miami, Florida (United States) | 01 Jan 2016 - 31 Dec 2017 | Population | Walker (2020) |
| 0 % | | Unspecified | 0 | 84 | Saint Lucia | May 2015 - Jan 2018 | Population | Morris (2020) |
| 0 % | | Unspecified | 0 | 3463 | Venezuela | May 2015 - Jan 2018 | Population | Morris (2020) |

continued on next page

continued from previous page

| Parameter value | Uncertainty | Naive or Adjusted | Numerator | Denom-inator | Population country | Dates | Population sample type | Reference |
|---|---|---|---|---|---|---|---|---|
| % | | Unspecified | 3 | 21 | Ho Chi Minh City (Vietnam) | Mar 2016 - Nov 2017 | Hospital | Grant (2021) |
| 0 % | | Unspecified | 0 | 286 | Virgin Islands (U.S.) | May 2015 - Jan 2018 | Population | Morris (2020) |

Table B.16: Overview of extracted CZS probability and pregnancy loss probability parameters.

### C PRISMA 2020 Checklists

| Section & Topic | Item # | Checklist item | Reported (Yes/No) |
|---|---|---|---|
| **Title** | | | |
| Title | 1 | Identify the report as a systematic review. | Yes |
| **Background** | | | |
| Objectives | 2 | Provide an explicit statement of the main objective(s) or question(s) the review addresses. | Yes |
| **Methods** | | | |
| Eligibility criteria | 3 | Specify the inclusion and exclusion criteria for the review. | Yes |
| Information sources | 4 | Specify the information sources (e.g. databases, registers) used to identify studies and the date when each was last searched. | Yes |
| Risk of bias | 5 | Specify the methods used to assess risk of bias in the included studies. | Yes |
| Synthesis of results | 6 | Specify the methods used to present and synthesise results. | Yes |
| **Results** | | | |
| Included studies | 7 | Give the total number of included studies and participants and summarise relevant characteristics of studies. | Yes |
| Synthesis of results | 8 | Present results for main outcomes, preferably indicating the number of included studies and participants for each. If meta-analysis was done, report the summary estimate and confidence/credible interval. If comparing groups, indicate the direction of the effect (i.e. which group is favoured). | Yes |
| **Discussion** | | | |
| Limitations of evidence | 9 | Provide a brief summary of the limitations of the evidence included in the review (e.g. study risk of bias, inconsistency and imprecision). | Yes |
| Interpretation | 10 | Provide a general interpretation of the results and important implications. | Yes |
| **Other** | | | |
| Funding | 11 | Specify the primary source of funding for the review. | Yes |
| Registration | 12 | Provide the register name and registration number. | Yes |

Table C.17: PRISMA 2020 Abstracts Checklist. ([14])

| Section & Topic | Item # | Checklist item | Location where item is reported |
|---|---|---|---|
| **Title** | | | |
| Title | 1 | Identify the report as a systematic review. | page 1 |
| **Abstract** | | | |
| Abstract | 2 | See the PRISMA 2020 for Abstracts checklist. | Table C.17 |
| **Introduction** | | | |
| Rationale | 3 | Describe the rationale for the review in the context of existing knowledge. | page 4 |
| Objectives | 4 | Provide an explicit statement of the objective(s) or question(s) the review addresses. | page 4 |
| **Methods** | | | |
| Eligibility criteria | 5 | Specify the inclusion and exclusion criteria for the review and how studies were grouped for the syntheses. | page 4/5 |

Table C.18: PRISMA 2020 Checklist. ([14])

| Section & Topic | Item # | Checklist item | Location where item is reported |
|---|---|---|---|
| Information sources | 6 | Specify all databases, registers, websites, organisations, reference lists and other sources searched or consulted to identify studies. Specify the date when each source was last searched or consulted. | page 4/5 |
| Search strategy | 7 | Present the full search strategies for all databases, registers and websites, including any filters and limits used. | page 4, Figure 1, Section appendix A.1 |
| Selection process | 8 | Specify the methods used to decide whether a study met the inclusion criteria of the review, including how many reviewers screened each record and each report retrieved, whether they worked independently, and if applicable, details of automation tools used in the process. | page 4, Section A.1 |
| Data collection process | 9 | Specify the methods used to collect data from reports, including how many reviewers collected data from each report, whether they worked independently, any processes for obtaining or confirming data from study investigators, and if applicable, details of automation tools used in the process. | page 5, Section A.1 |
| Data items | 10a | List and define all outcomes for which data were sought. Specify whether all results that were compatible with each outcome domain in each study were sought (e.g. for all measures, time points, analyses), and if not, the methods used to decide which results to collect. | page 4,5, Sections A.1 and A.2 |
| | 10b | List and define all other variables for which data were sought (e.g. participant and intervention characteristics, funding sources). Describe any assumptions made about any missing or unclear information. | page 4,5, Sections A.1 and A.2 |
| Study risk of bias assessment | 11 | Specify the methods used to assess risk of bias in the included studies, including details of the tool(s) used, how many reviewers assessed each study and whether they worked independently, and if applicable, details of automation tools used in the process. | page 6 |
| Effect measures | 12 | Specify for each outcome the effect measure(s) (e.g. risk ratio, mean difference) used in the synthesis or presentation of results. | page 4, 5 Section A.2 |
| Synthesis methods | 13a | Describe the processes used to decide which studies were eligible for each synthesis (e.g. tabulating the study intervention characteristics and comparing against the planned groups for each synthesis (item #5)). | page 5/6, Sections A.2 and A.3 |
| | 13b | Describe any methods required to prepare the data for presentation or synthesis, such as handling of missing summary statistics, or data conversions. | page 4page 5/6, Sections A.2 and A.3 |
| | 13c | Describe any methods used to tabulate or visually display results of individual studies and syntheses. | page 5/6, Sections A.2 and A.3 |
| | 13d | Describe any methods used to synthesize results and provide a rationale for the choice(s). If meta-analysis was performed, describe the model(s), method(s) to identify the presence and extent of statistical heterogeneity, and software package(s) used. | page 5/6, Sections A.2 and A.3 |
| | 13e | Describe any methods used to explore possible causes of heterogeneity among study results (e.g. subgroup analysis, meta-regression). | page 6 |
| | 13f | Describe any sensitivity analyses conducted to assess robustness of the synthesized results. | - |
| Reporting bias assessment | 14 | Describe any methods used to assess risk of bias due to missing results in a synthesis (arising from reporting biases). | page 6 |

Table C.18: PRISMA 2020 Checklist. ([14])

| Section & Topic | Item # | Checklist item | Location where item is reported |
|---|---|---|---|
| Certainty assessment | 15 | Describe any methods used to assess certainty (or confidence) in the body of evidence for an outcome. | page 4-6, Section A |
| **Results** | | | |
| Study selection | 16a | Describe the results of the search and selection process, from the number of records identified in the search to the number of studies included in the review, ideally using a flow diagram. | page 6, Figure 1 |
| | 16b | Cite studies that might appear to meet the inclusion criteria, but which were excluded, and explain why they were excluded. | Figure 1 and see epireview for list of excluded studies |
| Study characteristics | 17 | Cite each included study and present its characteristics. | pages 4-13 and Section A.1 |
| Risk of bias in studies | 18 | Present assessments of risk of bias for each included study. | page 6, list of included studies on epireview (Section A.1 |
| Results of individual studies | 19 | For all outcomes, present, for each study: (a) summary statistics for each group (where appropriate) and (b) an effect estimate and its precision (e.g. confidence/-credible interval), ideally using structured tables or plots. | pages 6-8, Appendix pages 44-117 |
| Results of syntheses | 20a | For each synthesis, briefly summarise the characteristics and risk of bias among contributing studies. | page 8, Sections B.5 and B.6 |
| | 20b | Present results of all statistical syntheses conducted. If meta-analysis was done, present for each the summary estimate and its precision (e.g. confidence/credible interval) and measures of statistical heterogeneity. If comparing groups, describe the direction of the effect. | page 8, Sections B.5 and B.6 |
| | 20c | Present results of all investigations of possible causes of heterogeneity among study results. | page 6-8 |
| | 20d | Present results of all sensitivity analyses conducted to assess the robustness of the synthesized results. | page 6-8 |
| Reporting biases | 21 | Present assessments of risk of bias due to missing results (arising from reporting biases) for each synthesis assessed. | pages 6-8 |
| Certainty of evidence | 22 | Present assessments of certainty (or confidence) in the body of evidence for each outcome assessed. | pages 6-8 |
| **Discussion** | | | |
| Discussion | 23a | Provide a general interpretation of the results in the context of other evidence. | page 8-10 |
| | 23b | Discuss any limitations of the evidence included in the review. | page 8-10 |
| | 23c | Discuss any limitations of the review processes used. | page 8-10 |
| | 23d | Discuss implications of the results for practice, policy, and future research. | page 8-10 |
| **Other Information** | | | |

Table C.18: PRISMA 2020 Checklist. ([14])

| Section & Topic | Item # | Checklist item | Location where item is reported |
|---|---|---|---|
| Registration and protocol | 24a | Provide registration information for the review, including register name and registration number, or state that the review was not registered. | page 4 |
| | 24b | Indicate where the review protocol can be accessed, or state that a protocol was not prepared. | page 4 |
| | 24c | Describe and explain any amendments to information provided at registration or in the protocol. | n/a |
| Support | 25 | Describe sources of financial or non-financial support for the review, and the role of the funders or sponsors in the review. | page 11 |
| Competing interests | 26 | Declare any competing interests of review authors. | page 11 |
| Availability of data, code and other materials | 27 | Report which of the following are publicly available and where they can be found: template data collection forms; data extracted from included studies; data used for all analyses; analytic code; any other materials used in the review. | page 10 |

Table C.18: PRISMA 2020 Checklist. ([14])

### D Pathogen Epidemiology Review Group (PERG) Membership

| First name | Surname | Affiliation |
|---|---|---|
| Aaron | Morris | University of Oxford |
| Alpha | Forna | University of Georgia |
| Amy | Dighe | Johns Hopkins |
| Anna | Vicco | Imperial College London |
| Anna-Maria | Hartner | Robert Koch Institute |
| Anne | Cori | Imperial College London |
| Arran | Hamlet | Imperial College London |
| Ben | Lambert | University of Oxford |
| Bethan | Cracknell Daniels | Imperial College London |
| Charles | Whittaker | Imperial College London |
| Christian | Morgenstern | Imperial College London |
| Cosmo | Santoni | Imperial College London |
| Cyril | Geismar | Imperial College London |
| Dariya | Nikitin | Imperial College London |
| David | Jorgensen | Imperial College London |
| Dominic | Dee | Imperial College London |
| Ed | Knock | Imperial College London |
| Gina | Cuomo-Dannenburg | Imperial College London |
| Hayley | Thompson | PATH |
| Ilaria | Dorigatti | Imperial College London |
| Isobel | Routledge | UCSF |
| Jack | Wardle | Imperial College London |
| Janetta | Skarp | Imperial College London |
| Joseph | Hicks | Imperial College London |
| Juliette | Unwin | University of Bristol |
| Kanchan | Parchani | Imperial College London |

| | | |
|---|---|---|
| Keith | Fraser | Imperial College London |
| Kelly | Charniga | Imperial College London |
| Kelly | McCain | Imperial College London |
| Kieran | Drake | Imperial College London |
| Lily | Geidelberg | Imperial College London |
| Lorenzo | Cattarino | UKHSA |
| Mantra | Kusumgar | Imperial College London |
| Mara | Kont | Imperial College London |
| Marc | Baguelin | Imperial College London |
| Natsuko | Imai-Eaton | Imperial College London |
| Pablo | Perez Guzman | Imperial College London |
| Patrick | Doohan | Imperial College London |
| Paul | Lietar | Imperial College London |
| Paula | Christen | Imperial College London |
| Rebecca | Nash | Imperial College London |
| Rich | Fitzjohn | Imperial College London |
| Richard | Sheppard | Imperial College London |
| Rob | Johnson | Imperial College London |
| Ruth | McCabe | Imperial College London |
| Sabine | van Elsland | Imperial College London |
| Sangeeta | Bhatia | Imperial College London |
| Sequoia | Leuba | Imperial College London |
| Shazia | Ruybal-Pesantez | Imperial College London |
| Sreejith | Radhakrishnan | University of Glasgow |
| Thomas | Rawson | Imperial College London |
| Tristan | Naidoo | Imperial College London |
| Zulma | Cucunuba Perez | Pontificia Universidad Javeriana |

Table D.19: Pathogen Epidemiology Review Group (PERG) membership as of July 2024.
